# Plasma multi-omics outlines association of urobilinogen with corticosteroid non-response, inflammation and leaky gut in Sever Alcoholic Hepatitis

**DOI:** 10.1101/2023.03.06.23286831

**Authors:** Manisha Yadav, Babu Mathew, Sadam H Bhat, Neha Sharma, Jitender Kumar, Pushpa Yadav, Gaurav Tripathi, Vasundhra Bindal, Nupur Sharma, Sushmita Pandey, Ravinder Singh, Ashima Bhaskar, Ved Prakash Dwivedi, Nirupama Trehanpati, Shvetank Sharma, Shiv Kumar Sarin, Jaswinder Singh Maras

## Abstract

**Background and Aims:** Severe alcoholic hepatitis (SAH) has a high mortality and corticosteroid therapy is effective in 60% patients. Reliable indicators of response to therapy and mortality in SAH are needed. A total of 223 SAH patients, 70 in derivative [50 responders (R) and 20 non-responders (NR)] and 153 in validation cohort [136R, 17NR] were subjected to plasma metabolic/meta-proteomic analysis using UHPLC-HRMS and validated using Machine-Learning (ML). Temporal metabolic changes were assessed using Weighted Metabolome Correlation Network Analysis (WMCNA). Functionality (inflammatory-nature, effect on membrane integrity and glucocorticoid receptor) of non-response indicator was assessed *in-vitro* on primary healthy neutrophils or mice enterocytes. Baseline plasma metabolomics and meta-proteomics clearly discriminated NR and showed significant increase in urobilinogen (3.6-fold), cholesterol sulfate (6.9-fold), Adenosine monophosphate (4.7-fold) and others (p<0.05, FC>1.5, FDR<0.01). Increase in alpha/beta diversity, biosynthesis of secondary metabolites was a characteristic feature of NR (p<0.05). NR were metabolically inactive however R showed temporal change in the metabolite expression post-corticosteroid therapy (p<0.05). Plasma urobilinogen predicted non-response [AUC=0.94] with a hazard-ratio of 1.5(1.2-1.6) and cut-off >0.07mg/ml segregated non-survivors (p<0.01) and showed >98% accuracy using ML. Plasma urobilinogen directly correlated with circulating bacterial peptides linked to bilirubin to urobilinogen metabolising bacteria (r^2^>0.7;p<0.05). Urobilinogen induced **neutrophil activation**, **oxidative-stress** and **pro-inflammatory cytokine**s (CXCR1, NGAL, NOXO1, NOX4, IL15, TNFα and others, p<0.05), promoted **corticosteroid resistance** by increasing the expression of GR-Beta and trans-repression genes under GR-alpha (inflammatory-NFkB, MAPK-MAP) and reducing GR-alpha, and transactivation (anti-inflammatory) gene levels. Urobilinogen also promoted leaky gut by deregulating intestinal membrane junction proteins.

**Conclusion:** Plasma metabolome/meta-proteome can stratify pre-therapy steroid response. Increase in plasma Urobilinogen pedals a vicious cycle of bacterial translocation and increase in inflammation and corticosteroid non-response in SAH patients.

## Introduction

Severe Alcoholic Hepatitis (SAH) is often associated with systemic inflammatory response syndrome, organ failure and high short-term mortality (1). Model for End Stage Liver Disease (MELD) score ≥20 and a Maddrey’s Discriminant Function (mDF) score ≥31 are used as cut-offs to deduce the severity in SAH (3, 4). For decades, liver transplant has been the only effective treatment. Corticosteroid therapy used as a standard medical care (5) improves short-term survival and is effective only in 60% of SAH patients (6). Over-exposure of corticosteroids increases predisposition to secondary bacterial infections, sepsis and early mortality in SAH (7, 8). Therefore, reliable indicators capable of segregating NR and R and can help us in determining the efficacies of corticosteroids in SAH are needed.

Further, untargeted plasma metabolomics is now actively promulgated to evaluate mechanisms underlying various pathologies (10, 11). Recently, Moreau et al. documented the utility of plasma metabolomics in showcasing that inflammation-associated mitochondrial dysfunction is a potential mechanism underlying ACLF patients (11). Javier Michelena et al. also worked on serum samples and showed the utility of metabolomic markers in accurate, non-invasive diagnosis and prognosis of alcoholic hepatitis (13). For SAH patients particularly on corticosteroid therapy such plasma based indicators which could predict response are of urgent need.

Dysbiosis is an unfavourable pathological event seen in SAH patients and presence of circulating microbial products can result in persistent stimulation of immune system (14) (15). Further prolong exposure of corticosteroids increases the risk of bacterial infection, sepsis and other complications (16). Recently, an increase in alcohol consumption has been proposed as the primary driver in changing circulating microbiomes (17). Therefore, robust analysis of the circulating bacterial peptide, microbial products and associated functions in corticosteroid responders and non-responders is warranted.

Increase in the circulatory bilirubin and urobilinoids are associated with high severity in SAH patients (18). Intestinal flora converts bilirubin to urobilinogen, 82% of which is transported to the kidney and excreted out through urine or converted to stercobilin or urobilin and then excreted out via feces (19). Recently, pro-inflammatory nature of stercobilin has been documented (10) but urobilinogen being the major urobilinoid has not been studied so far. Since bilirubin metabolism is bacterial driven and dysbiosis is often seen in the pathogenesis of SAH, it becomes necessary to gain insight into the overall metabolic state and the host-microbiome interactions in the plasma of SAH.

In the current study, the overall metabolic status and circulating microbiome of SAH patients was studied using plasma metabolomics and meta-proteomics respectively. Baseline plasma metabolic indicator which could segregate responders from non-responders correlates with disease severity and mortality at baseline was identified. Temporal changes in the metabolic phenotype post corticosteroid therapy in R and NR were also studied. Baseline meta-proteome (microbiome) was correlated with the baseline metabolome to understand the host-microbiome interactions. Results found an increase in bacteria associated with Urobilinogen (metabolic indicator of NR and outcome) in NR-SAH. Finally, functional properties (inflammatory nature, effect on membrane integrity, glucocorticoid receptor and downstream signalling) of identified marker were assessed in-vitro on primary healthy neutrophils (PHN) and its role in modulating intestinal permeability was studied on primary mice enterocytes (PME).

### Patients and Methods

SAH patients seen at Department of Hepatology, Institute of Liver and Biliary Science, New Delhi, India (2017-2020), and confirmed to have mDF >31, recent onset of jaundice, chronic alcohol abuse, and liver biochemistry and histologic features of SAH (n= 250) were screened for corticosteroid therapy (6). 27 patients were excluded due to presence of hepatitis B and C virus, hepatocellular carcinoma, portal vein thrombosis, or recent variceal bleed. Written informed consent was obtained from all the patients and the study was approved by the Institutional Ethics Committee.

Baseline demographic profile and clinical characteristics of all SAH patients were recorded. Plasma samples were collected at baseline and after day-3 and day-7 of prednisolone therapy (given as standard of care in SAH patients) (40mg/day) initiation. Patients were characterized as R and NR based on day-7 Lille’s score (6). SAH patients were randomly distributed into the derivative and validation cohort, where both the clinical and the laboratory personals were kept blind for grouping. In brief, of the 223 patients, 70 patients (30%) were grouped in derivative cohort, and the remaining 153 patients (70%) were grouped in validation cohort. Leads from the metabolomic studies in the derivative cohort were validated using high resolution MS and ML in the validation cohort.

### Untargeted Metabolomics and Meta-proteomics analysis using UHPLC High Resolution-MS

Untargeted metabolomic and meta-proteomic analyses of plasma samples were performed as detailed in **supplementary methods**.

#### Construction of Weighted Metabolome Co-Expression Network (WMCNA)

Method involved in the construction of WMCNA and Module Trait Relationship is detailed in **supplementary methods.**

#### Neutrophil Activation and oxidative Assay

PHN (>98% pure) were treated with urobilinogen (5 to120uM), LPS-10ng/ml and PMA-10ng/ml. CD66b+PE and CD11b+PECy7 (neutrophil markers) neutrophil populations were assessed for activation and inflammation markers using anti-CXCR1-BV510, and anti-TNFa-PerCPCy5.5 respectively. Intracellular ROS was measured by using 1uM 1’,7’-dichlorodihydrofluorescein diacetate (H2DCFDA). Mean fluorescence intensity (MFI) for CD66b+, CD11b+, CXCR1+, TNFa+, and (H2DCFDA) ROS was measured using Flow Cytometry.

### Semi-quantitative RT-PCR Analysis

Neutrophil activation and inflammation status was studied using a panel of 17 genes associated with activation (NGAL), intracellular antioxidants (CCS, GSR, SOD1, SOD1, and SOD3), ROS production (NOXO1, NOXA1, NOX4, and NOX1) and inflammatory cytokines (IL15, IL7, IL4, TNFa, IL8, IL6, and IL11) (11).

### Proteomic analysis of activated neutrophil and primary mice enterocytes

PHN and PME were isolated as detailed in supplementary methods. Neutrophils treated with urobilinogen (effective dose: 60uM), PMA, LPS, U+Pred, PMA+Pred, LPS+Pred, PMA+U, PMA+U+Pred, LPS+U and LPS+U+Pred were subjected to proteomic analysis as detailed in the **supplementary method**. PME were treated with urobilinogen (effective dose: 60uM) alone, LPS (30ng/ml) and Alcohol (5%) in presence and absence of U for 90 min and were subjected to proteomic analysis as detailed in the **supplementary method.**

### Statistical Analysis

Results are shown as mean and SD unless indicated otherwise. Statistical analysis was performed using Graph Pad Prism-version 6.0, and SPSS-version 10; and p-values <0.05 using Benjamini-Hochberg correction were considered significant. Unpaired (two-tailed) Student t-test and the Mann-Whitney U-test were performed for comparison of two groups. For temporal analysis a paired t-test was performed. For comparison among more than two groups, ANOVA and the Kruskal-Wallis tests were performed. All correlations were performed using Spearman correlation analysis, and R^1^ > 0.5 and p<0.05 were considered statistically significant. Potential metabolic candidates with highest AUC, FC, and significant p-value were identified at baseline to predict non-responsiveness. Comparative AUROC analysis was performed for top identified metabolite and other clinical variables such as DF, CTP, and MELD score in order to compute sensitivity and specificity and to obtain cut-offs for each variable. Based on the AUROC diagnostic cut-off, prediction of mortality was performed using Kaplan–Meier method and compared between the two groups (log-rank test, hazard ratio, and 95% confidence limit estimated using Cox regression Model). The metabolic indicator identified in the derivative cohort was then validated using MS and ML.

### Machine Learning Validation

Five different ML algorithms: LDA (Linear Discriminant Analysis), KNN (K-Nearest Neighbor), CART (Classification and Regression Tree), SVM (Support Vector Machine), and RF (Random Forest) were used to validate and compare the sensitivity and specificity of candidate indicators with clinical variables such as MELD, CTP and mDF as detailed in **supplementary methods**.

## RESULTS

### Patient Characterization

Baseline demographic profile of NR and R in both the derivative and validation cohorts was similar except for platelet count, leukocyte count and 90 day mortality, which were significantly higher in NR (Supplementary table-1).

### Characterization of Baseline Plasma Metabolome and Meta-Proteome

Plasma metabolomics (discovery cohort) led to the identification of 10164 and 13084 features in negative and positive electrospray ionization mode. Of them, 711 features were annotated based on m/z matching structure (spectral database), retention time and/or mass. Functionally these metabolites showed enrichment of amino acids, fatty acids, mono-saccharides, purines, pyrimidine, indoles, steroids, TCA, quinone, bilirubin, bile acids, and others super-classes (p<0.05) (Supplementary figure-1, Supplementary table-2). Plasma Meta-proteome analysis identified 349 unique peptide sequences belonging to different class of bacteria (Supplementary figure-2, supplementary table-3).

### Baseline Plasma Metabolome Robustly Distinguishes Non Responders

The flow chart in Figure 1A explains the study design. Untargeted metabolomics was performed in plasma samples of 223 SAH patients at baseline, day3, and day7, and temporal changes in the metabolic profile were studied at baseline, day3, and day7. Meta-proteome profile was studied at baseline and correlated with metabolome profile to identify metabolite-microbiome interactions in SAH. Metabolite indicator of non-response and early mortality was identified and validated using MS and ML (Figure 1A).

**Figure 1:**
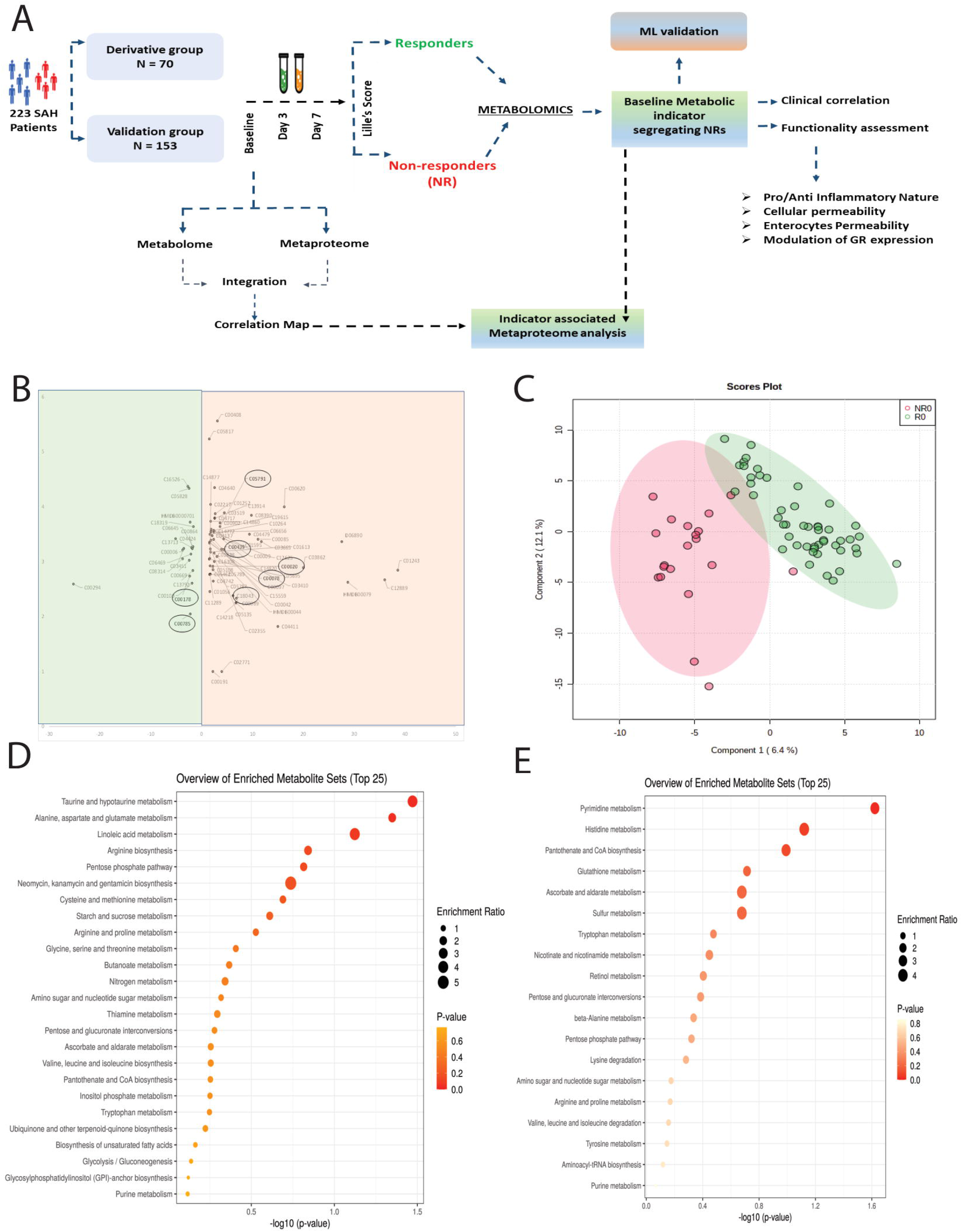
Baseline Plasma Metabolome Robustly Distinguishes Non Responders (NR) **1A.** Flow chart explains the blueprint of this study. 223 patients were enrolled and their metabolic profile was studied at baseline, day3 and day7. Baseline metabolomic and meta-proteomic profile were integrated to identify the host-microbiome interactions. Metabolomics was used to identify the baseline indicator capable of segregating NR and R at baseline. Functional properties of identified indicator of non-response as indicated were also investigated. **1B.** Volcano plot shows differentially expressed metabolites between NR and R at baseline. Y-axis corresponds to –log10 p-value, and x-axis corresponds to log1 fold change. The green colour in background indicates the differentially downregulated metabolites whereas red colour in background highlights the differentially upregulated metabolites at baseline. Encircled metabolites highlights the panel of metabolites which were selected to segregate responders and non-responders. **1C.** Partial least squares discriminant analysis (PLS-DA) score plot shows distinct metabotype of NR (red dot) and R (green dot) at baseline. **1D.** Dot plot shows KEGG pathway enrichment analysis of metabolites with significantly higher expression at baseline in NR compared to R (FC± 1.5, p-value<0.05). **1E.** Dot plot shows KEGG pathway enrichment analysis of metabolites with lower expression at baseline in NR compared to R (FC± 1.5, p-value<0.05).

A total of 193 (149 up-, 44 down-regulated) DEMs were identified at baseline in NR compared to R (p<0.05; Figure 1B, Supplementary table-4). PLS-DA analysis significantly segregated NR from R (Figure 1C, loading plot: Supplementary figure-3). NR showed a significant increase in metabolites associated with taurine/hypo-taurine metabolism, alanine, aspartate, and glutamate metabolism, linoleic acid metabolism, and others (p<0.05; Figure 1D) whereas metabolites linked to pyrimidine metabolism, histidine metabolism, and others, were decreased in NR (p<0.05; Figure 1E). Together these results suggest that the metabolic profile of NR is distinct from R and shows increased catabolism of amino acid, fatty acid, and others which functions as the fuel for the activation of immune cells. These differences could be exploited for the stratification of NR from R.

### WMCNA highlights Corticosteroid mediated changes in the metabolic profile are more pronounced in responders

To gain insights into the temporal changes that occurred upon corticosteroid administration in NR and R, plasma metabolome of baseline, day3 and day7 were compared. The expression of a plethora of metabolites got significantly downregulated in NR by day3 and day7 post steroid administration (Figure 2A, Supplement table-5). PLS-DA analysis showed a time-wise segregation of R from NR, suggesting metabolic variability in these patients (Figure 2B). Clustering analysis showed the inactive metabolic status of NR post-corticosteroid administration as the expression of metabolites barely changed over time whereas, in R, the metabolic profile changed in a temporal manner (Figure 2B).

**Figure 2.**
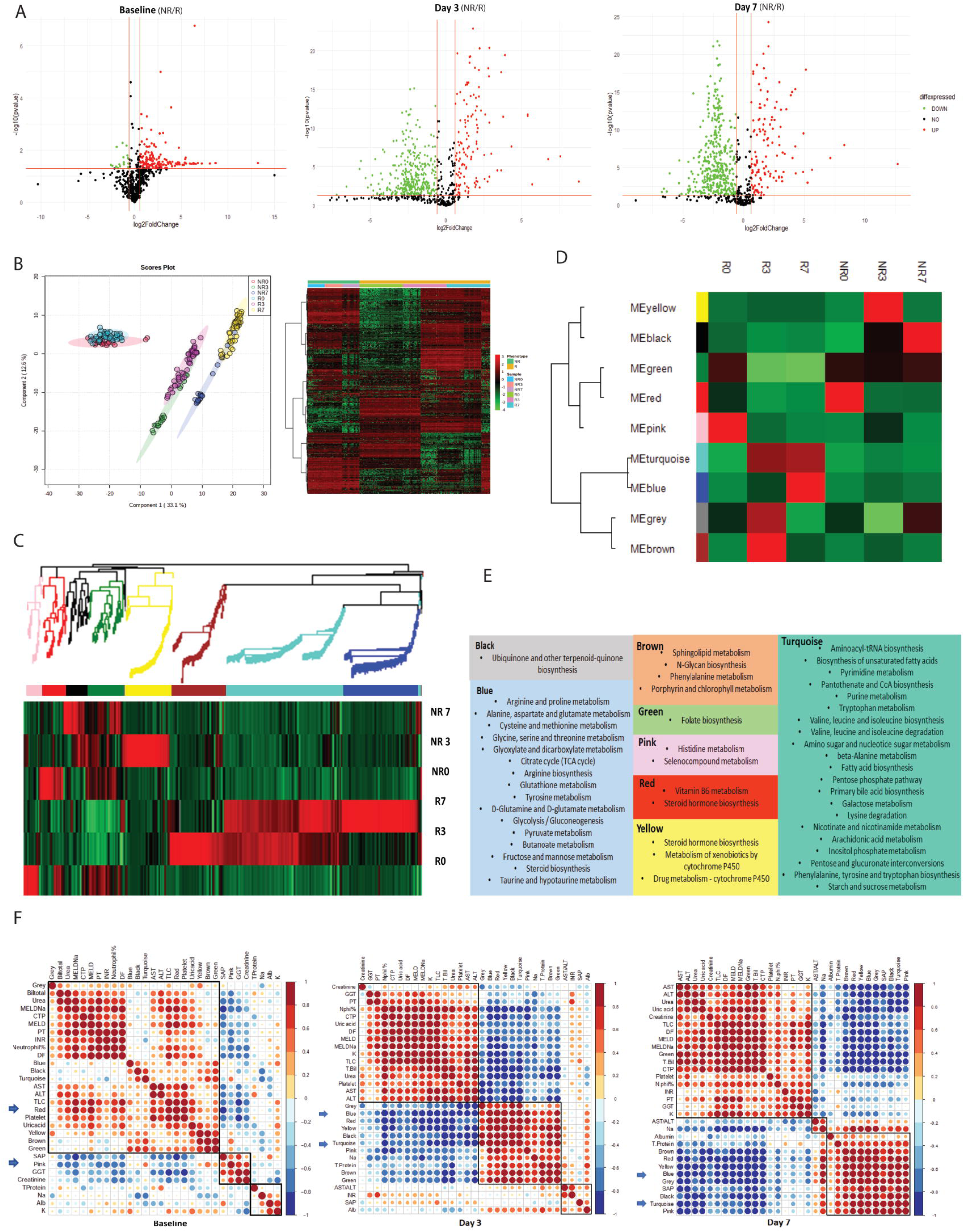
Corticosteroid mediated changes in the metabolic profile: **2A.** Volcano plot displaying temporal changes in expression of metabolites in NR vs. R at baseline and post-initiation of corticosteroid therapy at day 3, and day 7. Red dotes correspond to higher expression (FC>1.5, p-value<0.05) of metabolites in plasma, whereas green corresponds to lower expression (FC<1.5, p-value<0.05) in NR respectively. **2B.** Partial least squares – discriminant analysis score plot showing increased bifurcation in the metabolic profile of NR and R post initiation of corticosteroid therapy at day 3 and day 7. Multivariate heatmap showing the expression of metabolites at different days i.e. at baseline, day 3 and day 7 in NR and R. The R is represented by orange, and the NR is represented by turquoise, respectively. **2C.** Weighted Metabolite Co-expressed Network Analysis (WMCNA) followed by Hierarchical clustering showing 9 modules of co-expressed metabolites in NR and R highlighted by different colour bars. The heatmap shows the expression of modules (cluster of metabolites) in NR and R at baseline, Day 3 and Day 7. **2D.** Heatmap showing module trait (R, NR at baseline, Day 3 and Day 7) relationship identified by WMCNA. Each module is represented by the colour name. Expression of the module ranges from red (upregulated) to green (downregulated). Module red is NR specific (upregulated in NR at baseline) whereas pink module is R specific. **2E.** Pathway enrichment analysis. Each coloured-panel in the figure corresponds to pathways significantly associated with module of respective colour (p-value <0.05). **2F.** Correlation plot indicating the correlation of 9 identified modules with clinical parameters at baseline, day 3, and day 7 respectively.

Metabolic profiles of NR and R on day0, day3, and day7 were subjected to WMCNA to identify metabolic modules (metabolite clusters showing a similar trend in their expression over time) specific to R, NR, and severity indices. WMCNA clustered 711 metabolites into 9 modules (Figure 2C). The heatmap clearly shows activation and inhibition of a myriad of metabolites in R upon corticosteroid exposure and almost no effect of corticosteroid therapy in NR (Figure 2C). Module trait relationship (phenotype of R and NR at baseline, day3, and day7 are referred to as a trait) clustering analysis (Figure 2D) identified the ‘Pink’ module specific to R (up-regulated at baseline) which was linked to histidine and seleno-compound metabolism, whereas ‘Red’ module specific to NR (up-regulated at baseline) linked to steroid hormone biosynthesis and vitamin B6 metabolism (Figure 2D, and 2E). Metabolites in the ‘Blue’ and ‘Turquoise’ modules temporally increased in R (Figure 2D) and were associated with pathways linked to Citrate cycle (TCA cycle), Arginine biosynthesis, Glutathione metabolism, Tyrosine metabolism, Glycolysis/Gluconeogenesis, and others (Figure 2E). The statistical composition of each metabolite module is shown in Supplementary figure 7. At baseline NR specific ‘Red’ module correlated directly with the severity indices (MELD, CTP, DF, and others) whereas response-specific modules such as ‘Pink’, ‘Blue’, and ‘Turquoise’ correlated inversely with the severity indices from baseline to day 7 (Figure 2F). Hence WMCNA confirmed the notion that Corticosteroid therapy changes the metabolic profile only in R. The metabolic profile of NR remained almost unchanged upon corticosteroid administration with a predominant increase in steroid hormone biosynthesis. Further, up-regulation of the ‘Blue’ and ‘Turquoise’ modules reflects responsiveness to corticosteroid therapy in R.

### MS analysis and Machine learning validates that baseline plasma Level of Urobilinogen indicates non-response and correlates with Outcome in SAH Patients

The flow chart in Figure 3A explains the approach adopted to identify the metabolic indicator capable of segregating NR at baseline. In the derivative cohort, a panel of 8 metabolites (C05791-urobilinogen, C18043-cholestrol sulphate, C00020-AMP, C00439-formimidoyl-L-glutamate, C00078-tryptophan, C01835-maltotriose, C00785-urocanic acid, and C00178-thymine) based on significant p-value, high AUC and FC ±1.5 folds (Figure 3B) were selected which could segregate NR from R at baseline and were validated in a separate cohort of SAH patients (NR=17, R=136) (Figure 4B). C05791; Urobilinogen was selected as a metabolic indicator of non-response at baseline as it showed the highest AUROC value of 0.947 among all other selected metabolites (Figure 3C, Supplementary figure-8). COX multivariate analysis also identified urobilinogen as an independent predictor of mortality with a maximum Hazard Ratio (HR-1.469) (Figure 3C). Further, based on the AUROC of urobilinogen of 0.947 and the likelihood ratio of 3.1, a cut-off of 0.07 mg/ml was determined to assess the survival in SAH patients. Patients with urobilinogen levels higher than 0.07mg/ml showed higher mortality (log-rank <0.01, Figure 3D, Supplementary figure 9, and 10). When compared to other clinical factors (MELD, DF, and CTP), urobilinogen attained the highest AUROC of 0.83 (0.76-0.90) for the prediction of poor outcomes in SAH (Figure 3E). Together these results suggest urobilinogen as an independent predictor of corticosteroid response and short-term mortality in SAH patients.

**Figure 3.**
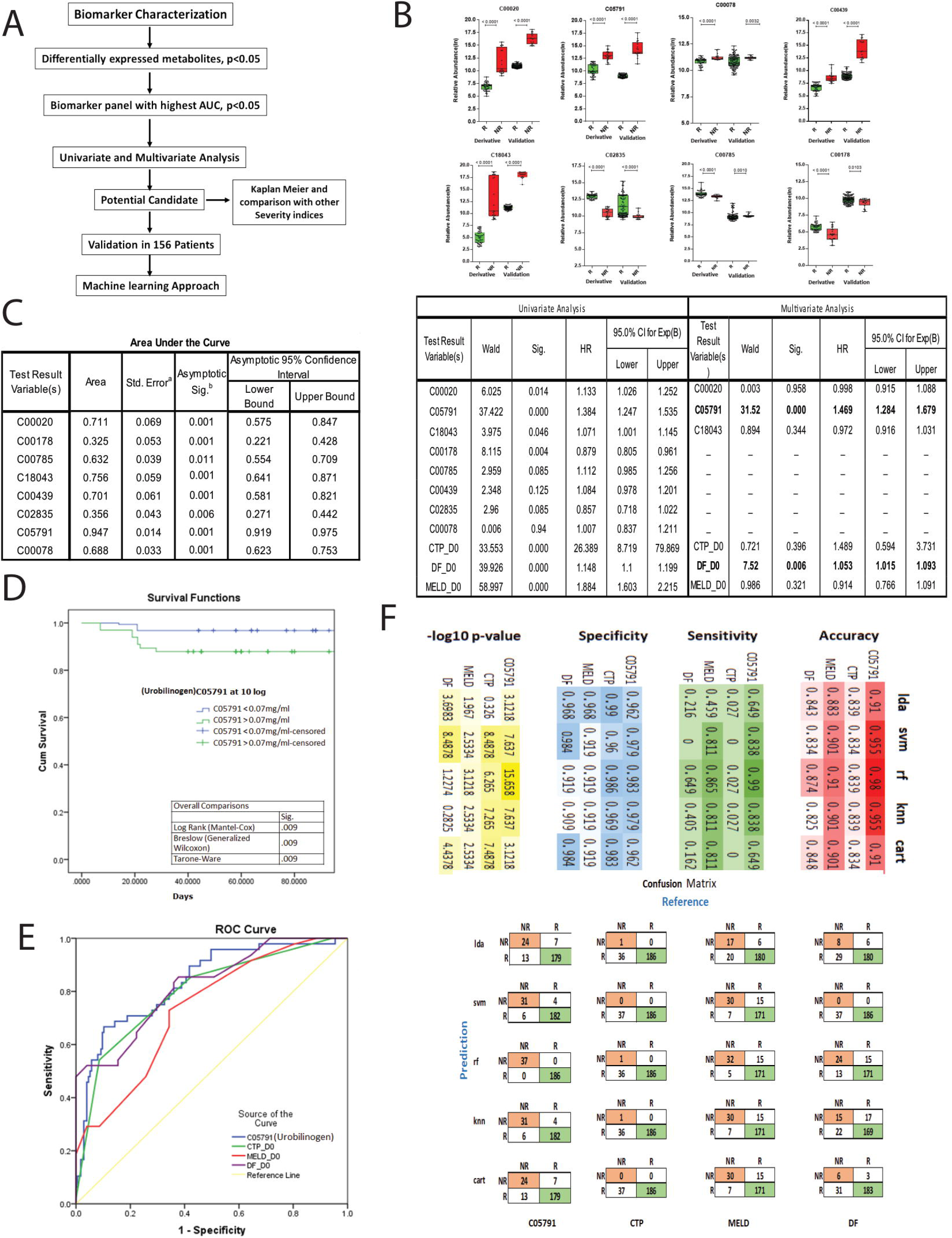
Baseline Plasma Level of Urobilinogen segregates R and NR: **3A.** Flow chart representing approach used to identify metabolic indicators of non-response at baseline and mortality prediction. **3B.** Whisker plot showing expression of top potential metabolites (p-value<0.05, FC±1.5) to predict non-response in derivative cohort and validation cohort at baseline. Metabolites C00020, C05791, C00078, C00439, and C18043 were upregulated and exhibited similar expression in derivative and validation cohorts, whereas C01835, C00785, and C00178 were downregulated in NRs. Red colour depicts NRs, and green colour depicts R. **3C**. Tables showing AUROC and univariate Cox regression analysis of the most significant metabolites capable of predicting non-response against corticosteroids based on their baseline levels respectively. AUC of C05791 (Urobilinogen) was found to be the highest and most significant for the determination of non-response in SAH patients **3D**. Kaplan Meier curve plotted for one-month mortality based on the cut of the level of Urobilinogen. Our results highlight that SAH patients with a plasma level of C05791 more than 0.07mg/ml were highly predisposed to non-response against corticosteroid treatment. Log-rank was found to be significant (p<0.05). **3E**. Comparison of AUROC of Urobilinogen with the severity scores for the prediction of mortality in SAH patients. The AUROC of Urobilinogen was better than CTP, MELD, and DF Score in SAH patients. **3F**. Machine learning-based method establishes that the accuracy, sensitivity, specificity, and p-value of urobilinogen as compared to CTP, DF, and MELD score. The confusion matrix of the random forest was the most appropriate model for segregating responders from non-responders.

**Figure 4:**
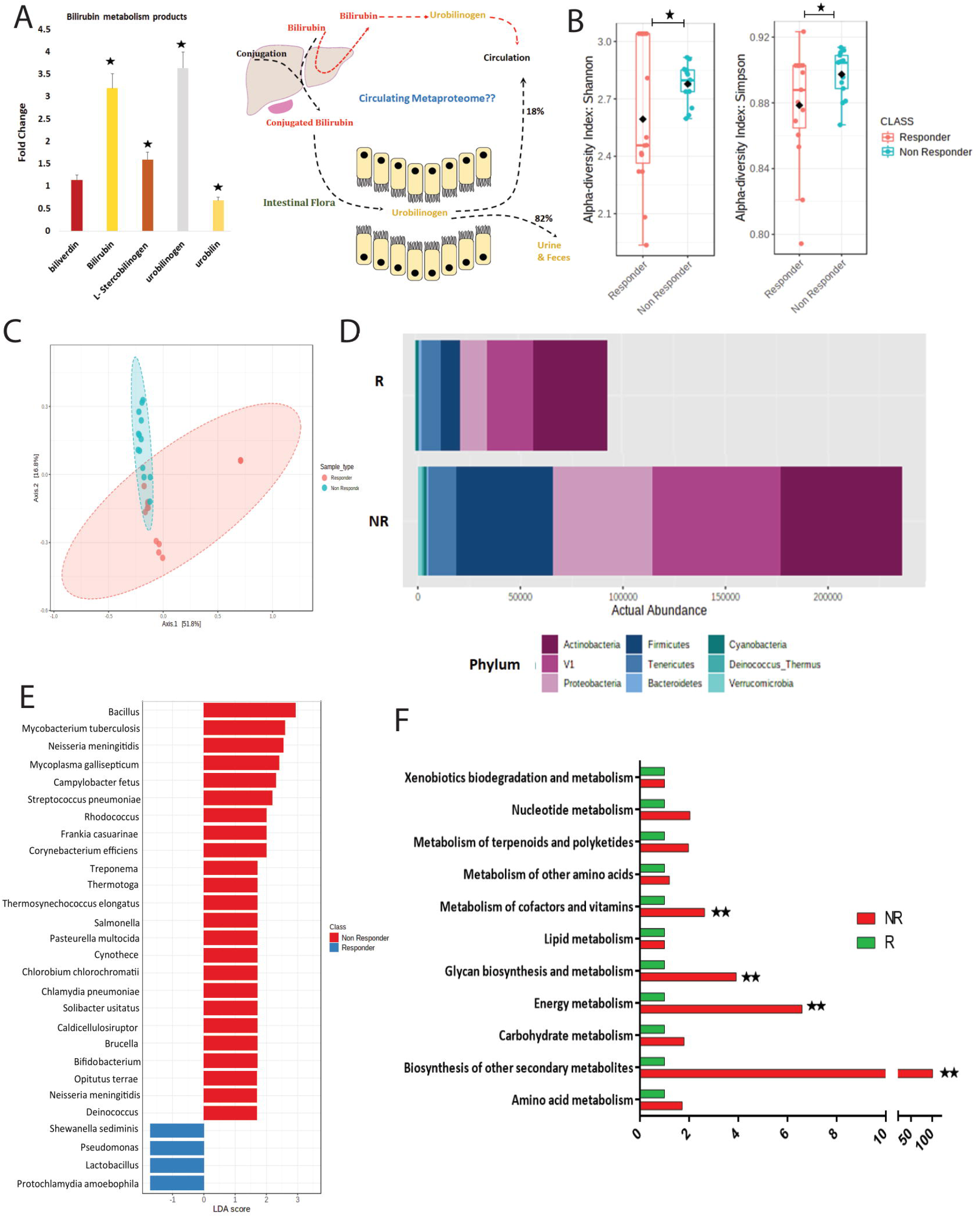
Baseline plasma meta-proteomics (microbiome) reflects high prevalence of bacteria in Non-Responders: **4A.** Bar plot showing significant increase in metabolites (p-value<0.05, FC±1.5) associated with bilirubin metabolism. Another panel depicts bilirubin and urobilinogen metabolism in the body. **4B.** Shannon and Simpson index showing an increase in alpha diversity in plasma of NR. **4C.** Beta diversity plot showing distinct metaproteome composition of NR and R. **4D.** Bar plot showing an increase in prevalence of metaproteome particularly, *Actinobacteria, Firmicutes* and *Proteobacteria* in the plasma of NR. **4E.** LDA (Least discriminant analysis) plot showing bacteria uniquely present in plasma of NR and R. **4F.** Bar plot showing functional assessment of identified metaproteome in R and NR. A significant increase in biosynthesis of secondary metabolites was found in NR (p-value<0.05, FC±1.5).

#### Machine Learning-based validation of urobilinogen as independent predictor of mortality in SAH

223 SAH patients were randomly divided into the training cohort (70% sample strength) and the test cohort (30% sample strength). A total of 5 ML algorithms LDA, KNN, RF, SVM, and CART were used (supplementary method). Four parameters - Accuracy, sensitivity, specificity, and p-value - were calculated for baseline Urobilinogen (C05791), baseline MELD, mDF, and CTP scores for mortality prediction in our study cohort. Twenty ML models were generated across the 4 parameters. Fourfold (outer) nested repeated (five times) tenfold (inner) cross-validation was used to train and test ML models, and the hyper-parameters of each algorithm were optimized. Accuracy and kappa of model development (training cohort) were significant for all the parameters (Supplementary figure-11). The prediction capability of Urobilinogen was found to be highest with accuracy (98%), sensitivity (99%), specificity (98.3%), and p<0.00001 as compared to MELD, mDF, and CTP scores (Figure 3F) validating the utility of urobilinogen as a candidate indicator and Random forest (RF) model as the model of choice for machine learning in SAH patients.

### Baseline plasma meta-proteomics (microbiome) of Non-Responders reflects increase in production of secondary metabolites

Certain intestinal bacteria belonging to *Clostridiales* are known to convert bilirubin to urobilinogen (10). Our data showed a significant increase in bilirubin metabolic by-products in the plasma of NR (p<0.05, Figure 4A). Thus circulating microbiome (metaproteomic) profile of NR was compared to R. The alpha, beta diversity (Shannon, Spimson index; PCoA) and relative abundance of bacteria in plasma of NR were significantly higher and distinct as compared to R (Figure 4B, 4C, and 4D respectively). Particularly, NR showed a significant increase in bacterial peptides associated with Bacillus, Mycobacterium, Neisseria, Mycoplasma, Streptococcus, Salmonella, Chlamydia, Bifidobacterium, and many others (Figure 4E) known to be associated with an increase in inflammation, infection, and severity. Functional enrichment of metaproteome showed a predominant increase in biosynthesis of secondary metabolites, energy, glycan, cofactors, and vitamin metabolism (p<0.05, Figure 4F). Together, these results show that the plasma of NR has higher bacterial diversity corresponding to an increase in bacterial peptides and functions (particularly an increase in secondary metabolite biosynthesis).

### Correlation of plasma metabolome and meta-proteome identifies meta-proteome associated with bacterial derived urobilinogen

As bacteria convert bilirubin to urobilinogen and in order to integrate the metabolome with meta-proteome, baseline differentially expressed metabolome (DEM) was correlated to differentially expressed meta-proteome (DEMp, Figure 5A). Correlation clustering identified 4 clusters showing a strong positive correlation between metabolites and meta-proteomes (Figure 5B; R^1^>0.5, p<0.05). Among these, Cluster 2 was the biggest cluster in which our candidate indicator of NR and mortality (urobilinogen) showed a strong positive correlation (R^1^ >0.5) with meta-proteins associated with bacteria; *Firmicutes* (*Clostridiales* and, Streptococcus pneumonia and Hungatei clostridium), and *Proteobacteria* (Neisseriaceae) (Figure 5C). Expression of these bacterial families and urobilinogen was significantly high in the plasma of NR as compared to R (p<0.05) (Figure 5D). These results clearly suggest that an increase in the plasma urobilinogen in NR could be due to an increase in the level of bilirubin metabolizing bacteria particularly belonging to *Clostridiales*.

**Figure 5.**
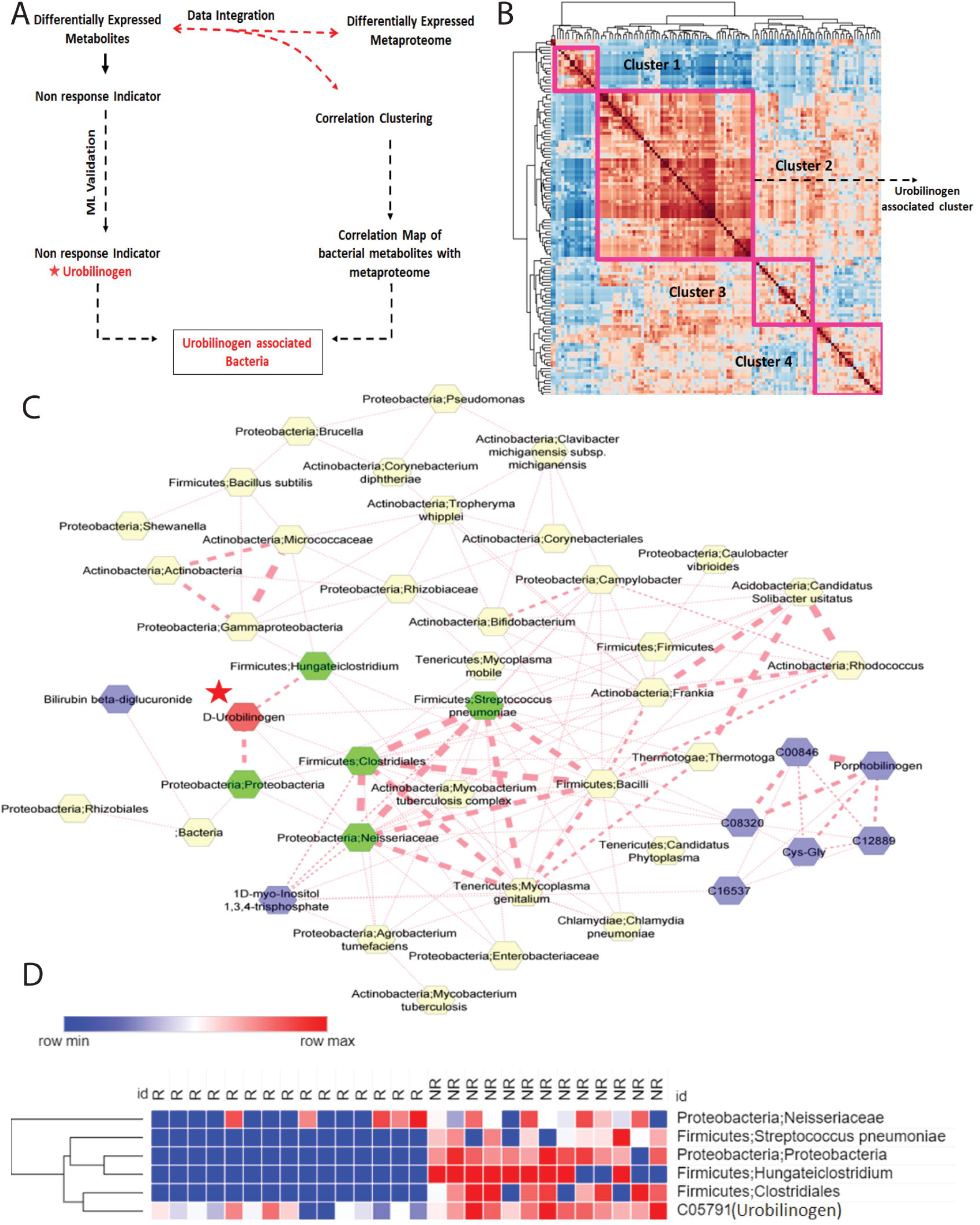
Integration of plasma metabolome and meta-proteome. **5A.** Flow diagram of method adopted to correlate metaproteome and metabolome. **5B.** Correlation clustering of baseline differentially expressed metabolites and metaproteins. Cluster 2 is urobilinogen specific cluster. **5C.** Correlation Network map displaying bacteria correlating with urobilinogen (R^1^ > 0.7). Bacteria belonging to *Firmicutes* and *Proteobacteria* correlated directly with urobilinogen. **5D.** Heatmap showing expression of urobilinogen and bacteria correlated with urobilinogen in NR and R at baseline.

### Urobilinogen activates neutrophils, induces respiratory burst and increases cellular permeability

Data until now clearly showed that plasma level of urobilinogen significantly increased in NR and is an independent predictor of poor outcomes in SAH. Our results also outlined that increased urobilinogen levels correlated directly with bilirubin metabolizing bacteria. We, therefore, speculated that this increase in circulatory urobilinogen may have immune-modulatory properties.

#### Urobilinogen activates neutrophils and induces respiratory burst

To ascertain the immune-modulatory properties of urobilinogen, primary healthy neutrophils (PHN) were exposed to different concentrations (5uM to 120uM) of pure urobilinogen (CAS No. 14684-37-8, MyBioSource) and mitogens like LPS (10ng/ml), PMA (10ng/ml) and H2O2 (0.001%) as control. CD66b+/CD11b+ dual positive neutrophils showed a significant increase in activation, pro-inflammatory and respiratory burst markers such as CXCR1, TNFα, and H2DCFDA expression (MFI) respectively at 60uM and 120uM doses of urobilinogen as seen in LPS, PMA and H2O2 (Figure 6A). 60uM urobilinogen was considered for further experimentation as cell death was observed at 120uM (Figure 6A).

**Figure 6:**
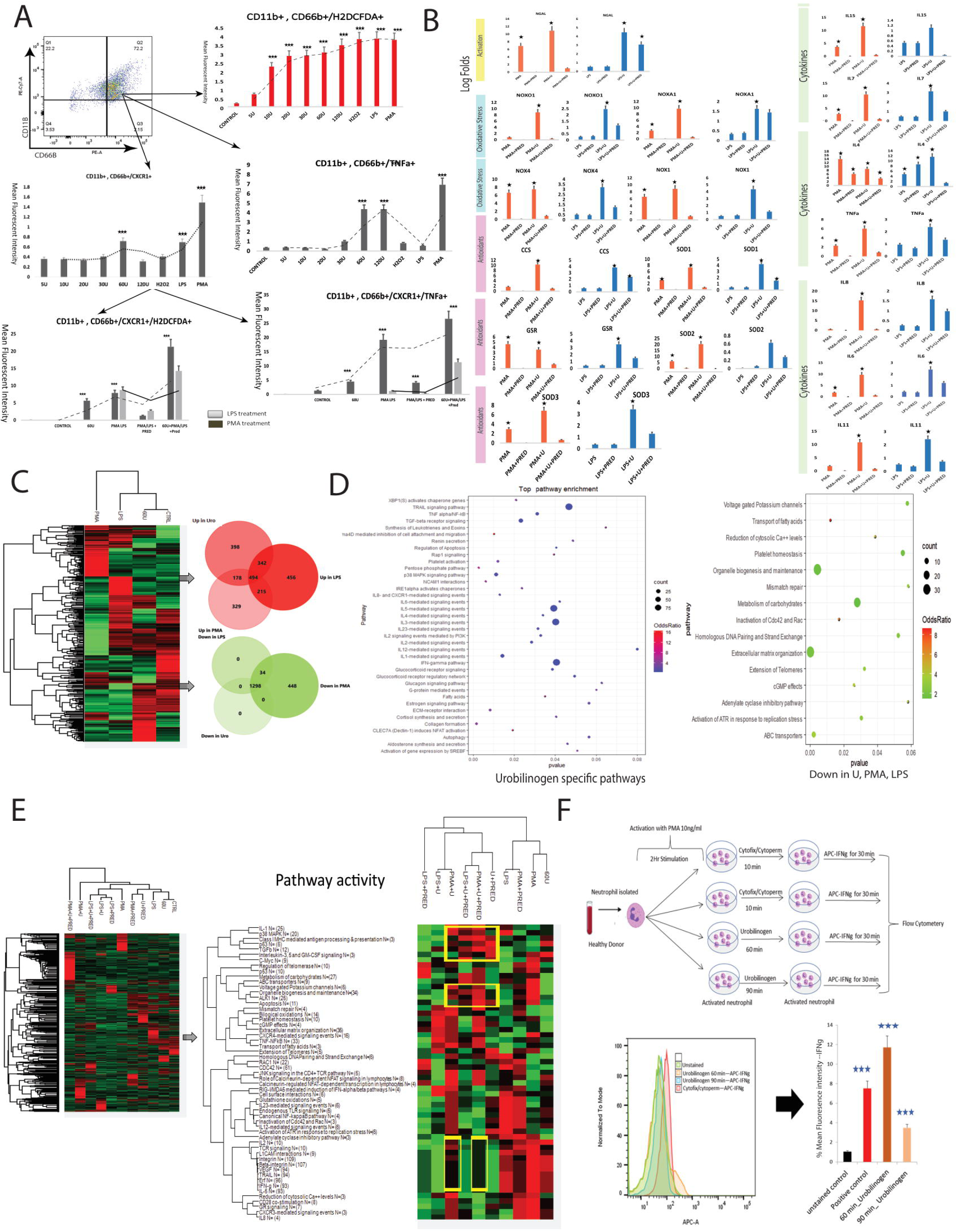
Urobilinogen activates neutrophils, induces respiratory burst and increases cellular permeability: **6A. Panel A)** Representative images of flow cytometry dot plots showing the neutrophil CD11b+ and CD66+ population. **Panel B, C, D)** Graphs representing increase in MFI expression of H2DCFDA (ROS generation), CXCR1+, and TNFa+ in a dose dependent treatment of urobilinogen on neutrophil. Highest MFI for ROS, neutrophil activation and inflammation was observed at 60uM urobilinogen treatment respectively. **Panel E,F)** Graphs representing increase in MFI post PMA/LPS/ Urobilinogen treatment on neutrophils and inefficiency of prednisolone to reduce inflammation (TNFa+ population) and ROS generation (H2DCFDA population) effectively in neutrophils treated with PMA/LPS and urobilinogen. **6B.** Panel showing expression of genes associated with activation of neutrophils, oxidative stress, antioxidants and inflammatory cytokines. Orange colour bar plots show expression of genes in PMA/Urobilinogen/Prednisolone treated PHN whereas blue bar plots shows expression of genes in LPS/Urobilinogen/Prednisolone treated PHN. Star mark highlights significant change in expression. **6C.** Heatmap shows expression of total enriched proteins in PMA, LPS and urobilinogen treated PHN. Red colour venn diagram represents the proteins which got upregulated post urobilinogen/PMA or LPS treatment respectively (p-value<0.05, FC±1.5). Green colour venn diagram represents the proteins which got upregulated post urobilinogen/PMA or LPS treatment respectively (p-value<0.05, FC±1.5). **6D.** Dot plot represents the pathways got significantly (p-value<0.05, FC>1.5) upregulated in PHN upon urobilinogen treatment alone and got downregulated (p-value<0.05, FC<-1.5) in PHN upon PMA/LPS/urobilinogen treatment. **6E.** Heatmap represents expression of proteins enriched globally in PHN treated under different conditions highlighted in the figure. Panel 1 shows the pathway activity of different pathways in different groups as mentioned in the figure. Numerical values in the bracket highlights the number of proteins enriched in each pathways. **6F.** Representative histogram and bar plot of IFNg-MFI measured post urobilinogen treatment on neutrophils to assess the permeability capacity of urobilinogen. Bar plot shows high MFI in urobilinogen treated samples compared to positive controls treated with cytofix and cytoperm.

Next, PHN treated with 60uM urobilinogen, PMA/LPS (positive controls), and in combination (PMA/LPS + 60uM-urobilinogen) were challenged with 10uM Prednisolone. PMA/LPS activated neutrophils (CD66b+/CD11b+/CXCR1+) showed a significant reduction in oxidative stress and inflammatory cytokine production post prednisolone treatment but in presence of urobilinogen (60uM) prednisolone couldn’t effectively reduce the inflammation suggesting that urobilinogen interferes with the activity of prednisolone (Figure 6A lower panels).

This observation was further validated using qRT-PCR. The expression of genes linked to neutrophil activation, oxidative stress, antioxidant response, and pro-inflammatory cytokines was overlooked in PHN treated with PMA/LPS and PMA/LPS+60uM-urobilinogen+prednisolone. On comparing PMA/LPS with PMA/LPS+60uM urobilinogen, we observed a significant increase in expression of neutrophil activation (NGAL), oxidative stress (NOXO1, NOXA1, NOX1, and NOX4), and release of pro-inflammatory cytokines (IL15, IL7, TNFα, IL6, IL8, and IL11) genes suggesting pro-inflammatory activity of urobilinogen (p<0.05). Similar to the previous findings (Flow Cytometry data), in presence of urobilinogen (60uM), prednisolone treatment was not able to significantly reduce the expression of neutrophil activation (NGAL), oxidative stress (NOXO1, NOXA1, and NOX4) and release of pro-inflammatory cytokines (IL7, IL8 and TNFα) again validating the role of urobilinogen in the interference of prednisolone functioning (p<0.05, Figure 6B).

To further validate the pro-inflammatory nature of urobilinogen, treated PHN (urobilinogen, PMA, LPS) were subjected to proteomics (Figure 6C). Urobilinogen specifically increased the expression of 398 proteins (Figure 6C) associated with inflammation (TNF/NF-kB, TGFb, p38MAPK, ILs, and IFNg), apoptosis (TRAIL), and other pathways (Figure 6D). Proteins commonly increased by urobilinogen, LPS, and PMA (178+494+341) proteins (Figure 6C) were also linked to pathways TRAIL, TNF, RAC1, p38 MAPK, mTOR, ILs, and others suggesting that urobilinogen shares inflammatory properties similar to LPS and PMA (Supplementary figure 14). Urobilinogen specifically did not down-regulate any proteins though proteins downregulated by urobilinogen, LPS, and PMA were linked to transport of fatty acids, organelle biogenesis, and maintenance, metabolism of carbohydrates, extracellular matrix organization, and other pathways (Figure 6D).

Next, to validate that urobilinogen modulates prednisolone functioning PHN treated with 60uM urobilinogen, PMA, LPS, and PMA/LPS + urobilinogen in combination and then challenged with Prednisolone were subjected to proteomics. Pathway activity (calculated based on the expression of genes belonging to a specific pathway - detailed in supplementary method) of differentially expressed proteins showed a significant increase in IFN-g, IL-5, ERF, TRAIL, VEGF, integrin, and B integrin known inflammatory pathways when treated with urobilinogen/PMA/LPS (Figure 6E). Interestingly, in presence of urobilinogen, prednisolone was not able to significantly reduce the expression of IL-1, P38-MAPK, TGF-B, IL-3, and 5, and GMCSF signalling, p63, C-MYC, biological oxidation, and apoptosis again validating the role of urobilinogen in hindering prednisolone functionality.

#### Urobilinogen increases cellular permeability

Our results validated that urobilinogen has pro-inflammatory nature and inflammation is more pronounced when urobilinogen is given along with LPS/PMA. Thus, we hypothesized that urobilinogen somehow increased the bioavailability of PMA/LPS. To validate, a permeability assay on PHN was performed (Figure 6F, further detailed in supplementary methods). We observed a significant increase in intracellular staining (MFI) of IFNg in PHN treated with urobilinogen alone as compared to cytofix/cytoperm-treated PHN suggesting that urobilinogen increased the cellular permeability which resulted in uptake of IFNg antibody (p<0.05, Figure 6F). This provided the first evidence that urobilinogen not only induces inflammation but also promotes cellular permeability leading to intracellular staining of IFNg Taken together, our results provide evidence and validate that urobilinogen is a pro-inflammatory molecule, modulates cell permeability, and also interferes with the functioning of prednisolone.

### Urobilinogen modulates glucocorticoid receptor expression and impairs gut integrity

#### Urobilinogen induces imbalance in GRa/b expression and hampers prednisolone activity

Glucocorticoids (prednisolone) by binding to GRα receptor are known to trans-activate and trans-repress certain genes to control inflammation. As already known, an increase in GRß expression inhibits the transactivation and trans-repression processes by a dominant negative feedback loop (Figure 7A). Our results showed that inflammation could not be suppressed after prednisolone treatment in PMA/LPS+U stimulated PHN effectively, thus we hypothesized that urobilinogen interferes with glucocorticoid receptor signalling. Therefore, we investigated the expression of two major isoforms of glucocorticoid receptors; GRα and GRß receptors using the DIA-MSMS approach. Analysis revealed that the expression of GRα got reduced and the expression of GRb got enhanced following urobilinogen treatment on neutrophils (Figure 7B). We then further checked the expression status of trans-activated and trans-repressed genes downstream of the glucocorticoid receptor. Targeted proteomics data revealed that the expression of genes known to get repressed such as NFkB, MAPK-MAP, IRF (Interferon regulatory factor) and CREB post prednisolone administration remained up-stimulated (Figure 7C) whereas the expression of trans-active genes such as an inhibitor of NFkB (IkB) remained suppressed (Figure 7C). To further validate, we enriched GR-associated genes (n=1590) using the Enrichr TRANSFAC library. Of 1590, our proteomic analysis identified 270 proteins. Expression-based pathway activity of these 270 proteins showed that urobilinogen enhanced inflammation (IL6, TNF, TGFb, p38 MAPK, IL1, IFNg, mTOR), apoptosis (RAC1), and other pathways and the inhibitory capacity of prednisolone for the pathways such as IFNg, mTOR signalling and cytokine signalling was found to be lower in presence of urobilinogen than that in absence of urobilinogen (as highlighted in the yellow box; expression of IFNg got significantly reduced when LPS was treated with prednisolone (red to green) but in presence of urobilinogen the expression remained high (red to red-black) respectively) (Figure 7D).

**Figure 7:**
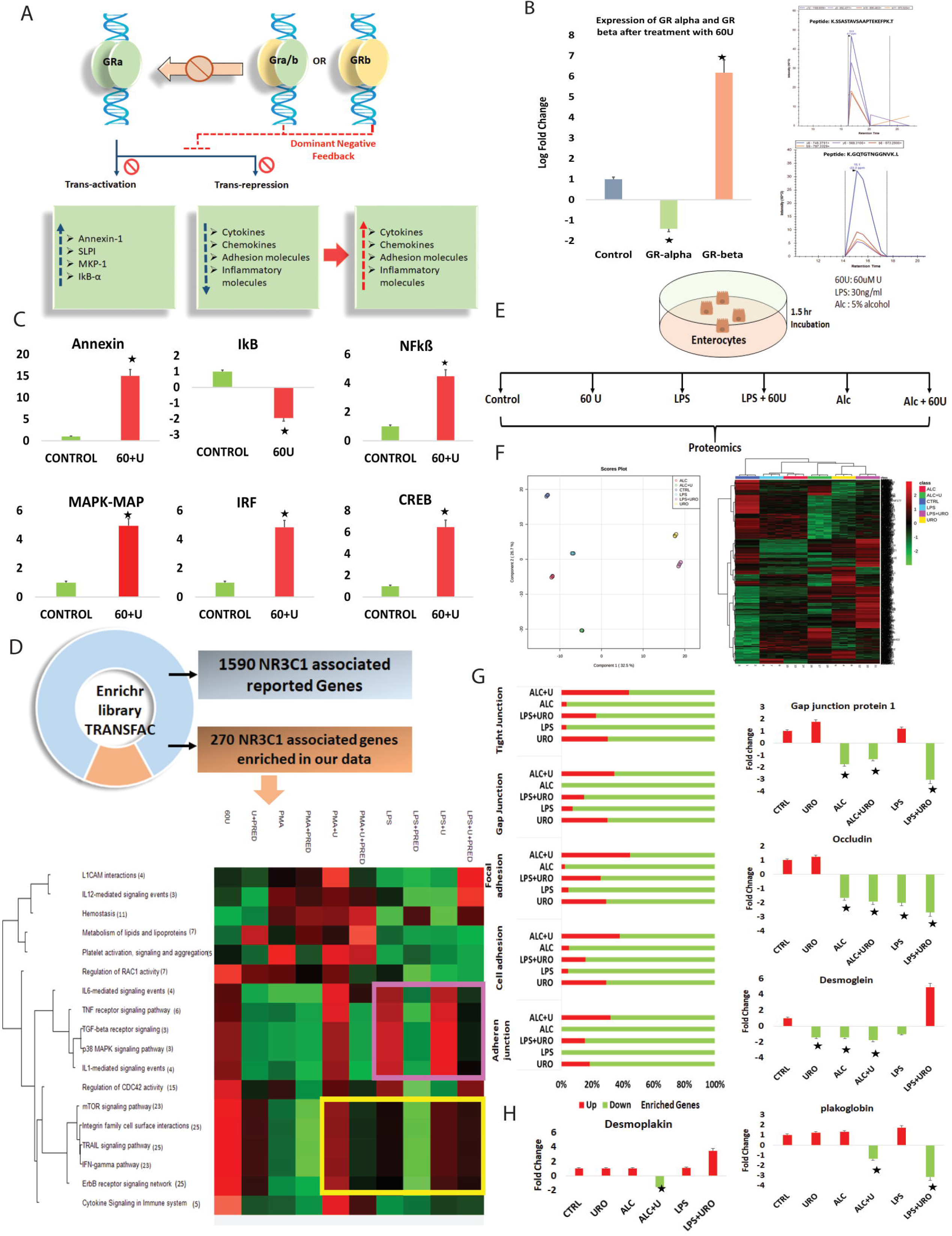
Urobilinogen modulates glucocorticoid receptor expression and impairs gut integrity. **7A.** Figure representing the functional role of GRα and GRβ receptor and its mechanism of regulation. Increase in GRβ expression dominantly repress the GRα. **7B.** Bar plot showing expression of GRα and GRβ in PHN treated with 60uM urobilinogen (p-value<0.05, FC>1.5). **7C.** Bar plot showing expression of molecules present in GRα downstream signalling. Y axis represents the fold change. **7D.** Pie chart here represents 270 genes enriched in our data out of 1590 NR3C1 associated reported genes. Heatmap represents the pathway activity of pathways associated with the enriched genes. Numerical values in the bracket represents number of genes enriched in the respective pathway. Pink and yellow box highlights the increase in inflammatory pathways upon PMA/LPS and urobilinogen administration and inefficiency of prednisolone to effectively downregulate inflammatory pathways. **7E.** Pictorial representation of permeability assay performed on enterocytes to assess the effect of urobilinogen on membrane integrity. **7F.** PLS-DA plot showing variability in the enterocytes treated with Urobilinogen, LPS, and alcohol. Heatmap represents the expression of proteins under different treatment conditions and highlights that urobilinogen has almost similar effects as LPS has on enterocytes. **7G.** Bar plot showing change in expression (p-value<0.05, FC>1.5) of proteins associated with Tight junction, gap junction, focal adhesion, cell adhesion, and adherent junction. Red colour represents the percent of proteins significantly upregulated post 60uM urobilinogen treatment and green colour represents the percent of proteins significantly downregulated post 60uM urobilinogen treatment. **7H.** Bar plot shows the expression of desmoplakin, gap junction protein 1, occludin, desmoglein, and plakoglobin post treatment with urobilinogen, LPS and alcohol. Green bar represents downregulation and red represents upregulation.

#### Urobilinogen modulates junction proteins and contributes to leaky gut

Our results showed that urobilinogen modulates cell permeability. Further urobilinogen is produced in the gut by bacterial action on bilirubin. An increase in urobilinogen in the gut could modulate gut integrity. To check, isolated Primary mouse enterocytes (PME) were treated with urobilinogen, LPS (30ng/ml), alcohol (5%), LPS+ 60uM U, and alcohol + 60uM U and subjected to proteomics (Figure 7E). PLS-DA and hierarchical clustering show that the proteomic expression in enterocytes under urobilinogen, LPS+urobiliniogen, and alcohol+urobilinogen was similar, whereas, proteomic expression of alcohol and LPS were similar (Figure 7F). Detailed proteomics analysis showed that the majority of proteins associated with tight junction, gap junction, adherent junction, focal adhesion, and cell adhesion were downregulated in comparison to controls (p<0.05, Figure 7G). Expression of proteins associated with the membrane integrity such as gap junction protein 1, occludin, desmoglein, plakoglobin, and desmoplakin got reduced when enterocytes were treated with both alcohol/LPS and urobilinogen suggesting a role of urobilinogen in modulating junction proteins and gut integrity (Figure 7H). Together, these results show the direct role of urobilinogen in modulating glucocorticoid receptor expression, junction proteins, and gut integrity resulting in inflammation inhibition failure and leaky gut respectively in such patients.

## DISCUSSION

In this pilot study, in order to identify the metabolic indicators capable of predicting non-responsiveness (baseline) against corticosteroid therapy in SAH patients, plasma metabolomics using high-resolution mass spectrometry-based (LC-MS) was performed. It is believed that SAH patients at baseline are similar, as also seen in our clinical parameters; but physiologically they appear to be different (WMCNA/heatmap). Therefore, metabolome provides a better insight into responders and non-responders at baseline. Results of our study show that plasma levels of urobilinogen, a major urobilinoid produced in the gut by bacterial action on bilirubin, is capable of segregating NR (independently) and also documented significant association with early mortality in SAH patients. Since urobilinogen is a bacterial metabolized product and was significantly high in circulation, we performed metaproteomics of baseline plasma samples to know bacterial prevalence in NR and found that the Phylum: *Actinobacteria, Proteobacteria* and *Firmicutes,* and class *Clostridiales* associated with the metabolism of urobilinogen were high in the circulation as well. Further, investigations suggested that increased plasma levels of urobilinogen not only activated neutrophils and induced the expression of inflammatory genes but also modulated the expression of glucocorticoid receptors thereby contributing to an increase in inflammation leading to a non-responsive phenotype in SAH. We also showed that urobilinogen contributed to an increase in membrane permeability in neutrophils and enterocytes by significantly reducing the expression of junction proteins and contributed to leaky gut phenotype in SAH patients. Thus early identification and clearance of urobilinogen could be one of the therapeutic strategies to prevent corticosteroid NR in SAH patients.

A total of 223 patients were enrolled in the study, with 70 patients in the derivative group and 153 in the validation group. At baseline, apart from the increase in bilirubin metabolic product, there was a significant increase in plasma metabolic products of amino acid metabolism, i.e. proteolysis, and fatty acid metabolism, i.e., lipolysis. Both proteolysis and lipolysis are known to be used as fuel by immune cells (11). Our results showed a significant increase in such catabolic processes in the NR at baseline suggesting a relatively more active immune phenotype compared to R. Previous studies have shown the accumulation of cholesterol sulfate (C18043) in the plasma of patients with liver disease. Cholesterol sulfate stabilizes the cell membrane to protect the cell from osmotic lysis (13). A higher accumulation of cholesterol sulfate was seen in the plasma of NR, stipulating more severe cell damage in NR. Further, a higher amount of ATP is produced from the TCA cycle than glucose oxidation (14), and downregulation of the TCA cycle and accumulation of AMP (C00020) (15) in NR reflects an energy-deprived microenvironment. This evidence provides insights into that plasma metabolome can segregate patients who are unlikely to respond to corticosteroid therapy.

The effect of prednisolone therapy was evaluated by studying temporal changes in the metabolic profile of R and NR. Our results showed that the prednisolone administration increases the metabolic activity of R throughout the treatment unlike in NR. In R there was an increase in energy metabolism, amino acid synthesis and metabolism, fatty acid metabolism, and vitamin metabolism highlighting functional improvement over time. This was validated using Weighted Metabolite Correlation Network Analysis (WMCNA). The WMCNA in an unsupervised manner, groups the metabolites in the modules sharing similarities in the biological pathways. WMCNA validated inactive metabolic state in NR post corticosteroid treatment. Modular analysis showed that NR has a baseline increase in steroid hormone biosynthesis and vitamin B6 metabolism. Baseline higher level of steroid hormone biosynthesis is one such prominent cause that could lead to feedback inhibition of the corticosteroid given to the patients leading to non-response (16). In addition, pathways such as citrate cycle (TCA cycle), arginine biosynthesis, glutathione metabolism, tyrosine metabolism, glycolysis/gluconeogenesis, and others were stimulated only in R. This observation suggests that corticosteroid exposure specifically induces energy metabolism, arginine metabolism, antioxidant response, and amino acid metabolism in R, which is directly associated with response. WMCNA Modules documented significant correlation with the severity such as baseline NR-specific module ‘RED’ (increased in NRs) showed direct correlation whereas module ‘PINK’ (increased in the Rs) showed an inverse correlation with the severity indices (MELD, CTP, DF, and others) respectively. Together the analysis validates the notion that Corticosteroid therapy is effective only in one subset of patients.

Based on the AUROC, p-value, FC, and mean decrease in accuracy, a panel of 8 metabolites that could segregate NR from R at baseline was identified. On validation in a separate cohort of 156 patients, similarities in the expression pattern of the eight metabolites were observed, thereby validating our results. C05791 (Urobilinogen) augmented the highest AUC of 0.94 and significantly predicted non-response against corticosteroid therapy and mortality with hazard-ratio of 1.5(1.1-1.6). Plasma urobilinogen at 0.07mg/ml cut-off reliably segregated non-survivors (p-value<0.01, log-rank test). In addition, validation by machine learning approach demonstrated the accuracy (98%), sensitivity (99%), and specificity (98.3%) of C05791 (Urobilinogen) as a candidate indicator (non-response and mortality) and Random forest (RF) model as the model of choice for machine learning in SAH patients.

As already known that urobilinogen, a bile pigment, is formed by bacterial action on conjugated bilirubin in the intestine (10). Knowing that it is a bacterial product and other bilirubin metabolized products were also high in the plasma of the NR, we looked down into the plasma metaproteomic profile of NR and R. Alpha and beta diversity were significantly high in NR and the level of urobilinogen correlated directly with the bilirubin and urobilinogen converting bacteria Bacteroides fragilis, Clostridium ramosum, Clostridium perfringens, and Clostridium difficile.

The physiological roles of these urobilinoids are not clear. It has been shown in one study that bilirubin has anti-oxidative activity, reduces adiposity and fatty liver (17). On the contrary, a study on stercobilin suggested that it not only has pro-inflammatory activity but also alters lipid metabolism in obesity/ diabetes mellitus (18) (10). Our study demonstrated that urobilinogen not only induces inflammation in the neutrophils but also alters the expression of glucocorticoid-alpha receptor diverging response for steroids toward non-response.

Prednisolone is known to reduce inflammation but when neutrophils were tweaked with PMA/LPS, urobilinogen, and prednisolone, prednisolone couldn’t effectively reduce the inflammation. This gave the clue that urobilinogen must be interfering with glucocorticoid signaling. To further introspect, we checked the expression of GRα and GRβ in neutrophils treated with urobilinogen. When compared to control, expression of GRα (essential for prednisolone response) got reduced post urobilinogen stimulation whereas the expression of GRβ (associated with dominant feedback inhibition of GRα response (19) got enhanced post-treatment with urobilinogen. This suggests that high and persistent levels of urobilinogen in the plasma of SAH patients may drive them to have severe inflammation and develop resistance to corticosteroid therapy.

Our data also showed that when neutrophils were treated in combination with both PMA/LPS and urobilinogen, inflammation increased to several folds. This suggested that maybe urobilinogen increases the bioavailability of inflammatory molecules and increases inflammation. Results of this study clearly demonstrated that urobilinogen modulates cellular permeability thereby inducing the labelling of the intracellular cytokine IFNg. These results provide the first evidence that overexposure to urobilinogen at a higher concentration is capable of modulating cellular permeability.

Since urobilinogen is produced in large amounts in the gut (10), intestinal cells are persistently exposed to a higher concentration of urobilinogen. This then may affect the membrane stability and contributes to leaky gut in such patients. Urobilinogen exposure at a higher concentration on the primary mice enterocytes showed a significant decrease in the tight junction, gap junction, adherent junction, focal adhesion, and cell adhesion proteins (p<0.05) specifically the expression of proteins associated with the membrane integrity such as gap junction protein 1, occludin, desmoglein, plakoglobin, and desmoplakin got reduced on treatment with urobilinogen in combination with alcohol/LPS suggesting a role of urobilinogen in the modulation of junction proteins and gut integrity.

In conclusion, in non-responders, a change in microbiome diversity; particularly an increase of bilirubin metabolizing microbiome (*Clostridiales*), converts the bilirubin to urobilinogen, in the gut. Increased gut urobilinogen levels, start a vicious cycle of dysbiosis by acting on the intestinal cells (enterocytes); impacting their cell membrane integrity by dysregulating tight junction, gap junction, adherent junction, focal adhesion, and cell adhesion proteins and promoting leaky gut. Increased dysbiosis (bilirubin metabolizing bacteria) contributes to the increase in circulatory urobilinogen level (increased bilirubin metabolism) which induces immune flare by activating neutrophils. Circulating urobilinogen also dysregulates the glucocorticoid receptors (increases GRß and decreases GRα) thereby contributing to non-response against corticosteroid therapy in patients with SAH (Figure 8).

**Figure 8:**
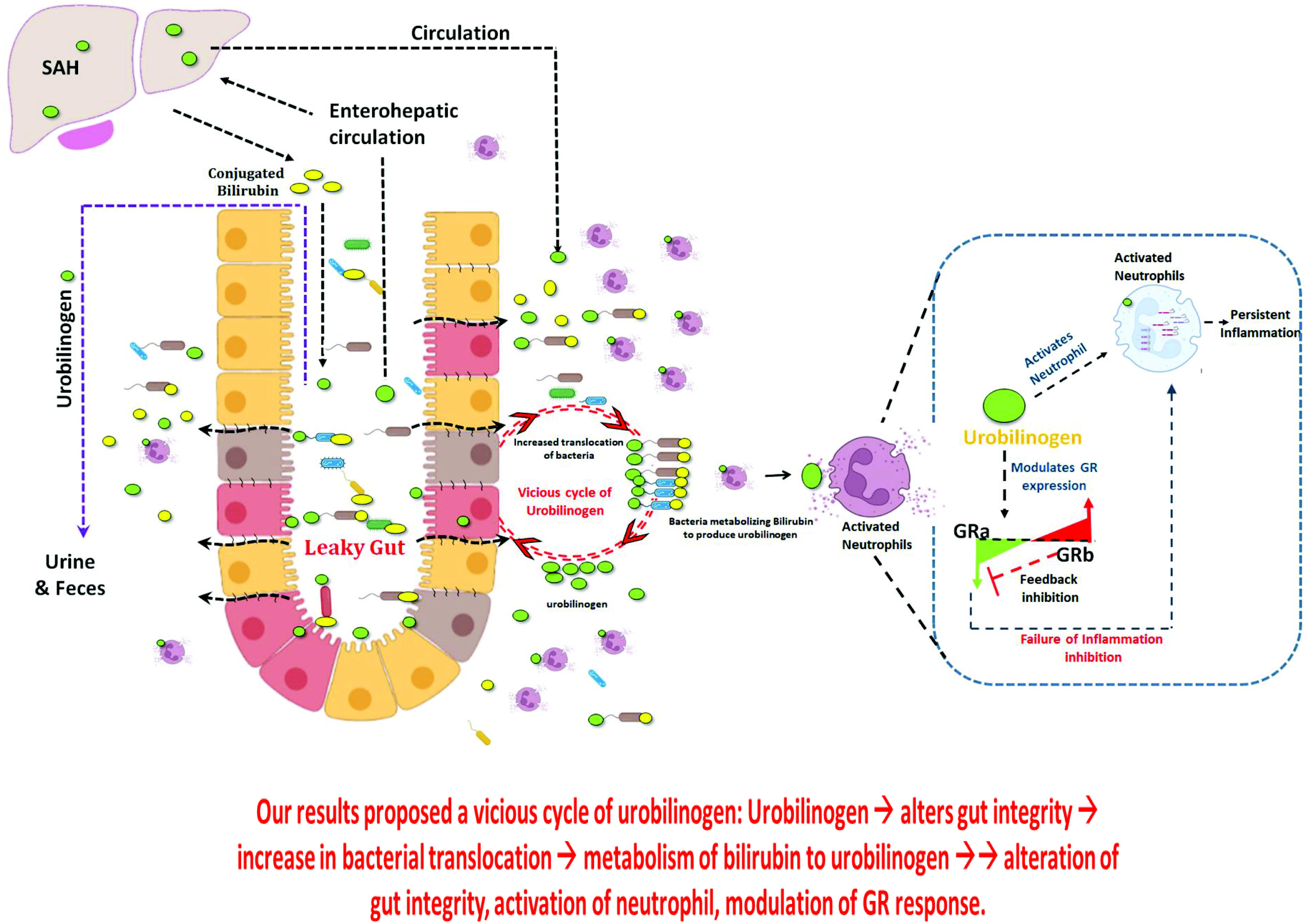
Graphical Abstract. Our study highlights that severe alcoholic hepatitis particularly among Non-responders has a high circulating bilirubin and its metabolic product urobilinogen. We also found an increase in bacteria specifically bacteria metabolizing bilirubin to urobilinogen in plasma of SAH-NR. This study highlights an increase in circulating urobilinogen in NR modulates: 1) gut integrity by downregulating junction proteins in the enterocytes. 2) Activates circulating neutrophils and induces flare of inflammation 3) Increases the expression of GRß which further inhibits GRα expression and dependent response. Urobilinogen inhibited trans-repression (inflammatory genes) and transactivation (anti-inflammatory genes) of genes under GRα thereby generating resistance against corticosteroid therapy seen in SAH patients. Finally, our results proposed a vicious cycle of urobilinogen: Urobilinogen ➔ alters gut integrity ➔ increase in bacterial translocation ➔ metabolism of bilirubin to urobilinogen ➔➔ alteration of gut integrity, activation of neutrophil, modulation of GR response.

This study has the limitation of being monocentric. Future multicentre studies should be performed with metabolomics and meta-proteomics in large series of patients with SAH.

To conclude, baseline plasma metabolome and meta-proteome discriminate corticosteroid NRs from Rs. Further, bacterial-derived urobilinogen induces inflammation, modulates intestinal permeability, and predicts outcomes in Severe Alcoholic Hepatitis.

## Data Availability

Data associated to the manuscript could be obtained from Dr jaswinder singh maras on request

## Abbreviations

SAH: Severe Alcoholic Hepatitis
R: Responders
NR: Non-responders
DF: Discriminant Factor
DEMs: Differentially Expressed Metabolites
GR: Glucocorticoid Receptor
H2DCFDA: 1’,7’-dichlorodihydrofluorescein diacetate
MFI: Mean fluorescence intensity
ML: Machine Learning
MS: Mass Spectrometry
PLS-DA: Partial Least Square Discriminant Analysis
PHN: Primary Human Neutrophils
PME: Primary Mice Enterocytes
Pred: Prednisolone
U: Urobilinogen
WMCNA: Weighted Metabolome Correlation Network Analysis

