## supplementary method for "Plasma multi-omics outlines association of urobilinogen with corticosteroid non-response, inflammation and leaky gut in Sever Alcoholic Hepatitis"

**Supplementary Methods**

**Patients**

The laboratory personnel were blinded to clinical or laboratory details of the patients. The patients were treated as per the standard treatment guidelines including intensive care monitoring, intravenous albumin, broad-spectrum antibiotics, and a high-calorie diet (35‐40 call/kg/day). mDF, MELD, and Child-Pugh scores were used to assess the severity at the initial presentation.

**Untargeted Metabolomics using UHPLC-HRMS**

Untargeted metabolomics was performed in plasma samples of derivative cohort, then validated in the validation cohort using Ultra high-pressure liquid chromatography (UHPLC) coupled to high-resolution mass spectrometry (HRMS). Early morning plasma samples were collected and were stored at –80°C. The metabolic extracts were enriched from 100µl of plasma samples following the organic phase extraction method (1). Sample and methanol were mixed in 1:4 ratios and were kept at –20°C overnight to precipitate down the protein content in the samples. The supernatant was collected after a high spin at 13000rpm for 10 minutes. The supernatant was vacuum dried followed by reconstitution in 5% acetonitrile: 95% water spiked with known concentration of internal and external standards. Samples were subjected to reverse‐phase chromatography on an ultra‐high performance liquid chromatographic system followed by high‐resolution mass spectrometry (HRMS) in the C18 column as detailed in the our recent publication (2)

**Construction of Weighted Metabolome Co-Expression Network (WMCNA):**

The Weighted Metabolome co-expression network analysis (WMCNA) was performed using Perseus software integrated with the WGCNA R package. Pre-processed metabolome data was log normalized followed by z-scoring. Finally, WMCNA (Network type - signed; Correlation function – bicor and Power- 9 was used as soft-threshold) was performed in 6 groups (R at day0, day3, and day7; NR at day0, day3, and day7) followed by hierarchical clustering. As a result, 712 metabolites were clustered into 9 modules (clusters of metabolites) using an average linkage hierarchical clustering algorithm (3).

**Construction of Module Trait Relationship:**

WMCNA was constructed by correlation of phenotypic traits (R at day0, day3, and day7; NR at day0, day3, and day7 are referred as traits) with modules (clustered metabolites) in each group of samples to establish module trait relationship heatmap (4).

**Correlation Analysis:**

Correlation analysis between each module and clinical parameters at day0, day3 and day7 was performed using R.

Differentially expressed metabolites at baseline were correlated with meta-proteins using Spearman correlation algorithm (R2>0.5, p<0.05). Correlation network between DEMs and meta-proteins was built using MetScape version 3.0.

**Machine Learning Validation:**

The Machine learning approach was employed using five different ML (Linear discriminant analysis; LDA, Random forest; RF, Classification and Regression Trees; CART, Support Vector Machine; SVM, K-Nearest Neighbor; KNN) algorithms to validate and compare the sensitivity and specificity of candidate indicators with the clinical variables (severity indices). In total, we implemented 20 ML models comprising of 5 ML algorithms along with 4 parameters (used for multivariate COX regression analysis). Fourfold (outer) nested repeated (five times) tenfold (inner) cross-validation (with randomized stratified splitting) was done on training and test cohort in R with the Caret package. In this way, repeated tenfold cross-validation was performed 20 times, and the models obtained the best results. Besides, the overall cross-validation prediction performance was summarized by the accuracy, sensitivity, and specificity performance measures. The equations used to quantify these performance measures are presented below (in which TP represents true positives, TN represents true negatives, FP represents false positives, and FN represents false negatives):

Accuracy=TP+TNTP+TN+FP+FN

Sensitivity=TPTP+FN

Specificity=TNTN+FP

**Untargeted Metaproteomics using Nano-LCMS**

The metaproteome study was performed as detailed in our published paper (ref). In brief, low abundant protein fraction was collected after depleting albumin from plasma samples of SAH patients. Isolated proteins were then reduced and alkylated, followed by trypsin digestion. Digested proteins were purified using C18 spin columns and subjected to LC-MS as mentioned in the protocol paper (5, 6). Acquired MS/MS data was analysed using Proteome Discoverer (version 2.0, Thermo Fisher Scientific, Waltham, MA, United States) with database of the bacterial sequences (UniprotSwP_20170609, with sequences 467231 and MG_BG_UPSP with sequences 2019194). This was further validated using Mascot algorithm (Mascot 2.4, Matrix Science). Significant peptide groups were enriched at (p-value <0.05) and q values (p-value <0.05) and false discovery rate of 0.01. Only peptides with rank-1and Peptide Sequence Match (PSM)>3 were subjected to biodiversity and functional analysis on unipept. Enriched bacterial species were subjected to statistical, functional and biodiversity analysis.

**Metabolome-metaproteome Integration Analysis:** Differentially expressed metabolites at baseline were correlated with baseline metaproteome to understand the host microbiome interactions in SAH. Metaboanalyst version 5.0 was used for correlation clustering between metabolome and metaproteome. Cytoscape-MetaScape was used to establish the correlation network map between members of identified clusters.

**Neutrophil Activation and Oxidative Burst Assessment:**

**Primary Healthy Neutrophil (PHN) isolation:** Peripheral blood was obtained from the healthy individuals and the neutrophils were isolated using Polymorphprep™ (Progen Cat. No. 1895). 10ml of anticoagulated blood was carefully layered on 10mL of Polymorphprep™ without disturbing the Polymorphprep™ media. The sample medium was then centrifuged at 550g for 30” without brakes. After centrifugation, cells separated into two distinct layers. The upper layer contained mononuclear cells whereas the lower band contained polymorphonuclear cells. The PMNs were collected and washed with PBS. Finally culture media RPMI with 10% FBS media was added to the PMNs (6).

**Neutrophil viability:** Neutrophil viability was calculated using trypan blue staining method. Isolated neutrophils were stained with trypan blue and were observed under microscope. Live cells remained unstained whereas dead cells got stained with trypan stain. 99% of the neutrophil population obtained were live.

**Neutrophil Activation:** ~10^6^ PHN (>98% pure) were first treated with different concentrations of urobilinogen (5, 10, 20, 30, 60, and 120uM) for 3 hr at 37^0^ C and 5% CO_2_ . LPS (10ng/ml), PMA (10ng/ml) and H2O2 (0.002%) were taken as positive control. After 3 hr of treatment, cells were collected and washed with PBS.

**Flow Cytometry Analysis:** Neutrophils were characterized following the treatment of cells using CD66b+PE (Clone G10F5, PE, BioLegend) and anti-CD11b+PECy7 (Clone ICRF44, PE, BioLegend) (neutrophil markers), and anti- CXCR1-BV510, and anti-TNFa-PerCPCy5.5 (neutrophil activation markers). The stained cells were acquired using BD FACS ARIA III flow cytometer, and the recorded data was analysed using FlowJo software version 10.

**Oxidative Burst Assessment:** To estimate the effects of different treatments on ROS generation, 2,7-dichloro dihydro fluorescein diacetate (H2DCFDA) dye was used. The stained cells were further incubated with 1uM of H2DCFDA for 10” in dark (7).

**Primary Mice Enterocytes (PMEs) Isolation:** Mice were euthanized per institutional guidelines followed by isolation of small intestines which were opened, washed gently, and placed in ice-cold PBS. After mincing into 1mm fragments, the single cell suspension was prepared from the intestine using frosted glass slides. The cell pellet was re-suspended in complete DMEM (10% FBS) and seeded at 37˚C, 5% CO2 at the density of 1x106 per ml for further experiments (8).

**Proteomics:**

Treated PHNs (samples shown in figure 8C, E and F) and PMEs (Figure 9B, 9E and 9G) were subjected to proteomics analysis. Media was removed from the samples and pelleted cell were washed with PBS. Cells were then lysed using cell lysis buffer and subjected to protein quantification. 50ug of protein was taken from each sample. Samples were treated with DDT, IAA followed by trypsin digestion. Trypsin digested peptide were passes through C18 columns and lyophilized. Samples were reconstituted in 0.1% formic acid before running on mass spectrometry. Global and targeted proteomics was performed on the samples (5). Targeted analysis using Data independent acquisition was performed to determine the expression of glucocorticoid receptor and downstream signalling molecules directly involved in its regulation (GRα, GRß, Annexin, Interferon regulatory factor, IkB, NF-kB and CREB), and of gap junction proteins such as gap junction protein 1, desmoglobin, desmoplakin, plakoglobin, and occluding.

**Pathway Activity Analysis:** All the enriched proteins were subjected to enrichr and pathways associated with the uploaded proteins were enriched. Pathways that showed enrichment of more than 3 proteins with significant p-value of <0.05 were considered for further analysis. Expression of all the proteins belonging to a specific pathway in different treatment groups such as 60uM urobilinogen, LPS, PMA and others were retrieved from the raw files and average was taken for each group. These average values for each group reflected the pathway activity of the respective groups and were then compared to assess the effect of different treatment (60uM urobilinogen, LPS, PMA and others) on the PHNs.

**Permeability Assay:**

**Effect of urobilinogen treatment on membrane integrity of Primary Healthy Neutrophils:** To assess the effect of urobilinogen on the cell permeability, 4 groups of PHN were taken and stimulated with 10ng/ml PMA for two hours and were further treated as:

Group 1 (Unstained): Cells were treated with cytofix and cytoperm for 10 min followed by PBS wash and acquisition.

Group 2 (Control): Cells were treated with cytofix and cytoperm for 10 min followed by staining with APC-IFNg for 30 min and acquisition.

Group 3 (Treatment): Cells were treated with 60uM urobilinogen for 60min followed by staining with APC-IFNg for 30 min and acquisition.

Group 4 (Treatment): Cells were treated with 60uM urobilinogen for 90min followed by staining with APC-IFNg for 30 min and acquisition.

The stained cells were acquired using BD FACS ARIA III flow cytometer, and the recorded data was analysed using FlowJo software version 10.

**Effect of urobilinogen treatment on membrane integrity of Primary Mice Enterocytes:** Since urobilinigen is produced in large amount in gut by intestinal flora and is in persistent exposure of high concentration of urobilinogen, we wanted to investigate the effect of urobilinogen on the membrane integrity of enterocytes. To assess this, enterocytes isolated from mice were treated as:

Group 1: Control group.

Group 2: Cells were treated with 60uM urobilinogen for 1.5 hr.

Group 3: Cells were treated with 30ng/ml LPS for 1.5 hr.

Group 4: Cells were treated with 30ng/ml LPS along with 60uM urobilinogen for 1.5 hr.

Group 5: Cells were treated with 5% alcohol for 1.5 hr.

Group 6: Cells were treated with 5% alcohol along with 60uM urobilinogen for 1.5 hr.

Post treatment cells were collected and were further prepared for proteomics analysis.

6.	Oh H, Siano B, Diamond S. Neutrophil isolation protocol. J Vis Exp 2008.

7.	Wu D, Yotnda P. Production and detection of reactive oxygen species (ROS) in cancers. J Vis Exp 2011.

8.	Ren HJ, Zhang CL, Liu RD, Li N, Li XG, Xue HK, Guo Y, et al. Primary cultures of mouse small intestinal epithelial cells using the dissociating enzyme type I collagenase and hyaluronidase. Braz J Med Biol Res 2017;50:e5831.
