## supplementary figures for "Plasma multi-omics outlines association of urobilinogen with corticosteroid non-response, inflammation and leaky gut in Sever Alcoholic Hepatitis"

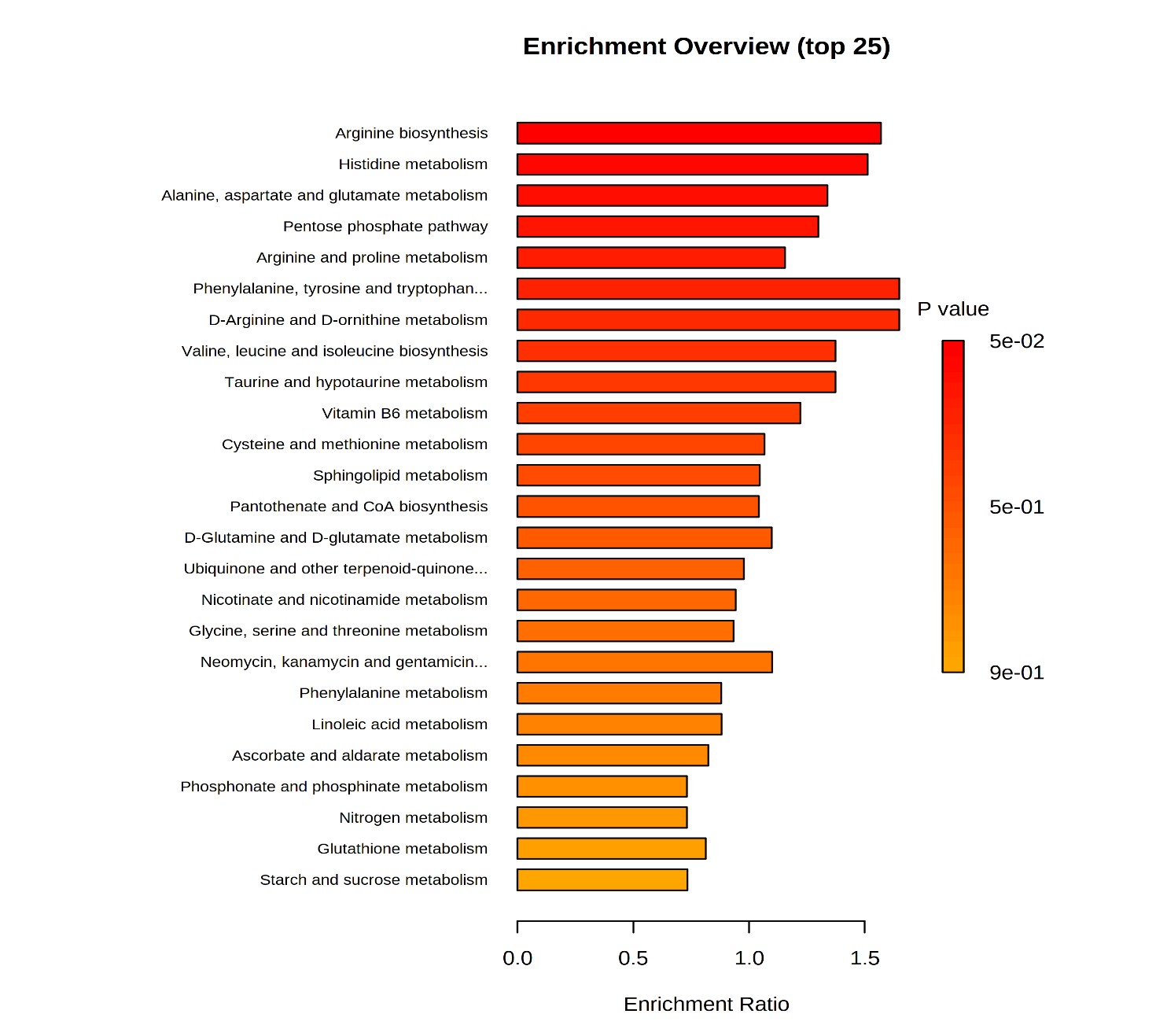
**Supplementary figure 1**: the bar plot here represents the pathways associated with the total enriched metabolites in the plasma of SAH patients. Enriched metabolites were subjected to enrichment analysis and it was found that these metabolites belonged to super-class: amino acids, fatty acid, monosaccharides, purines, pyrimidine, indoles, steroids, TCA acids, quinone, bilirubin, and bile acids (metabolite enrichment > metabolites per super class, p>0.05)

**Supplementary figure 2:** Bar plot showing species abundance in responders and non-responders at baseline.

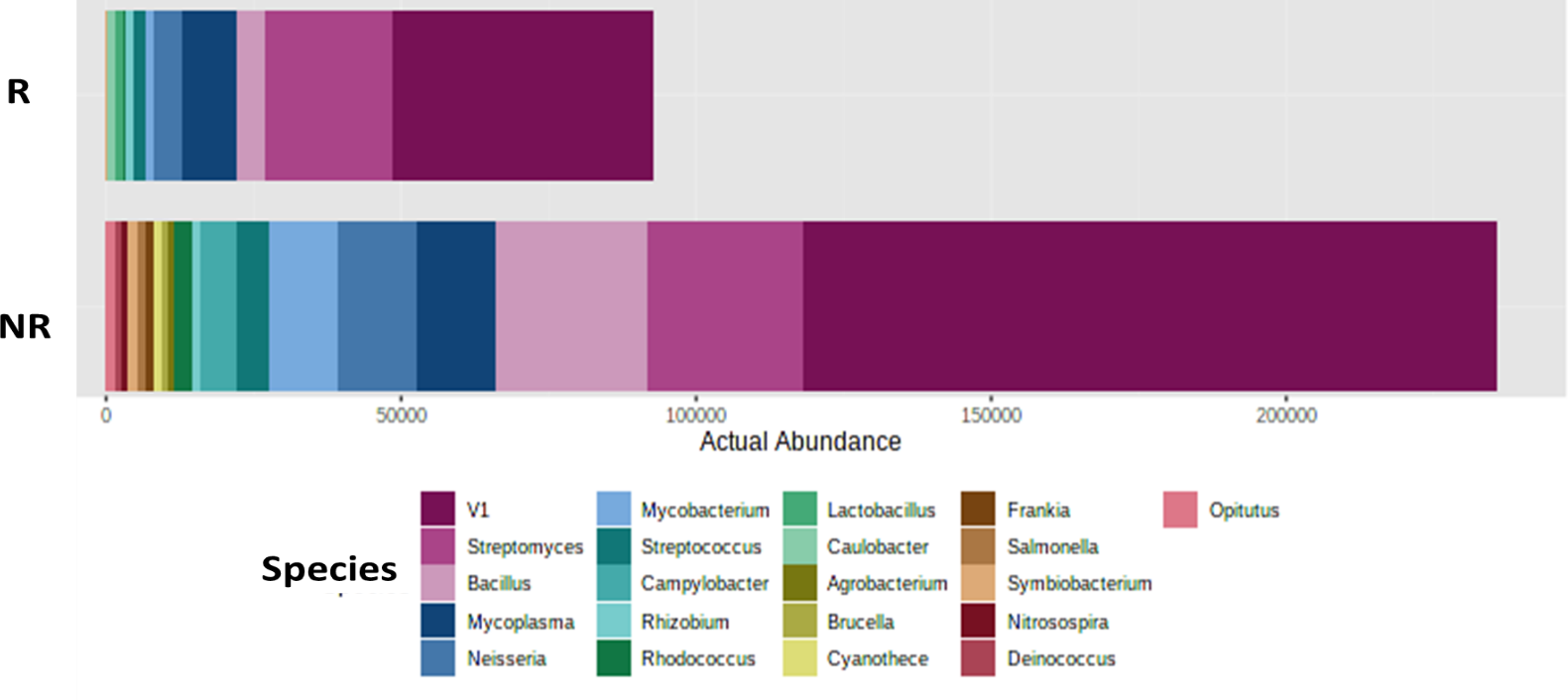

**Supplementary figure 3**: Loading plot of PLSDA showing featured metabolites at baseline.

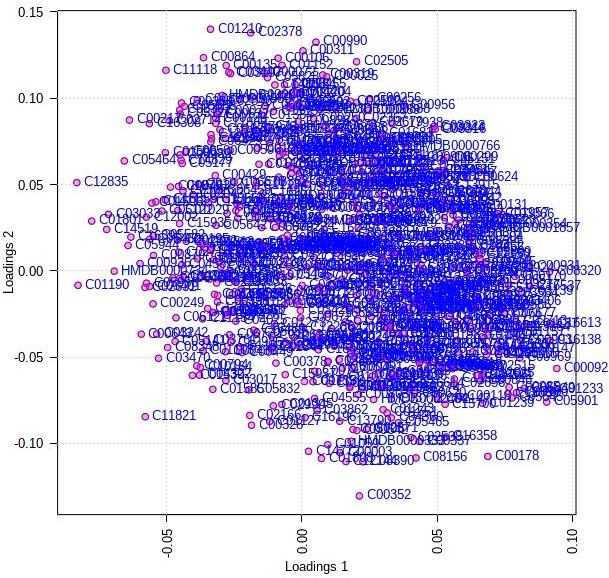

**Supplementary Figure 4**: Loading plot of PLSDA showing featured metabolites in temporal change analysis.

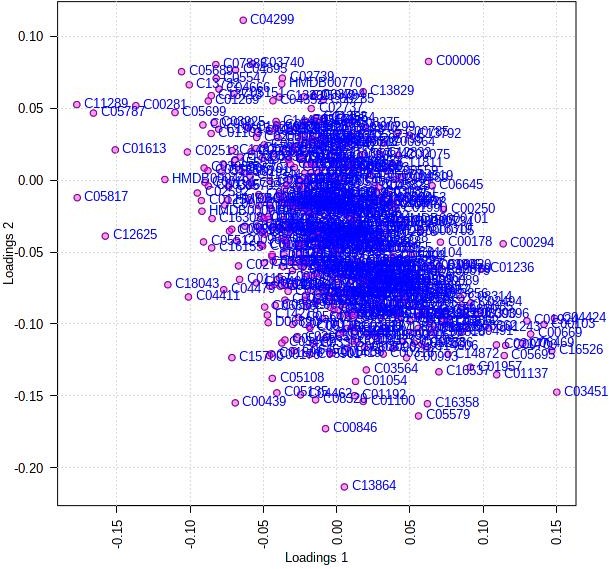

**Supplementary Figure 5:** Venn diagram showing number of metabolites (differentially upregulated) and associated pathways at baseline, day 3, and day 7 respectively in NR.

Metabolites associated with amino acid, secondary bile acid, beta-oxidation, sphingolipid metabolism, steroid hormone biosynthesis, glycerol-phospholipid metabolism, phenylalanine, tyrosine, and tryptophan biosynthesis remained highly concentrated in the plasma of NR. Interestingly we observed a temporal and radical increase in the metabolic by-product of bilirubin such as urobilin, and urobilinogen in the plasma of NR. A significant increase in bilirubin and its metabolic by-products in NR could be due to the inefficiency of hepatocytes to clear bilirubin or increased metabolism of bilirubin by the intestinal microbiome in NR (intestinal flora converts bilirubin to urobilinogen), though this needs to be validated.

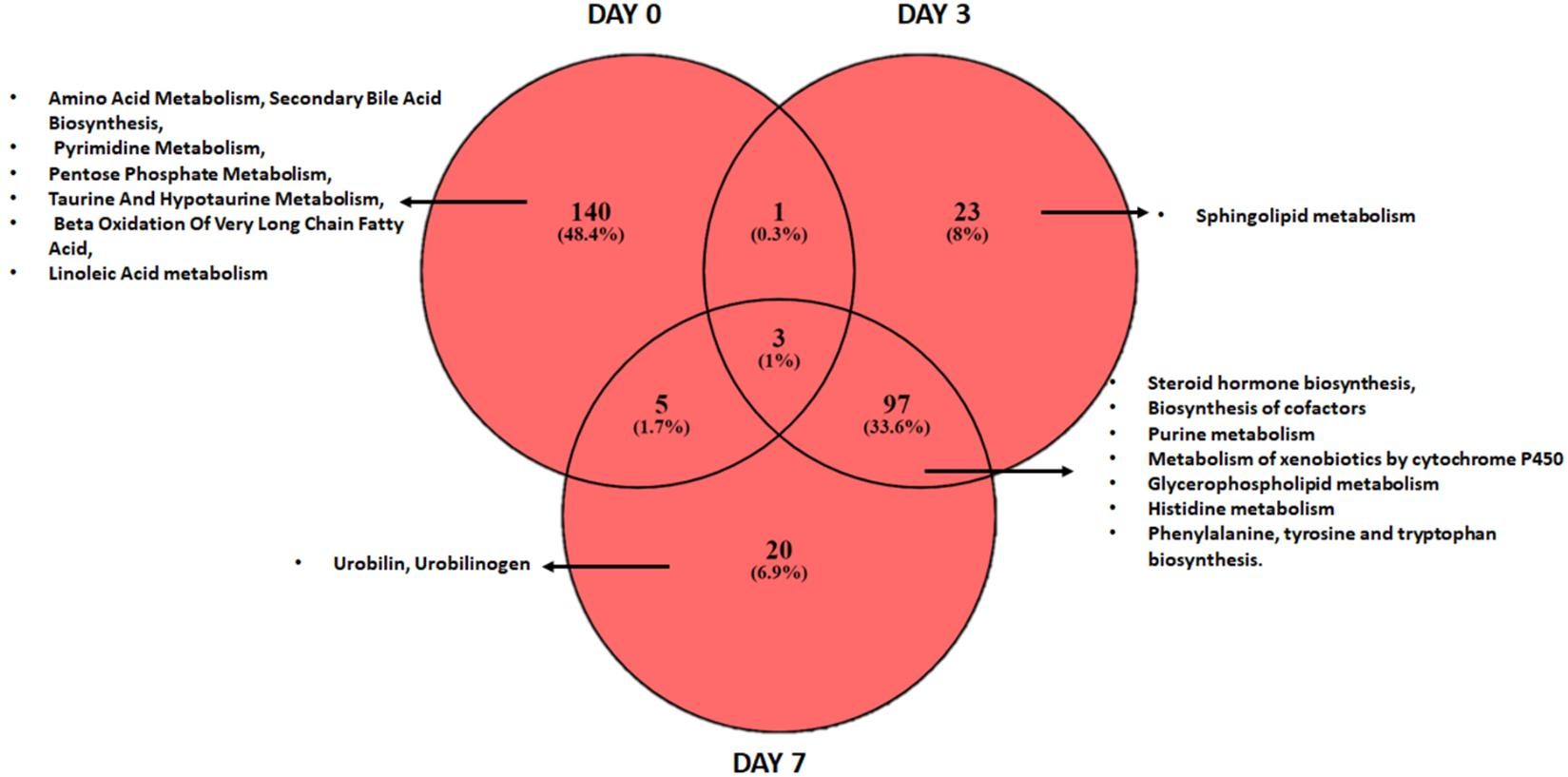

**Supplementary Figure 6:** Venn diagram showing number of significantly downregulated metabolites and associated pathways at baseline, day 3, and day 7 respectively in NR. The lower expression in NR means that these metabolites have higher expression in R. Therefore, we can say that R showed a basal increase in pyrimidine, pantothenate CoA biosynthesis, and others; metabolites linked to nicotinate, and nicotinamide metabolism, valine, leucine, and isoleucine biosynthesis, and others at day3 whereas butanoate and alanine metabolism, purine metabolism, various amino acid metabolism, biosynthesis of amino-Acyl t-RNA, unsaturated fatty acid, and energy metabolism (pentose phosphate pathway, TCA cycle) were increased at day7. Based on these results, we infer that in response to glucocorticoid therapy the metabolic profile of the patient changes toward metabolic, regenerative, and energy pathways (TCA cycle, amino acid synthesis, and translational machinery) associated with metabolites in R and the failure of these pathways in NR could be one of the reasons for unresponsiveness.

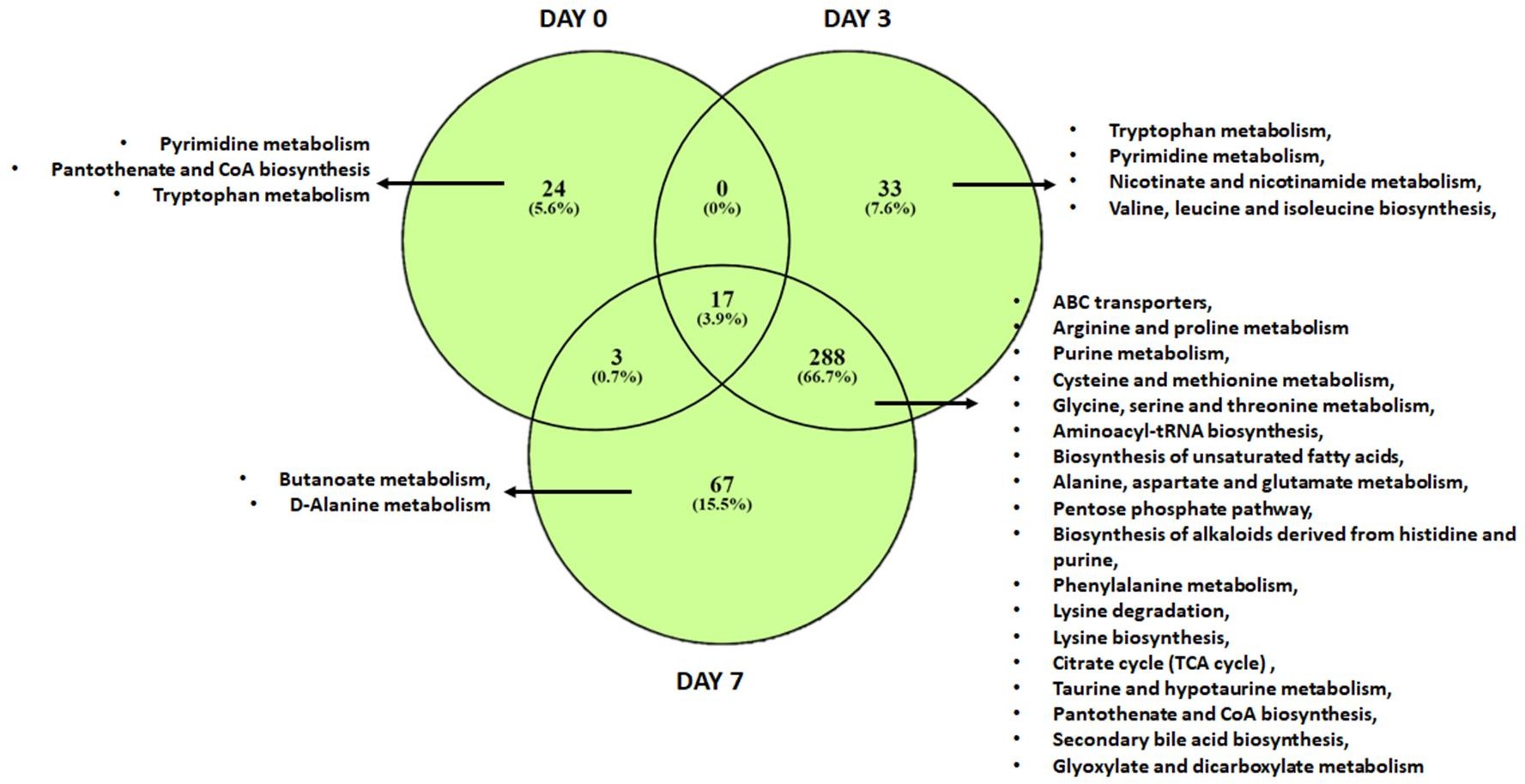

**Supplementary Figure 7:** Bar plots showing the change in the number of metabolites (up = red, down = green, and with no change = blue color, respectively) in different modules in NRs compared to Rs during treatment.

Black, Brown, Green, Pink, Red, Turquoise, Yellow, and blue module show an increase in the number of metabolites suppressed in NRs over time compared to Rs.

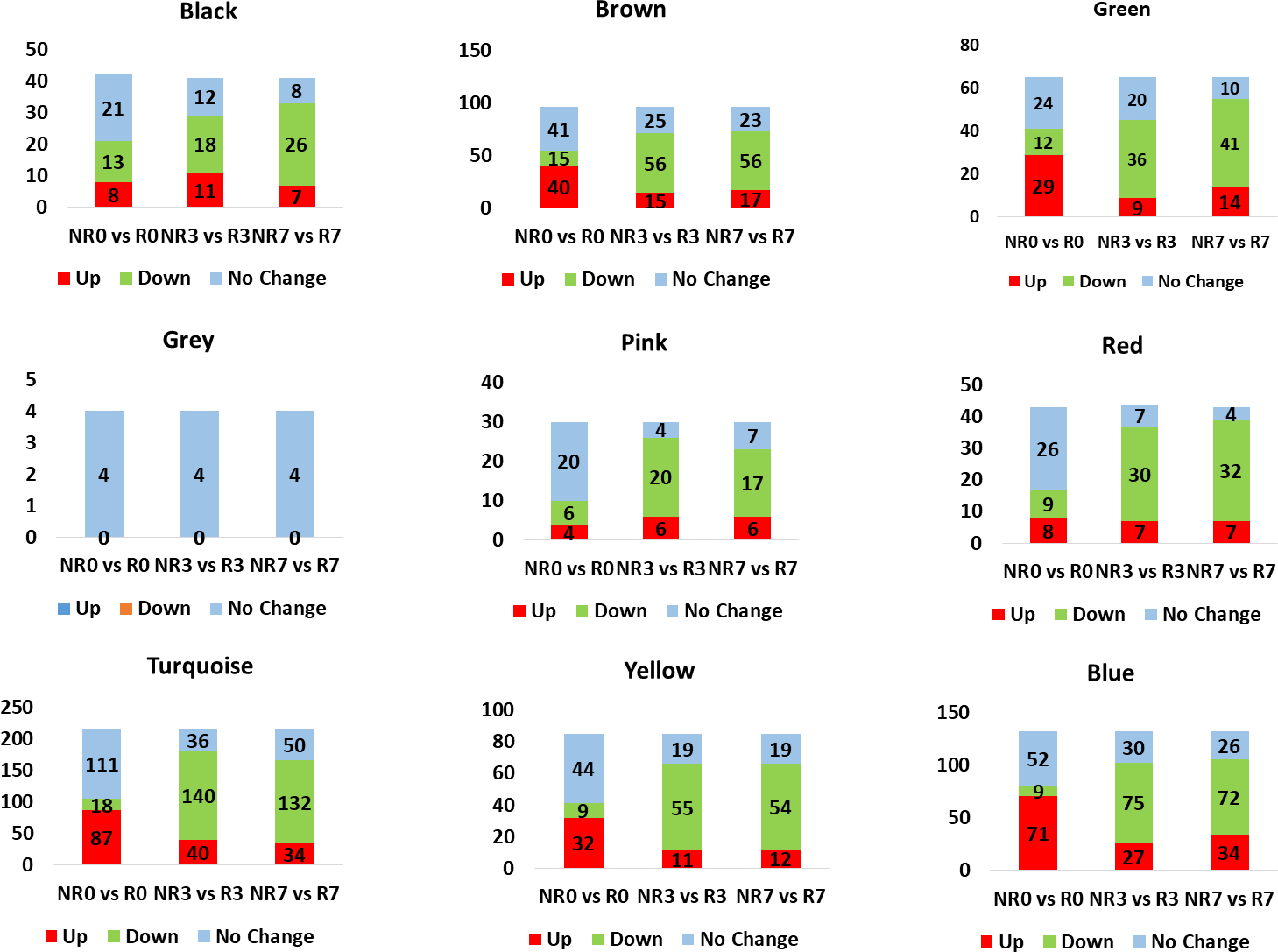

**Supplementary Figure 8**: AUC univariate analysis showing panel of metabolites capable of segregating R and NR (p<0.05, FC>1.5<). Of all Urobilinogen (C05791) augmented highest Area under the curve (AUC).

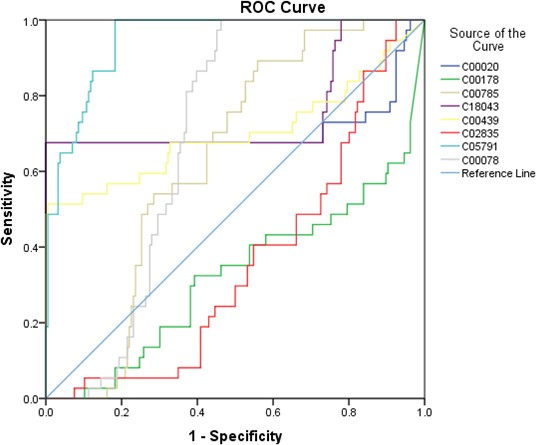

**Supplementary Figure 9**: Standard curve for calculating plasma urobilinogen concentration in R and NR. Bar plot showing concentration of urobilinogen in R and NR respectively in mg/ml. A cut off of above than 0.07mg/ml segregated NR from R.

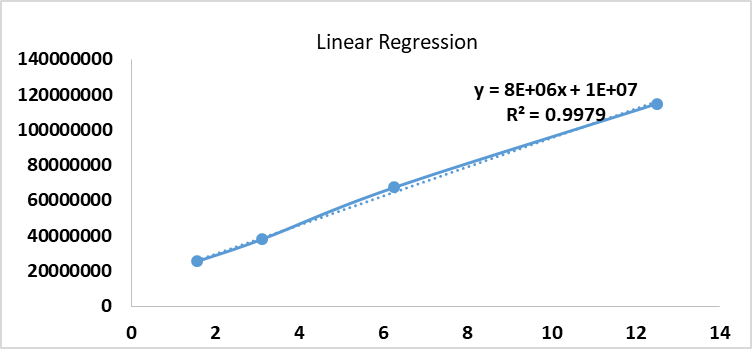

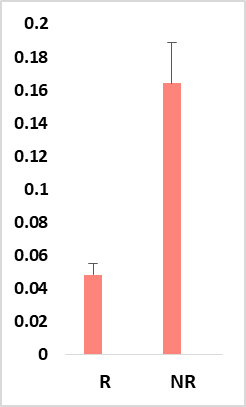

**Supplementary Figure 10**: Spectra showing Urobilinogen standard peaks at different dilutions (1:1, 1:2, 1:4, and 1:8).

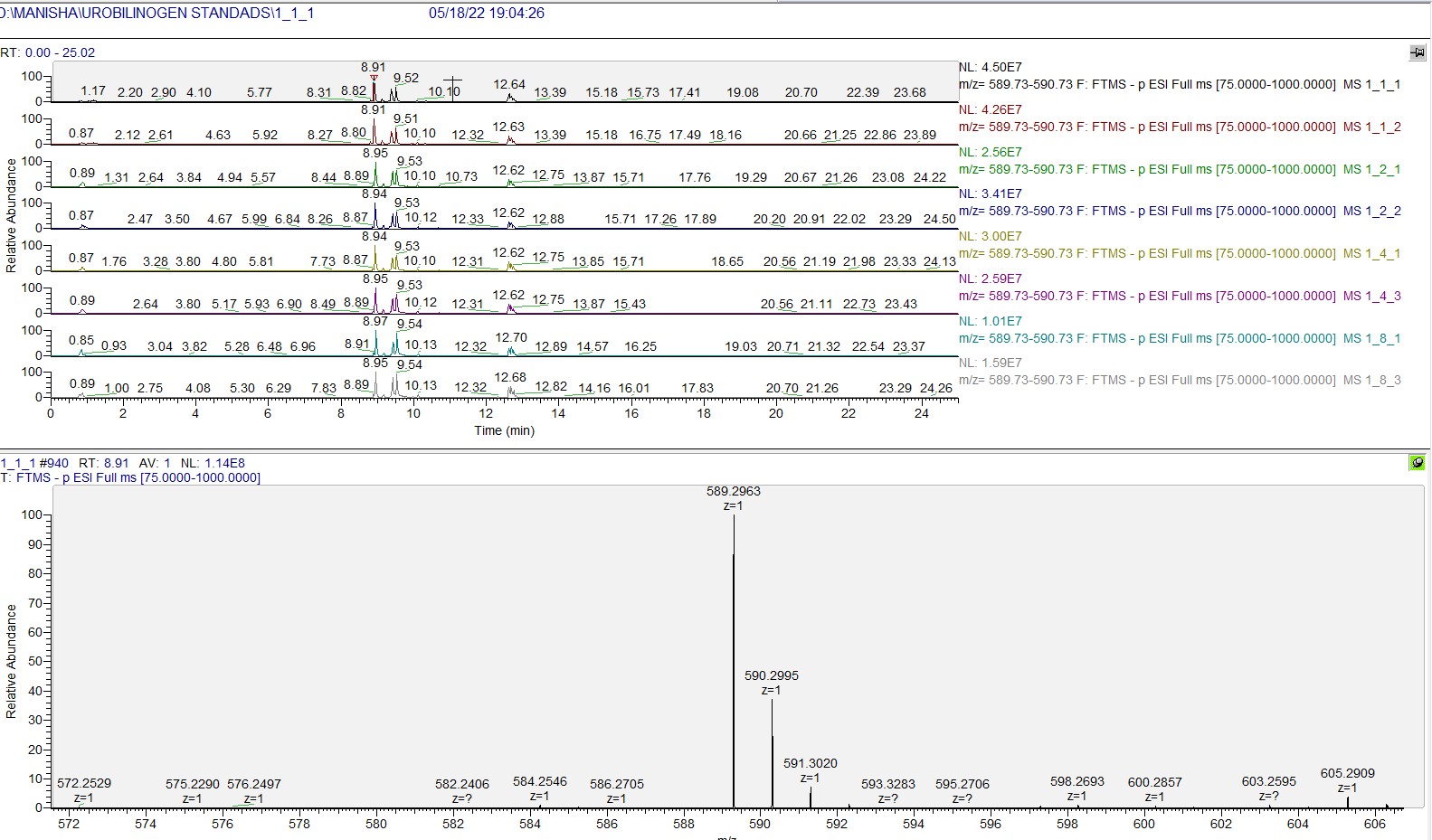

**Supplementary Figure 11:** Table displaying area under the curve for C05791, CTP, MELD, and DF. Urobilinogen attained the highest AUC and showed highest capability to predict severity at baseline.

**Area Under the Curve**

| Test Result Variable(s) | Area | Std. Error^a^ | Asymptotic Sig.^b^ | Asymptotic 95% Confidence  Interval | |
| --- | --- | --- | --- | --- | --- |
|  |  |  |  | Lower Bound | Upper Bound |
| C05791(Urobilinogen) | 0.834 | 0.033 | 0 | 0.768 | 0.9 |
| CTP_D0 | 0.799 | 0.037 | 0 | 0.725 | 0.872 |
| MELD_D0 | 0.732 | 0.039 | 0 | 0.656 | 0.809 |
| DF_D0 | 0.821 | 0.035 | 0 | 0.753 | 0.89 |

**Supplementary Figure 12:** Machine learning-based validation of Urobilinogen as the marker for mortality in SAH patients. The figure documents the model efficiency in terms of model accuracy and the kappa value of the model.

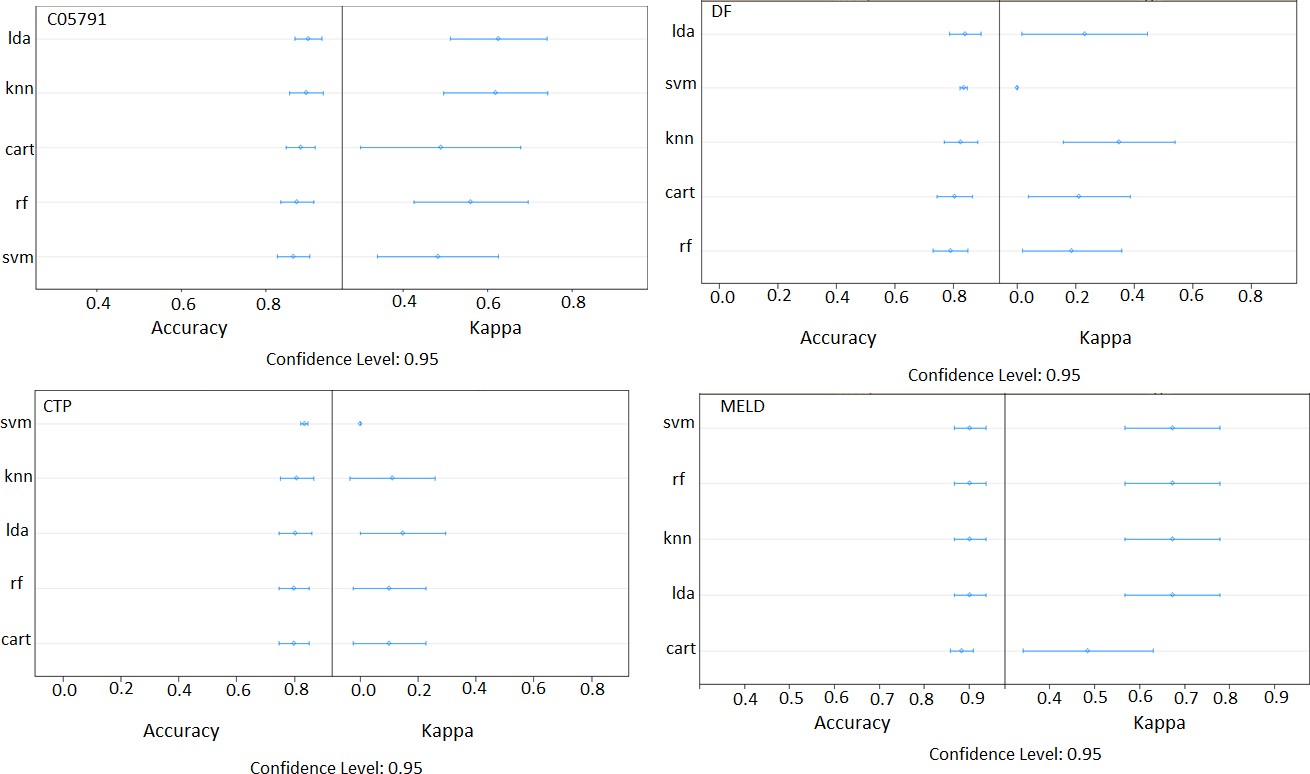

**Supplementary Figure 13:** Shannon and Simpson index showing microbial diversity.

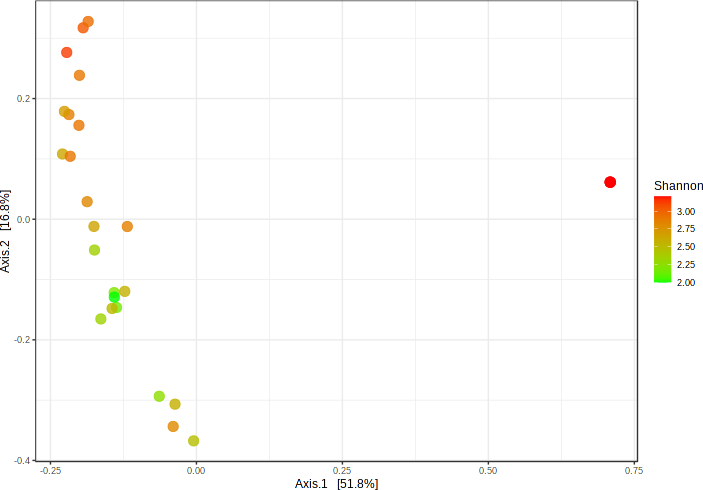

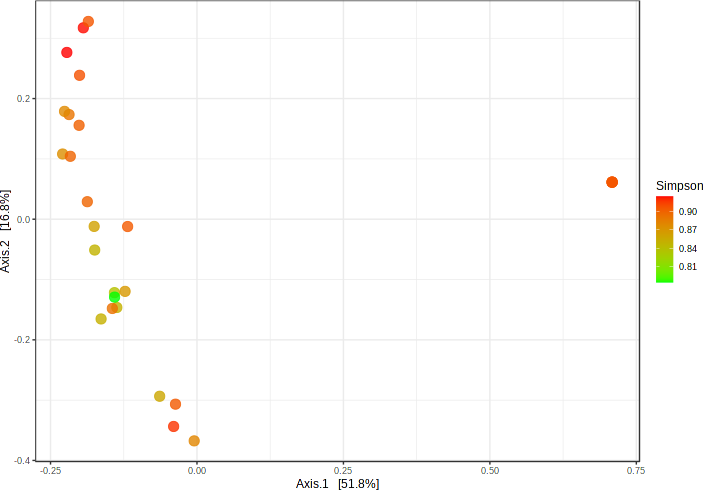

**Supplementary Figure 14:** Dot plot showing pathways associated with the proteins commonly upregulated by PMA, LPS and urobilinogen. When the neutrophils treated with PMA, LPS and Urobilinogen were subjected to proteomics analysis, it was observed that pathways associated with TRAIL, RAC1,TNFa, TGFb, MAPK, mTOR, JNK, IL8, IL6, IFNg, and others got highly expressed post treatment suggesting that urobilinogen induces inflammation in a similar manner as PMA/LPS induces.

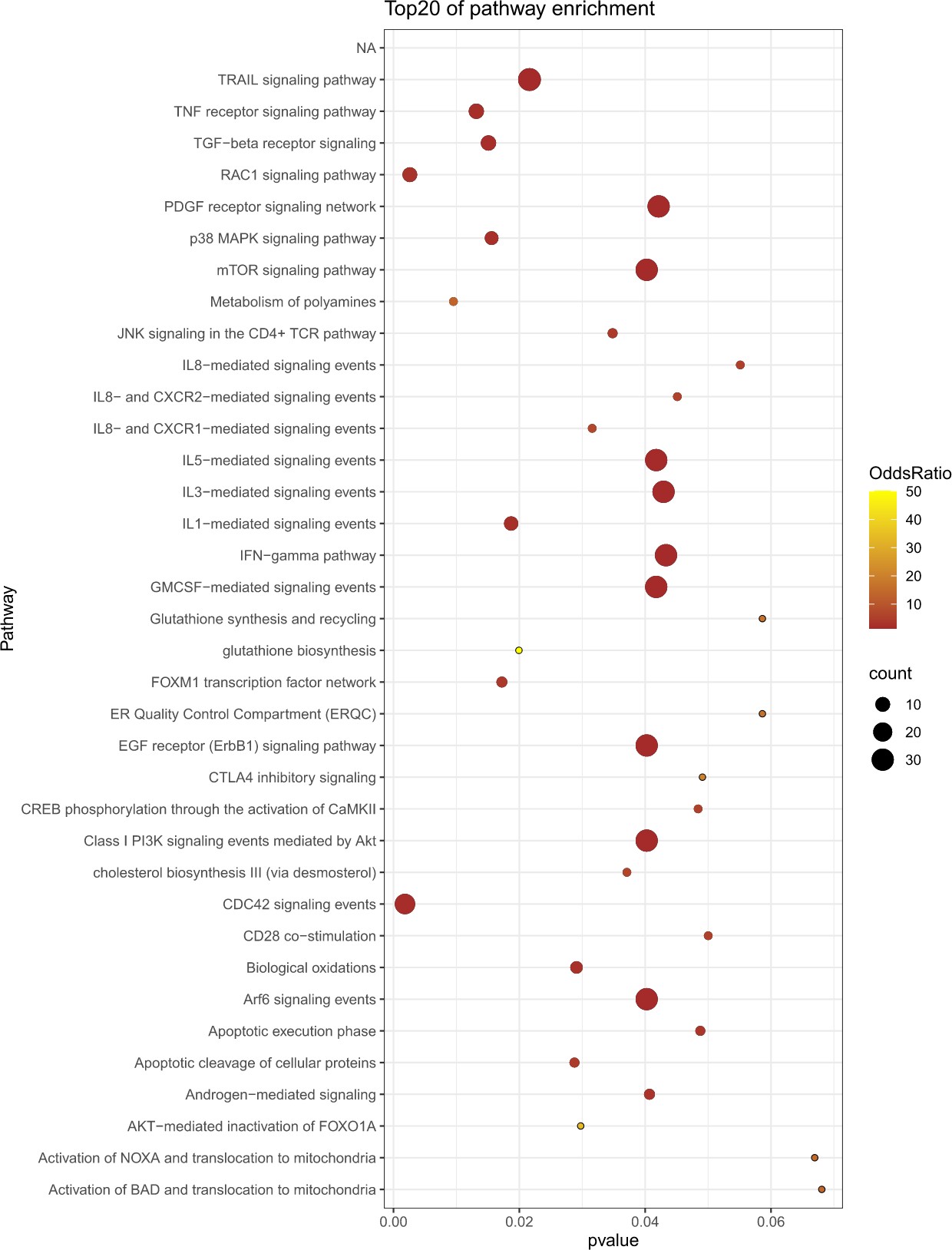
