## supplementary tables for "Plasma multi-omics outlines association of urobilinogen with corticosteroid non-response, inflammation and leaky gut in Sever Alcoholic Hepatitis"

**Supplementary table 1**

| **Demographic Profile** | **Discovery Cohort** | | | | | **Validation Cohort** | | | | |
| --- | --- | --- | --- | --- | --- | --- | --- | --- | --- | --- |
|  | Responders | | Non-responders (n=20) | | 'p' value | Responders | | Non-responders (n=17) | | 'p' value |
|  | (n=50) | |  |  |  | (n=135) | |  |  |  |
|  | Median | Range | Median | Range |  | Median | Range | Median | Range |  |
|  |  | (Min – max) |  | (Min – max) |  |  | (Min – max) |  | (Min – max) |  |
| Age (years) | 39 | 29-59 | 37 | 25-68 | 0.62 | 34 | 26-65 | 30 | 24-71 | 0.52 |
| BMI (kg/m2) | 25.12 | 16.48-37.23 | 27.39 | 18.64-39.51 | 0.72 | 22.5 | 12.16-38.03 | 31 | 19.01-45.91 | 0.24 |
| Age of onset of alcohol (yr) | 27 | 15-51 | 26 | 20-36 | 0.76 | 23 | 16-50 | 28 | 21-39 | 0.55 |
| Jaundice duration (days) | 29 | 11-86.0 | 40 | 20-63 | 0.46 | 28 | 10-95.0 | 37 | 29-51 | 0.63 |
| Ascites duration (days) | 10 | 0-69 | 20 | 3-45.0 | 0.35 | 15 | 0-76 | 23 | 1-52.0 | 0.54 |
| Jaundice to ascites interval(days) | 13 | 0-86 | 3.5 | 0-62 | 0.36 | 16 | 0-83 | 4 | 0-51.0 | 0.64 |
| Alcohol to steroid interval (days) | 33 | 1-90.0 | 27 | 9-58.0 | 0.52 | 31 | 2-73.0 | 30 | 13-50.0 | 0.15 |
| T. Bilirubin (mg/dl) | 18.23 | 6-49.56 | 23.65 | 9.1-36.7 | 0.72 | 20 | 4.03-45.39 | 26.63 | 10.1-46.3 | 0.24 |
| Direct bilirubin (mg/dl) | 11.29 | 1.9-33.5 | 11.56 | 3.9-21.36 | 0.63 | 16 | 2-31.5 | 12 | 2.65-26.53 | 0.28 |
| AST (IU) | 123 | 60-401 | 194 | 61-329 | 0.06 | 120.5 | 50-391 | 185 | 59-370 | 0.05 |
| ALT(IU) | 93.5 | 8-173 | 60.9 | 32-147 | 0.29 | 43.26 | 12-160 | 64.6 | 33-141 | 0.89 |
| AST/ALT ratio | 2.36 | 1.67-11.85 | 2.42 | 1.46-5.13 | 0.72 | 3.66 | 2.35-12.36 | 2.96 | 1.5-6.32 | 0.24 |
| T. Protein (g/dl) | 7.13 | 2.9-8.19 | 7.1 | 5.6-7.8 | 0.18 | 6.5 | 3.5-8.0 | 6.3 | 5.5-7.9 | 0.18 |
| S. Albumin (g/dl) | 2.53 | 1.4-4.1 | 2.23 | 1.9-3.33 | 0.13 | 2.6 | 2-3.0 | 2.3 | 1.7-3.4 | 0.14 |
| INR | 1.57 | 1.5-4.0 | 2 | 1.56-3.1 | 0.72 | 2.02 | 1.56-4.04 | 1.98 | 1.65-3.05 | 0.44 |
| Hb (g/dl) | 9.9 | 6.3-14.8 | 9.6 | 7.2-11.9 | 0.83 | 7.9 | 6.8-14.1 | 9.8 | 6.4-13.3 | 0.32 |
| TLC (cells/micro liter) | 11.63 | 5.0-36.2 | 14.89 | 8-34.0 | 0.01 | 12.05 | 5.3-33.02 | 14.9 | 9-36.0 | 0.03 |
| Neutrophil % | 68 | 52-93 | 77 | 70-93 | 0.71 | 79 | 47-89 | 89 | 67-93.0 | 0.13 |
| Platelet count (cells/micro liter) | 137.6 | 39-384 | 210.36 | 31-419 | 0.04 | 143.5 | 46-155 | 237.09 | 40-388 | 0.02 |
| Urea (mg/dl) | 18.5 | 11.5-76 | 29.5 | 9-70.5 | 0.62 | 18.5 | 3.5-85.0 | 35 | 7-80.5 | 0.20 |
| S.Creatinine (mg/dl) | 0.53 | 0.1-1.33 | 0.56 | 0.04-1.16 | 0.83 | 0.44 | 0.07-1.49 | 0.54 | 0.09-1.43 | 0.32 |
| eGFR (mL/min/1.73m^2^) | 99 | 95.4-119.56 | 103 | 97-109 | 0.15 | 107 | 103.7-113.3 | 103 | 97.4-109.4 | 0.19 |
| S. Sodium (mEq/L) | 133.6 | 110-155.31 | 131.5 | 120-139 | 0.29 | 133.8 | 117-136.56 | 130 | 115-142 | 0.89 |
| S. Potassium (mEq/L) | 5.11 | 2.93-6.11 | 4.56 | 3.50-5.68 | 0.42 | 4.06 | 3.48-5.89 | 5.02 | 4.0-5.75 | 0.20 |
| S. TNF alfa | 10.4 | 1.26-516 | 13.5 | 06-273 | 0.58 | 12 | 0.5-710 | 7.06 | 0.5-681 | 0.76 |
| HVPG (mmHg) n=46 | 20 | 10-29.3 | 21.5 | 17-33 | 0.95 | 20 | 10.0-32.0 | 22 | 15-35.0 | 0.51 |
| CP score | 11 | 11-16.0 | 13.5 | 10-18.0 | 0.32 | 12 | 12-16.0 | 11 | 12-20.0 | 0.24 |
| MELD score | 23 | 19-32 | 26 | 17-34 | 0.94 | 25 | 20-34 | 26 | 15-32 | 0.09 |
| MELDNa | 26 | 19-36 | 31 | 19-40 | 0.83 | 27 | 21-40 | 35 | 17-45 | 0.30 |
| GAH score | 7 | 7-18.0 | 9 | 7-18.0 | 0.12 | 10 | 9-15.0 | 9.5 | 8-11.0 | 0.12 |
| MDF | 69.5 | 37-175 | 75.5 | 57-149 | 0.73 | 68 | 36-163 | 74 | 53-159 | 0.29 |
| Lille score | 0.1 | 0.03-0.46 | 0.8 | 0.56-0.994 | 0.00 | 0.15 | 0.07-0.49 | 0.73 | 0.45-1.15 | 0.01 |
| 90 day Mortality | 4/50(8%) | - | 10/20(50%) | - | 0.01 | 13/135(9.63%) | - | 12/17 (70%) | - | 0.01 |
| Demographic sketch for the Derivative and Validation cohort of SAH patients (Values documented in median and range), BMI: Body Mass Index, AST: Aspartate Aminotransferase Test , ALT: Alanine Aminotransferase Test, INR: International Normalized Ratio, Hb: Haemoglobine levels, TLC: Total leukocytes count, TNFalpha: Tumor necrosis factor alpha, CP: Child-Pugh score, MELD: Model of End Stage Liver Diseases, eGFR= Estimated glomerular filtration rate. GAH: Glassgo alcoholic hepatitis score, MDF: Maddery discriminant function , HVPG: Hepatic vein pressure gradient. | | | | | | | | | | |

**Supplementary table 2**

|  | **Annotation final** | **Molecular Weight** | **peak intensity** | **rt** | **rtmin** | **rtmax** | **Kegg/HMDB** | Name | Identification | Average expression of metabolites of Rs at baseline | Average expression of metabolites at day 3 of Rs | Average expression of metabolites at day 7 of Rs | Average expression of metabolites at baseline of NRs | Average expression of metabolites at day 3 of NRs | Average expression of metabolites at day 7 of NRs |
| --- | --- | --- | --- | --- | --- | --- | --- | --- | --- | --- | --- | --- | --- | --- | --- |
| 1 | C00003 | 665.1305542 | 9667.184532 | 1566.42326 | 1559.521102 | 1566.90051 | C00003 | NAD | Mass, RT and Spectra | 824.0211963 | 56.83221739 | 17533.35122 | 8208.401325 | 2921.500649 | 3731.458357 |
| 2 | C00006 | 745.0842213 | 16892.33139 | 1073.796412 | 1070.612802 | 1076.20937 | C00006 | NADP | Mass, RT and Spectra | 14104.77095 | 23240.11007 | 18886.85052 | 3777.028744 | 11761.63681 | 9472.320059 |
| 3 | C05151 | 617.408394 | 12064.77576 | 690.2584538 | 687.8156815 | 691.7784724 | C05151 | phorbol 13-acetate 12-myristate | Mass, RT and Spectra | 19536.58381 | 4144.174332 | 2436.856811 | 19493.4375 | 23908.59125 | 39607.2546 |
| 4 | C05177 | 597.2980579 | 263905.8481 | 756.5202346 | 747.6225187 | 758.7183741 | C05177 | Berbamunine | Mass, RT and Spectra | 310729.9492 | 197401.277 | 196281.1017 | 311453.165 | 367468.0353 | 384443.4879 |
| 5 | C00008 | 428.0333914 | 49641.31661 | 147.9524325 | 139.0987726 | 154.302756 | C00008 | ADP | Mass, RT and Spectra | 56352.44275 | 54415.97538 | 47811.67993 | 57060.22191 | 49379.82419 | 48616.17432 |
| 6 | C00009 | 98.98418789 | 2303874.189 | 90.01489963 | 86.11991911 | 114.3058125 | C00009 | Hydrogen phosphate | Mass, RT and Spectra | 8155.378409 | 5005.751595 | 5185.748194 | 19340.83703 | 7470.904505 | 14097.71153 |
| 7 | C00020 | 345.9982246 | 11351.68356 | 481.7763577 | 479.6314274 | 484.0110973 | C00020 | Adenosine monophosphate | Mass, RT and Spectra | 1192.829308 | 23700.96555 | 23050.81556 | 2673.689584 | 2487.526823 | 4381.144899 |
| 8 | C00022 | 87.00840443 | 364808.9339 | 468.829667 | 467.6452012 | 470.5178683 | C00022 | Pyruvic acid | Mass, RT and Spectra | 34990.99707 | 717923.412 | 735543.0486 | 49710.70582 | 112868.5787 | 115393.9252 |
| 9 | C05239 | 82.04071458 | 5046.913866 | 1163.862961 | 1160.091074 | 1168.136451 | C05239 | 5-Aminoimidazole | Mass, RT and Spectra | 97.55529376 | 10259.54675 | 10722.96736 | 464.2901484 | 787.4277641 | 2395.02486 |
| 10 | C00025 | 146.0453365 | 1124939.262 | 468.259162 | 466.5166905 | 469.8889483 | C00025 | L-Glutamic acid | Mass, RT and Spectra | 174015.234 | 1763615.161 | 2771956.673 | 352019.7912 | 378876.9377 | 232807.8544 |
| 11 | C00037 | 76.03924797 | 4116869.269 | 104.5551558 | 102.1737052 | 113.6161793 | C00037 | Glycine | Mass, RT and Spectra | 2541217.743 | 5559035.33 | 5718444.763 | 2985777.766 | 3446676.148 | 2871375.49 |
| 12 | C00041 | 90.05491358 | 29072505.45 | 109.2052118 | 91.00134715 | 119.8393172 | C00041 | L-Alanine | Mass, RT and Spectra | 21407556.92 | 38739300.59 | 38495680.32 | 20254500.56 | 29216222.9 | 22098961.74 |
| 13 | C00042 | 117.0189847 | 715715.5031 | 475.4541937 | 473.2189912 | 486.2754152 | C00042 | Succinic acid | Mass, RT and Spectra | 84227.91659 | 1139334.95 | 1451656.815 | 201116.4673 | 271571.0924 | 321988.587 |
| 14 | C00047 | 145.0977916 | 930948.5189 | 545.9769774 | 538.1133905 | 556.1100725 | C00047 | L-Lysine | Mass, RT and Spectra | 166785.3576 | 1833839.958 | 1804212.12 | 221176.8226 | 405767.6046 | 398457.1444 |
| 15 | C00049 | 132.0297623 | 390468.1107 | 470.1370333 | 467.2155626 | 485.2972453 | C00049 | L-Aspartic acid | Mass, RT and Spectra | 61166.35765 | 768956.0646 | 802912.9076 | 69630.95125 | 179398.7824 | 170291.7526 |
| 16 | C00053 | 506.9848882 | 815715.5258 | 889.744238 | 888.3175367 | 890.5062128 | C00053 | Phosphoadenosine phosphosulfate | Mass, RT and Spectra | 13114.59233 | 9804.816416 | 23986.5391 | 2561.935266 | 1835.608894 | 5495.636474 |
| 17 | C00062 | 173.103647 | 1035979.96 | 571.2765134 | 557.0887237 | 584.4932161 | C00062 | L-Arginine | Mass, RT and Spectra | 211059.3441 | 1986940.341 | 2417010.38 | 183942.5365 | 328863.6842 | 294051.563 |
| 18 | C00063 | 483.9950152 | 14264.58905 | 889.3080276 | 888.4345985 | 890.2487114 | C00063 | Cytidine triphosphate | Mass, RT and Spectra | 14099.97731 | 221471.5562 | 174594.6935 | 19650.67758 | 28328.00233 | 26511.50831 |
| 19 | C00065 | 104.0349948 | 1276495.196 | 429.9112093 | 419.390084 | 433.6183328 | C00065 | L-Serine | Mass, RT and Spectra | 215701.3092 | 2456865.228 | 2618350.246 | 253751.5093 | 437280.9684 | 462349.6024 |
| 20 | C05290 | 303.1946554 | 13624.16486 | 9.309203618 | 4.224452529 | 12.12357261 | C05290 | 19-Hydroxyandrost-4-ene-3,17-dione | Mass, RT and Spectra | 15872.14454 | 7246.485172 | 7167.854784 | 7606.814169 | 48400.86434 | 63360.65991 |
| 21 | C17432 | 428.2788255 | 1410281.074 | 832.8473715 | 827.5118624 | 833.8625474 | C17432 | all-trans-Decaprenyl diphosphate | Mass, RT and Spectra | 2308783.975 | 2146597.455 | 1404535.526 | 2861119.456 | 2553029.439 | 2382879.222 |
| 22 | C00072 | 203.0819259 | 226540.7933 | 102.152885 | 96.87342109 | 104.4466872 | C00072 | Ascorbate | Mass, RT and Spectra | 104527.2879 | 52517.42209 | 49881.62 | 91319.08851 | 81284.6614 | 79415.50844 |
| 23 | C00073 | 148.0433372 | 3400051.026 | 234.5332321 | 212.8205789 | 249.2690555 | C00073 | L-Methionine | Mass, RT and Spectra | 337046.2239 | 8128706.247 | 7153894.01 | 530781.4005 | 2183594.203 | 3564538.244 |
| 24 | C05350 | 180.0415774 | 1429359.632 | 137.7119639 | 131.6073891 | 145.201175 | C05350 | 2-Hydroxy-3-(4-hydroxyphenyl)propenoic acid | Mass, RT and Spectra | 90784.11475 | 68892.16792 | 61800.10844 | 95530.76948 | 91848.52607 | 87698.24857 |
| 25 | C05381 | 528.0886936 | 53785.27912 | 320.6592926 | 303.0945265 | 328.1526672 | C05381 | 3-Carboxy-1-hydroxypropylthiamine diphosphate | Mass, RT and Spectra | 86393.74968 | 25390.39141 | 25966.7709 | 69154.06646 | 47541.00675 | 55580.02364 |
| 26 | C00075 | 484.9730189 | 20826.65565 | 889.4905304 | 888.0648508 | 890.6914952 | C00075 | Uridine triphosphate | Mass, RT and Spectra | 18615.08227 | 15937.4931 | 32138.2289 | 6475.873606 | 5205.281507 | 9007.222302 |
| 27 | C05400 | 281.0857274 | 8820.23349 | 514.8565432 | 512.993699 | 515.7083847 | C05400 | Isomaltose | Mass, RT and Spectra | 43.21640664 | 19914.85225 | 20448.71352 | 4113.707228 | 413.9292954 | 1813.520778 |
| 28 | C05403 | 341.110202 | 189675.4463 | 549.2238003 | 545.063005 | 551.1375531 | C05403 | 3'-Ketolactose | Mass, RT and Spectra | 1769.713757 | 44747.09832 | 60913.7609 | 6447.101459 | 7139.199884 | 9192.294415 |
| 29 | C00077 | 133.0966727 | 1193190.201 | 107.3564758 | 99.9220957 | 117.4375394 | C00077 | Ornithine | Mass, RT and Spectra | 1246501.742 | 1145210.903 | 1139930.477 | 1022150.228 | 1426454.136 | 1287203.091 |
| 30 | C00078 | 113.035554 | 73318.69443 | 264.259595 | 249.866065 | 276.2618903 | C00078 | L-Tryptophan | Mass, RT and Spectra | 49847.95293 | 102089.3891 | 109060.3844 | 68276.57813 | 68971.87882 | 72445.04879 |
| 31 | C00079 | 164.0711855 | 3462405.941 | 185.7153448 | 163.0270625 | 201.0076447 | C00079 | L-Phenylalanine | Mass, RT and Spectra | 656903.2061 | 6551352.568 | 7097693.961 | 622392.6084 | 1279698.529 | 1390948.4 |
| 32 | C00082 | 180.0661176 | 3374862.157 | 311.061209 | 288.739864 | 330.2060148 | C00082 | L-Tyrosine | Mass, RT and Spectra | 780778.6035 | 6445197.889 | 7556631.259 | 631761.5651 | 878746.74 | 975625.6037 |
| 33 | C00085 | 259.0209183 | 14971.20592 | 491.3824395 | 481.5523833 | 518.3677575 | C00085 | Fructose 6-phosphate | Mass, RT and Spectra | 372.7758956 | 23810.55515 | 38563.35469 | 4149.334032 | 7540.984494 | 5103.019194 |
| 34 | C00089 | 341.1080369 | 35158.87936 | 486.7098363 | 480.2283398 | 493.3616731 | C00089 | Sucrose | Mass, RT and Spectra | 2351.702786 | 69505.88558 | 63731.66417 | 16127.50187 | 25552.02336 | 18111.43884 |
| 35 | C00092 | 318.9811508 | 5635.914409 | 314.5705045 | 311.4523463 | 320.0909584 | C00092 | Glucose 6-phosphate | Mass, RT and Spectra | 24.67107329 | 12771.48873 | 13700.43526 | 83.81879447 | 409.0063221 | 887.8152664 |
| 36 | C00097 | 122.0267947 | 43965.7637 | 104.8440404 | 99.98971339 | 110.083666 | C00097 | L-Cysteine | Mass, RT and Spectra | 24845.34776 | 51769.78403 | 59266.14752 | 39612.29327 | 43183.08626 | 59520.93628 |
| 37 | C00103 | 241.0119795 | 14782.46054 | 477.2663553 | 475.9543673 | 479.2491357 | C00103 | Glucose 1-phosphate | Mass, RT and Spectra | 1959.658903 | 31004.3456 | 34615.53756 | 1043.472141 | 1025.511897 | 1980.990207 |
| 38 | C00104 | 427.007753 | 187046.0122 | 529.89018 | 519.1061374 | 537.7486506 | C00104 | IDP | Mass, RT and Spectra | 19543.35509 | 346205.9166 | 391932.1366 | 63584.44482 | 65633.45111 | 73253.6401 |
| 39 | C00106 | 111.0201052 | 1509667.119 | 1047.343906 | 1043.707212 | 1057.243086 | C00106 | Uracil | Mass, RT and Spectra | 2481306.74 | 7580125.607 | 7065366.92 | 3580972.534 | 4530176.98 | 3719586.441 |
| 40 | C00109 | 305.1868109 | 13104.16888 | 100.4589335 | 97.24515696 | 102.3926485 | C00109 | 2-Ketobutyric acid | Mass, RT and Spectra | 18941.26784 | 254901.2243 | 288407.1596 | 24722.03205 | 48932.01097 | 56060.77889 |
| 41 | C00110 | 786.5976821 | 46684835.02 | 1079.151134 | 1073.487364 | 1083.053839 | C00110 | Dolichol phosphate | Mass, RT and Spectra | 10961.06752 | 189496.7557 | 149121.0477 | 21159.76154 | 26700.70317 | 49352.75549 |
| 42 | C00111 | 168.9903195 | 463384.3707 | 422.600512 | 417.6472858 | 444.0350777 | C00111 | Dihydroxyacetone phosphate | Mass, RT and Spectra | 102981.3527 | 955473.5934 | 741604.101 | 177515.6538 | 247831.3703 | 209408.8603 |
| 43 | C05462 | 448.306529 | 16084615.48 | 108.2887748 | 96.39602107 | 120.340192 | C05462 | Chenodeoxyglycocholic acid | Mass, RT and Spectra | 3711888.768 | 31945674.48 | 25066622.71 | 6456817.414 | 5661469.03 | 9421837.619 |
| 44 | C00118 | 149.0084391 | 30130.99773 | 518.025898 | 514.6893931 | 527.2342865 | C00118 | D-Glyceraldehyde 3-phosphate | Mass, RT and Spectra | 303.6516354 | 59430.90688 | 67166.73767 | 1258.535658 | 12034.1316 | 11794.07606 |
| 45 | C05463 | 498.0534401 | 13393.02329 | 463.0304918 | 459.4025952 | 465.7543061 | C05463 | Taurodeoxycholic acid | Mass, RT and Spectra | 433.9208182 | 24324.04887 | 32906.40491 | 3545.05647 | 3623.295656 | 2646.218455 |
| 46 | C16613 | 185.0115018 | 15939.60543 | 236.3809824 | 232.7308497 | 245.9156382 | C16613 | 6-Thiourate | Mass, RT | 48.05537017 | 27416.92448 | 42312.11321 | 28.55094405 | 284.9197794 | 2344.834698 |
| 47 | C00121 | 149.044848 | 174135.0876 | 263.4667916 | 250.956552 | 283.0792375 | C00121 | D-Ribose | Mass, RT and Spectra | 20390.68096 | 364497.7252 | 301929.1473 | 34920.34536 | 69059.41396 | 71672.14456 |
| 48 | C00122 | 115.0032544 | 459439.9828 | 472.7285688 | 468.6032313 | 475.4195847 | C00122 | Fumaric acid | Mass, RT and Spectra | 118977.1945 | 743481.6514 | 874748.1556 | 193062.1242 | 217793.6078 | 205288.5253 |
| 49 | C00123 | 130.0869036 | 5249732.254 | 231.5879076 | 186.0665667 | 246.381373 | C00123 | L-Leucine | Mass, RT and Spectra | 565104.8307 | 10331245.01 | 11470228.59 | 1071154.38 | 1671933.076 | 1537151.937 |
| 50 | C00129 | 247.0154615 | 1257057.371 | 137.4427134 | 131.8112165 | 144.0206862 | C00129 | Isopentenyl pyrophosphate | Mass, RT and Spectra | 1517598.01 | 826610.1326 | 1209536.163 | 1580676.041 | 1332774.927 | 1477367.942 |
| 51 | C05464 | 450.3201611 | 50023601.79 | 736.039593 | 732.4954711 | 740.4330452 | C05464 | Deoxycholic acid glycine conjugate | Mass, RT | 44164567.44 | 60608277.11 | 49062966.7 | 43554835.32 | 35603371.58 | 40093266.56 |
| 52 | C00131 | 411.8999911 | 8683.377037 | 315.0632951 | 310.4129259 | 323.1153591 | C00131 | Deoxyadenosine triphosphate | Mass, RT and Spectra | 689.957067 | 17734.86906 | 19499.73063 | 551.8497574 | 1818.263677 | 2019.523948 |
| 53 | C05465 | 248.640385 | 5020.355397 | 106.7965279 | 91.15495716 | 108.3160205 | C05465 | Tetrahydrocortisol | Mass, RT and Spectra | 82.75733098 | 13079.4758 | 7918.020909 | 3480.744928 | 2800.372035 | 3508.36282 |
| 54 | C00135 | 154.0616826 | 1977040.35 | 398.3220452 | 388.443189 | 403.9976984 | C00135 | L-Histidine | Mass, RT and Spectra | 413888.653 | 3609832.461 | 3852996.549 | 561547.6417 | 839631.1914 | 841888.1857 |
| 55 | C05466 | 448.2311811 | 7950.97866 | 107.2655452 | 105.2684894 | 108.9013811 | C05466 | Chenodeoxycholic acid glycine conjugate | Mass, RT and Spectra | 550.7687606 | 19871.98211 | 12746.97045 | 1379.422198 | 504.50844 | 2376.526696 |
| 56 | C00140 | 256.0589057 | 240698.4794 | 342.6215502 | 328.1716733 | 358.6695927 | C00140 | N-Acetyl-D-glucosamine | Mass, RT and Spectra | 40228.652 | 409772.2799 | 522148.0003 | 94526.44367 | 100178.301 | 82679.61882 |
| 57 | C00141 | 115.0396398 | 2112273.864 | 113.5912271 | 84.49231391 | 129.915626 | C00141 | Alpha-ketoisovaleric acid | Mass, RT and Spectra | 547655.565 | 3790981.277 | 4128433.298 | 658834.548 | 753885.5359 | 1037150.808 |
| 58 | C00147 | 134.0467917 | 9214.674435 | 208.2469653 | 200.4470372 | 217.2967024 | C00147 | Adenine | Mass, RT and Spectra | 534.3132949 | 23200.36881 | 13789.80802 | 901.0734145 | 19454.14899 | 20033.42423 |
| 59 | C05471 | 365.2358452 | 66332.69312 | 647.6109601 | 646.1659162 | 648.5526022 | C05471 | Dihydrocortisol | Mass, RT | 1390.304929 | 1079.284274 | 1087.76519 | 1208.604473 | 4015.203078 | 9991.957922 |
| 60 | C05476 | 174.1152832 | 11931.46099 | 101.1384471 | 98.54747242 | 103.5592107 | C05476 | Tetrahydrocorticosterone | Mass, RT and Spectra | 296.4750613 | 23742.5446 | 28076.4434 | 569.3398498 | 937.5924005 | 3643.487871 |
| 61 | C00148 | 114.0556715 | 1870924.532 | 356.670634 | 342.8524141 | 371.698104 | C00148 | L-Proline | Mass, RT and Spectra | 157290.489 | 4135081.929 | 3706175.876 | 262547.6573 | 557886.6656 | 474314.5263 |
| 62 | C00149 | 133.0136842 | 68796.37992 | 353.7741348 | 339.7063762 | 356.9665488 | C00149 | L-Malic acid | Mass, RT and Spectra | 2551.693833 | 152918.6429 | 122513.3475 | 16226.82293 | 41445.67531 | 35582.48891 |
| 63 | C00152 | 133.0603441 | 1265314.736 | 102.3138554 | 97.95714976 | 107.7450122 | C00152 | L-Asparagine | Mass, RT and Spectra | 1384261.516 | 1084207.456 | 1080448.502 | 1456649.857 | 1733590.512 | 1602861.895 |
| 64 | C05504 | 465.2090785 | 159181.6885 | 751.9260328 | 746.9227759 | 761.7039591 | C05504 | Estriol-16-Glucuronide | Mass, RT and Spectra | 168975.975 | 146682.9632 | 146256.4541 | 163096.5931 | 170394.7412 | 156270.5979 |
| 65 | C05512 | 253.0920881 | 25982.32407 | 522.7263947 | 517.9112603 | 527.226405 | C05512 | Deoxyinosine | Mass, RT and Spectra | 13407.6057 | 26426.41042 | 11468.75362 | 112965.6267 | 49368.56529 | 85328.98804 |
| 66 | C00157 | 734.5677121 | 3665030.336 | 1160.228807 | 1151.209288 | 1165.71224 | C00157 | PC(16:0/16:0) | Mass, RT and Spectra | 4544764.332 | 1878944.559 | 2780350.231 | 4983564.221 | 4180896.927 | 5887690.454 |
| 67 | C05516 | 128.0453045 | 138507.2397 | 1583.899537 | 1578.08476 | 1587.259611 | C05516 | 5-Amino-4-imidazole carboxylate | Mass, RT and Spectra | 155643.6062 | 117113.0162 | 115751.5867 | 150612.0557 | 154310.5514 | 143922.1461 |
| 68 | C00158 | 111.0084213 | 491354.233 | 551.6205383 | 542.3253725 | 558.658131 | C00158 | Citric acid | Mass, RT and Spectra | 150015.0375 | 890165.6304 | 843379.8753 | 211172.6068 | 227958.6775 | 217308.4013 |
| 69 | C17467 | 121.0291547 | 144800.2505 | 133.9529818 | 107.8311139 | 156.3354769 | C17467 | 4-Hydroxybenzyl alcohol | Mass, RT and Spectra | 25814.95021 | 334329.8193 | 225968.3087 | 29297.04449 | 50078.51716 | 43532.99042 |
| 70 | C00160 | 75.00842328 | 840611.8473 | 143.6865942 | 111.6091838 | 160.8960091 | C00160 | Glycolate | Mass, RT and Spectra | 139137.3294 | 1486832.738 | 1650679.955 | 160453.1949 | 230290.5716 | 354650.4782 |
| 71 | C05547 | 325.161618 | 222758.3491 | 637.7803396 | 634.9612059 | 638.7597434 | C05547 | Galactosylhydroxylysine | Mass, RT and Spectra | 22132.03018 | 2026.787279 | 1648.85692 | 23224.68284 | 12282.14748 | 14034.5434 |
| 72 | C00166 | 135.0447398 | 34387.61296 | 132.4350758 | 122.3742511 | 141.0951078 | C00166 | Phenylpyruvic acid | Mass, RT and Spectra | 1575.266765 | 78993.58182 | 68362.45124 | 2493.728057 | 8230.586411 | 8566.867348 |
| 73 | C00167 | 579.0272677 | 1512774.952 | 513.7576211 | 508.437103 | 534.6825406 | C00167 | Uridine diphosphate glucuronic acid | Mass, RT and Spectra | 68168.43695 | 52861.91517 | 59895.76524 | 19773.42118 | 15923.14111 | 2780.559354 |
| 74 | C00169 | 139.9750441 | 2228458.331 | 488.7209827 | 485.6228915 | 491.9228809 | C00169 | Carbamoyl phosphate | Mass, RT and Spectra | 227611.0133 | 4539963.358 | 4008059.373 | 504650.6246 | 943611.8479 | 1180315.837 |
| 75 | C00170 | 296.0844035 | 10547.47361 | 514.2739906 | 511.7511732 | 517.7842242 | C00170 | 5'-Methylthioadenosine | Mass, RT and Spectra | 556.0206396 | 21477.42433 | 48793.78643 | 1984.551418 | 10856.7842 | 11506.24623 |
| 76 | C05560 | 491.1249978 | 15157.10169 | 889.863921 | 885.8551457 | 890.687338 | C05560 | L-2-Aminoadipate adenylate | Mass, RT and Spectra | 13816.68204 | 9963.008055 | 23889.28749 | 3106.222032 | 1599.091728 | 4467.454639 |
| 77 | C00178 | 125.0346363 | 8591.101241 | 178.7331237 | 174.6108777 | 190.8241372 | C00178 | Thymine | Mass, RT and Spectra | 311.21893 | 19572.47457 | 18459.21763 | 128.001799 | 2175.085549 | 2144.412971 |
| 78 | C00180 | 121.0297163 | 65990.57828 | 1162.951692 | 1157.625431 | 1182.784616 | C00180 | 3,5-Dihydroxybenzoic acid | Mass, RT and Spectra | 30.87215686 | 26051.20527 | #DIV/0! | 198.7117741 | 396.2323525 | 2201.443946 |
| 79 | C00183 | 117.0746935 | 249958.3571 | 297.7977524 | 272.3265243 | 320.8501546 | C00183 | L-Valine | Mass, RT and Spectra | 28442.9071 | 487749.1109 | 542505.2302 | 52264.7824 | 93551.2151 | 113107.2187 |
| 80 | C17938 | 191.0214262 | 1610653.603 | 223.2473658 | 217.8528978 | 231.0319161 | C17938 | 5,6-Indolequinone-2-carboxylic acid | Mass, RT and Spectra | 24516.81631 | 10079.57447 | 7251.602183 | 24864.97522 | 27303.66673 | 26016.34152 |
| 81 | C00187 | 387.36137 | 765027.5235 | 12.31053663 | 5.289900972 | 20.51470162 | C00187 | Cholesterol | Mass, RT and Spectra | 121465.5037 | 1559399.174 | 1410643.979 | 149334.9698 | 374805.5481 | 249192.3769 |
| 82 | C00191 | 193.0347833 | 1649539.777 | 490.729065 | 489.1770436 | 493.5553512 | C00191 | D-Glucuronic acid | Mass, RT and Spectra | 441247.0133 | 2927501.297 | 2059196.512 | 983597.0848 | 1301852.827 | 1483686.521 |
| 83 | C00195 | 538.5183954 | 167971.8816 | 1254.439273 | 1202.464734 | 1280.78894 | C00195 | Ceramide (d18:1/12:0) | Mass, RT and Spectra | 274353.4823 | 180841.3726 | 223074.3975 | 241508.2836 | 252398.0702 | 182399.0827 |
| 84 | C00197 | 186.9998001 | 19589.04956 | 85.42378577 | 83.32231852 | 90.15228149 | C00197 | 3-Phosphoglyceric acid | Mass, RT and Spectra | 19217.89199 | 20009.18345 | 21482.36663 | 15556.23865 | 18452.29072 | 36227.75566 |
| 85 | C00198 | 177.0397809 | 294036.5918 | 379.0183128 | 368.7184008 | 390.9471771 | C00198 | Gluconolactone | Mass, RT and Spectra | 55442.43647 | 494362.0376 | 474424.0289 | 137558.4763 | 319818.9304 | 297381.926 |
| 86 | C00209 | 88.98772707 | 1008068.243 | 528.0848721 | 523.4172439 | 532.3128287 | C00209 | Oxalic acid | Mass, RT and Spectra | 85653.83598 | 2038182.079 | 2276510.005 | 144430.0178 | 282302.2591 | 360454.3252 |
| 87 | C05565 | 171.0404434 | 19649.80704 | 464.5574195 | 462.1712641 | 482.2955589 | C05565 | Hydantoin-5-propionic acid | Mass, RT and Spectra | 74685.43174 | 50642.8461 | 30175.69006 | 183776.3029 | 402671.8511 | 901035.1619 |
| 88 | C00212 | 268.1022644 | 14687.55359 | 92.92039999 | 83.18024821 | 99.85620735 | C00212 | Adenosine | Mass, RT and Spectra | 28907.20317 | 4473.447767 | 12199.11311 | 27701.55664 | 10484.03832 | 15430.28484 |
| 89 | C00213 | 88.04008926 | 4465987.983 | 409.6851004 | 386.2449239 | 414.4233934 | C00213 | Sarcosine | Mass, RT and Spectra | 244166.5489 | 9599721.152 | 9806584.497 | 569725.3007 | 1409046.83 | 1466945.289 |
| 90 | C00214 | 120.0378893 | 31143.23988 | 283.1977664 | 269.7935853 | 288.7326206 | C00214 | Thymidine | Mass, RT | 2298.029603 | 52383.55303 | 53164.57197 | 8890.406991 | 37539.86761 | 56573.67503 |
| 91 | C00216 | 149.0449286 | 392540.1844 | 379.5152133 | 360.2510859 | 396.2124271 | C00216 | D-Arabinose | Mass, RT and Spectra | 95552.96767 | 575496.6147 | 985628.6901 | 100130.7044 | 210818.0008 | 264283.6519 |
| 92 | C05576 | 171.0659822 | 341521.494 | 1553.280272 | 1550.986381 | 1555.863801 | C05576 | 3,4-Dihydroxyphenylglycol | Mass, RT and Spectra | 252490.708 | 387128.8808 | 421633.1414 | 279420.3096 | 233190.7475 | 304499.4195 |
| 93 | C05579 | 146.0242462 | 15299.72059 | 118.7915242 | 115.1668683 | 120.7442964 | C05579 | 19-Hydroxytestosterone | Mass, RT and Spectra | 933.5077336 | 28543.63646 | 27306.97255 | 6734.33504 | 1229.709098 | 2353.906594 |
| 94 | C00217 | 146.0453124 | 120210.857 | 339.6151918 | 333.9045396 | 358.5223265 | C00217 | D-Glutamic acid | Mass, RT and Spectra | 106715506.6 | 109905481.9 | 108309718.8 | 101331436 | 90254856.64 | 103879009.1 |
| 95 | C00219 | 303.2320552 | 1505151.665 | 91.18586851 | 90.13643798 | 92.27870473 | C00219 | Arachidonic acid | Mass, RT and Spectra | 284211.7123 | 2447641.956 | 2466226.024 | 492140.639 | 617055.2658 | 496299.8171 |
| 96 | C05582 | 183.0659412 | 169870.4826 | 1553.176491 | 1550.351421 | 1556.535646 | C05582 | Homovanillic acid | Mass, RT and Spectra | 107690.4705 | 222213.8694 | 222202.4865 | 135179.9068 | 150868.3413 | 132205.9797 |
| 97 | C18043 | 467.3183811 | 2977550.934 | 664.8129981 | 662.9016425 | 665.5050448 | C18043 | Cholesterol sulfate | Mass, RT and Spectra | 97.08279193 | 2758.36245 | 6045.650454 | 619.9720774 | 7541.945999 | 54980.48149 |
| 98 | C00233 | 131.0701786 | 184228.4416 | 15.19489195 | 10.25076382 | 19.98982184 | C00233 | Ketoleucine | Mass, RT and Spectra | 149700.7672 | 223787.6845 | 221624.8455 | 150266.6516 | 162351.069 | 163689.9265 |
| 99 | C05588 | 198.1140081 | 22896.90144 | 11.79858402 | 4.720976144 | 16.35515915 | C05588 | Metanephrine | Mass, RT and Spectra | 16094.97821 | 33856.86481 | 31736.03326 | 9249.33391 | 28749.62505 | 13857.3808 |
| 100 | C00245 | 124.006974 | 1147740.728 | 400.7854918 | 392.0852001 | 408.8604504 | C00245 | Taurine | Mass, RT and Spectra | 278700.2918 | 1490532.604 | 2199905.099 | 449260.0831 | 556943.2834 | 482033.5129 |
| 101 | C00248 | 333.1335898 | 14368.1514 | 103.8483623 | 102.7045593 | 104.2141033 | C00248 | Lipoamide | Mass, RT and Spectra | 15628.98626 | 18422.46587 | 19608.59485 | 17680.91874 | 16694.47529 | 18738.34835 |
| 102 | C16614 | 167.0370017 | 9649.00435 | 96.21406937 | 90.24447071 | 102.7203787 | C16614 | 6-Methylthiopurine | Mass, RT and Spectra | 762.1152156 | 19445.46796 | 21431.46188 | 3180.448208 | 3310.322301 | 3423.251391 |
| 103 | C00249 | 255.2323893 | 28547075.64 | 93.32761465 | 92.66187071 | 93.99489178 | C00249 | Palmitic acid | Mass, RT and Spectra | 4948760.886 | 47332803.51 | 52533545.57 | 10478857.45 | 9373298.489 | 11987108.76 |
| 104 | C00250 | 168.0653119 | 122816.1999 | 289.766601 | 283.0501352 | 304.71822 | C00250 | Pyridoxal | Mass, RT and Spectra | 339424.9388 | 44209.81341 | 38921.93452 | 292924.9007 | 320691.1997 | 27441.89459 |
| 105 | C05635 | 190.0505333 | 7565.755239 | 207.7821041 | 203.3040762 | 219.0352271 | C05635 | 5-Hydroxyindoleacetic acid | Mass, RT and Spectra | 647.7149258 | 17635.83175 | 16674.27477 | 480.2866181 | 8201.118215 | 5736.190404 |
| 106 | C00256 | 91.03815673 | 952164.2814 | 236.7466843 | 227.0866951 | 244.1500946 | C00256 | D-Lactic acid | Mass, RT | 85617.98071 | 1541318.872 | 2074042.444 | 104519.9179 | 249182.3436 | 368410.7006 |
| 107 | C00257 | 195.0503603 | 708224.4386 | 426.2799348 | 416.5432259 | 439.6531879 | C00257 | Gluconic acid | Mass, RT and Spectra | 115998.4546 | 1212568.606 | 1244735.546 | 414670.5323 | 526819.7319 | 688916.0551 |
| 108 | C00258 | 105.0189381 | 144225.8914 | 333.4282809 | 312.8533288 | 340.4572159 | C00258 | Glyceric acid | Mass, RT and Spectra | 22378.41825 | 251845.1197 | 283128.299 | 36769.07518 | 52834.36175 | 56514.25719 |
| 109 | C00262 | 135.0307898 | 523198.1001 | 255.5637207 | 236.6240966 | 267.3985973 | C00262 | Hypoxanthine | Mass, RT and Spectra | 124260.2232 | 652116.4187 | 1252037.906 | 104762.9233 | 93844.12554 | 157169.5237 |
| 110 | C00263 | 118.0505633 | 2823405.162 | 390.2076756 | 376.9285664 | 397.8918848 | C00263 | L-Homoserine | Mass, RT and Spectra | 278522.4054 | 5865204.34 | 5953522.945 | 540955.6534 | 929199.5951 | 1125938.648 |
| 111 | C00281 | 291.0457381 | 16775.77055 | 105.5189252 | 100.0727832 | 106.6459986 | C00281 | D-Sedoheptulose 7-phosphate | Mass, RT and Spectra | 10498.52498 | 530.6094377 | 218.4076581 | 14494.6765 | 22309.97688 | 31114.68405 |
| 112 | C18319 | 147.0657826 | 43443.91847 | 161.5967474 | 153.6892341 | 173.9530638 | C18319 | Ethylmethylacetic acid | Mass, RT and Spectra | 14260.52812 | 74316.06669 | 83852.92627 | 8839.077264 | 32589.4178 | 24392.53285 |
| 113 | C00292 | 94.06509264 | 1399837.404 | 1553.173198 | 1550.812659 | 1555.417283 | C00292 | Aniline | Mass, RT and Spectra | 1209261.524 | 1511355.807 | 1688144.782 | 1239272.749 | 1293581.374 | 1180359.352 |
| 114 | C00294 | 267.073086 | 17286.72021 | 293.6707441 | 276.4418658 | 299.819659 | C00294 | Inosine | Mass, RT and Spectra | 47688.08747 | 14304.82833 | 28387.29298 | 731.463776 | 614.3649459 | 734.318661 |
| 115 | C05636 | 181.0964248 | 38356.93198 | 542.6769007 | 541.4160171 | 545.4580757 | C05636 | 3-Hydroxykynurenamine | Mass, RT and Spectra | 19409.68512 | 38599.77691 | 16611.55662 | 122116.8309 | 10627.98361 | 8201.319043 |
| 116 | C16660 | 263.1677061 | 1654488.396 | 488.5015712 | 483.5085285 | 493.7979385 | C16660 | 2-Ethyl-5-methyl-3,3-diphenyl-1-pyrroline | Mass, RT and Spectra | 25971.78056 | 8605.617121 | 9661.318648 | 27773.53493 | 35242.67635 | 50298.84939 |
| 117 | C00295 | 155.0085959 | 9108.49945 | 471.3634687 | 468.3287169 | 473.36353 | C00295 | Orotic acid | Mass, RT and Spectra | 1668.96242 | 19062.9535 | 14392.69955 | 2286.512016 | 1280.423502 | 4401.082907 |
| 118 | C00296 | 191.0554323 | 87706.85089 | 389.6297935 | 383.3428361 | 397.6725788 | C00296 | Quinic acid | Mass, RT and Spectra | 7193.467096 | 125933.4498 | 205495.6079 | 14032.37563 | 49318.73001 | 56922.50084 |
| 119 | C05642 | 263.1034778 | 43079.39529 | 113.5426982 | 102.3551299 | 119.3650096 | C05642 | Acetyl-N-formyl-5-methoxykynurenamine | Mass, RT | 9785.239439 | 71652.90307 | 96495.75031 | 18136.84752 | 20616.42003 | 37835.6818 |
| 120 | C05645 | 223.0461008 | 10098.27766 | 147.5927719 | 144.1112868 | 157.611041 | C05645 | 4-(2-Amino-3-hydroxyphenyl)-2,4-dioxobutanoic acid | Mass, RT and Spectra | 71.83016425 | 32283.28397 | 21710.78066 | 198.9545727 | 7165.338528 | 14422.71138 |
| 121 | C00299 | 245.074621 | 41979.71275 | 100.562712 | 93.23590308 | 117.4200143 | C00299 | Uridine | Mass, RT and Spectra | 63006.79261 | 27781.2722 | 21627.53717 | 44742.06523 | 56933.21139 | 56705.99678 |
| 122 | C00300 | 132.076477 | 3245281.1 | 125.7979666 | 120.7688228 | 139.9762459 | C00300 | Creatine | Mass, RT and Spectra | 4706418.798 | 1240632.055 | 1587516.88 | 2799726.251 | 5291165.103 | 12058882.08 |
| 123 | C00311 | 191.0192505 | 6262177.766 | 552.005856 | 541.9480855 | 558.9659655 | C00311 | Isocitric acid | Mass, RT and Spectra | 1124671.695 | 12325910.35 | 11287800.9 | 2212749.088 | 2011349.114 | 1930904.882 |
| 124 | C00315 | 146.1649716 | 64993.0434 | 1774.940747 | 1747.568327 | 1794.481088 | C00315 | Spermidine | Mass, RT and Spectra | 54942.04669 | 68910.26371 | 75482.57671 | 60421.3888 | 54254.8088 | 43436.85253 |
| 125 | C05655 | 226.0352082 | 28837.91361 | 199.9945607 | 191.0371863 | 213.4520507 | C05655 | 5-(2'-Carboxyethyl)-4,6-Dihydroxypicolinate | Mass, RT and Spectra | 3693.826898 | 46170.29544 | 82666.57586 | 1628.794384 | 3574.042956 | 4433.287927 |
| 126 | C05656 | 254.0297753 | 27048.93696 | 448.9714244 | 445.072824 | 451.5475301 | C05656 | 5-(3'-Carboxy-3'-oxopropyl)-4,6-dihydroxypicolinate | Mass, RT and Spectra | 1178.505975 | 66628.29228 | 51313.49069 | 4577.476762 | 13782.64712 | 13121.33077 |
| 127 | C00318 | 198.0710585 | 68911.57669 | 394.6634027 | 375.536639 | 398.6456984 | C00318 | L-Carnitine | Mass, RT and Spectra | 8097.779473 | 140398.4862 | 126856.9638 | 15513.80088 | 31648.65679 | 23593.12683 |
| 128 | C00319 | 300.2886959 | 138707.4464 | 974.7237406 | 953.225894 | 979.128208 | C00319 | Sphingosine | Mass, RT and Spectra | 164017.3141 | 85807.49283 | 103401.4168 | 133596.5101 | 136901.9731 | 176926.0574 |
| 129 | C05658 | 134.059818 | 17214.99031 | 448.9109183 | 447.9956265 | 451.1746 | C05658 | Indoxyl | Mass, RT and Spectra | 1869652.205 | 612462.4059 | 633724.1672 | 1743247.899 | 1578132.463 | 1583923.165 |
| 130 | C00327 | 174.0878288 | 455387.9142 | 444.716574 | 442.5026063 | 447.3466377 | C00327 | Citrulline | Mass, RT and Spectra | 63533.83189 | 787786.9616 | 1078420.126 | 76143.39243 | 110316.0176 | 129570.3218 |
| 131 | C00328 | 207.0784448 | 51436.55866 | 207.3040667 | 196.2460273 | 221.747818 | C00328 | L-Kynurenine | Mass, RT and Spectra | 3336.311202 | 76468.3229 | 110360.1852 | 2968.930051 | 11895.19708 | 55571.51768 |
| 132 | C05659 | 189.103469 | 9705.626188 | 93.57707871 | 92.10219882 | 102.983278 | C05659 | 5-Methoxytryptamine | Mass, RT and Spectra | 586.98043 | 15234.42821 | 24344.10593 | 1743.846075 | 1150.971277 | 2849.563535 |
| 133 | C00334 | 102.0556973 | 27549.99748 | 158.5168407 | 153.7712719 | 164.7321722 | C00334 | Gamma-Aminobutyric acid | Mass, RT and Spectra | 444.8348245 | 73016.74212 | 46239.48431 | 2840.214822 | 8468.042954 | 8767.190001 |
| 134 | C05660 | 101.5293437 | 1876537.305 | 236.3159792 | 220.8165018 | 253.3436387 | C05660 | 5-Methoxyindoleacetate | Mass, RT and Spectra | 37551.22694 | 146790.4346 | 138196.7287 | 51074.89227 | 65041.12503 | 63325.85796 |
| 135 | C19615 | 287.2207558 | 17455.04981 | 855.8425374 | 851.829871 | 860.3111832 | C19615 | Hexadecanedioic acid | Mass, RT and Spectra | 16457.47942 | 13299.23068 | 9915.715503 | 30242.62013 | 13651.88989 | 11238.57542 |
| 136 | C05673 | 430.064872 | 1982971.471 | 526.436578 | 519.965458 | 530.3322574 | C05673 | CMPciliatine | Mass, RT and Spectra | 318870.0555 | 127783.9541 | 125299.5088 | 144717.6938 | 188561.7488 | 166553.7145 |
| 137 | C00337 | 157.026724 | 9103.070114 | 296.0144707 | 292.783796 | 302.3844636 | C00337 | 4,5-Dihydroorotic acid | Mass, RT and Spectra | 235.8128892 | 15973.68384 | 13137.77936 | 224.3006336 | 3761.029556 | 6192.128043 |
| 138 | C05683 | 516.3104231 | 33864.34214 | 700.3639637 | 694.6804561 | 703.1167686 | C05683 | Ciliatocholate | Mass, RT and Spectra | 40609.37773 | 21612.58102 | 29289.31079 | 32010.0598 | 42526.01131 | 41908.37561 |
| 139 | C00345 | 277.0324368 | 347301.7062 | 422.1149099 | 418.455859 | 426.7975901 | C00345 | 6-Phosphogluconic acid | Mass, RT | 77174.30865 | 754913.3146 | 533778.6389 | 136769.1749 | 185903.8394 | 138684.6884 |
| 140 | C00346 | 142.0260872 | 117309.2282 | 91.86032461 | 87.57508422 | 95.22636308 | C00346 | O-Phosphoethanolamine | Mass, RT and Spectra | 203909.7661 | 54570.0049 | 77140.02429 | 156248.8211 | 186503.6633 | 145107.8014 |
| 141 | C00352 | 260.0523086 | 126886.7856 | 401.3170699 | 385.0445825 | 403.5590265 | C00352 | Glucosamine 6-phosphate | Mass, RT and Spectra | 19.67521309 | 35339.26399 | 33110.60658 | 838.30658 | 1762.643932 | 5109.919125 |
| 142 | C00355 | 198.0766948 | 275034.4174 | 1553.287264 | 1550.187437 | 1555.275312 | C00355 | L-Dopa | Mass, RT and Spectra | 213244.7941 | 309151.9202 | 355521.7606 | 181950.4401 | 220352.7677 | 252902.008 |
| 143 | C00362 | 346.0547889 | 31252.75772 | 441.982584 | 439.323713 | 443.7748508 | C00362 | 2'-Deoxyguanosine 5'-monophosphate | Mass, RT and Spectra | 21837.73782 | 8453.09985 | 10376.16349 | 28912.58194 | 14753.52924 | 17831.3884 |
| 144 | C00364 | 322.05761 | 2013010.875 | 452.928898 | 450.0654499 | 455.9731905 | C00364 | 5-Thymidylic acid | Mass, RT and Spectra | 15363.64429 | 10677.1091 | 6265.492619 | 6394.678055 | 7064.787569 | 4013.5905 |
| 145 | C00366 | 167.0204021 | 13286244.13 | 380.739087 | 366.4561626 | 392.3173071 | C00366 | Uric acid | Mass, RT and Spectra | 2852123.796 | 22723799.19 | 26203349.9 | 4149139.367 | 6821958.963 | 6842717.218 |
| 146 | C00378 | 266.1211572 | 86541.11794 | 454.2262085 | 449.9456221 | 455.9731905 | C00378 | Thiamine | Mass, RT and Spectra | 119898.2348 | 41185.7521 | 53369.11552 | 100334.5993 | 102103.024 | 332374.0303 |
| 147 | C05689 | 183.9877045 | 1406062.885 | 1740.508843 | 1717.726067 | 1746.653743 | C05689 | Se-Methylselenocysteine | Mass, RT and Spectra | 83631.4264 | 389.02817 | 204.1338642 | 111919.5986 | 73164.3883 | 63493.5677 |
| 148 | C00380 | 112.050418 | 119140.9644 | 12.09630912 | 7.604087311 | 15.54073971 | C00380 | Cytosine | Mass, RT and Spectra | 56690.19197 | 181908.7462 | 168226.7329 | 66946.17537 | 101154.5657 | 116022.7307 |
| 149 | C05695 | 311.0159378 | 64134.36131 | 571.1405561 | 557.4474755 | 585.0815209 | C05695 | Gamma-Glutamyl-Se-methylselenocysteine | Mass, RT and Spectra | 5448.611058 | 134955.6935 | 151028.509 | 8247.596199 | 17461.59272 | 14487.31731 |
| 150 | C00385 | 151.0255439 | 208811.0299 | 348.9835871 | 332.1133398 | 354.7674672 | C00385 | Xanthine | Mass, RT and Spectra | 23893.52628 | 267666.3588 | 609096.9856 | 29291.69345 | 42041.77254 | 57346.89193 |
| 151 | C05699 | 271.021712 | 4713.581519 | 346.8529062 | 344.1810311 | 350.5155 | C05699 | Selenocystathionine | Mass, RT and Spectra | 18511.34559 | 5270.403199 | 6704.713642 | 23946.87059 | 9658.401527 | 12309.1128 |
| 152 | C19691 | 324.2014752 | 2076366.198 | 404.6501723 | 398.0969482 | 408.8661112 | C19691 | Farnesylcysteine | Mass, RT and Spectra | 24384.63494 | 12540.38078 | 11453.5153 | 21594.9807 | 23189.84613 | 21458.53571 |
| 153 | C05787 | 935.318536 | 2118610.686 | 655.4319793 | 647.7224726 | 666.2566626 | C05787 | Bilirubin diglucuronide | Mass, RT and Spectra | 68144.07007 | 25591.77477 | 18368.44738 | 114946.495 | 127555.1284 | 121037.9035 |
| 154 | C00386 | 227.1034738 | 7966.866429 | 428.0285964 | 423.096881 | 430.2867164 | C00386 | Carnosine | Mass, RT and Spectra | 604.9059196 | 13588.54604 | 15237.09634 | 3003.199256 | 3517.922045 | 2193.093479 |
| 155 | C00388 | 112.0868572 | 5781432.558 | 1556.491567 | 1546.593101 | 1570.025179 | C00388 | Histamine | Mass, RT and Spectra | 6660447.502 | 4885851.417 | 5118259.53 | 5999958.921 | 5707080.697 | 6234325.507 |
| 156 | C00392 | 183.0865622 | 1116545.174 | 93.10812496 | 90.06093217 | 97.79175891 | C00392 | Mannitol | Mass, RT and Spectra | 1617967.925 | 253589.0065 | 244071.8972 | 1855084.539 | 3167356.271 | 4714574.913 |
| 157 | C00396 | 81.04480503 | 104563565.8 | 1553.146583 | 1551.896761 | 1555.00951 | C00396 | Pyrimidine | Mass, RT and Spectra | 8117.051059 | 82517.56164 | 96254.69373 | 5707.102411 | 15722.11873 | 14985.34087 |
| 158 | C00398 | 161.1069851 | 185450.2461 | 650.6896 | 637.3061442 | 665.5738852 | C00398 | Tryptamine | Mass, RT and Spectra | 92760.90914 | 283581.6121 | 281550.566 | 119486.3016 | 154354.448 | 130251.5491 |
| 159 | C00408 | 128.0712664 | 179671.8698 | 335.7141476 | 319.3364401 | 348.0465639 | C00408 | Pipecolic acid | Mass, RT and Spectra | 10813.07764 | 391798.2388 | 335251.1925 | 33845.90703 | 59158.94149 | 78275.90933 |
| 160 | C05789 | 597.3705429 | 42867.78879 | 1080.530741 | 1069.287427 | 1087.485357 | C05789 | L-Urobilinogen | Mass, RT and Spectra | 59676.12879 | 93651.98367 | 30380.52662 | 95210.05417 | 43651.8077 | 68025.45245 |
| 161 | C00418 | 145.0500159 | 41301.33454 | 203.5824787 | 198.5841435 | 208.8296053 | C00418 | Mevalonic acid | Mass, RT and Spectra | 1363.890328 | 92951.53167 | 73990.55245 | 3311.479008 | 13992.85484 | 17228.01921 |
| 162 | C05791 | 591.3159952 | 176946.934 | 714.8020035 | 696.4931264 | 726.2246479 | C05791 | D-Urobilinogen | Mass, RT and Spectra | 126371.8628 | 134356.1745 | 105971.1414 | 459196.5313 | 43783.02661 | 673829.7148 |
| 163 | C00429 | 113.0352785 | 751829.3707 | 185.3332061 | 163.9207157 | 200.5032258 | C00429 | Dihydrouracil | Mass, RT and Spectra | 142667.6348 | 1423909.413 | 1459442.517 | 140444.9523 | 368607.9427 | 357168.7938 |
| 164 | C05793 | 595.3541574 | 12284.47614 | 772.9773194 | 757.5698159 | 776.4946676 | C05793 | L-Urobilin | Mass, RT and Spectra | 15259.73979 | 4785.997078 | 5424.10444 | 16349.5127 | 11540.27323 | 13204.71614 |
| 165 | C00431 | 118.0848184 | 325533.3055 | 593.3298105 | 588.7975649 | 595.889297 | C00431 | 5-Aminopentanoic acid | Mass, RT and Spectra | 336784.1139 | 309716.4238 | 293627.7974 | 357543.4287 | 457975.9033 | 363549.1365 |
| 166 | C00433 | 129.0189683 | 806366.5842 | 551.5224579 | 542.1664532 | 559.2302761 | C00433 | 2,5-Dioxopentanoate | Mass, RT and Spectra | 208877.8037 | 1460400.969 | 1435916.715 | 359412.9958 | 398051.627 | 387424.0981 |
| 167 | C00437 | 173.0915396 | 42069.8964 | 379.4873163 | 361.1799037 | 398.4647726 | C00437 | N-Acetylornithine | Mass, RT and Spectra | 5915.172463 | 68527.68619 | 87847.64267 | 8269.190603 | 24421.38115 | 23335.98907 |
| 168 | C05795 | 589.3013667 | 68562.13025 | 739.7536184 | 724.133911 | 751.5508227 | C05795 | D-Urobilin | Mass, RT and Spectra | 50788.54114 | 23739.06154 | 21278.9631 | 34508.28067 | 25878.9788 | 33852.50053 |
| 169 | C05804 | 561.4255953 | 11391.69786 | 1254.077397 | 1244.23738 | 1260.225809 | C05804 | 2-Hexaprenyl-3-methyl-6-methoxy-1,4 benzoquinone | Mass, RT and Spectra | 18469.7709 | 2933.969679 | 6655.300917 | 9449.633916 | 11921.40817 | 5938.909032 |
| 170 | C00439 | 173.0561184 | 47822.4569 | 366.3228161 | 355.1706575 | 377.8469168 | C00439 | Formiminoglutamic acid | Mass, RT and Spectra | 2252.225334 | 77998.73676 | 92280.28277 | 9743.925048 | 36556.30616 | 36577.89887 |
| 171 | C00445 | 457.1738429 | 6394.400025 | 793.1149551 | 791.1769444 | 794.2228875 | C00445 | 5,10-Methenyltetrahydrofolic acid | Mass, RT and Spectra | 8480.261283 | 4851.254141 | 4704.136467 | 7679.995503 | 7871.085108 | 6346.791566 |
| 172 | C05809 | 683.5432328 | 2162.435913 | 1016.571776 | 1014.921802 | 1017.50191 | C05809 | 3-Octaprenyl-4-hydroxybenzoate | Mass, RT and Spectra | 5926.133566 | 169.0597687 | 94.07672177 | 597.2832063 | 125.8907778 | 4671.590811 |
| 173 | C00449 | 277.1401837 | 443134.358 | 821.9610906 | 817.7732349 | 824.4700177 | C00449 | Saccharopine | Mass, RT and Spectra | 341298.0165 | 537622.2259 | 562805.2695 | 314475.5098 | 350426.7033 | 290973.5687 |
| 174 | C19910 | 308.0990277 | 42311.78333 | 425.1571524 | 416.7768106 | 428.4067954 | C19910 | N-Acetylneuraminic acid | Mass, RT and Spectra | 6754.570233 | 77694.16973 | 71300.91971 | 22852.78355 | 33700.49 | 32276.47719 |
| 175 | C00450 | 126.0557958 | 10978.56148 | 150.3475643 | 145.8660347 | 156.5505414 | C00450 | (S)-2,3,4,5-Tetrahydropiperidine-2-carboxylate | Mass, RT and Spectra | 242.364615 | 23169.01542 | 24300.19898 | 585.8195395 | 4674.117559 | 5244.172217 |
| 176 | C00455 | 333.0497691 | 2235143.706 | 101.7420788 | 99.57822503 | 103.842136 | C00455 | Nicotinamide ribotide | Mass, RT and Spectra | 331985.6478 | 285505.1222 | 357268.9262 | 332694.4682 | 243240.0114 | 348895.0585 |
| 177 | C00463 | 118.0649886 | 984073.1538 | 1553.307217 | 1546.593101 | 1556.858152 | C00463 | Indole | Mass, RT and Spectra | 816019.971 | 930173.8922 | 1141745.741 | 1053362.767 | 785783.3093 | 986447.9119 |
| 178 | C00474 | 151.0604463 | 34040.56629 | 356.5117147 | 342.8524141 | 370.7783735 | C00474 | Ribitol | Mass, RT and Spectra | 2089.130367 | 66339.49973 | 61453.35721 | 13362.42542 | 19893.36531 | 22769.11376 |
| 179 | C00487 | 161.1072028 | 12621.19847 | 102.5893342 | 97.26549129 | 104.0418297 | C00487 | Malonyl-Carnitin | Mass, RT and Spectra | 401.2472495 | 24284.62694 | 30377.21721 | 1279.706058 | 1347.894971 | 4095.647687 |
| 180 | C00491 | 239.0158152 | 211076.0762 | 476.4990365 | 473.6868322 | 479.2491357 | C00491 | L-Cystine | Mass, RT and Spectra | 41767.07428 | 411327.3684 | 481295.6174 | 26979.55913 | 30525.02975 | 53911.34972 |
| 181 | C05817 | 241.0693462 | 7444551.012 | 433.1902978 | 426.7852753 | 436.9359956 | C05817 | (1R,6R)-6-Hydroxy-2-succinylcyclohexa-2,4-diene-1-carboxylate | Mass, RT and Spectra | 5719507.175 | 6248013.169 | 7332413.709 | 8746590.699 | 11732298.55 | 5256597.674 |
| 182 | C00493 | 175.0578599 | 116857.7359 | 96.54087208 | 91.67031403 | 100.9155504 | C00493 | Shikimic acid | Mass, RT and Spectra | 163798.966 | 72705.13997 | 66706.4532 | 164033.0747 | 189030.6382 | 223993.3652 |
| 183 | C05828 | 141.0655949 | 224477.8851 | 271.9668023 | 267.3275675 | 277.2935703 | C05828 | Methylimidazoleacetic acid | Mass, RT and Spectra | 593542.8821 | 123757.0051 | 90977.0369 | 242617.5351 | 224189.8681 | 306953.6203 |
| 184 | C21298 | 153.0188247 | 430052.4945 | 103.9556256 | 81.2853141 | 110.4760107 | C21298 | 2,6-Dihydroxybenzoic acid | Mass, RT and Spectra | 130070.2587 | 746001.6698 | 844091.4107 | 134152.5947 | 236745.4248 | 347460.7138 |
| 185 | C00499 | 87.01966424 | 11831.90006 | 1162.296159 | 1158.306521 | 1166.263404 | C00499 | Allantoic acid | Mass, RT | 129.0998575 | 25912.9022 | 23302.31864 | 1067.833817 | 2157.283701 | 5698.15168 |
| 186 | C05832 | 249.0874052 | 33751.38756 | 1553.507872 | 1547.877978 | 1554.959132 | C05832 | 5-Hydroxyindoleacetylglycine | Mass, RT and Spectra | 18260.87848 | 44095.62706 | 48819.11199 | 24522.0961 | 22797.77106 | 24773.50804 |
| 187 | C00500 | 581.2387818 | 23322.71971 | 102.6543814 | 97.66391782 | 104.8600675 | C00500 | Biliverdin | Mass, RT and Spectra | 7467331.141 | 8189013.755 | 8283737.776 | 8479141.502 | 8379470.497 | 7636705.971 |
| 188 | C00502 | 165.0397567 | 177927.2403 | 426.2739993 | 410.9574064 | 431.6791043 | C00502 | D-Xylonate | Mass, RT and Spectra | 23849.24704 | 318071.4726 | 354006.5975 | 54390.08246 | 108282.487 | 127909.3939 |
| 189 | C05849 | 467.347816 | 17424305.08 | 735.9763922 | 732.4954711 | 737.1389764 | C05849 | Vitamin K1 2,3-epoxide | Mass, RT and Spectra | 273.2912342 | 38747.49875 | 32319.30511 | 2948.174838 | 3769.407137 | 3326.441844 |
| 190 | C00503 | 121.0501935 | 234372.2433 | 299.4461862 | 283.5728513 | 309.268541 | C00503 | Erythritol | Mass, RT and Spectra | 15855.49682 | 404777.5216 | 457690.5547 | 28780.14418 | 88754.64206 | 126710.928 |
| 191 | C05901 | 245.0818571 | 33257.63728 | 94.5060222 | 91.37032863 | 101.2426864 | C05901 | 3,3',4'5-Tetrahydroxystilbene | Mass, RT | 52.93288058 | 6157.502423 | 16064.8576 | 184.9367333 | 41.18058442 | 399.6441403 |
| 192 | C00504 | 442.1441825 | 4779.96568 | 889.6983419 | 888.6659308 | 890.383085 | C00504 | Folic acid | Mass, RT and Spectra | 3851.22406 | 2198.418974 | 6855.387175 | 71.3652437 | 233.379203 | 436.5568857 |
| 193 | C00506 | 170.0111888 | 14394.96766 | 94.79851415 | 94.27362022 | 95.49590048 | C00506 | Cysteic acid | Mass, RT and Spectra | 7719.202356 | 8755.699831 | 11697.61565 | 9381.809012 | 4315.697751 | 8176.013125 |
| 194 | C00515 | 131.0821077 | 42181.33815 | 570.979757 | 562.4310916 | 582.6201815 | C00515 | D-Ornithine | Mass, RT and Spectra | 34906.09092 | 84.76307487 | 11.38146999 | 20672.01017 | 37548.10968 | 75077.84299 |
| 195 | C05933 | 191.1131465 | 11696.55074 | 100.181789 | 95.92278982 | 102.1234337 | C05933 | N-(o)-Hydroxyarginine | Mass, RT and Spectra | 177.1753245 | 21931.09582 | 27993.73061 | 972.8421588 | 1952.490548 | 3466.160201 |
| 196 | C00523 | 291.2311858 | 13773.1742 | 9.038940242 | 6.159311919 | 12.19494913 | C00523 | Androsterone | Mass, RT and Spectra | 518.5747999 | 29889.8583 | 26311.63946 | 2030.985796 | 7292.166584 | 5834.724518 |
| 197 | C00525 | 203.0819134 | 2675300.125 | 241.2150345 | 220.4843394 | 254.2235703 | C00525 | D-Tryptophan | Mass, RT and Spectra | 316996.6363 | 6036635.326 | 6644803.983 | 296714.0007 | 806164.92 | 906282.6135 |
| 198 | C00532 | 153.0746847 | 62539.11737 | 93.26891991 | 90.92396109 | 116.3965926 | C00532 | L-Arabitol | Mass, RT and Spectra | 62974.49579 | 61713.46481 | 52762.02485 | 75264.08464 | 83574.50437 | 108405.1944 |
| 199 | C00534 | 169.0958964 | 56406.00679 | 268.9079321 | 255.2918933 | 271.4597306 | C00534 | Pyridoxamine | Mass, RT and Spectra | 39848.49093 | 70712.80447 | 64481.6912 | 44302.59725 | 96863.19592 | 51432.46593 |
| 200 | C00544 | 168.0414856 | 2310785.812 | 120.2633926 | 101.4777099 | 123.9243953 | C00544 | Homogentisic acid | Mass, RT and Spectra | 9307.453391 | 18370.5957 | 17090.38813 | 4885.86208 | 4969.30701 | 10934.6987 |
| 201 | C00555 | 88.07571317 | 5624310.126 | 143.538651 | 136.9637558 | 148.4099526 | C00555 | 4-Aminobutyraldehyde | Mass, RT and Spectra | 3144641.957 | 8920321.955 | 8468169.414 | 3531859.416 | 4080063.484 | 3671521.903 |
| 202 | C00559 | 252.1066686 | 22701.67789 | 103.9084816 | 99.8127897 | 110.4315609 | C00559 | Deoxyadenosine | Mass, RT and Spectra | 42147.47595 | 13291.73009 | 13190.76297 | 35263.34263 | 13741.62531 | 22131.26913 |
| 203 | C00575 | 328.0456147 | 10279.62406 | 564.4183802 | 552.0163217 | 578.713165 | C00575 | Cyclic AMP | Mass, RT and Spectra | 1085.126177 | 19884.314 | 17748.58538 | 1481.355204 | 3364.447789 | 5971.161689 |
| 204 | C00581 | 176.0017247 | 59798.3674 | 491.3691373 | 490.0320363 | 492.5274297 | C00581 | Guanidoacetic acid | Mass, RT and Spectra | 3518.215624 | 130998.7963 | 180777.6263 | 3142.934824 | 19671.60946 | 20516.40972 |
| 205 | C05942 | 112.0397315 | 83791.24062 | 214.7491805 | 211.0549506 | 217.0249485 | C05942 | Pyrrole-2-carboxylic acid | Mass, RT and Spectra | 29345.50524 | 127628.2082 | 120722.9334 | 41448.65344 | 102563.3246 | 148754.2718 |
| 206 | C00587 | 137.0239192 | 35475.51806 | 1173.625401 | 1161.589972 | 1178.594625 | C00587 | 3-Hydroxybenzoic acid | Mass, RT and Spectra | 16266.03024 | 5453.891151 | 6264.419613 | 13641.22677 | 12012.4259 | 13462.7122 |
| 207 | C00596 | 113.0240445 | 2354771.092 | 422.280492 | 418.9685102 | 427.4582939 | C00596 | 2-Hydroxy-2,4-pentadienoate | Mass, RT and Spectra | 422829.6015 | 5449204.496 | 3754355.925 | 786574.9312 | 1037235.433 | 875653.0878 |
| 208 | C05944 | 206.1393549 | 518772.5977 | 1553.73704 | 1548.116139 | 1555.605924 | C05944 | Pantothenol | Mass, RT and Spectra | 585336.0249 | 446574.2524 | 497934.4344 | 470985.407 | 506469.5575 | 592495.9405 |
| 209 | C05945 | 255.087144 | 2674.677641 | 447.1715004 | 430.110857 | 447.7830655 | C05945 | L-Phosphoarginine | Mass, RT and Spectra | 1404.75552 | 3520.916759 | 2218.255366 | 2274.520668 | 4718.236951 | 3602.963577 |
| 210 | C00597 | 184.9849674 | 29633.07895 | 521.4624805 | 517.2130369 | 524.6771254 | C00597 | 3-Phosphoglyceric acid | Mass, RT and Spectra | 654.1482279 | 50553.72257 | 63307.88731 | 4816.396801 | 3458.20204 | 11077.26414 |
| 211 | C00599 | 385.3459025 | 3618865.002 | 13.76713412 | 6.466627609 | 24.5675422 | C00599 | Cholestenone | Mass, RT and Spectra | 1043643.899 | 6746342.043 | 6318517.406 | 1326682.126 | 2668328.18 | 1444019.483 |
| 212 | C00606 | 153.0095683 | 10832.38694 | 435.2686874 | 426.5448875 | 439.8395962 | C00606 | 3-Sulfinoalanine | Mass, RT | 230.9447668 | 24126.01037 | 22206.47763 | 1400.945371 | 1397.662714 | 2208.222305 |
| 213 | C00620 | 229.0111677 | 9247.12274 | 489.5222211 | 482.8286295 | 491.0005072 | C00620 | Ribose 1-phosphate | Mass, RT and Spectra | 291.8238438 | 19399.79087 | 14860.32847 | 4749.40775 | 3741.442958 | 3986.050888 |
| 214 | C00624 | 188.0558119 | 37316.22225 | 196.1433583 | 189.0420145 | 207.0421039 | C00624 | N-Acetylglutamic acid | Mass, RT and Spectra | 6866.082296 | 57011.48126 | 53158.40042 | 13878.06311 | 50507.68888 | 82805.72639 |
| 215 | C00631 | 187.000646 | 70076.29998 | 421.5532917 | 418.2259817 | 426.8465926 | C00631 | 2-Phospho-D-glyceric acid | Mass, RT | 9927.964926 | 147756.253 | 117718.6814 | 21323.18982 | 35418.58129 | 29555.51105 |
| 216 | C00632 | 152.0358478 | 2361835.406 | 238.1696756 | 223.8697739 | 270.0059261 | C00632 | 3-Hydroxyanthranilic acid | Mass, RT and Spectra | 4426374.598 | 3136944.123 | 2982571.633 | 4504303.118 | 4655818.719 | 4681872.109 |
| 217 | C05949 | 335.2215685 | 65584.64486 | 490.222831 | 488.4780986 | 498.20926 | C05949 | 12-Keto-leukotriene B4 | Mass, RT and Spectra | 130900.3576 | 22690.37273 | 22859.16289 | 63670.87238 | 59365.38693 | 33062.33979 |
| 218 | C00633 | 123.0441893 | 25029.21797 | 322.2640197 | 308.9910068 | 324.8445329 | C00633 | 4-Hydroxybenzaldehyde | Mass, RT and Spectra | 25413.97904 | 31799.21913 | 32048.79963 | 18738.59924 | 19101.4041 | 18779.5667 |
| 219 | C05956 | 183.1034804 | 2379975.139 | 164.9663352 | 158.88552 | 170.7050831 | C05956 | Prostaglandin G2 | Mass, RT and Spectra | 81026.32325 | 85993.24901 | 75054.0039 | 86783.16028 | 90040.32042 | 75284.82462 |
| 220 | C05964 | 367.2149946 | 3793.318852 | 1177.023943 | 1174.672939 | 1178.578449 | C05964 | 11-Dehydro-thromboxane B2 | Mass, RT and Spectra | 2091.549669 | 6241.355695 | 7061.282083 | 213.3187016 | 17261.21687 | 11608.51987 |
| 221 | C00642 | 151.0394763 | 58637.03873 | 227.6071579 | 217.5306343 | 237.3666556 | C00642 | p-Hydroxyphenylacetic acid | Mass, RT and Spectra | 10234.94193 | 120670.8343 | 118781.6195 | 5854.397501 | 34911.81919 | 105197.8346 |
| 222 | C00643 | 221.0916471 | 82302.10553 | 318.2297114 | 308.5159938 | 323.6389209 | C00643 | 5-Hydroxy-L-tryptophan | Mass, RT and Spectra | 83201.8816 | 79186.65041 | 81396.93145 | 70294.51468 | 78497.36377 | 110368.3606 |
| 223 | C06034 | 178.0705324 | 13591.65952 | 92.76097792 | 87.11645682 | 96.18366208 | C06034 | 4-Hydroxy-4-methylglutamate | Mass, RT and Spectra | 24521.19747 | 6480.684233 | 4953.826812 | 36649.30415 | 9225.548438 | 16425.68156 |
| 224 | C06054 | 214.996562 | 104630.8796 | 214.5296917 | 204.6671511 | 233.63593 | C06054 | 2-Oxo-3-hydroxy-4-phosphobutanoic acid | Mass, RT and Spectra | 44490.67118 | 193747.814 | 196449.7738 | 47278.24312 | 52739.71724 | 54719.06891 |
| 225 | C06121 | 298.2736031 | 167720.6707 | 13.50520163 | 8.145622788 | 19.26872098 | C06121 | 3-ketosphingosine | Mass, RT and Spectra | 55056.34402 | 306469.3541 | 291654.2808 | 66464.95977 | 121737.4327 | 103094.7905 |
| 226 | C00647 | 247.0500106 | 96554.47107 | 380.8237003 | 367.6809116 | 388.619931 | C00647 | Pyridoxamine 5'-phosphate | Mass, RT and Spectra | 10545.40602 | 146089.6898 | 422089.231 | 4469.30068 | 718.9971033 | 29618.78252 |
| 227 | C06126 | 888.6475179 | 13860.06935 | 1217.224962 | 1211.373679 | 1220.291917 | C06126 | Galabiosylceramide (d18:1/9Z-18:1) | Mass, RT and Spectra | 837199.9815 | 617298.4244 | 758888.7397 | 785133.3837 | 896350.1318 | 720145.9767 |
| 228 | C06231 | 143.0812025 | 347556.9862 | 9.172154261 | 5.19185791 | 14.42046294 | C06231 | Ectoine | Mass, RT and Spectra | 60201.34481 | 714050.9876 | 1090749.801 | 96283.31241 | 109779.248 | 156087.409 |
| 229 | C06313 | 238.0915151 | 12354.57513 | 95.23058811 | 87.09014514 | 103.0139713 | C06313 | Biopterin | Mass, RT and Spectra | 24271.55788 | 4217.505821 | 5449.84937 | 15475.27959 | 7413.834717 | 12046.25076 |
| 230 | C06323 | 130.0649695 | 243762.4095 | 1553.06686 | 1547.310041 | 1569.332153 | C06323 | Isoquinoline | Mass, RT and Spectra | 173753.5356 | 258456.8559 | 290989.5858 | 205302.2628 | 260676.885 | 247411.197 |
| 231 | C00651 | 158.0451345 | 35554.24331 | 516.1257233 | 512.5004799 | 520.676646 | C00651 | L-2-Amino-4-methylenepentanedioic acid | Mass, RT and Spectra | 2428.408802 | 74675.4805 | 78589.93387 | 16029.32792 | 7267.151472 | 11860.11992 |
| 232 | C06424 | 227.2011817 | 2722680.944 | 93.86386956 | 92.66187071 | 94.8786 | C06424 | Myristic acid | Mass, RT and Spectra | 496123.7326 | 4142750.66 | 5120445.499 | 1047016.306 | 652514.3925 | 965308.7374 |
| 233 | C00655 | 363.0352912 | 2564482.689 | 1785.590878 | 1767.180357 | 1793.58155 | C00655 | Xanthylic acid | Mass, RT and Spectra | 231664.7135 | 68402.14913 | 65059.67065 | 224334.3838 | 165941.2776 | 216464.9502 |
| 234 | C22112 | 428.3270633 | 2683326.025 | 722.0458047 | 719.3704759 | 723.6007003 | C22112 | 4alpha-Carboxy-5alpha-cholesta-8,24-dien-3beta-ol | Mass, RT and Spectra | 19264.74729 | 14759.31642 | 12666.20042 | 5171.067905 | 17038.3626 | 17690.15771 |
| 235 | C00659 | 145.050149 | 21408.35941 | 94.1332293 | 77.19163617 | 95.68791246 | C00659 | Acetohydroxybutyrate | Mass, RT and Spectra | 776.3193821 | 43166.63816 | 35175.69326 | 4333.377058 | 6706.144879 | 11501.12427 |
| 236 | C06426 | 277.2165493 | 1605836.094 | 92.01720708 | 90.58961178 | 93.2359208 | C06426 | Gamma-Linolenic acid | Mass, RT and Spectra | 365217.6839 | 2512066.457 | 2704474.076 | 486411.0938 | 283090.4192 | 1079029 |
| 237 | C06427 | 279.2311266 | 352069.5686 | 1042.579086 | 1037.599551 | 1046.308687 | C06427 | Alpha-Linolenic acid | Mass, RT and Spectra | 11636.11413 | 204188.3326 | 243607.6185 | 34544.10247 | 40316.45942 | 39238.1524 |
| 238 | C00666 | 191.1017831 | 153645.4314 | 96.99613854 | 93.88101841 | 103.7511816 | C00666 | Diaminopimelic acid | Mass, RT and Spectra | 172938.5535 | 121067.2068 | 106243.0821 | 163434.0232 | 218393.2183 | 224702.0785 |
| 239 | C00669 | 120.0115738 | 14262.02817 | 478.1281477 | 475.2646394 | 483.2748515 | C00669 | gamma-Glutamylcysteine | Mass, RT and Spectra | 3411.882877 | 28331.30158 | 32887.91466 | 1403.39677 | 2361.986328 | 4061.208501 |
| 240 | C00672 | 213.0165391 | 138874.6655 | 430.8989756 | 425.1475114 | 436.8415353 | C00672 | Deoxyribose 1-phosphate | Mass, RT and Spectra | 18684.47295 | 293721.3322 | 257446.9362 | 37516.73009 | 44681.61084 | 46287.12379 |
| 241 | C00673 | 215.0332525 | 21455.42646 | 251.4760684 | 245.2270682 | 263.158461 | C00673 | Deoxyribose 5-phosphate | Mass, RT and Spectra | 26768.33518 | 18563.23182 | 21130.96372 | 21721.55634 | 16519.90659 | 18036.32597 |
| 242 | C06428 | 301.2196491 | 108465.4095 | 92.05816716 | 90.29534851 | 92.9247492 | C06428 | Eicosapentaenoic acid | Mass, RT and Spectra | 16947.84478 | 184345.4451 | 200186.1422 | 25002.22627 | 34728.53141 | 30111.21178 |
| 243 | C00682 | 143.0332824 | 77016.39397 | 168.1556291 | 143.6791857 | 186.7603633 | C00682 | 2-Hydroxymuconate semialdehyde | Mass, RT and Spectra | 13617.32142 | 150109.7289 | 134446.3064 | 34220.89931 | 77141.46676 | 35493.29128 |
| 244 | C00695 | 407.27973 | 120235.9367 | 99.85029039 | 93.99765235 | 113.3277297 | C00695 | Cholic acid | Mass, RT and Spectra | 22529.23046 | 191618.4412 | 307606.4982 | 12243.46374 | 29416.25901 | 39472.29817 |
| 245 | C00700 | 260.9793534 | 18961.61375 | 318.4548667 | 313.6996869 | 327.8646189 | C00700 | Xanthosine 5-triphosphate | Mass, RT | 2109.28366 | 42322.69996 | 38090.12541 | 2274.161864 | 6413.742547 | 11954.58948 |
| 246 | C00716 | 106.0495212 | 3067527.881 | 103.4582423 | 99.8127897 | 108.6065854 | C00716 | 2-Amino-3-hydroxypropanoic acid | Mass, RT and Spectra | 2970432.329 | 3167643.609 | 3264865.309 | 2920697.625 | 3179966.347 | 2763741.025 |
| 247 | C00718 | 371.1527991 | 14085.75643 | 627.5304099 | 626.2514404 | 628.1297237 | C00718 | Amylose | Mass, RT and Spectra | 7116.714317 | 106664.9204 | 104856.7127 | 13175.17351 | 20782.99547 | 26446.58723 |
| 248 | C06429 | 327.2314912 | 477303.5994 | 90.70734864 | 89.20357046 | 100.028788 | C06429 | Docosahexaenoic acid | Mass, RT and Spectra | 102487.2198 | 794367.527 | 812193.3597 | 130128.5715 | 162644.066 | 173006.1243 |
| 249 | C00719 | 154.0449311 | 57771.84066 | 293.1252594 | 276.4418658 | 317.3310097 | C00719 | Betaine | Mass, RT and Spectra | 4193.799411 | 119268.8084 | 94506.36501 | 6064.59344 | 30273.44562 | 47848.97434 |
| 250 | C06451 | 125.0005883 | 456956.5043 | 370.984273 | 341.4704828 | 382.3074658 | C06451 | 2-Hydroxyethylphosphonate | Mass, RT and Spectra | 2118.822219 | 872648.6093 | 1017930.013 | 49112.77234 | 73705.46573 | 127747.2136 |
| 251 | C06459 | 169.0855262 | 164105.4955 | 477.6131068 | 468.3710535 | 497.5009961 | C06459 | Phosphorylcholine | Mass, RT and Spectra | 171193.7156 | 153127.3143 | 151263.02 | 172146.9542 | 172506.7006 | 172328.3209 |
| 252 | C06469 | 266.0874995 | 45541.52643 | 447.0257058 | 437.4723959 | 449.1912971 | C06469 | Neuraminic acid | Mass, RT and Spectra | 11701.6451 | 99258.13963 | 81437.70906 | 5589.351691 | 11188.51777 | 10094.95693 |
| 253 | C00735 | 363.2158501 | 122912.6175 | 578.439454 | 570.0679957 | 589.7495284 | C00735 | Cortisol | Mass, RT and Spectra | 81612.33017 | 114718.9113 | 87530.56673 | 48188.71368 | 473033.8744 | 186461.6905 |
| 254 | C00747 | 80.04955468 | 4462957.133 | 1553.258978 | 1551.734601 | 1555.417283 | C00747 | Pyridine | Mass, RT and Spectra | 16815.23752 | 381801.7875 | 255333.2322 | 61955.36857 | 86261.68271 | 94600.12992 |
| 255 | C00750 | 203.222178 | 13638.56828 | 95.63437677 | 94.12893291 | 98.35885442 | C00750 | Spermine | Mass, RT and Spectra | 7726.86772 | 29863.49547 | 27044.46023 | 1841.544832 | 6623.443123 | 6530.95021 |
| 256 | C00777 | 301.2191578 | 37000.27294 | 821.7593525 | 819.9241359 | 823.7073445 | C00777 | All-trans-retinoic acid | Mass, RT and Spectra | 48966.14803 | 24505.24013 | 26068.27794 | 40630.45452 | 43423.53016 | 39118.96097 |
| 257 | C06470 | 250.0923655 | 163362.4426 | 421.2326864 | 416.981076 | 424.8889085 | C06470 | Muramic acid | Mass, RT and Spectra | 85421.82249 | 45954.15825 | 45348.26434 | 79869.34088 | 77367.75956 | 79294.75043 |
| 258 | C06501 | 260.0317349 | 10433.18464 | 90.28015892 | 88.63084634 | 91.2622649 | C06501 | O-Phosphotyrosine | Mass, RT and Spectra | 1007.111497 | 10457.97106 | 19192.38318 | 2548.634094 | 11295.26658 | 21489.748 |
| 259 | C00785 | 139.0498931 | 798259.0215 | 356.7817318 | 343.7546714 | 371.1392295 | C00785 | Urocanic acid | Mass, RT and Spectra | 1389941.035 | 384577.8271 | 713419.2416 | 628985.39 | 732491.2847 | 650962.5972 |
| 260 | C00788 | 184.0966963 | 28922.17691 | 388.6743569 | 385.9545489 | 393.8706593 | C00788 | Epinephrine | Mass, RT and Spectra | 46674.16698 | 30716.59078 | 26189.36115 | 34615.43441 | 36111.20858 | 14588.63027 |
| 261 | C06554 | 128.0110487 | 11938.27029 | 551.2355129 | 542.7045879 | 558.9659655 | C06554 | Cyanuric acid | Mass, RT and Spectra | 592.0325112 | 25798.66794 | 21036.71432 | 4170.191398 | 2546.970946 | 2767.65193 |
| 262 | C00791 | 112.0512579 | 1508250.201 | 212.8464525 | 188.5666933 | 228.0060283 | C00791 | Creatinine | Mass, RT and Spectra | 229533.8475 | 2779325.108 | 2735339.386 | 229954.7105 | 768485.4086 | 804944.9303 |
| 263 | C06579 | 199.0945737 | 24687.6701 | 13.03875503 | 9.582791558 | 16.54527777 | C06579 | cis-2,3-Dihydroxy-2,3-dihydro-p-cumate | Mass, RT and Spectra | 14070.72341 | 43883.86769 | 30414.83267 | 16281.94142 | 22037.51366 | 20781.98261 |
| 264 | C00792 | 173.1046928 | 2730848.986 | 890.2011329 | 886.3333537 | 891.975235 | C00792 | D-Arginine | Mass, RT and Spectra | 66312.15416 | 63742.25828 | 77941.18918 | 60113.44008 | 56206.7884 | 61054.99952 |
| 265 | C00794 | 217.0289681 | 1037390.567 | 374.8589842 | 370.7479913 | 382.6194632 | C00794 | Sorbitol | Mass, RT and Spectra | 108550.6963 | 2693889.014 | 1989303.536 | 153403.6148 | 262522.8341 | 193309.4361 |
| 266 | D06890 | 193.0611835 | 9625.210269 | 128.9030086 | 109.9694978 | 134.2455427 | D06890 | 4-Aminohippuric acid | Mass, RT and Spectra | 680.6117869 | 155092.4884 | 136033.6446 | 18794.67776 | 34110.40876 | 29725.3772 |
| 267 | C00798 | 794.1031222 | 10991.20059 | 466.9980532 | 466.284752 | 467.793339 | C00798 | Formyl-CoA | Mass, RT and Spectra | 406.8870385 | 23209.18314 | 21472.90967 | 2315.563021 | 1950.272876 | 2829.874496 |
| 268 | C00803 | 101.0604031 | 195579.6815 | 135.3337419 | 110.4977789 | 162.2668113 | C00803 | Valeric acid | Mass, RT and Spectra | 21637.07377 | 405112.6273 | 401803.3416 | 19343.27241 | 54656.01711 | 47666.55612 |
| 269 | C00818 | 209.0297246 | 78881.02847 | 525.2264619 | 516.4695644 | 536.1356922 | C00818 | Glucaric acid | Mass, RT and Spectra | 11709.41426 | 140219.1127 | 170213.3794 | 28501.31812 | 33286.41997 | 44881.03315 |
| 270 | C00819 | 146.0647358 | 267951.2223 | 421.8039719 | 404.2218789 | 425.7140118 | C00819 | D-Glutamine | Mass, RT and Spectra | 58007.12708 | 538392.3055 | 508123.6883 | 65842.4305 | 109365.5462 | 97028.95919 |
| 271 | C00828 | 445.3120001 | 2831875.377 | 933.9588308 | 931.8619673 | 936.5951272 | C00828 | Vitamin K2 | Mass, RT and Spectra | 24522.23385 | 13920.91058 | 14377.80128 | 25044.018 | 26192.11598 | 17611.59453 |
| 272 | C06645 | 380.9214288 | 39866.76055 | 96.06298165 | 93.89527243 | 102.5542435 | C06645 | 6-Oxo-2-hydroxy-7-(4'-chlorophenyl)-3,8,8-trichloroocta-2E,4E,7E-trienoate | Mass, RT and Spectra | 46197.04761 | 55342.34574 | 37980.55916 | 27545.16841 | 37644.45796 | 18621.53797 |
| 273 | C00836 | 302.3059643 | 13197.58073 | 100.3895291 | 99.01615727 | 102.3926485 | C00836 | Sphinganine | Mass, RT and Spectra | 60.36516863 | 14113.8116 | 14772.79883 | 228.5774053 | 587.7257863 | 2084.433608 |
| 274 | C00846 | 159.0292324 | 25240.38983 | 473.882267 | 467.5078653 | 482.4876187 | C00846 | 3-Oxoadipic acid | Mass, RT and Spectra | 16.06212542 | 13467.05503 | 18129.32415 | 713.6922467 | 731.3779512 | 818.900853 |
| 275 | C00847 | 182.0453759 | 529832.2898 | 112.1574906 | 89.99064179 | 126.2657949 | C00847 | 4-Pyridoxic acid | Mass, RT and Spectra | 488895.2914 | 659260.0461 | 776845.0757 | 340904.2737 | 401839.1222 | 74624.26743 |
| 276 | C06656 | 227.1131573 | 10635.66064 | 101.0449904 | 94.29682868 | 107.3224978 | C06656 | Deoxyguanidinoproclavaminic acid | Mass, RT and Spectra | 968.2169864 | 18682.09974 | 30465.6666 | 1766.563241 | 638.0373397 | 2758.254604 |
| 277 | C00848 | 169.1220604 | 201145.6346 | 13.15175499 | 6.873391124 | 15.27453726 | C00848 | 6-oxocineole | Mass, RT and Spectra | 169271.4713 | 239308.0103 | 242520.2433 | 174335.0268 | 188292.8863 | 183369.6345 |
| 278 | C06670 | 190.9645704 | 79692.86796 | 235.04838 | 221.7562449 | 251.8716952 | C06670 | 2,4-Dichlorobenzoate | Mass, RT and Spectra | 21770.28903 | 145083.8033 | 125986.0517 | 34263.36567 | 53535.01587 | 64506.59646 |
| 279 | C00857 | 665.0967515 | 16132.77509 | 1734.552473 | 1731.715753 | 1737.389192 | C00857 | Nicotinic acid adenine dinucleotide | Mass, RT and Spectra | 43030.02654 | 344.4571524 | 2392.090112 | 40383.67857 | 36617.17704 | 6408.181691 |
| 280 | C06671 | 171.0116 | 240.284227 | 115.2207879 | 113.7593377 | 117.6171791 | C06671 | 2,4-Dichlorobenzoyl-CoA | Mass, RT and Spectra | 220.0835665 | 12877.10895 | 25488.08046 | 1358.755563 | 1611.174105 | 3433.973734 |
| 281 | C06677 | 171.0116415 | 240.284227 | 115.2207879 | 113.7593377 | 117.6171791 | C06677 | p-Toluenesulfonic acid | Mass, RT and Spectra | 0 | 202.6607432 | 267.3919811 | 86.87611522 | 103.9945726 | 36.71489377 |
| 282 | C06772 | 106.086155 | 94682.12919 | 114.7518443 | 113.2633681 | 115.8620553 | C06772 | Diethanolamine | Mass, RT and Spectra | 19695.96694 | 131718.9687 | 97303.56595 | 18421.63197 | 47601.99137 | 45699.59907 |
| 283 | C00860 | 140.0819085 | 18529.56818 | 108.1347296 | 105.7917141 | 112.2419622 | C00860 | L-Histidinol | Mass, RT and Spectra | 4926.137221 | 22032.2249 | 69129.35324 | 15942.16213 | 8381.267 | 2747.596232 |
| 284 | C00864 | 220.1175183 | 587275.183 | 294.5526007 | 283.7018577 | 308.2948045 | C00864 | Pantothenic acid | Mass, RT and Spectra | 903501.0037 | 304864.2596 | 300602.9636 | 499558.8987 | 633251.0415 | 600432.1224 |
| 285 | C00869 | 297.2430443 | 597012.2666 | 91.23444 | 88.93961408 | 93.55778975 | C00869 | 2-oxo-octadecanoic acid | Mass, RT and Spectra | 126678.4759 | 1118330.446 | 1085848.579 | 141158.6346 | 193251.2158 | 254151.0105 |
| 286 | C00870 | 138.0198893 | 14729.05675 | 118.9877614 | 97.20888788 | 123.7660609 | C00870 | 4-Nitrophenol | Mass, RT and Spectra | 13087.13692 | 20797.57483 | 21269.95572 | 32220.80047 | 21270.31405 | 2936.979099 |
| 287 | C07326 | 163.0607113 | 2830202.408 | 314.053606 | 291.3361977 | 332.4398447 | C07326 | 1,5-Anhydrosorbitol | Mass, RT and Spectra | 365669.9229 | 5112925.445 | 6025201.513 | 804617.8227 | 1024978.557 | 1108599.629 |
| 288 | C07888 | 261.0319877 | 4471.744057 | 162.8823759 | 158.529935 | 165.6125371 | C07888 | Cyclophosphamide | Mass, RT and Spectra | 12627.79923 | 564.4618912 | 808.4706029 | 8353.531712 | 7755.127433 | 6017.589304 |
| 289 | C00879 | 211.0446403 | 3990.896076 | 592.387816 | 578.5003506 | 594.4077057 | C00879 | Galactaric acid | Mass, RT and Spectra | 2585234.283 | 3554523.618 | 3118915.558 | 2541185.534 | 2079347.878 | 2515223.227 |
| 290 | C00882 | 437.0132971 | 11271.23418 | 422.1146345 | 409.6320207 | 424.2408973 | C00882 | Dephospho-CoA | Mass, RT and Spectra | 180.2898863 | 25767.69168 | 21983.2177 | 2160.026191 | 4423.634738 | 2915.533903 |
| 291 | C00902 | 129.0553732 | 5631555.76 | 109.0462532 | 90.7092791 | 119.5915216 | C00902 | 2-Ketohexanoic acid | Mass, RT and Spectra | 641.2560754 | 19801.70446 | 14703.66587 | 2337.056702 | 8182.084681 | 5667.627665 |
| 292 | C08156 | 411.2895872 | 15490.4516 | 95.29161717 | 94.72898381 | 96.72359053 | C08156 | Testosterone cypionate | Mass, RT and Spectra | 2651.790039 | 36241.31203 | 26742.56486 | 905.2400797 | 1647.831664 | 1627.855738 |
| 293 | C00906 | 127.0507727 | 81181.68157 | 138.6832731 | 132.0970378 | 163.1838382 | C00906 | Dihydrothymine | Mass, RT and Spectra | 1906.884284 | 164782.5976 | 172580.9981 | 11145.12721 | 21081.08022 | 18526.8611 |
| 294 | C00921 | 315.1219394 | 17882.6142 | 10.86579183 | 8.207649533 | 13.72587156 | C00921 | 7,8-Dihydropteroic acid | Mass, RT and Spectra | 37164.94441 | 1932.502137 | 1530.225484 | 24039.47023 | 28742.19433 | 29263.53777 |
| 295 | C08285 | 289.2360889 | 29218.47211 | 1084.916304 | 1078.407154 | 1090.42896 | C08285 | (S)-10,16-Dihydroxyhexadecanoic acid | Mass, RT and Spectra | 42679.36446 | 20980.52393 | 22619.71049 | 18021.11013 | 30381.37252 | 27781.48157 |
| 296 | C00931 | 285.0461301 | 13220.06294 | 515.1896213 | 512.993699 | 517.7842242 | C00931 | Porphobilinogen | Mass, RT and Spectra | 10.22378155 | 33143.53599 | 29493.00379 | 4284.501237 | 981.1276096 | 2155.14741 |
| 297 | C08302 | 189.138061 | 595256.4593 | 833.3661868 | 807.3167895 | 845.8266219 | C08302 | Dimethyltryptamine | Mass, RT and Spectra | 303145.1996 | 877282.0871 | 868526.3062 | 390619.0971 | 479716.6972 | 408524.1785 |
| 298 | C00936 | 215.0320812 | 18700279.44 | 419.4086511 | 411.9693442 | 425.0261056 | C00936 | D-Mannose | Mass, RT and Spectra | 3324450.07 | 40850384.46 | 32909640.22 | 6106113.411 | 7585456.327 | 6606411.446 |
| 299 | C00940 | 144.0297447 | 263425.0919 | 281.2456482 | 262.8250913 | 294.8303509 | C00940 | 2-Keto-glutaramic acid | Mass, RT and Spectra | 45036.76132 | 469738.8031 | 546889.6334 | 80442.2134 | 156940.1502 | 133573.6376 |
| 300 | C00944 | 189.0398025 | 512381.8553 | 285.9006525 | 251.7132334 | 313.2073924 | C00944 | 3-Dehydroquinate | Mass, RT and Spectra | 135193.1959 | 888504.1651 | 937121.7748 | 176566.8975 | 240981.9411 | 313027.6008 |
| 301 | C00954 | 174.0554534 | 207219.6585 | 187.0719271 | 164.8166415 | 201.5115552 | C00954 | Indoleacetic acid | Mass, RT and Spectra | 31281.34196 | 495939.0061 | 465587.8064 | 25061.96862 | 99475.55836 | 119837.0049 |
| 302 | C00955 | 160.0761277 | 54851.15023 | 1177.702682 | 1173.222 | 1179.9372 | C00955 | Tryptophanol | Mass, RT and Spectra | 4237.882621 | 28979.07342 | 34315.57046 | 4339.643802 | 2792.000989 | 3311.605113 |
| 303 | G13036 | 486.3930165 | 2838693.015 | 1273.207251 | 1232.046634 | 1299.707615 | G13036 | (GlcA)2 (GlcN)1 (GlcNAc)1 (S)3 | Mass, RT and Spectra | 127899.5465 | 169614.4279 | 246594.9852 | 164343.6458 | 252813.6531 | 148155.1205 |
| 304 | C00956 | 160.0609755 | 277721.4527 | 236.3886244 | 223.5775248 | 250.6194261 | C00956 | Aminoadipic acid | Mass, RT and Spectra | 33733.57381 | 502935.8348 | 614703.1725 | 51744.69675 | 174738.0883 | 203376.91 |
| 305 | C00969 | 75.04391395 | 18334.28252 | 101.4643039 | 99.18811956 | 103.0289756 | C00969 | 3-Hydroxypropanal | Mass, RT and Spectra | 48.57861972 | 29019.40123 | 38958.16187 | 95.67952095 | 4358.662627 | 4317.957367 |
| 306 | C00979 | 146.0453298 | 383588.78 | 292.5832328 | 275.8944115 | 315.1813094 | C00979 | O-Acetylserine | Mass, RT and Spectra | 82859.43481 | 658564.016 | 705565.5653 | 127669.5674 | 250859.2279 | 248196.7725 |
| 307 | C00984 | 89.02409912 | 8884668.8 | 421.6525593 | 418.8320063 | 424.8230523 | C00984 | D-Galactose | Mass, RT and Spectra | 1291574.326 | 18272729.24 | 16105645.22 | 2533061.277 | 3656432.369 | 3628309.775 |
| 308 | C00988 | 155.9826833 | 11666.37186 | 419.6239009 | 414.9874815 | 425.3409277 | C00988 | Phosphoglycolic acid | Mass, RT | 376.337086 | 27649.89617 | 18799.72699 | 3341.560218 | 6550.75317 | 5581.825947 |
| 309 | C08314 | 315.2535073 | 21056.77838 | 95.23396896 | 93.46366676 | 98.57029871 | C08314 | threo-9,10-Dihydroxystearic acid | Mass, RT and Spectra | 2544.228002 | 41647.43051 | 43840.47471 | 673.4010589 | 6718.678024 | 2555.214814 |
| 310 | C08316 | 337.3102192 | 194025.1932 | 89.63582841 | 87.12989477 | 90.8325241 | C08316 | Erucic acid | Mass, RT and Spectra | 54886.8091 | 376974.0546 | 434138.0787 | 46413.8934 | 25157.57338 | 18604.81388 |
| 311 | C00989 | 103.0397266 | 4436562.682 | 180.3503006 | 158.1650086 | 205.2628373 | C00989 | Gamma Hydroxybutyric Acid | Mass, RT and Spectra | 822717.1186 | 8644809.418 | 8339008.61 | 2047312.009 | 1942576.785 | 2183149.551 |
| 312 | C08317 | 217.1792681 | 178599.874 | 1105.941028 | 1093.75441 | 1110.89508 | C08317 | 12-Hydroxydodecanoic acid | Mass, RT and Spectra | 205175.3146 | 161046.6367 | 146413.4663 | 126374.8347 | 158667.3812 | 156608.1631 |
| 313 | C00990 | 117.1021623 | 245040.2024 | 116.7626429 | 115.1050216 | 118.3749213 | C00990 | 5-Aminopentanamide | Mass, RT and Spectra | 107027.5001 | 427231.7174 | 454770.7803 | 124034.0509 | 131211.1998 | 130035.2811 |
| 314 | C00993 | 159.0770442 | 10854.44563 | 232.335768 | 224.9395106 | 244.5226566 | C00993 | D-Alanyl-D-alanine | Mass, RT and Spectra | 48.81102879 | 140375.3038 | 243325.7876 | 13238.0324 | 10036.52119 | 18381.93775 |
| 315 | C08320 | 367.3565779 | 11626.85291 | 87.56420922 | 84.84925776 | 92.07533918 | C08320 | Tetracosanoic acid | Mass, RT and Spectra | 27.23059688 | 28609.44606 | 32895.52587 | 721.3429252 | 648.5847033 | 1546.32859 |
| 316 | C08322 | 225.185358 | 604608.6068 | 93.6806528 | 91.91929008 | 94.72036242 | C08322 | Myristoleic acid | Mass, RT and Spectra | 104284.7076 | 809100.147 | 979469.7029 | 251951.3381 | 129334.2942 | 189662.0144 |
| 317 | HMDB0000333 | 182.0536124 | 54753.79683 | 197.3639947 | 185.6758441 | 211.4814722 | HMDB0000333 | Isohomovanillic acid | Mass, RT and Spectra | 17079.74508 | 99689.63073 | 111152.1966 | 11651.39633 | 24156.25375 | 29754.68169 |
| 318 | C08323 | 365.3412661 | 78601.59094 | 88.67223826 | 86.36294843 | 90.62513918 | C08323 | Nervonic acid | Mass, RT and Spectra | 70782.32204 | 94601.10315 | 93942.67203 | 62628.32282 | 76982.37278 | 86963.51082 |
| 319 | C01005 | 186.0177315 | 46888.51468 | 104.713 | 101.4643039 | 111.6956828 | C01005 | Phosphoserine | Mass, RT and Spectra | 30773.62431 | 62033.38205 | 68430.86818 | 25467.06086 | 56635.75883 | 66015.85366 |
| 320 | C08348 | 129.0189546 | 98291.5438 | 496.8859184 | 491.9464815 | 499.8422615 | C08348 | Galacturonic acid | Mass, RT and Spectra | 20988.31528 | 182544.3582 | 161909.7357 | 52946.53598 | 85640.0085 | 84982.93316 |
| 321 | C08357 | 353.1451833 | 3757.661112 | 434.9204514 | 430.6326576 | 437.40413 | C08357 | Estradiol-17beta 3-sulfate | Mass, RT and Spectra | 7320.14525 | 1384.731461 | 1174.41954 | 8229.852196 | 3900.568068 | 6462.271696 |
| 322 | C01010 | 103.0145533 | 2556812.252 | 237.6397953 | 231.3368466 | 250.0837377 | C01010 | Urea-1-carboxylate | Mass, RT and Spectra | 49786.48562 | 5471454.539 | 5715811.642 | 184590.7482 | 1580637.504 | 2097297.144 |
| 323 | C08362 | 255.2309881 | 322982.6287 | 89.04138754 | 73.35246433 | 94.61078501 | C08362 | Palmitoleic acid | Mass, RT and Spectra | 44083.28355 | 361602.9766 | 270978.1716 | 28804.23561 | 62748.97302 | 47396.89101 |
| 324 | HMDB0000354 | 117.0552742 | 177121.8615 | 212.3547675 | 206.7476842 | 219.1354907 | HMDB0000354 | 2-Methyl-3-hydroxybutyric acid | Mass, RT and Spectra | 14140.12437 | 342658.1625 | 361160.812 | 23939.25916 | 75692.40752 | 60074.00813 |
| 325 | HMDB0000407 | 117.0553155 | 3447058.689 | 145.9712243 | 130.2073505 | 171.2558954 | HMDB0000407 | 2-Hydroxy-3-methylbutyric acid | Mass, RT and Spectra | 715622.5295 | 6067827.029 | 6325883.595 | 529221.3891 | 1455243.446 | 1201043.072 |
| 326 | C01017 | 219.0770099 | 6708.157322 | 148.8560787 | 145.7690698 | 163.8403803 | C01017 | 5-Hydroxytryptophan | Mass, RT and Spectra | 172586.2881 | 72977.76404 | 76097.32498 | 145013.5669 | 129808.8969 | 141884.4771 |
| 327 | C08375 | 181.1582387 | 26118.6929 | 1106.949456 | 1101.278897 | 1111.475129 | C08375 | (3Z,6Z,9Z)-dodecatrienol | Mass, RT and Spectra | 37473.08385 | 19476.39517 | 16895.04303 | 16932.00013 | 19015.11738 | 22414.54647 |
| 328 | C01026 | 102.0556836 | 201049.4168 | 313.7760212 | 291.6710683 | 332.5040301 | C01026 | Dimethylglycine | Mass, RT and Spectra | 9197.228664 | 428708.119 | 447340.4437 | 33371.19594 | 56916.89162 | 95022.68995 |
| 329 | C08390 | 75.02774357 | 9935.896379 | 439.0505159 | 437.1696044 | 441.2850366 | C08390 | 1-Propanethiol | Mass, RT and Spectra | 161.0646222 | 19777.29103 | 22403.64848 | 1582.15803 | 1452.920129 | 1792.146139 |
| 330 | C08486 | 195.1384022 | 201628.7241 | 95.2483294 | 93.54541421 | 99.02345817 | C08486 | ethyl 2E,4Z-decadienoate | Mass, RT and Spectra | 29334.99713 | 368000.9606 | 343948.8549 | 56455.27551 | 53252.72324 | 97626.4891 |
| 331 | C01029 | 188.1752184 | 734336.4676 | 475.120536 | 445.512163 | 508.6498392 | C01029 | N8-Acetylspermidine | Mass, RT and Spectra | 963229.9325 | 606971.6807 | 542698.7318 | 866011.5003 | 1165696.915 | 895898.1114 |
| 332 | C01041 | 174.0167586 | 6431.294106 | 392.7857569 | 384.5552188 | 400.296215 | C01041 | Ascorbic acid | Mass, RT and Spectra | 156.3816068 | 14080.70232 | 12653.89896 | 897.4756164 | 2466.689217 | 3621.220083 |
| 333 | C01042 | 112.0400657 | 206332.6408 | 461.9902106 | 459.3580347 | 466.6163314 | C01042 | N-Acetyl-L-aspartic acid | Mass, RT and Spectra | 23640.48012 | 470583.4614 | 428827.6058 | 51547.35044 | 65171.13178 | 71537.23777 |
| 334 | C01044 | 160.0244905 | 165085.4547 | 479.1147693 | 477.2330378 | 481.3819244 | C01044 | N-Formyl-L-aspartate | Mass, RT and Spectra | 11463.29695 | 382804.6423 | 429863.5047 | 11456.50034 | 69362.37663 | 63553.6919 |
| 335 | C01045 | 172.0245359 | 84888.74171 | 469.4375098 | 463.7824568 | 474.0748928 | C01045 | N-Formyl-L-glutamic acid | Mass, RT and Spectra | 92800.16528 | 64914.54696 | 65608.60114 | 93665.22333 | 87842.43123 | 115332.6292 |
| 336 | C01054 | 427.3894548 | 1130.012127 | 1024.469061 | 1011.867519 | 1042.386241 | C01054 | (S)-2,3-Epoxysqualene | Mass, RT and Spectra | 306.1165396 | 29058.88603 | 53992.04623 | 4189.964897 | 3914.260962 | 5801.145288 |
| 337 | C16682 | 286.2285981 | 2842454.486 | 1037.120227 | 1034.745816 | 1040.417585 | C16682 | 9-cis-Retinol | Mass, RT and Spectra | 24677.27111 | 11096.06084 | 21822.40454 | 6434.724253 | 19947.77097 | 16770.81953 |
| 338 | C10164 | 124.0392271 | 90035.39822 | 13.51527668 | 9.891844157 | 15.69223417 | C10164 | Picolinic acid | Mass, RT and Spectra | 77753.29683 | 100485.3735 | 105860.7344 | 80807.52745 | 79982.4108 | 88404.98681 |
| 339 | C10264 | 339.1613704 | 25548.04093 | 1047.484852 | 1045.33504 | 1050.724453 | C10264 | 6-Hydroxy-4-methoxy-3-(3-methyl-2-butenyl)-2-(2-phenylethenyl)benzoic acid | Mass, RT and Spectra | 22963.92077 | 21753.55695 | 19986.04337 | 38059.82526 | 23043.57222 | 34123.15892 |
| 340 | C10544 | 417.1517652 | 3601.620349 | 756.4672084 | 753.4226504 | 758.3745845 | C10544 | 8-Acetoxypinoresinol | Mass, RT and Spectra | 5235.610077 | 1755.755365 | 1927.359238 | 5002.535764 | 4176.437618 | 4556.24201 |
| 341 | C10998 | 170.0958972 | 8151.157955 | 910.8536294 | 901.9244814 | 913.8424271 | C10998 | 4-Aminobiphenyl | Mass, RT and Spectra | 4792.310611 | 11247.04075 | 10956.47795 | 6083.822621 | 6852.972691 | 5632.252047 |
| 342 | C01056 | 179.1170742 | 19149.98998 | 603.2874135 | 582.3292383 | 623.7237018 | C01056 | (S)-6-Hydroxynicotine | Mass, RT and Spectra | 26089.58814 | 13021.94034 | 12127.74077 | 42632.69443 | 23423.48836 | 56392.6605 |
| 343 | C11045 | 293.1133682 | 13276.23406 | 241.2754555 | 236.6163889 | 253.3054432 | C11045 | Aspartame | Mass, RT and Spectra | 62857.16588 | 154915.0684 | 175440.2713 | 75868.21474 | 97128.87043 | 95338.57515 |
| 344 | C01059 | 110.0244008 | 3985.868152 | 1167.237855 | 1164.898863 | 1171.720193 | C01059 | 2,5-Dihydroxypyridine | Mass, RT and Spectra | 83.52569026 | 8741.94053 | 7805.511582 | 524.5892333 | 1300.882066 | 1646.69014 |
| 345 | C01061 | 198.016936 | 2883915.559 | 89.51137869 | 86.02683965 | 110.4150745 | C01061 | 4-Fumarylacetoacetic acid | Mass, RT and Spectra | 79685.29212 | 17624.65995 | 15213.3944 | 79322.53006 | 82334.31715 | 63042.3918 |
| 346 | C01074 | 222.0970956 | 27012.95189 | 95.20701974 | 91.73614585 | 101.0613308 | C01074 | N-Acetylgalactosamine | Mass, RT and Spectra | 34610.78975 | 20955.15175 | 13838.22747 | 35480.82313 | 39859.23828 | 42619.42069 |
| 347 | C01092 | 188.1275142 | 135820.8877 | 14.96460748 | 9.779840624 | 18.77726177 | C01092 | 8-Amino-7-oxononanoate | Mass, RT and Spectra | 135745.6732 | 151360.0477 | 123621.7856 | 130439.7471 | 121712.5851 | 130879.7191 |
| 348 | C11111 | 289.0571236 | 6127.679569 | 1181.007704 | 1174.014684 | 1181.656833 | C11111 | Isoprothiolane | Mass, RT and Spectra | 1938.004348 | 10045.1667 | 10142.41763 | 2119.819998 | 4475.366721 | 4102.045076 |
| 349 | C01100 | 220.0479443 | 24665.72519 | 404.8481996 | 399.2910572 | 407.8030135 | C01100 | L-Histidinol phosphate | Mass, RT and Spectra | 4033.659772 | 21733.82993 | 37566.16394 | 4240.86014 | 9246.756387 | 15680.78401 |
| 350 | C01102 | 200.0323636 | 86457.64896 | 66.34000279 | 52.90321189 | 74.34901891 | C01102 | O-Phosphohomoserine | Mass, RT and Spectra | 13272.67368 | 7393.359248 | 15537.24893 | 4154.738754 | 4093.643961 | 4271.493964 |
| 351 | C01103 | 289.0686051 | 15952.28905 | 491.9860535 | 489.9628039 | 495.7367865 | C01103 | Orotidylic acid | Mass, RT and Spectra | 452379.9884 | 38462.9902 | 28026.57436 | 497194.4902 | 775280.4478 | 974332.3885 |
| 352 | C01104 | 76.07572671 | 488553.7407 | 206.1622226 | 186.1219798 | 216.8910985 | C01104 | Trimethylamine N-oxide | Mass, RT and Spectra | 623895.6963 | 384468.0446 | 687508.0568 | 197534.3175 | 255383.4385 | 280452.1488 |
| 353 | C01108 | 125.0239478 | 115090.685 | 422.3050144 | 417.4505498 | 429.4787918 | C01108 | 1,2,3-Trihydroxybenzene | Mass, RT and Spectra | 5291.472492 | 293569.6842 | 197483.0842 | 24869.06831 | 39228.83946 | 34826.61578 |
| 354 | C01118 | 220.0813875 | 993.9137823 | 101.08882 | 97.20529076 | 102.2250551 | C01118 | O-Succinyl-L-homoserine | Mass, RT and Spectra | 2187.434857 | 902.9997269 | 252.2052888 | 757.4348525 | 1578.353237 | 2476.004907 |
| 355 | C01120 | 382.2709257 | 200102.3333 | 855.5099867 | 851.4978382 | 869.3812634 | C01120 | Sphinganine 1-phosphate | Mass, RT and Spectra | 378976.4793 | 301629.5375 | 325185.4296 | 365645.7956 | 308139.2573 | 385326.895 |
| 356 | C01127 | 161.0086185 | 40257.52422 | 267.444262 | 249.3642277 | 278.5802436 | C01127 | 4-Hydroxy-2-oxoglutaric acid | Mass, RT and Spectra | 16368.70041 | 70556.7978 | 74673.1291 | 17790.84464 | 15913.39022 | 10705.71533 |
| 357 | C01136 | 250.0915187 | 942.8561172 | 98.94956462 | 98.30308378 | 99.61602208 | C01136 | S-Acetyldihydrolipoamide | Mass, RT and Spectra | 1983.370471 | 298.0509312 | 1807.413479 | 1298.937455 | 82.62483892 | 0 |
| 358 | C11118 | 100.0757907 | 11651490.13 | 247.1509293 | 240.5625455 | 252.0970739 | C11118 | 2-Pyrrolidinone | Mass, RT and Spectra | 1123477.548 | 3448713.714 | 3734550.817 | 1502032.475 | 1997212.648 | 2065776.188 |
| 359 | HMDB0000422 | 281.0263302 | 68547.82176 | 435.5360837 | 434.1636474 | 437.6935298 | HMDB0000422 | 2-Methylglutaric acid | Mass, RT and Spectra | 29178.15452 | 87962.0379 | 146680.5835 | 42446.18511 | 4084.839275 | 149143.9095 |
| 360 | C01137 | 354.1506549 | 22411.77667 | 444.8911443 | 442.0209808 | 447.071183 | C01137 | S-Adenosylmethioninamine | Mass, RT and Spectra | 1176.813573 | 51548.27317 | 52644.49006 | 1523.016363 | 2513.379928 | 2284.035338 |
| 361 | C01152 | 170.0918142 | 575600.4211 | 1554.722843 | 1552.111234 | 1569.294512 | C01152 | 1-Methylhistidine | Mass, RT and Spectra | 602932.6143 | 484488.3843 | 461962.6053 | 637892.4169 | 661555.9724 | 649689.8804 |
| 362 | C01157 | 176.055767 | 7986.54938 | 426.0257419 | 423.5539134 | 432.463042 | C01157 | 4-Hydroxyproline | Mass, RT and Spectra | 183.7544652 | 12912.28259 | 12230.98983 | 2793.872932 | 4072.945971 | 4408.700753 |
| 363 | C01158 | 331.0693588 | 75782.95594 | 242.0711564 | 226.2074953 | 272.8217989 | C01158 | beta-Glucogallin | Mass, RT and Spectra | 7567.670937 | 64074.83948 | 463473.6073 | 1236.594301 | 474.3931286 | 22276.55196 |
| 364 | HMDB00005 | 101.0240588 | 2833952.628 | 422.6894214 | 418.4677426 | 427.583434 | HMDB00005 | 2-Ketobutyric acid | Mass, RT and Spectra | 351142.0197 | 6939369.626 | 4603285.894 | 822914.5986 | 1160489.537 | 994964.4628 |
| 365 | C11132 | 477.2082856 | 414911.746 | 664.9730671 | 664.0805123 | 667.3985676 | C11132 | 2-Methoxyestrone 3-glucuronide | Mass, RT and Spectra | 368165.2534 | 446915.3384 | 475160.2921 | 347642.9618 | 362428.442 | 343879.2792 |
| 366 | C11133 | 447.1997949 | 4374.105116 | 742.5833515 | 739.3291717 | 743.0694603 | C11133 | Estrone glucuronide | Mass, RT and Spectra | 4346.479248 | 3727.923199 | 4770.219642 | 4706.196584 | 5128.764199 | 4315.956387 |
| 367 | C01169 | 306.0857678 | 19898.1805 | 403.2231794 | 400.4469994 | 406.3380877 | C01169 | (S)-Succinyldihydrolipoamide | Mass, RT and Spectra | 1748.798437 | 141.1094689 | 92.97908153 | 96.79167002 | 110.3786833 | 0 |
| 368 | C11134 | 465.2480674 | 29881.52147 | 540.5005498 | 533.9774254 | 551.2839681 | C11134 | Testosterone glucuronide | Mass, RT and Spectra | 45058.33657 | 13713.30998 | 14883.21377 | 43880.27411 | 40747.46325 | 39056.65036 |
| 369 | C11136 | 467.2618714 | 12714.52893 | 735.0596621 | 732.4348893 | 736.5986509 | C11136 | Etiocholanolone glucuronide | Mass, RT | 19649.94241 | 3401.946967 | 4171.851838 | 18300.91255 | 13847.90826 | 14951.31303 |
| 370 | C01176 | 331.225932 | 27878.23388 | 8.939603272 | 4.037333918 | 11.29146796 | C01176 | 17-Hydroxyprogesterone | Mass, RT and Spectra | 5879.22273 | 63877.8 | 49080.44612 | 5967.249988 | 15329.96289 | 16784.25892 |
| 371 | C01187 | 239.0769641 | 141896.8757 | 536.3397945 | 531.8877452 | 539.2171172 | C01187 | 3-Deoxy-D-manno-octulosonate | Mass, RT and Spectra | 61689.07849 | 74617.33343 | 60276.42146 | 78393.81601 | 435855.7917 | 1188484.042 |
| 372 | C11150 | 161.9046317 | 21802.58312 | 375.4689957 | 364.981912 | 382.3074658 | C11150 | Trichloroacetic acid | Mass, RT and Spectra | 53.53804055 | 45191.65003 | 47772.24172 | 4438.552974 | 3720.048174 | 5667.160099 |
| 373 | C01190 | 700.572305 | 235151.898 | 1172.603766 | 1139.538459 | 1194.740983 | C01190 | Glucosylceramide | Mass, RT and Spectra | 83221250.74 | 37803860.08 | 33460139.03 | 96512040.35 | 92240894.7 | 105346856.5 |
| 374 | C01192 | 407.2196435 | 11441.1827 | 101.6116398 | 86.70229436 | 103.7797237 | C01192 | 1-palmitoylglycerone 3-phosphate | Mass, RT and Spectra | 161.6253785 | 18608.34222 | 26093.12655 | 403.2188744 | 378.2711908 | 668.0429003 |
| 375 | C11154 | 370.186172 | 34661.7221 | 784.090878 | 780.9275268 | 785.1147568 | C11154 | Trimetrexate | Mass, RT and Spectra | 53276.19222 | 15812.46444 | 15086.94086 | 48558.03884 | 46461.89487 | 46555.79244 |
| 376 | C01210 | 156.0417105 | 28636933.07 | 117.4933317 | 112.9230291 | 135.5999958 | C01210 | N-Methylethanolamine phosphate | Mass, RT and Spectra | 29789189.85 | 27395530.8 | 21961116.47 | 36835432.76 | 36344033.02 | 35121119.85 |
| 377 | C01225 | 335.0734058 | 15907.13089 | 80.26338324 | 76.83357591 | 99.69999501 | C01225 | 1-(sn-Glycero-3-phospho)-1D-myo-inositol | Mass, RT and Spectra | 23507.34831 | 6615.373869 | 9789.393874 | 21305.14655 | 20988.7568 | 25498.35988 |
| 378 | C11263 | 226.1080149 | 462889.8958 | 1553.372561 | 1550.663894 | 1566.288904 | C11263 | Cytokinin B | Mass, RT and Spectra | 370101.3692 | 518945.3183 | 531630.4175 | 388490.3888 | 518169.1365 | 402313.1145 |
| 379 | C01226 | 291.1989554 | 586513.8807 | 1178.469166 | 1172.452768 | 1179.3336 | C01226 | 12-OPDA | Mass, RT and Spectra | 180560.0835 | 949129.1407 | 1204168.556 | 189691.4017 | 200111.6303 | 297949.6449 |
| 380 | C11289 | 528.1701078 | 8705.14854 | 521.1405404 | 518.5490121 | 525.1118725 | C11289 | 17-beta-estradiol 3-sulfate-17-(beta-D-glucuronide) | Mass, RT and Spectra | 9689.435946 | 4789.300421 | 1879.179895 | 15793.42199 | 20335.43743 | 21707.66119 |
| 381 | C11301 | 513.2802321 | 1057.434695 | 1596.69812 | 1594.385652 | 1598.634325 | C11301 | Sulfolithocholylglycine | Mass, RT and Spectra | 1673.023267 | 503.8273718 | 471.6642299 | 1447.210342 | 1555.213496 | 941.7887254 |
| 382 | C01227 | 289.2154048 | 28294.12938 | 9.765575989 | 7.126695731 | 15.66772232 | C01227 | Dehydroepiandrosterone | Mass, RT and Spectra | 4677.682227 | 61573.84718 | 54805.67304 | 6085.487181 | 14799.7596 | 15350.6333 |
| 383 | C11352 | 148.9932483 | 7839.584867 | 1640.489443 | 1635.762278 | 1643.946024 | C11352 | 5,6-Dichloro-1,3-cyclohexadiene | Mass, RT and Spectra | 13289.86981 | 3157.456181 | 2802.011888 | 11491.5386 | 11130.55258 | 9933.190048 |
| 384 | C11404 | 186.1235791 | 1274.592263 | 1214.192938 | 1211.658817 | 1216.727059 | C11404 | (3R)-6-Hydroxy-3-isopropenyl-heptanoate | Mass, RT and Spectra | 3352.327063 | 357.617692 | 325.7903542 | 965.5092253 | 1431.813721 | 1112.061374 |
| 385 | C01230 | 587.3314884 | 13090.80224 | 616.4767611 | 613.0373918 | 619.0477892 | C01230 | All-trans-hexaprenyl diphosphate | Mass, RT and Spectra | 19195.08497 | 6704.053098 | 7136.740141 | 16883.29689 | 15270.77894 | 17620.7593 |
| 386 | C11434 | 217.0466009 | 71415.46733 | 135.3803653 | 125.0613724 | 144.8817681 | C11434 | 2-C-Methyl-D-erythritol 4-phosphate | Mass, RT and Spectra | 80003.66944 | 53080.6109 | 54970.65115 | 87970.22739 | 73740.74878 | 75614.95932 |
| 387 | C01233 | 216.0626054 | 7179.95872 | 446.7158858 | 446.4101987 | 448.1448136 | C01233 | Glycerylphosphorylethanolamine | Mass, RT | 288.5013585 | 16935.77214 | 15417.01431 | 376.7886498 | 3281.265993 | 3535.159722 |
| 388 | C11457 | 165.0550749 | 218546.8091 | 155.4774844 | 136.8376926 | 166.0129293 | C11457 | 3-(3-Hydroxyphenyl)propanoic acid | Mass, RT and Spectra | 1267.873523 | 56040.57383 | 47714.28546 | 6399.223686 | 9618.261719 | 11542.67741 |
| 389 | C11516 | 347.0388588 | 10605.95636 | 508.9249145 | 506.5013297 | 513.4632359 | C11516 | 2-(alpha-D-Mannosyl)-3-phosphoglycerate | Mass, RT and Spectra | 233.4482853 | 13057.42581 | 36857.77033 | 161.6536925 | 128.8062274 | 369.3794249 |
| 390 | C01234 | 100.0400283 | 67846.95042 | 470.0359106 | 464.2368207 | 476.3326654 | C01234 | 1-Aminocyclopropanecarboxylic acid | Mass, RT and Spectra | 932.7298563 | 144669.9425 | 147373.1914 | 13099.13755 | 16369.12042 | 19390.84069 |
| 391 | C01236 | 258.0141372 | 13672.08854 | 313.818441 | 310.6628667 | 319.0581593 | C01236 | 6-Phosphonoglucono-D-lactone | Mass, RT | 1008.851928 | 28991.13239 | 33905.07737 | 328.6563657 | 554.3680866 | 1459.727601 |
| 392 | C01239 | 221.1129396 | 5041.322049 | 7.732541149 | 4.647799801 | 10.78802215 | C01239 | N-Acetyl-b-glucosaminylamine | Mass, RT and Spectra | 71.62234304 | 8693.595235 | #DIV/0! | 2549.897976 | 4207.149314 | 3408.089756 |
| 393 | C11811 | 263.0075382 | 217091.7797 | 98.2507348 | 95.21368912 | 103.9668558 | C11811 | 1-Hydroxy-2-methyl-2-butenyl 4-diphosphate | Mass, RT and Spectra | 310935.4123 | 211678.9371 | 133373.4481 | 217072.9455 | 228535.6555 | 143381.8531 |
| 394 | C01243 | 420.9680319 | 1641.07976 | 160.0230572 | 155.4831902 | 163.1173381 | C01243 | Inositol 1,3,4-trisphosphate | Mass, RT and Spectra | 109.1314383 | 24866.26819 | #DIV/0! | 4225.601695 | 949.6945569 | 2094.91058 |
| 395 | C16741 | 143.0817725 | 57660.45323 | 517.2565915 | 514.0231235 | 522.0509349 | C16741 | 5-Hydroxylysine | Mass, RT and Spectra | 8417.85704 | 109946.7802 | 125224.0672 | 25181.12152 | 16357.37242 | 23566.06494 |
| 396 | C01250 | 172.060919 | 8256.045959 | 153.6425872 | 149.4521954 | 160.2530624 | C01250 | N-Acetyl-L-glutamate 5-semialdehyde | Mass, RT and Spectra | 1726.438604 | 17959.15481 | 11535.35002 | 686.0289162 | 16059.77695 | 12467.86915 |
| 397 | C01252 | 208.0619526 | 17707.63578 | 592.5862023 | 591.5922305 | 593.5293422 | C01252 | 4-(2-Aminophenyl)-2,4-dioxobutanoic acid | Mass, RT and Spectra | 4250.674323 | 41748.0771 | 26661.02166 | 11454.70412 | 12334.40021 | 20777.87476 |
| 398 | C01269 | 325.0338984 | 8967.356905 | 99.45445865 | 96.46833792 | 100.5345668 | C01269 | 5-O-(1-Carboxyvinyl)-3-phosphoshikimate | Mass, RT and Spectra | 17295.20203 | 1632.181889 | 2302.615239 | 14517.62425 | 6789.465027 | 15373.20102 |
| 399 | C11821 | 183.0171385 | 282022.6059 | 421.6908772 | 401.4514954 | 426.3444817 | C11821 | 5-Hydroxyisourate | Mass, RT and Spectra | 71525.07532 | 518719.2699 | 521201.9183 | 81666.07124 | 126436.2667 | 138308.3187 |
| 400 | C01273 | 217.0522835 | 78555.55434 | 251.0504703 | 236.0980597 | 261.5217337 | C01273 | 2-Hydroxy-6-oxo-6-phenylhexa-2,4-dienoate | Mass, RT and Spectra | 29877.24764 | 134225.4295 | 148157.3419 | 21913.24887 | 38998.92735 | 55593.69352 |
| 401 | C01290 | 806.5676941 | 1229800.658 | 1084.351359 | 1073.322533 | 1112.762934 | C01290 | Lactosylceramide (d18:1/12:0) | Mass, RT and Spectra | 49815.5434 | 13553.66739 | 41554.95111 | 57177.45731 | 35741.02905 | 16964.51993 |
| 402 | C01292 | 407.2780103 | 52020.18184 | 912.4387217 | 909.4783252 | 915.4929766 | C01292 | 3,7-Dihydroxy-12-oxocholanoic acid | Mass, RT and Spectra | 51479.9 | 56231.36906 | 49045.11496 | 43600.95949 | 46518.51338 | 45005.9049 |
| 403 | C01302 | 348.0514183 | 6308.081062 | 287.4291421 | 281.9975245 | 290.8890144 | C01302 | 1-(2-Carboxyphenylamino)-1-deoxy-D-ribulose 5-phosphate | Mass, RT and Spectra | 757.1592481 | 26019.40321 | 7487.515445 | 325.6767838 | 3500.795695 | 10830.7052 |
| 404 | C11839 | 214.1079941 | 1019450.798 | 1553.398056 | 1551.90499 | 1555.013576 | C11839 | N-(3-oxo-hexanoyl)-homoserine lactone | Mass, RT and Spectra | 852047.9713 | 1140236.21 | 1012861.489 | 808070.4865 | 1034035.466 | 923285.281 |
| 405 | C01344 | 412.0164719 | 2910359.34 | 115.3490244 | 112.9639073 | 118.0154388 | C01344 | dIDP | Mass, RT and Spectra | 26969.83746 | 16354.35031 | 8245.278607 | 35746.79437 | 45814.12541 | 40509.97625 |
| 406 | C01345 | 244.9844582 | 182844.7012 | 314.9438521 | 292.6073965 | 333.5556741 | C01345 | 2'-Deoxyinosine triphosphate | Mass, RT | 23079.55482 | 340953.0395 | 346194.01 | 34821.64577 | 51713.84296 | 91436.22241 |
| 407 | C01367 | 348.8883313 | 25161.65579 | 931.4971221 | 894.0202739 | 958.6064327 | C01367 | 3'-AMP | Mass, RT and Spectra | 9845.398377 | 31061.53962 | 58580.74392 | 5321.669766 | 11273.60505 | 7477.206627 |
| 408 | C01384 | 115.003319 | 418006.4167 | 490.9387254 | 484.9701049 | 493.657277 | C01384 | Maleic acid | Mass, RT and Spectra | 108977.9038 | 666232.9897 | 797099.3787 | 170448.3132 | 202159.6798 | 190489.4957 |
| 409 | C11844 | 214.1437835 | 29726.21927 | 12.13085197 | 7.03049596 | 17.64262501 | C11844 | N-heptanoyl-homoserine lactone | Mass, RT and Spectra | 22984.219 | 37533.14683 | 32544.73542 | 27903.29951 | 28839.49119 | 25824.75528 |
| 410 | C11849 | 285.2416995 | 17545.48868 | 1178.436693 | 1172.452768 | 1181.415651 | C11849 | 3-hydroxypalmitic acid methyl ester | Mass, RT and Spectra | 1249.198462 | 36184.37873 | 35195.87984 | 4929.796015 | 8741.096965 | 7083.243586 |
| 411 | C01419 | 88.01223633 | 35440.50683 | 236.8417084 | 231.8871233 | 249.6219589 | C01419 | Cysteinylglycine | Mass, RT | 56.58554438 | 64263.8607 | 83121.41817 | 173.6556599 | 6468.342702 | 12856.44413 |
| 412 | C01475 | 144.0661094 | 13173.25758 | 341.4369236 | 335.1582996 | 343.3775264 | C01475 | Allysine | Mass, RT and Spectra | 248558.3205 | 576315.1726 | 571085.5269 | 325964.2965 | 290393.6017 | 332869.7573 |
| 413 | C11896 | 329.2475432 | 289968.6023 | 90.72193953 | 89.71216493 | 93.6806528 | C11896 | Taxa-4(20),11(12)-dien-5alpha-yl acetate | Mass, RT and Spectra | 62428.41207 | 488038.1744 | 455210.3662 | 98646.96511 | 102141.357 | 120176.5413 |
| 414 | C11924 | 165.0915476 | 855917.7475 | 100.5187846 | 94.63884532 | 106.7642134 | C11924 | Perillic acid | Mass, RT and Spectra | 125527.301 | 1571936.816 | 1585581.047 | 226296.2739 | 417398.8466 | 494473.062 |
| 415 | HMDB0000555 | 399.0769808 | 23193.40923 | 108.3313385 | 94.5413598 | 112.6051213 | HMDB0000555 | 3-Methyladipic acid | Mass, RT and Spectra | 3797.450707 | 49155.21785 | 30210.52589 | 8881.954617 | 14836.66257 | 10973.42985 |
| 416 | C01479 | 165.0550863 | 344646.9917 | 118.402002 | 116.051522 | 135.3976705 | C01479 | Phenyllactic acid | Mass, RT and Spectra | 63114.45837 | 542916.3028 | 889812.5104 | 93764.23719 | 112034.8536 | 137586.5162 |
| 417 | C01512 | 377.1413756 | 233982.7618 | 426.6723449 | 424.2113645 | 439.6186473 | C01512 | Loganate | Mass, RT and Spectra | 252583.5942 | 215103.8008 | 205556.0269 | 236088.7002 | 281088.8671 | 287551.0104 |
| 418 | C11989 | 373.2618001 | 11201.82165 | 408.5015917 | 406.1499951 | 410.4157386 | C11989 | 8,8a-Deoxyoleandolide | Mass, RT and Spectra | 3576.30125 | 17969.73312 | 16615.09963 | 5751.933723 | 8038.755541 | 6729.882262 |
| 419 | C11992 | 531.3133759 | 100685.6232 | 682.1999788 | 679.9550015 | 684.541563 | C11992 | L-Oleandrosyl-oleandolide | Mass, RT and Spectra | 130613.4703 | 71285.24334 | 75242.83407 | 108469.3153 | 110158.5101 | 115266.6244 |
| 420 | C01530 | 283.2636183 | 12698104.78 | 93.20238969 | 92.77940146 | 93.99765235 | C01530 | Stearic acid | Mass, RT and Spectra | 2289250.329 | 23248053.74 | 24370421.97 | 3615158.307 | 4913247.31 | 6227075.912 |
| 421 | C01545 | 129.1272112 | 79197.75219 | 1764.0857 | 1755.462643 | 1780.51607 | C01545 | Octanal | Mass, RT and Spectra | 48490.29598 | 112390.1001 | 104544.4384 | 59034.21767 | 64195.94162 | 58352.81337 |
| 422 | C01551 | 261.0009987 | 318205.4989 | 421.4004723 | 413.0798358 | 425.8928717 | C01551 | Allantoin | Mass, RT and Spectra | 22526.06272 | 693515.784 | 567998.5705 | 83042.36521 | 141385.5846 | 136827.3168 |
| 423 | C01571 | 171.1385539 | 737622.5777 | 97.51116359 | 89.76343617 | 103.0083117 | C01571 | Capric acid | Mass, RT and Spectra | 96236.68067 | 1347950.724 | 1515405.992 | 149029.7144 | 375165.9643 | 196026.9476 |
| 424 | C01585 | 115.076002 | 172471.142 | 113.0059646 | 93.23059318 | 114.7093341 | C01585 | Caproic acid | Mass, RT and Spectra | 23717.30601 | 318150.4098 | 395120.6968 | 24043.94882 | 54580.91304 | 50225.90369 |
| 425 | C01586 | 178.0504009 | 316828.2966 | 138.8240374 | 128.7814765 | 159.6456403 | C01586 | Hippuric acid | Mass, RT and Spectra | 44969.11521 | 1007816.909 | 349367.1771 | 37529.47553 | 101515.367 | 69949.52352 |
| 426 | C01595 | 279.2323563 | 16100897.09 | 91.93672529 | 91.07321178 | 95.28858649 | C01595 | Linoleic acid | Mass, RT and Spectra | 3424571.5 | 24340441.04 | 25318870.59 | 6431339.688 | 4399806.485 | 11496409.77 |
| 427 | C12002 | 598.3528833 | 55021.79451 | 633.5578601 | 629.6704249 | 641.3884775 | C12002 | 5-O-beta-D-Mycaminosyltylonolide | Mass, RT and Spectra | 73101.96338 | 39611.20878 | 37746.29295 | 54688.32974 | 57147.67454 | 65889.5771 |
| 428 | C01596 | 114.0192659 | 639442.7759 | 145.6648914 | 128.6847893 | 170.6236627 | C01596 | Maleamate | Mass, RT and Spectra | 91577.00763 | 1210083.89 | 1277596.002 | 99645.8853 | 232970.8921 | 338210.623 |
| 429 | C01606 | 165.018599 | 50216.14005 | 425.8521215 | 413.8142348 | 432.9301124 | C01606 | Phthalic acid | Mass, RT and Spectra | 5649.977373 | 91054.21221 | 113549.3996 | 9016.745292 | 12564.68097 | 22557.55971 |
| 430 | C01613 | 667.2278643 | 55007.73436 | 834.3812094 | 825.202962 | 851.6398685 | C01613 | Stachyose | Mass, RT and Spectra | 25081.83397 | 81973.03663 | 29586.20542 | 44327.94165 | 111796.0478 | 69689.25065 |
| 431 | C01674 | 425.1785911 | 30517.6945 | 662.2258284 | 658.733413 | 682.5089915 | C01674 | Chitobiose | Mass, RT and Spectra | 40374.4023 | 21687.08163 | 19603.58329 | 38939.77936 | 41528.87268 | 41631.74114 |
| 432 | C01683 | 263.1236539 | 1578.294074 | 98.68786526 | 98.68784493 | 98.68788559 | C01683 | Nopalinic acid | Mass, RT and Spectra | 2191.306446 | 1505.511828 | 864.5926617 | 5190.916397 | 563.6918212 | 2108.56258 |
| 433 | C01801 | 90.02739038 | 5386624.179 | 233.2108454 | 210.1248017 | 246.381373 | C01801 | Deoxyribose | Mass, RT and Spectra | 644477.8989 | 9476542.742 | 11520434.38 | 676580.2854 | 1718871.672 | 2430966.746 |
| 434 | C01838 | 269.2828517 | 20737.19442 | 955.8482883 | 953.7326718 | 956.6248898 | C01838 | Stearaldehyde | Mass, RT and Spectra | 5175.347597 | 31858.89754 | 34286.98838 | 12810.90734 | 23501.90175 | 11510.41304 |
| 435 | C12027 | 216.1238715 | 1052773.683 | 1553.559345 | 1551.061289 | 1554.979514 | C12027 | 2-amino-8-oxo-9,10-epoxy-decanoic acid | Mass, RT and Spectra | 993614.294 | 1158240.813 | 1157763.281 | 873226.6431 | 1341246.777 | 896236.398 |
| 436 | C01879 | 130.0494813 | 2372856.569 | 102.5495905 | 98.51204338 | 109.7408314 | C01879 | Pyroglutamic acid | Mass, RT and Spectra | 3499727.484 | 1920239.574 | 1808275.602 | 2690193.805 | 2730214.256 | 2095093.068 |
| 437 | C01921 | 464.2996162 | 2006989.905 | 111.7477909 | 98.25480193 | 124.0562236 | C01921 | Glycocholic acid | Mass, RT and Spectra | 567929.1041 | 3759271.704 | 3314225.412 | 588640.7915 | 796348.496 | 1055906.241 |
| 438 | C01924 | 187.1192935 | 15329.39516 | 594.1061707 | 573.7829039 | 612.1611407 | C01924 | Homo-L-arginine | Mass, RT and Spectra | 2675.094185 | 34263.28779 | 26675.40473 | 2424.796407 | 2709.379581 | 392.4987072 |
| 439 | C01933 | 188.0445969 | 388387.872 | 231.9907336 | 219.8122002 | 245.8470893 | C01933 | L-Norleucine | Mass, RT and Spectra | 128935.6901 | 543384.673 | 724231.1234 | 140540.4304 | 205024.8355 | 229547.8044 |
| 440 | HMDB0000622 | 263.0763032 | 16836.75856 | 441.9415438 | 440.6776842 | 443.6857613 | HMDB0000622 | Ethylmalonic acid | Mass, RT and Spectra | 151.8059534 | 37412.09824 | 36308.33532 | 3822.763617 | 3823.025072 | 4610.204281 |
| 441 | C01957 | 373.1119912 | 2172.792492 | 542.0265516 | 535.3964447 | 543.0395771 | C01957 | Secologanate | Mass, RT and Spectra | 280.833659 | 5633.357558 | 4249.683103 | 223.5157922 | 316.4053564 | 45.2628122 |
| 442 | C01984 | 210.993929 | 9181.470572 | 156.836362 | 152.6771479 | 162.8051549 | C01984 | Mandelic acid | Mass, RT and Spectra | 2166.697391 | 14055.83832 | 14547.0904 | 995.7178964 | 8534.519643 | 10761.98743 |
| 443 | C01990 | 207.013341 | 125237.5137 | 106.2808402 | 104.3756278 | 109.2558791 | C01990 | 3-Oxalomalate | Mass, RT and Spectra | 165306.9846 | 120868.7003 | 110000.3176 | 126297.639 | 134953.2952 | 93590.8352 |
| 444 | C01991 | 105.054496 | 672810.4434 | 175.1575356 | 168.359349 | 183.2448262 | C01991 | 4-Hydroxybutyric acid | Mass, RT and Spectra | 268308.8757 | 737694.9342 | 703042.5868 | 483212.7722 | 426153.7056 | 359316.2818 |
| 445 | C02037 | 131.0457508 | 557722.1366 | 414.5694831 | 412.2943153 | 417.2495465 | C02037 | Glycyl-glycine | Mass, RT and Spectra | 103052.879 | 1161454.561 | 1115234.452 | 132993.4815 | 239385.477 | 247258.9011 |
| 446 | C02043 | 204.0658835 | 306409.4248 | 161.8762746 | 142.8937298 | 175.1380029 | C02043 | Indolelactic acid | Mass, RT and Spectra | 64415.29525 | 570791.337 | 660516.8256 | 55889.67292 | 193188.9594 | 252763.9371 |
| 447 | C12100 | 267.2325293 | 291082.5686 | 92.15795931 | 90.08133934 | 93.2359208 | C12100 | Cyclohexaneundecanoic acid | Mass, RT and Spectra | 54136.05175 | 396417.145 | 471006.1318 | 123976.7302 | 69054.76408 | 92603.17633 |
| 448 | C12106 | 286.0344123 | 10320.41414 | 515.1896213 | 512.7158784 | 516.9942343 | C12106 | AminoDAHP | Mass, RT and Spectra | 186.9038158 | 23871.02712 | 23682.90727 | 4405.529072 | 1098.651604 | 2029.915349 |
| 449 | C12144 | 316.2843029 | 2986483.587 | 790.1740724 | 783.2743661 | 791.6708116 | C12144 | Phytosphingosine | Mass, RT and Spectra | 4204.01373 | 27703.63412 | 45389.39733 | 3478.26565 | 1321.944446 | 992.3046974 |
| 450 | C02059 | 451.3564383 | 11701.24355 | 889.4594664 | 887.9491501 | 890.6238282 | C02059 | Vitamin K1 | Mass, RT and Spectra | 232116.8962 | 205928.3692 | 150911.6877 | 325957.601 | 208329.0782 | 140847.551 |
| 451 | C02067 | 243.0613422 | 294534.7105 | 333.6116177 | 307.4392137 | 348.0465639 | C02067 | Pseudouridine | Mass, RT and Spectra | 69921.77177 | 504427.1903 | 517715.4586 | 108861.0681 | 193750.5641 | 184276.928 |
| 452 | C02076 | 209.0664285 | 20984.48441 | 376.9905151 | 367.6621025 | 390.5764407 | C02076 | Sedoheptulose | Mass, RT and Spectra | 171632.1051 | 508939.8851 | 473062.8468 | 197004.7127 | 257793.2799 | 218353.5481 |
| 453 | C12286 | 193.1582982 | 32069.89915 | 11.38811586 | 6.883007426 | 15.12358053 | C12286 | alpha-Ionone | Mass, RT and Spectra | 12092.88081 | 57478.62925 | 51433.7663 | 14314.90118 | 20952.90928 | 18806.0092 |
| 454 | C12294 | 183.137394 | 53053.59033 | 83.99778996 | 81.70769862 | 90.01489963 | C12294 | Neryl formate | Mass, RT and Spectra | 38053.7589 | 71013.54835 | 66484.37343 | 43461.11841 | 48084.58561 | 41973.43291 |
| 455 | C02140 | 347.2211059 | 7252.926844 | 8.544468281 | 6.344724351 | 13.41506568 | C02140 | Corticosterone | Mass, RT and Spectra | 2174.269604 | 16007.32925 | 12062.42778 | 2566.868414 | 2074.524302 | 2956.706419 |
| 456 | C02147 | 207.0504404 | 18562.12265 | 383.2483896 | 368.7734679 | 392.1375626 | C02147 | Dihydrolipoate | Mass, RT and Spectra | 575.7303278 | 27887.7961 | 47700.02255 | 855.3334466 | 3097.998417 | 5215.287924 |
| 457 | C12452 | 184.1329245 | 37299.37546 | 12.58962846 | 7.99733596 | 15.73383465 | C12452 | Acetyltropine | Mass, RT and Spectra | 31049.61365 | 42838.49186 | 40077.84556 | 40916.3024 | 37414.13318 | 35357.56361 |
| 458 | C02166 | 624.2932433 | 3076730.465 | 961.2290341 | 958.7881436 | 964.6125543 | C02166 | Leukotriene C4 | Mass, RT and Spectra | 69183.86506 | 46589.57801 | 70989.84489 | 55484.84416 | 64359.22393 | 44454.94286 |
| 459 | C12625 | 531.1194424 | 5362.381062 | 150.3324842 | 145.4123147 | 158.0657229 | C12625 | Biochanin A 7-(6-malonylglucoside) | Mass, RT and Spectra | 302.0904293 | 10499.29605 | 5468.378085 | 1312.6364 | 4957.647801 | 4639.472477 |
| 460 | C02170 | 119.0335667 | 52402.48698 | 246.6627058 | 233.0047637 | 262.6456967 | C02170 | Methylmalonic acid | Mass, RT and Spectra | 3965.927381 | 95337.1198 | 77662.18797 | 16802.31303 | 33135.27477 | 17541.06177 |
| 461 | C12832 | 214.9425262 | 26236.68959 | 94.64459086 | 81.15275965 | 96.95734081 | C12832 | 3,4,6-Trichloro-cis-1,2-dihydroxycyclohexa-3,5-diene | Mass, RT and Spectra | 25840.43377 | 28532.79338 | 31202.19061 | 21040.75689 | 24414.44325 | 23302.37225 |
| 462 | C02197 | 299.1642735 | 7937.308977 | 104.5229348 | 98.53965192 | 105.4354408 | C02197 | Testolactone | Mass, RT and Spectra | 336.2199055 | 16104.08534 | 16031.10406 | 829.0708096 | 2064.630253 | 2333.703654 |
| 463 | C02217 | 185.1175663 | 179543.2584 | 98.28328674 | 95.93754663 | 105.0395066 | C02217 | 10-oxo-decanoic acid | Mass, RT and Spectra | 23368.15943 | 334036.4442 | 368895.2751 | 41482.45159 | 82855.58845 | 96517.23032 |
| 464 | C02237 | 129.038255 | 345330.5782 | 264.4084522 | 246.1614055 | 274.6256426 | C02237 | Pyrrolidonecarboxylic acid | Mass, RT and Spectra | 61716.36936 | 589377.363 | 702975.1678 | 63225.37741 | 122497.3866 | 95893.21258 |
| 465 | C02242 | 166.0719332 | 271692.9388 | 269.8258476 | 262.5157558 | 279.182128 | C02242 | 7-Methylguanine | Mass, RT and Spectra | 369376.6796 | 258974.6672 | 194111.2197 | 415168.9001 | 399912.7348 | 455365.9872 |
| 466 | C02272 | 91.05818884 | 31557088.29 | 109.5070438 | 106.1761331 | 121.5513161 | C02272 | Diethyl sulfide | Mass, RT and Spectra | 22106396.77 | 40064095.96 | 39198598.35 | 25086893.65 | 29123139.74 | 26069939.68 |
| 467 | C02287 | 118.9987703 | 285459.6629 | 515.7066701 | 513.5388579 | 518.9939304 | C02287 | Hydroxypropanedioic acid | Mass, RT and Spectra | 29192.24147 | 542447.9435 | 617131.9478 | 43678.32666 | 74197.23324 | 87085.56508 |
| 468 | C02291 | 221.0613267 | 25430.43672 | 481.512397 | 469.2242412 | 483.4663171 | C02291 | L-Cystathionine | Mass, RT and Spectra | 895.7160811 | 55979.97791 | 55323.33689 | 6455.226335 | 13877.18497 | 11930.09302 |
| 469 | C02355 | 305.0165949 | 31021.26804 | 470.2163831 | 467.6779319 | 471.3264783 | C02355 | Uridine 2',3'-cyclic phosphate | Mass, RT and Spectra | 8539.242767 | 60841.71908 | 44206.01735 | 14749.24609 | 10503.92338 | 16192.97701 |
| 470 | HMDB0000678 | 158.0816389 | 17370.76405 | 146.8546411 | 139.0716957 | 156.5195408 | HMDB0000678 | Isovalerylglycine | Mass, RT and Spectra | 513.9717957 | 34330.85605 | 42427.52017 | 906.5874594 | 4937.040525 | 5842.09073 |
| 471 | C12835 | 226.9506606 | 48778760.31 | 123.5076799 | 118.8318727 | 139.1740997 | C12835 | 2,5-Dichloro-4-oxohex-2-enedioate | Mass, RT and Spectra | 57704673.22 | 45209513.91 | 41254498.52 | 52086595.96 | 51408562.13 | 46098206.71 |
| 472 | HMDB0000695 | 177.0668234 | 63581.02155 | 114.139106 | 101.3137005 | 118.7417098 | HMDB0000695 | Ketoleucine | Mass, RT and Spectra | 5329.269374 | 130490.9867 | 110625.5077 | 19375.52678 | 26650.01767 | 46102.5966 |
| 473 | C02356 | 104.0706108 | 6354496.644 | 11.80520093 | 2.993151608 | 19.64263107 | C02356 | L-Alpha-aminobutyric acid | Mass, RT and Spectra | 1951812.913 | 12168793.93 | 11030698.3 | 2684675.555 | 3938796.474 | 4331500.673 |
| 474 | C02367 | 213.1489893 | 94367.18987 | 97.19141822 | 95.01401115 | 99.42216534 | C02367 | 3-Oxododecanoic acid | Mass, RT and Spectra | 16561.55612 | 157941.1545 | 161613.535 | 36366.14749 | 29580.87889 | 31629.71837 |
| 475 | C02376 | 126.0659604 | 4442550.407 | 1553.46195 | 1549.94279 | 1575.825177 | C02376 | 5-Methylcytosine | Mass, RT and Spectra | 5712104.725 | 3783542.407 | 3647340.672 | 4369873.974 | 4670809.335 | 5515003.044 |
| 476 | C16884 | 123.0663494 | 304939.9933 | 8.021769968 | 4.720976144 | 13.34010088 | C16884 | D-Threitol | Mass, RT and Spectra | 112806.4227 | 532896.6839 | 420618.7234 | 154406.3399 | 218543.3619 | 240386.7119 |
| 477 | C02378 | 132.1018582 | 106617481.1 | 358.3064733 | 345.2787252 | 370.0935387 | C02378 | Aminocaproic acid | Mass, RT and Spectra | 90570827.38 | 136267479.8 | 137355409.2 | 83134517.27 | 95533882.41 | 87598929.6 |
| 478 | C12888 | 463.2316157 | 31062.35206 | 770.6149339 | 767.2114118 | 773.5992504 | C12888 | Hydrocortisone succinate | Mass, RT and Spectra | 13783.17303 | 55661.20934 | 36908.23262 | 17096.04075 | 23877.71931 | 31835.76463 |
| 479 | C12889 | 186.1498946 | 9447.463291 | 1178.956031 | 1173.222 | 1181.805449 | C12889 | n-decanohydroxamic acid | Mass, RT and Spectra | 62.53635318 | 22280.67777 | 16665.07553 | 4135.224246 | 7645.430616 | 4787.475353 |
| 480 | C02427 | 188.1033746 | 6674.172587 | 430.8431454 | 429.2477941 | 432.6490809 | C02427 | Homocitrulline | Mass, RT and Spectra | 1495.465287 | 12134.49471 | 15809.96424 | 2082.450922 | 3456.755827 | 4227.719773 |
| 481 | C02470 | 365.0063411 | 46494.29381 | 464.912732 | 463.3740618 | 466.1704256 | C02470 | Xanthurenic acid | Mass, RT and Spectra | 0 | 103721.1579 | 99338.58638 | 9382.444208 | 6312.580113 | 12685.95497 |
| 482 | C02479 | 147.0294186 | 243459.7903 | 400.8022439 | 394.8863759 | 419.5625527 | C02479 | L-Arabinose | Mass, RT and Spectra | 52963.28646 | 439836.7317 | 456268.46 | 80827.51933 | 111825.5437 | 125691.872 |
| 483 | C12986 | 204.0858571 | 353517.4775 | 279.9941942 | 253.1109732 | 291.7635202 | C12986 | N2-Acetyl-L-aminoadipate | Mass, RT and Spectra | 70772.29138 | 903301.3308 | 563984.3161 | 116333.3348 | 236913.2465 | 175754.7387 |
| 484 | C02494 | 282.1189265 | 265626.8939 | 287.4397772 | 276.4372036 | 300.0999715 | C02494 | 1-Methyladenosine | Mass, RT and Spectra | 10268.24126 | 240940.5925 | 268515.4874 | 13757.753 | 38745.18745 | 26135.64821 |
| 485 | C02502 | 96.04432201 | 62937.25764 | 163.7163488 | 158.9831239 | 167.8321211 | C02502 | 2-Hydroxypyridine | Mass, RT and Spectra | 65691.7092 | 66916.78254 | 59101.66955 | 45695.6913 | 40209.08327 | 45541.7903 |
| 486 | C02504 | 177.074738 | 88125.82141 | 1233.075851 | 1225.373573 | 1254.612662 | C02504 | 2-Isopropylmalic acid | Mass, RT and Spectra | 90752.65299 | 83947.52855 | 85142.62804 | 81542.99706 | 90383.84635 | 89722.00573 |
| 487 | C02505 | 136.0753824 | 485021.2434 | 321.161982 | 302.4415425 | 327.6670429 | C02505 | 2-Phenylacetamide | Mass, RT and Spectra | 658131.5431 | 486553.3922 | 504019.3645 | 404816.5196 | 336790.2025 | 381023.1581 |
| 488 | C02513 | 463.3034036 | 24596.57801 | 724.8490322 | 723.5384647 | 725.9028328 | C02513 | 3-Dehydroecdysone | Mass, RT and Spectra | 15395.42635 | 39714.79726 | 32946.80744 | 20928.08827 | 35957.79535 | 26112.89031 |
| 489 | C13309 | 437.3454061 | 12049.43708 | 13.15330922 | 10.1512286 | 17.28251085 | C13309 | Demethylphylloquinone | Mass, RT and Spectra | 19574.24335 | 2667.295202 | 5025.736664 | 16489.28569 | 18329.86069 | 25298.03127 |
| 490 | C16939 | 473.3614487 | 85648.60309 | 942.2301365 | 939.3582092 | 951.6902434 | C16939 | Maslinic acid | Mass, RT and Spectra | 108.1129043 | 85884.40334 | 67427.57225 | 16051.0455 | 24837.90603 | 15376.00089 |
| 491 | C02515 | 307.9796034 | 121.5197881 | 156.8003681 | 153.3838787 | 164.3530558 | C02515 | Iodotyrosine | Mass, RT and Spectra | 161.627662 | 92.49540508 | 98.84682265 | 140.1968725 | 0 | 101.3080844 |
| 492 | C02538 | 348.0796367 | 2816.765937 | 430.1095758 | 428.8808454 | 431.4230723 | C02538 | Estrone sulfate | Mass, RT and Spectra | 298.2702804 | 4045.211488 | 4712.043812 | 434.0714638 | 1302.386286 | 1031.059947 |
| 493 | C02571 | 202.1078382 | 27538.7018 | 295.6711453 | 289.9438063 | 304.4902174 | C02571 | L-Acetylcarnitine | Mass, RT and Spectra | 1353.374033 | 46333.46475 | 39427.2928 | 4480.537047 | 13266.63884 | 13824.77907 |
| 494 | C02592 | 482.2908378 | 19251.79132 | 88.50165873 | 85.35061335 | 96.10817229 | C02592 | Lithocholyltaurine | Mass, RT and Spectra | 4635.779643 | 38731.62578 | 32233.78435 | 6950.29811 | 18504.07354 | 23876.54891 |
| 495 | C13358 | 487.2657604 | 120018.5656 | 112.265472 | 98.32075403 | 129.915626 | C13358 | Hydrocortisone butyrate propionate | Mass, RT and Spectra | 22287.73224 | 241260.6878 | 191786.9035 | 32894.55263 | 33812.79035 | 53268.63405 |
| 496 | C13713 | 334.2504015 | 3255016.943 | 916.476237 | 913.5024113 | 918.5148919 | C13713 | Tetrahydrodeoxycorticosterone | Mass, RT and Spectra | 2199.985808 | 4207.113285 | 15367.38292 | 1116.246446 | 1833.170698 | 2046.238149 |
| 497 | C02631 | 159.066157 | 940741.7651 | 1553.439728 | 1552.016444 | 1555.082797 | C02631 | Isopropylmaleate | Mass, RT and Spectra | 713513.138 | 1050129.78 | 1096226.157 | 841394.2042 | 970257.8577 | 931118.8861 |
| 498 | C13732 | 312.1447905 | 11064.44631 | 100.4390285 | 97.70661084 | 102.3926485 | C13732 | Domoic acid | Mass, RT and Spectra | 16849.50528 | 3043.714558 | 4982.446972 | 20780.14642 | 9546.716475 | 11895.7847 |
| 499 | C02632 | 89.0597967 | 202045.8767 | 19.19887532 | 3.775316242 | 23.52114189 | C02632 | Isobutyric acid | Mass, RT and Spectra | 188168.4728 | 231867.5112 | 224895.8814 | 179018.2157 | 194445.858 | 179780.7395 |
| 500 | C13790 | 243.1964215 | 30314.2462 | 1177.737 | 1172.452768 | 1181.115 | C13790 | alpha-hydroxy myristic acid | Mass, RT and Spectra | 1370.944838 | 64099.62696 | 62495.18784 | 6725.931121 | 13262.6507 | 11993.44983 |
| 501 | C02647 | 130.097356 | 398294.741 | 1654.244884 | 1647.695283 | 1665.646974 | C02647 | 4-Guanidinobutanal | Mass, RT and Spectra | 337029.0034 | 492954.4893 | 495880.5981 | 333383.0987 | 349233.5755 | 339227.3528 |
| 502 | C02656 | 205.0712854 | 24160.15683 | 353.2011205 | 343.6585414 | 358.5985841 | C02656 | Pimelic acid | Mass, RT and Spectra | 116.8115562 | 53631.71719 | 47562.10034 | 5400.929053 | 12934.78025 | 10852.75084 |
| 503 | C02678 | 231.1586339 | 20294.80764 | 12.58905496 | 7.604087311 | 17.15945486 | C02678 | Dodecanedioic acid | Mass, RT and Spectra | 18992.41602 | 23473.76746 | 21553.097 | 11752.11675 | 19375.26243 | 22378.26943 |
| 504 | C02679 | 199.1697745 | 620292.751 | 1178.8125 | 1172.452768 | 1180.8465 | C02679 | Dodecanoic acid | Mass, RT and Spectra | 73309.3492 | 1237461.498 | 1086523.812 | 221996.9999 | 437502.9701 | 322424.4463 |
| 505 | C02700 | 235.0715421 | 11839.02248 | 223.0128277 | 216.3107377 | 233.2645773 | C02700 | L-Formylkynurenine | Mass, RT and Spectra | 1016.605003 | 15909.55529 | 35727.31218 | 3373.340289 | 270.8896672 | 440.9185259 |
| 506 | C02713 | 292.102871 | 19241.22545 | 303.5885387 | 297.5231057 | 311.8694827 | C02713 | N-Acetylmuramic acid | Mass, RT and Spectra | 380.5522539 | 35090.51295 | 24936.99263 | 270.8318507 | 1969.811622 | 2624.437552 |
| 507 | C13792 | 285.2414652 | 49588.58793 | 954.6931919 | 951.5482799 | 957.8798836 | C13792 | 2-methoxy-5Z-hexadecenoic acid | Mass, RT and Spectra | 49996.76648 | 59631.59438 | 51065.86017 | 29151.63209 | 40544.95349 | 46156.66644 |
| 508 | HMDB0000701 | 218.1026636 | 298275.4882 | 197.805747 | 179.9947928 | 210.4958499 | HMDB0000701 | Hexanoylglycine | Mass, RT and Spectra | 97049.46265 | 493066.2248 | 663608.3957 | 43179.87717 | 99028.67926 | 91091.37742 |
| 509 | C13822 | 651.6324034 | 11248.63684 | 101.7686451 | 99.8127897 | 103.7252681 | C13822 | 2S,3R-didecanoyl-docosane-2,3-diol | Mass, RT and Spectra | 15218.00686 | 9843.056369 | 8689.759955 | 11067.93808 | 11087.09121 | 8016.045829 |
| 510 | C02714 | 131.1175957 | 153425.5841 | 211.9454571 | 207.934891 | 217.8425948 | C02714 | N-Acetylputrescine | Mass, RT and Spectra | 215825.1363 | 103896.34 | 95609.03595 | 221869.769 | 314663.1718 | 290694.8786 |
| 511 | C13829 | 376.317431 | 10063.32766 | 1040.846241 | 1037.011152 | 1045.359261 | C13829 | Adrenoyl ethanolamide | Mass, RT and Spectra | 20700.33526 | 1420.503694 | 3188.732472 | 11882.97207 | 6655.535073 | 3601.704114 |
| 512 | C02718 | 103.0501065 | 815654.2317 | 1776.511939 | 1762.682468 | 1785.355535 | C02718 | N-Formiminoglycine | Mass, RT and Spectra | 486333.4338 | 1172286.576 | 1180309.38 | 573409.4829 | 667295.023 | 583037.9482 |
| 513 | C13831 | 200.2003566 | 65471.16547 | 877.6800516 | 873.4539491 | 879.2826886 | C13831 | dodecanamide | Mass, RT and Spectra | 46655.56581 | 82963.05318 | 84227.17479 | 50723.35893 | 56239.1246 | 50133.9532 |
| 514 | C02737 | 708.4800574 | 10618.49162 | 1115.748853 | 1112.944196 | 1118.578099 | C02737 | PS(16:0/16:0) | Mass, RT and Spectra | 6533.110348 | 2906.164287 | 2533.232951 | 3909.205444 | 5221.033717 | 4411.837502 |
| 515 | C02739 | 720.0172017 | 41736.80985 | 87.4405901 | 83.63669158 | 94.52903484 | C02739 | Phosphoribosyl-ATP | Mass, RT and Spectra | 78763.51244 | 8007.639245 | 7230.240986 | 56350.45579 | 57154.50207 | 55069.61233 |
| 516 | C13846 | 284.2941758 | 291940.1542 | 13.3286132 | 6.929908215 | 19.01051875 | C13846 | Octadecanamide | Mass, RT and Spectra | 38514.30878 | 623556.8294 | 561863.159 | 96078.10448 | 168604.7567 | 143380.0862 |
| 517 | C13856 | 377.2691574 | 18243.39072 | 97.94824094 | 95.44757726 | 102.1048245 | C13856 | 2-Arachidonylglycerol | Mass, RT and Spectra | 1361.409837 | 37489.132 | 42241.7051 | 1176.52058 | 1049.091104 | 4827.497965 |
| 518 | C02771 | 247.1335279 | 29871.12639 | 94.11234076 | 92.09942682 | 100.6379712 | C02771 | 1,2-Dihydrosantonin | Mass, RT and Spectra | 2994.820532 | 59403.6602 | 52846.50235 | 11912.78317 | 20130.09138 | 30376.70751 |
| 519 | C13864 | 551.5021751 | 110719.3601 | 17.38646795 | 12.88328233 | 30.5178217 | C13864 | DG(P-14:0/18:1(9Z)) | Mass, RT and Spectra | 81.94712582 | 229370.8696 | 218782.9936 | 32876.5595 | 51520.56235 | 44263.14022 |
| 520 | C02774 | 187.1334209 | 192316.7778 | 97.18174862 | 94.1372194 | 99.81746849 | C02774 | 10-hydroxy capric acid | Mass, RT and Spectra | 28331.84942 | 304276.8759 | 399769.5221 | 51588.63641 | 76006.91563 | 70910.91609 |
| 521 | C02794 | 225.0876334 | 217549.8527 | 1553.14678 | 1550.509016 | 1555.591498 | C02794 | Hydroxykynurenine | Mass, RT and Spectra | 175435.4865 | 275538.9325 | 264463.1461 | 174800.141 | 167688.2935 | 176340.0991 |
| 522 | C02835 | 127.0499313 | 327403.0351 | 1552.958412 | 1547.185519 | 1557.057818 | C02835 | Imidazoleacetic acid | Mass, RT and Spectra | 460433.8042 | 235520.4477 | 230263.9571 | 365842.9632 | 324188.0565 | 350628.7828 |
| 523 | C02838 | 322.1779741 | 14286.90628 | 147.7679994 | 143.8606638 | 155.9244342 | C02838 | L-Octanoylcarnitine | Mass, RT and Spectra | 30659.23746 | 150.5578349 | 471.5277398 | 23.60741734 | 0 | 1 |
| 524 | C02846 | 122.0963871 | 4990595.849 | 573.4101132 | 541.2825705 | 598.0514665 | C02846 | N,N-Dimethylaniline | Mass, RT and Spectra | 4873094.634 | 4986023.613 | 4995042.961 | 4832893.197 | 5108421.372 | 4929262.881 |
| 525 | C02862 | 232.154021 | 1425084.378 | 734.2693429 | 716.9522257 | 756.480813 | C02862 | Butyrylcarnitine | Mass, RT and Spectra | 5254.87548 | 8093.710685 | 16950.71419 | 3241.315655 | 120.4171002 | 14399.98314 |
| 526 | HMDB0000705 | 304.1780432 | 20283.42942 | 158.9744562 | 153.4423476 | 180.1663881 | HMDB0000705 | Hexanoylcarnitine | Mass, RT and Spectra | 7098.875413 | 31715.64605 | 42604.54171 | 6008.383056 | 8408.803516 | 18403.69542 |
| 527 | C02915 | 353.1599997 | 745758.6654 | 104.2467114 | 98.68879567 | 111.2454261 | C02915 | (S)-N-methylcanadine | Mass, RT and Spectra | 893114.1075 | 813411.4811 | 589317.0217 | 465358.9055 | 138866.3187 | 143365.8526 |
| 528 | C02946 | 144.0660423 | 70355.20137 | 221.101356 | 193.8985824 | 235.4853458 | C02946 | 4-Acetamidobutanoic acid | Mass, RT and Spectra | 9168.833222 | 113628.6549 | 109681.458 | 20533.31735 | 107466.5561 | 132541.4578 |
| 529 | C13914 | 328.3201819 | 522723.8405 | 14.68725797 | 2.258199084 | 45.5395592 | C13914 | N,N-Dimethylsphingosine | Mass, RT and Spectra | 76137.94012 | 1055674.44 | 847213.2557 | 195836.7783 | 375207.1417 | 273866.0688 |
| 530 | C02979 | 171.0072899 | 9397.803493 | 256.5478559 | 251.2605058 | 264.141329 | C02979 | Beta-Glycerophosphoric acid | Mass, RT and Spectra | 16258.73065 | 259889.0583 | 276003.3809 | 18486.16971 | 42217.26596 | 54223.69467 |
| 531 | C17076 | 213.1854599 | 146324.0042 | 1178.631857 | 1172.452768 | 1181.001724 | C17076 | Tridecanoic acid | Mass, RT and Spectra | 3400564.711 | 4385688.211 | 5554918.271 | 4095443.37 | 4171761.442 | 4113635.666 |
| 532 | C13915 | 274.2729898 | 257472.6827 | 1019.968808 | 1008.44449 | 1028.551141 | C13915 | C16 Sphinganine | Mass, RT and Spectra | 5492.694755 | 631462.3938 | 643958.4089 | 4367.753272 | 82011.0569 | 7271.437534 |
| 533 | C02990 | 400.3405818 | 251377.0794 | 1077.625391 | 1071.342636 | 1082.749744 | C02990 | L-Palmitoylcarnitine | Mass, RT and Spectra | 19155.90733 | 4961.946467 | 6679.252303 | 25029.44472 | 9570.435987 | 13040.8789 |
| 534 | C02997 | 196.074037 | 232556.2686 | 394.9844696 | 374.7553664 | 400.9055458 | C02997 | N-Acetylhistidine | Mass, RT and Spectra | 33565.15227 | 466622.1637 | 419316.898 | 58136.6924 | 111833.9517 | 83011.53946 |
| 535 | C14040 | 172.0395722 | 3384986.744 | 143.0705235 | 135.3914289 | 152.0765751 | C14040 | 1-Nitronaphthalene | Mass, RT and Spectra | 45740.21438 | 43543.74586 | 30238.26759 | 50753.29525 | 67465.52784 | 80679.34099 |
| 536 | C03017 | 218.1384052 | 4723840.491 | 545.0599621 | 534.6374119 | 548.9914258 | C03017 | Propionylcarnitine | Mass, RT and Spectra | 4078791.067 | 3977635.303 | 3824299.728 | 4395825.073 | 9024838.856 | 8129254.153 |
| 537 | C03033 | 361.110028 | 119661.2611 | 416.9130466 | 412.0465501 | 428.4993879 | C03033 | 3-Methoxy-4-hydroxyphenylglycol glucuronide | Mass, RT and Spectra | 26583174.39 | 61016604.17 | 61075913.29 | 34993325.79 | 35313731.85 | 23760890.94 |
| 538 | C03042 | 299.2583292 | 178059.6801 | 92.6410409 | 90.22315074 | 94.3959 | C03042 | 2R-hydroxy-stearic acid | Mass, RT and Spectra | 36666.14339 | 280990.6415 | 343304.9849 | 55648.605 | 76907.42887 | 72089.2836 |
| 539 | C03044 | 89.06047013 | 7605.981793 | 141.6559463 | 135.9554909 | 146.8141223 | C03044 | (2R,3R)-2,3-Butanediol | Mass, RT and Spectra | 84.20020518 | 11407.24831 | 20674.66074 | 100.4979704 | 556.1964208 | 1996.433537 |
| 540 | C14112 | 187.0972404 | 1256824.405 | 1553.464205 | 1552.431846 | 1554.951615 | C14112 | cis-2-Carboxycyclohexyl-acetic acid | Mass, RT and Spectra | 1005725.51 | 1460819.492 | 1420802.923 | 1102942.253 | 1156310.662 | 1051602.136 |
| 541 | C03087 | 160.0964122 | 117375.5476 | 318.3432702 | 308.4197068 | 324.5570547 | C03087 | 5-Acetamidovalerate | Mass, RT and Spectra | 139236.7744 | 109226.8304 | 91159.4045 | 84018.16182 | 205488.4358 | 75393.63625 |
| 542 | C03089 | 181.0523071 | 48316.72252 | 141.4422178 | 138.1479473 | 146.6822357 | C03089 | 5-Methylthioribose | Mass, RT | 52684.50397 | 31078.08083 | 12249.28286 | 77688.55387 | 139827.9455 | 100847.2037 |
| 543 | C03090 | 230.0429553 | 21957.31054 | 232.908256 | 225.6001057 | 241.783338 | C03090 | 5-Phosphoribosylamine | Mass, RT and Spectra | 375.7084141 | 51478.95777 | 47942.09235 | 912.2315701 | 4651.160372 | 3636.829923 |
| 544 | C14179 | 138.9701429 | 46215.03308 | 534.618587 | 529.4763828 | 540.1499591 | C14179 | Sulfoacetate | Mass, RT and Spectra | 3766.314241 | 87982.71128 | 104542.837 | 10695.02812 | 10719.16712 | 19082.19031 |
| 545 | C14218 | 145.1228501 | 10527.56947 | 1181.468301 | 1172.452768 | 1187.21558 | C14218 | octane-1,8-diol | Mass, RT and Spectra | 166.527853 | 21922.07315 | 22224.24668 | 1156.648502 | 1834.960642 | 4672.879132 |
| 546 | C03139 | 176.068067 | 3518.361431 | 105.1296521 | 103.0411073 | 106.6459986 | C03139 | Guanidinosuccinic acid | Mass, RT and Spectra | 873.2566375 | 4473.497047 | 5343.478292 | 1105.810013 | 1896.19229 | 11747.5167 |
| 547 | C14240 | 371.3151532 | 86595.03855 | 1164.963386 | 1138.830056 | 1177.655642 | C14240 | Diethylhexyl adipate | Mass, RT and Spectra | 83620.90822 | 96226.32345 | 93272.50761 | 80636.10906 | 75259.43349 | 77739.29618 |
| 548 | C03145 | 176.0380343 | 72127.45468 | 149.0218662 | 133.6863784 | 176.2677535 | C03145 | N-Formyl-L-methionine | Mass, RT and Spectra | 10921.19479 | 132553.0261 | 167336.3712 | 9196.495128 | 31664.48313 | 25447.42075 |
| 549 | C03187 | 549.0886471 | 75643.06348 | 577.6718908 | 566.5460667 | 585.4071366 | C03187 | dTDP-6-deoxy-beta-L-talose | Mass, RT and Spectra | 128302.1037 | 32082.63066 | 30570.42514 | 63820.5646 | 128150.0809 | 84652.03894 |
| 550 | C03197 | 103.039722 | 2875289.69 | 235.5196851 | 212.7897987 | 251.1631408 | C03197 | (S)-3-Hydroxybutyric acid | Mass, RT and Spectra | 392732.5603 | 4680892.463 | 4433512.406 | 500977.5448 | 1041552.853 | 1098955.695 |
| 551 | C14422 | 103.0865163 | 474385.4106 | 75.64139046 | 70.52443573 | 79.41104394 | C14422 | Diethylamine | Mass, RT and Spectra | 228881.5432 | 755757.9046 | 819579.7544 | 266657.4896 | 291342.4489 | 316528.858 |
| 552 | C14472 | 343.1170849 | 11347.25654 | 352.8323511 | 345.1223206 | 374.8260232 | C14472 | 4',5,6,7-Tetramethoxyflavone | Mass, RT and Spectra | 19252.67643 | 3706.700873 | 3675.652088 | 16229.97521 | 16022.63396 | 14910.03849 |
| 553 | C03223 | 199.024678 | 5591.585023 | 96.3360455 | 90.31872822 | 109.8734047 | C03223 | 3,4-Dihydroxyphthalate | Mass, RT and Spectra | 6916.543613 | 5209.824888 | 5447.891297 | 5654.02076 | 5063.010573 | 4590.703424 |
| 554 | HMDB0000734 | 188.0701797 | 5109105.075 | 576.3031805 | 565.9101884 | 579.7665496 | HMDB0000734 | Indoleacrylic acid | Mass, RT and Spectra | 3759261.939 | 7205749.895 | 7898846.357 | 2925555.752 | 3366434.398 | 3756468.728 |
| 555 | C14519 | 219.0821265 | 273244.9656 | 151.8591399 | 133.132771 | 165.7523179 | C14519 | 1-Hydroxypyrene | Mass, RT and Spectra | 421657.9942 | 204223.2846 | 205448.8203 | 250778.9536 | 268049.6045 | 244340.7623 |
| 556 | C03226 | 781.6112535 | 160468.8737 | 1080.6074 | 1074.098466 | 1084.729599 | C03226 | 3-demethylubiquinone-9 | Mass, RT and Spectra | 180776.5575 | 130956.4877 | 123743.6617 | 193311.7515 | 167537.9485 | 174510.745 |
| 557 | C03232 | 184.9851772 | 9136053.542 | 1735.682228 | 1715.856737 | 1736.898653 | C03232 | Phosphohydroxypyruvic acid | Mass, RT and Spectra | 239209.9191 | 244176.9904 | 162711.2895 | 224697.8211 | 155122.9586 | 181457.0121 |
| 558 | C03242 | 305.2472798 | 333732.2481 | 91.05278502 | 89.11413934 | 92.08351096 | C03242 | 8,11,14-Eicosatrienoic acid | Mass, RT and Spectra | 63569.49187 | 539376.9982 | 569996.7071 | 121209.8472 | 95013.55013 | 134855.5563 |
| 559 | C03277 | 153.0300401 | 4051.530906 | 332.4454258 | 329.4589632 | 335.6922675 | C03277 | Imidazol-5-yl-pyruvate | Mass, RT and Spectra | 547.3794365 | 7257.272848 | 7928.930287 | 451.4150833 | 2623.578718 | 2087.830112 |
| 560 | C03283 | 119.0812227 | 343033.9432 | 1661.825321 | 1660.00361 | 1665.753834 | C03283 | L-2,4-diaminobutyric acid | Mass, RT and Spectra | 224174.0915 | 497113.7844 | 501223.0705 | 237562.8414 | 269200.3223 | 241031.8244 |
| 561 | HMDB0000750 | 281.0298334 | 61416.74147 | 235.3109129 | 230.5473316 | 244.6320066 | HMDB0000750 | 3-Hydroxymandelic acid | Mass, RT and Spectra | 4572.637334 | 132720.0814 | 166429.2717 | 5537.672964 | 21056.95069 | 28899.71709 |
| 562 | C14561 | 313.1957017 | 431143.0505 | 117.7778627 | 99.34856338 | 140.1794974 | C14561 | (+/-)-2,4,6-Triphenyl-1-hexene | Mass, RT and Spectra | 1213187.438 | 51371.06488 | 314880.7473 | 716641.5549 | 556.3703308 | 18623.34998 |
| 563 | C03287 | 226.0113563 | 3410251.179 | 1736.212703 | 1730.663322 | 1738.264379 | C03287 | L-Glutamic acid 5-phosphate | Mass, RT and Spectra | 295364.9573 | 200804.3144 | 213317.9451 | 308055.49 | 298595.9073 | 291418.221 |
| 564 | C03365 | 193.0711066 | 27480.8085 | 418.8268741 | 414.951646 | 430.9080516 | C03365 | Sequoyitol | Mass, RT and Spectra | 658.1077084 | 51735.05426 | 48402.29235 | 1794.797105 | 9401.544044 | 26520.01645 |
| 565 | C03366 | 243.0736521 | 558.4484299 | 748.9361334 | 748.0274616 | 750.3983219 | C03366 | 5-phosphonooxy-L-lysine | Mass, RT and Spectra | 539.0089116 | 1416.526594 | 981.5598217 | 0 | 100.8250795 | 36.72933276 |
| 566 | C03401 | 105.065869 | 116668.5375 | 117.2392181 | 115.2230439 | 118.6835951 | C03401 | 2,3-Diaminopropionic acid | Mass, RT and Spectra | 1505918.352 | 940194.1817 | 982552.0325 | 2639794.677 | 2156749.068 | 2329170.951 |
| 567 | HMDB0000766 | 131.0538871 | 83330.69749 | 214.6258003 | 205.9894769 | 220.5364106 | HMDB0000766 | N-Acetyl-L-alanine | Mass, RT and Spectra | 10561.35561 | 165725.9279 | 150042.8456 | 10451.30245 | 32223.54204 | 38812.25392 |
| 568 | C03406 | 271.1044783 | 5462.767337 | 514.2986202 | 513.0227573 | 516.2143922 | C03406 | Argininosuccinic acid | Mass, RT and Spectra | 39.79262634 | 11969.42433 | 13248.23066 | 3147.18414 | 64.49617396 | 1003.98282 |
| 569 | C03410 | 324.092661 | 3952.980699 | 440.8924991 | 440.1240944 | 442.9153386 | C03410 | N-Glycolylneuraminic acid | Mass, RT and Spectra | 4173.83831 | 83417.15045 | 93327.28869 | 16453.05152 | 59922.46326 | 70050.19587 |
| 570 | C03413 | 287.2436243 | 8448.356848 | 1651.384965 | 1634.982952 | 1661.202514 | C03413 | N1,N12-Diacetylspermine | Mass, RT and Spectra | 524.9229814 | 32280.13373 | 27045.16809 | 5435.252816 | 10485.87587 | 6918.536065 |
| 571 | C03428 | 585.3098388 | 3457399.865 | 723.6804647 | 702.7732438 | 728.499206 | C03428 | Presqualene diphosphate | Mass, RT and Spectra | 81745.66997 | 29996.29432 | 30683.57997 | 74737.09683 | 73653.57756 | 74224.74333 |
| 572 | HMDB0000888 | 217.1446222 | 8807.860882 | 592.9086157 | 592.1729426 | 595.5715969 | HMDB0000888 | Undecanedioic acid | Mass, RT and Spectra | 14504.92527 | 3441.102981 | 2476.331883 | 10620.77971 | 11593.84259 | 10024.09475 |
| 573 | C03440 | 132.0651545 | 1310132.582 | 101.3702735 | 97.22497695 | 105.6821029 | C03440 | cis-4-Hydroxy-D-proline | Mass, RT and Spectra | 1420602.18 | 1267563.7 | 814075.6993 | 1779441.516 | 1728043.54 | 1570028.338 |
| 574 | C14606 | 307.2261677 | 14640.07171 | 10.13359164 | 6.164236861 | 13.28720642 | C14606 | 11-Hydroxyandrosterone | Mass, RT and Spectra | 1168.040786 | 34068.33959 | 58480.14684 | 1944.155351 | 6738.624855 | 8962.242279 |
| 575 | C14771 | 159.1095824 | 3472793.159 | 172.705295 | 155.5449003 | 189.9282468 | C14771 | 14,15-Epoxy-5,8,11-eicosatrienoic acid | Mass, RT | 130661.5279 | 154877.9782 | 164288.8156 | 139782.8106 | 147708.3806 | 144529.2686 |
| 576 | C03451 | 378.0947283 | 23778.44185 | 384.8632778 | 377.9795538 | 392.3173071 | C03451 | S-Lactoylglutathione | Mass, RT and Spectra | 6479.618056 | 64403.33389 | 44108.42996 | 1857.860509 | 2294.98148 | 2230.149689 |
| 577 | C14772 | 168.1146875 | 23982.42863 | 100.4878565 | 95.73758048 | 102.5866454 | C14772 | 5,6-DHET | Mass, RT | 1217.202141 | 45594.19649 | 52078.3385 | 3058.232777 | 4724.440695 | 7144.315459 |
| 578 | C14791 | 224.5629279 | 75717.84325 | 98.52955243 | 94.29482503 | 101.8977652 | C14791 | (1R)-Hydroxy-(2R)-glutathionyl-1,2-dihydronaphthalene | Mass, RT and Spectra | 8465.420903 | 135097.3136 | 130171.4004 | 18228.42403 | 36840.7794 | 35604.85394 |
| 579 | C03458 | 126.0192176 | 64529.01374 | 281.6848716 | 262.8250913 | 300.3283657 | C03458 | 2,3,6-Trihydroxypyridine | Mass, RT and Spectra | 14863.3338 | 109076.5606 | 129072.0558 | 22112.33403 | 50616.99427 | 38964.82529 |
| 580 | C03470 | 451.3769427 | 189345.6855 | 1080.416439 | 1073.714417 | 1089.693245 | C03470 | 3-alpha(S)-Strictosidine | Mass, RT and Spectra | 184646.7245 | 184531.0647 | 183811.3739 | 145315.0214 | 176093.8453 | 166407.9758 |
| 581 | C03519 | 206.0814953 | 40741.74045 | 112.2775999 | 100.4129992 | 114.5353812 | C03519 | N-Acetyl-L-phenylalanine | Mass, RT and Spectra | 3871.838469 | 83743.103 | 68585.77091 | 17335.25469 | 20804.86735 | 23377.34156 |
| 582 | C03557 | 78.95874419 | 501624.0724 | 496.7519302 | 494.3327889 | 507.2249883 | C03557 | Ciliatine | Mass, RT and Spectra | 127515.3032 | 845259.23 | 1060280.502 | 188728.0099 | 284391.452 | 273716.221 |
| 583 | C03564 | 112.0400159 | 48073.69715 | 526.1869797 | 521.197199 | 535.85295 | C03564 | 1-Pyrroline-2-carboxylic acid | Mass, RT and Spectra | 175.8684686 | 108498.9498 | 103477.7434 | 10774.0971 | 12155.18111 | 14690.21509 |
| 584 | C14800 | 188.034679 | 3629721.597 | 179.8955734 | 170.1923121 | 195.9086588 | C14800 | 1-Nitronaphthalene-5,6-oxide | Mass, RT and Spectra | 26579.58183 | 12656.34587 | 8774.581314 | 30744.86456 | 32269.16788 | 25929.2697 |
| 585 | HMDB0001624 | 131.0708849 | 359785.7271 | 124.3901947 | 104.5614597 | 140.6173186 | HMDB0001624 | 2-Hydroxycaproic acid | Mass, RT and Spectra | 74028.16859 | 650318.3928 | 719713.0046 | 90117.75879 | 143441.3556 | 94332.32912 |
| 586 | C03577 | 456.2437333 | 104559.4855 | 519.3646502 | 513.8951737 | 538.8257189 | C03577 | 20-Hydroxy-leukotriene E4 | Mass, RT and Spectra | 140718.7497 | 71535.40198 | 70932.72485 | 126282.208 | 119211.9131 | 124312.6974 |
| 587 | C14820 | 337.234192 | 22895.95515 | 811.8165778 | 806.4230066 | 829.3570381 | C14820 | 11(R)-HPETE | Mass, RT and Spectra | 38341.79841 | 291828.9812 | 304503.9572 | 92169.62418 | 76278.41705 | 56876.86845 |
| 588 | C14829 | 313.2378287 | 180713.4407 | 98.5648698 | 93.99765235 | 103.7673287 | C14829 | 12,13-DHOME | Mass, RT and Spectra | 19697.49835 | 5415.638537 | 4827.449131 | 19404.76537 | 17313.20962 | 18653.32705 |
| 589 | C03592 | 242.1144778 | 225812.9971 | 1554.429943 | 1552.064652 | 1557.107314 | C03592 | 5-Methyldeoxycytidine | Mass, RT and Spectra | 223304.6179 | 234099.6589 | 234487.0076 | 213562.7786 | 244563.8915 | 220626.8144 |
| 590 | C14847 | 480.0420985 | 6791.97757 | 104.3042918 | 100.5014462 | 106.9627138 | C14847 | 3,4-Dihydro-3-hydroxy-4-S-glutathionyl bromobenzene | Mass, RT and Spectra | 2133.255205 | 335.4835818 | 608.0960291 | 4948.327597 | 1274.04121 | 23499.66262 |
| 591 | C14853 | 303.1042086 | 45336.48785 | 352.0802348 | 346.0197352 | 359.6470122 | C14853 | Benzo[a]pyrene-7,8-dihydrodiol-9,10-oxide | Mass, RT and Spectra | 57961.6122 | 29929.35211 | 29592.08228 | 44503.70046 | 85578.58535 | 122337.3504 |
| 592 | C14854 | 285.0936191 | 3394.346307 | 828.1337366 | 826.5580381 | 831.9905299 | C14854 | 9-Hydroxybenzo[a]pyrene-4,5-oxide | Mass, RT and Spectra | 5702.43992 | 1362.922795 | 1297.827181 | 4649.288006 | 5334.943782 | 5094.55178 |
| 593 | HMDB0001857 | 196.0575564 | 7208.762526 | 178.9060625 | 170.7694359 | 179.6258019 | HMDB0001857 | 1,3-Dimethyluric acid | Mass, RT and Spectra | 5989.347405 | 1511.819031 | #DIV/0! | 2655.121522 | 158.971348 | 53.91165203 |
| 594 | C14860 | 130.9648274 | 352747.2582 | 1732.331956 | 1716.760067 | 1736.837183 | C14860 | 2,2-Dichloro-1,1-ethanediol | Mass, RT and Spectra | 104164.2954 | 549984.5617 | 533327.1317 | 186731.3883 | 264179.9214 | 186827.533 |
| 595 | C03594 | 402.3482176 | 3676025.427 | 1228.176994 | 1206.896682 | 1235.444399 | C03594 | 7a-Hydroxycholesterol | Mass, RT and Spectra | 32498.90518 | 29890.47861 | 47139.84304 | 23418.81618 | 28357.1796 | 24215.66548 |
| 596 | C14872 | 148.9909123 | 13496.57309 | 486.1668078 | 484.984441 | 487.4541519 | C14872 | Thiodiacetic acid | Mass, RT and Spectra | 403.8929102 | 30688.04884 | 26900.95371 | 6671.843174 | 8689.088557 | 11199.29411 |
| 597 | C03626 | 201.134937 | 9520.582743 | 477.2055837 | 474.858789 | 478.9083611 | C03626 | Asymmetric dimethylarginine | Mass, RT and Spectra | 812708.4258 | 11558980.63 | 12342753.19 | 1030005.006 | 1830515.1 | 1678137.928 |
| 598 | C03640 | 466.3570631 | 36305.16136 | 891.3903813 | 888.2900985 | 897.8702254 | C03640 | LysoSM(d18:1) | Mass, RT and Spectra | 34554.29016 | 29714.29174 | 39627.52162 | 27902.59624 | 19759.27263 | 31468.76129 |
| 599 | C03646 | 499.1210821 | 972.2124786 | 104.5486937 | 104.1590682 | 104.9383192 | C03646 | Bis-gamma-L-glutamyl-L-cystine | Mass, RT and Spectra | 1570.485893 | 734.8949288 | 843.1753308 | 920.2785954 | 787.3461486 | 293.892954 |
| 600 | C14877 | 341.1057174 | 19797.44308 | 112.4543411 | 108.5999848 | 116.6230171 | C14877 | S-[2-(N7-Guanyl)ethyl]-N-acetyl-L-cysteine | Mass, RT and Spectra | 25017.00055 | 9740.892934 | 7006.386798 | 40365.40895 | 37786.91689 | 37173.30768 |
| 601 | C03664 | 220.9751971 | 12466.24902 | 594.9678845 | 593.6424172 | 600.8357159 | C03664 | 2,4-Dichlorophenoxyacetic acid | Mass, RT and Spectra | 19915.34868 | 5944.375996 | 4381.758661 | 15752.93902 | 16235.11976 | 14680.4249 |
| 602 | C15532 | 216.0981536 | 40110.38941 | 351.6680375 | 341.681202 | 363.6767829 | C15532 | N-a-Acetylcitrulline | Mass, RT and Spectra | 16258.02016 | 293551.1532 | 261403.5372 | 47121.3954 | 95152.9936 | 72679.92069 |
| 603 | C15556 | 185.0224539 | 837644.7081 | 236.1335816 | 221.5439683 | 252.855674 | C15556 | 3,4-Dihydroxy-2-butanone 4-phosphate | Mass, RT and Spectra | 964138.4259 | 736735.4752 | 724368.8529 | 938677.0634 | 814586.5139 | 840403.4176 |
| 604 | C15557 | 434.3267182 | 43860.49539 | 841.160586 | 840.0893817 | 842.8580047 | C15557 | Lithocholic acid glycine conjugate | Mass, RT and Spectra | 33830.63662 | 45995.96546 | 90471.75776 | 55969.48112 | 9507.425837 | 34355.48437 |
| 605 | C03665 | 102.055681 | 86686.09723 | 377.8266048 | 371.3978095 | 385.1175962 | C03665 | 2-Aminoisobutyric acid | Mass, RT and Spectra | 2116.924931 | 186895.0879 | 163695.7375 | 24933.8265 | 38849.74863 | 59745.17606 |
| 606 | C15559 | 530.2770394 | 463225.7277 | 611.9977648 | 607.8666168 | 616.8021116 | C15559 | N-[(3a,5b,7a)-3-hydroxy-24-oxo-7-(sulfooxy)cholan-24-yl]-Glycine | Mass, RT and Spectra | 251328.0351 | 636717.1116 | 635207.567 | 458403.8473 | 378918.7896 | 351478.581 |
| 607 | C03690 | 389.2684228 | 72533.55498 | 96.18271559 | 93.99765235 | 102.5351152 | C03690 | DEHP | Mass, RT and Spectra | 11636.07249 | 148478.6193 | 144129.7151 | 9399.559627 | 11977.64601 | 28393.38217 |
| 608 | C03691 | 631.1545674 | 108385.9107 | 101.940884 | 99.57822503 | 104.4103409 | C03691 | CMP-N-glycoloylneuraminate | Mass, RT and Spectra | 119045.8674 | 95888.41337 | 99669.86868 | 117981.4164 | 98956.6099 | 110668.1946 |
| 609 | C03722 | 122.0244349 | 24174.93959 | 469.9665191 | 468.9681063 | 471.4222246 | C03722 | Quinolinic acid | Mass, RT and Spectra | 6223.749436 | 35783.44602 | 32953.31199 | 11044.5579 | 34481.11793 | 31319.06044 |
| 610 | C03740 | 217.082327 | 11607.77875 | 407.3284908 | 405.0108779 | 418.3135329 | C03740 | gamma-Glutamylalanine | Mass, RT and Spectra | 117831.6112 | 5925.610694 | 4919.629995 | 89181.15528 | 73793.79061 | 72170.98761 |
| 611 | C15606 | 161.0278456 | 5835.358118 | 478.7507811 | 478.1969368 | 497.1118105 | C15606 | 1,2-Dihydroxy-3-keto-5-methylthiopentene | Mass, RT and Spectra | 150.6640853 | 15449.47481 | 16316.71847 | 52.39817927 | 1613.606772 | 1685.945995 |
| 612 | C03758 | 154.0859301 | 86845.15111 | 13.12885818 | 5.204336144 | 26.48729974 | C03758 | Dopamine | Mass, RT and Spectra | 39769.31276 | 125232.0843 | 129553.1854 | 55825.24084 | 67723.94624 | 78460.5276 |
| 613 | HMDB0001863 | 99.0448041 | 112600.8642 | 162.4952218 | 132.2285499 | 191.9530611 | HMDB0001863 | 2-Hydroxyvaleric acid | Mass, RT and Spectra | 22318.64254 | 231059.8394 | 162981.1039 | 62271.58884 | 65208.65853 | 71732.3758 |
| 614 | C03761 | 161.0449984 | 2070155.731 | 422.3689622 | 419.4687641 | 429.9526044 | C03761 | 3-Hydroxymethylglutaric acid | Mass, RT and Spectra | 188219.3746 | 5195601.31 | 3423869.904 | 538828.0777 | 764477.1998 | 676674.2021 |
| 615 | C15700 | 215.1027838 | 7656.177339 | 202.0923345 | 197.378283 | 212.6889256 | C15700 | gamma-Glutamyl-gamma-aminobutyraldehyde | Mass, RT and Spectra | 452.7211266 | 7265.668984 | 18275.00647 | 552.454795 | 20422.54147 | 13418.4282 |
| 616 | C15799 | 435.3823335 | 10908.10229 | 1166.444813 | 1162.104045 | 1170.808604 | C15799 | 6-Deoxoteasterone | Mass, RT and Spectra | 7835.726473 | 9708.086323 | 18470.69492 | 5110.120605 | 10125.5716 | 10037.14511 |
| 617 | C03771 | 174.0867621 | 53679.4072 | 419.4835073 | 417.8864167 | 422.634178 | C03771 | 2-Oxoarginine | Mass, RT and Spectra | 126558.7147 | 28357.50137 | 11903.71009 | 73132.43458 | 34791.91554 | 4465.54757 |
| 618 | C03772 | 143.0969113 | 4129219.877 | 157.8722796 | 150.548131 | 178.4379252 | C03772 | Etiocholanedione | Mass, RT and Spectra | 15472.93389 | 57132.94871 | 55529.111 | 17812.41578 | 24486.90542 | 18716.81428 |
| 619 | C15802 | 449.3632272 | 147882.5075 | 90.81187838 | 88.93172031 | 92.7124649 | C15802 | 6-Deoxocastasterone | Mass, RT and Spectra | 37687.00677 | 265977.0797 | 270520.2096 | 34959.44074 | 47968.94396 | 65262.19356 |
| 620 | C15877 | 567.4610299 | 104980.7605 | 1055.407903 | 1046.640983 | 1073.676264 | C15877 | Anhydrorhodovibrin | Mass, RT and Spectra | 128938.3481 | 76834.45825 | 99775.50497 | 98354.85776 | 101336.2474 | 112121.8842 |
| 621 | C03793 | 189.159148 | 959542.7467 | 165.2208848 | 153.5301972 | 186.9886283 | C03793 | N6,N6,N6-Trimethyl-L-lysine | Mass, RT and Spectra | 165535.7038 | 254636.3457 | 157963.4599 | 162365.6363 | 422994.9023 | 229693.6898 |
| 622 | HMDB0002404 | 196.060216 | 11368.69551 | 386.2300853 | 384.9512613 | 402.7574008 | HMDB0002404 | Alpha-Hydroxyhippuric acid | Mass, RT and Spectra | 18055.70991 | 5216.993907 | 8917.35247 | 7747.749196 | 9522.021463 | 24992.547 |
| 623 | C03794 | 464.0838079 | 7962.189585 | 104.9633832 | 103.0570258 | 107.9631438 | C03794 | Adenylsuccinic acid | Mass, RT and Spectra | 11304.55974 | 4346.577316 | 4076.645302 | 9233.466647 | 9305.015374 | 7826.437044 |
| 624 | C03805 | 435.2506723 | 50689.29091 | 101.1329518 | 95.22699997 | 107.0577676 | C03805 | DHAP(18:0) | Mass, RT and Spectra | 6434.682824 | 103381.4668 | 115257.1288 | 4750.89992 | 6220.806465 | 6552.221369 |
| 625 | C03838 | 285.0479755 | 4449872.37 | 89.44841984 | 85.91772134 | 94.53204107 | C03838 | Glycineamideribotide | Mass, RT and Spectra | 78034.16067 | 12645.36745 | 12553.52885 | 73174.63089 | 57093.33274 | 55943.1094 |
| 626 | C03862 | 132.9459057 | 59529.83612 | 496.1887327 | 494.3327889 | 498.6761888 | C03862 | Dolichyl phosphate D-mannose | Mass, RT and Spectra | 684.1055193 | 128439.4293 | 142734.517 | 13773.36556 | 8679.295498 | 15351.41918 |
| 627 | C03872 | 227.0431032 | 20450.26762 | 546.1953402 | 538.0356062 | 556.1100725 | C03872 | L-2-Aminoethyl seryl phosphate | Mass, RT and Spectra | 839.6322503 | 46424.10204 | 41739.97418 | 3472.194003 | 6551.653921 | 7019.820331 |
| 628 | C03925 | 294.8623323 | 8962.450595 | 1767.912431 | 1760.153322 | 1771.847411 | C03925 | 3,5-Dibromo-4-hydroxybenzoate | Mass, RT and Spectra | 10290.29153 | 162.1390843 | 206.4665284 | 9170.654847 | 6895.542453 | 6291.09098 |
| 629 | C03955 | 203.1029815 | 29913.25103 | 515.3492377 | 513.058988 | 518.8652111 | C03955 | N6-Acetyl-N6-hydroxy-L-lysine | Mass, RT and Spectra | 4908.54427 | 57108.93025 | 64581.58593 | 13736.27972 | 7958.645501 | 12586.28268 |
| 630 | C04025 | 103.003354 | 170721.1579 | 491.3504656 | 488.528409 | 498.491285 | C04025 | Malonic acid | Mass, RT and Spectra | 20752.57824 | 329955.7237 | 321875.8816 | 47443.06032 | 67529.02823 | 82989.21824 |
| 631 | C15886 | 569.4761774 | 122660.122 | 1082.543863 | 1073.581402 | 1085.535791 | C15886 | Dihydroanhydrorhodovibrine/ Dihydromethoxylycopene | Mass, RT and Spectra | 176437.3009 | 75812.51444 | 88047.96498 | 115720.9809 | 122810.8323 | 124527.6189 |
| 632 | C04137 | 247.1383347 | 66351.92357 | 1032.348185 | 1031.378762 | 1034.181075 | C04137 | N2-(D-1-Carboxyethyl)-L-arginine | Mass, RT and Spectra | 26297.79504 | 73252.78965 | 161086.2693 | 55045.8422 | 68044.67095 | 38755.93399 |
| 633 | C04148 | 263.1032664 | 133371.8146 | 145.2096119 | 129.682815 | 169.3533564 | C04148 | Alpha-N-Phenylacetyl-L-glutamine | Mass, RT and Spectra | 30796.09572 | 235177.3252 | 320053.9643 | 28840.64115 | 64961.8565 | 119017.2863 |
| 634 | C04188 | 260.0127809 | 4455878.058 | 233.7407867 | 212.1993265 | 259.562236 | C04188 | 5-Methylthioribose 1-phosphate | Mass, RT and Spectra | 43464.97651 | 151556.2714 | 136858.2864 | 61555.06344 | 85544.08171 | 76615.02727 |
| 635 | HMDB00044 | 175.0241437 | 74158.08361 | 104.2891662 | 97.26549129 | 106.7830408 | HMDB00044 | Ascorbic acid | Mass, RT and Spectra | 17697.7001 | 113828.3457 | 95088.45415 | 53611.5555 | 80936.48198 | 103552.7765 |
| 636 | C04218 | 817.5653649 | 23377.3318 | 93.29644373 | 92.01132169 | 94.19278885 | C04218 | alpha,alpha'-Trehalose 6-mycolate | Mass, RT and Spectra | 2477.502065 | 35933.79493 | 74691.60734 | 4305.086389 | 6175.603008 | 6489.913633 |
| 637 | C15935 | 743.5233715 | 168844.6102 | 1079.46201 | 1059.240874 | 1099.896696 | C15935 | (2'S)-Deoxymyxol 2'-(2,4-di-O-methyl-alpha-L-fucoside) | Mass, RT and Spectra | 178043.1925 | 66368.48678 | 127200.8825 | 244254.5909 | 132646.2571 | 125556.0005 |
| 638 | C04271 | 257.0803094 | 117385.3968 | 357.9833962 | 343.374808 | 361.7758139 | C04271 | 3,9-Dihydroxypterocarpan | Mass, RT and Spectra | 183598.4922 | 55484.61153 | 55905.44677 | 167097.7149 | 143269.9481 | 143799.6454 |
| 639 | C04280 | 220.0934803 | 315.0894635 | 290.3207253 | 288.8239982 | 291.5695601 | C04280 | Amidino-scyllo-inosamine | Mass, RT and Spectra | 10.2045705 | 418.8351383 | #DIV/0! | 109.2127106 | 293.2468058 | 1017.8646 |
| 640 | C15937 | 759.5213079 | 43750.39166 | 1302.09848 | 1281.886211 | 1330.089844 | C15937 | (3R,2'S)-Myxol 2'-(2,4-di-O-methyl-alpha-L-fucoside) | Mass, RT and Spectra | 42008.18369 | 44272.90746 | 47085.0523 | 41993.89084 | 46142.80201 | 35754.90659 |
| 641 | C04299 | 243.026296 | 22843.46964 | 96.41089259 | 79.75791864 | 103.8805511 | C04299 | Inositol cyclic phosphate | Mass, RT and Spectra | 6073.611158 | 267.9197375 | 78.61450201 | 6612.106791 | 338.0941924 | 351.9653881 |
| 642 | C15974 | 512.126103 | 13946.85178 | 889.5441151 | 889.0366586 | 890.3221487 | C15974 | 3-Methyl-1-hydroxybutyl-ThPP | Mass, RT and Spectra | 12916.67374 | 9614.323116 | 22591.38124 | 2183.601292 | 1483.323787 | 4074.808902 |
| 643 | C04322 | 114.0548289 | 399739.1893 | 17.56632274 | 12.25608677 | 20.37907631 | C04322 | 1-Pyrroline-5-carboxylic acid | Mass, RT and Spectra | 992561.6256 | 695451.5894 | 614366.9035 | 1064102.796 | 1896829.462 | 1959189.946 |
| 644 | C04327 | 224.0150292 | 13771.38517 | 1613.512096 | 1611.110949 | 1618.567253 | C04327 | 4-Methyl-5-(2-phosphoethyl)-thiazole | Mass, RT and Spectra | 18085.19736 | 6086.324018 | 6359.662175 | 15925.90381 | 14062.46377 | 15778.41033 |
| 645 | HMDB00079 | 127.0508732 | 28827.78886 | 530.0574919 | 526.5515284 | 537.5200894 | HMDB00079 | Dihydrothymine | Mass, RT and Spectra | 250.3797145 | 61723.98293 | 61898.22085 | 7236.925268 | 6970.133943 | 6842.364863 |
| 646 | C04352 | 401.0801693 | 4476346.005 | 906.6505148 | 881.6140333 | 911.6739351 | C04352 | 4-Phosphopantothenoylcysteine | Mass, RT and Spectra | 468034.4093 | 176449.3631 | 161335.7053 | 121981.5958 | 258109.3652 | 140694.237 |
| 647 | C16038 | 223.1010622 | 20897.77887 | 397.2877779 | 379.409672 | 400.1078799 | C16038 | 1-Methyl-6-phenyl-1H-imidazo[4,5-b]pyridin-2-amine | Mass, RT and Spectra | 639.5057828 | 43021.59329 | 38755.9029 | 3145.195873 | 12317.15975 | 13801.06611 |
| 648 | C16138 | 254.1531313 | 4692.077854 | 90.70733062 | 88.09709477 | 93.19480502 | C16138 | L-Pyrrolysine | Mass, RT and Spectra | 28.77230384 | 5655.014799 | 5615.08798 | 3156.228704 | 3407.252155 | 2380.87924 |
| 649 | C04360 | 637.1085945 | 508.4758476 | 102.5632125 | 101.3104169 | 103.3862988 | C04360 | 1,2,6-Trigalloyl-beta-D-glucopyranose | Mass, RT and Spectra | 508.5994761 | 234.8071401 | 340.3685829 | 155.0764366 | 408.924866 | 1321.279329 |
| 650 | C04404 | 145.0858344 | 136925.9687 | 15.02217392 | 8.814952812 | 19.6162829 | C04404 | 4-Hydroxycyclohexylcarboxylic acid | Mass, RT | 109861.6423 | 174905.9638 | 166549.5088 | 110923.6061 | 119248.9703 | 118733.6592 |
| 651 | C04409 | 185.0310133 | 385.9334542 | 405.0989445 | 401.8714168 | 406.0975127 | C04409 | 2-Amino-3-carboxymuconic acid semialdehyde | Mass, RT and Spectra | 48.7054551 | 543.6818776 | 718.3242749 | 105.7777871 | 27.84656301 | 93.20228517 |
| 652 | C04411 | 175.0604623 | 16743.38161 | 395.740259 | 385.3506915 | 415.7986757 | C04411 | 3-Isopropylmalate | Mass, RT and Spectra | 371.2045785 | 32289.51943 | 34594.9417 | 5600.403704 | 11153.14627 | 10872.94792 |
| 653 | C04424 | 276.1081601 | 4342.654404 | 373.4571779 | 367.9534188 | 382.5963195 | C04424 | S-(2-Methylpropionyl)-dihydrolipoamide-E | Mass, RT and Spectra | 1030.83616 | 11333.14372 | 6623.871933 | 192.2753499 | 543.3788079 | 274.0736723 |
| 654 | C04431 | 139.0031667 | 3902.22605 | 153.3981788 | 148.9238869 | 157.5510853 | C04431 | cis-4-Carboxymethylenebut-2-en-4-olide | Mass, RT and Spectra | 706.2050045 | 3341.147834 | 15928.64996 | 26.64323158 | 22.30135935 | 150.8644458 |
| 655 | C04462 | 288.075117 | 15727.30488 | 427.7156969 | 422.1444995 | 429.4744498 | C04462 | N-Succinyl-2-amino-6-ketopimelate | Mass, RT and Spectra | 345.3702812 | 24912.7742 | 38005.32894 | 8580.274372 | 6491.290884 | 15827.02461 |
| 656 | C04479 | 213.0396701 | 4456.662692 | 386.9929201 | 381.8909057 | 390.2584455 | C04479 | 2-Hydroxy-6-oxonona-2,4-diene-1,9-dioate | Mass, RT and Spectra | 152.7923462 | 10401.70604 | 6985.504451 | 1445.586001 | 1070.977127 | 1997.300941 |
| 657 | HMDB00140 | 812.6953142 | 177558.7904 | 1370.102147 | 1346.23119 | 1415.635914 | HMDB00140 | Glucosylceramide | Mass, RT and Spectra | 222625.5466 | 99284.14639 | 121771.7312 | 197824.5943 | 281857.1677 | 194561.1229 |
| 658 | C16146 | 433.3142055 | 2901654.377 | 735.8195721 | 732.4954711 | 737.7910146 | C16146 | 4,4'-Diaponeurosporenic acid | Mass, RT and Spectra | 2633022.672 | 3472279.323 | 2754956.471 | 2588975.348 | 2019243.3 | 2291683.707 |
| 659 | C04483 | 391.2846842 | 799238.5007 | 104.3234728 | 93.14803617 | 118.8411431 | C04483 | Deoxycholic acid | Mass, RT and Spectra | 136486.2726 | 1939148.823 | 1885736.817 | 146701.708 | 171132.1634 | 280755.9995 |
| 660 | C04489 | 718.2775006 | 51045.31808 | 747.4532665 | 720.1540358 | 756.295388 | C04489 | 5-Methyltetrahydropteroyltri-L-glutamate | Mass, RT and Spectra | 26829.29088 | 87350.23693 | 70122.8023 | 15809.47734 | 35658.33268 | 36473.75029 |
| 661 | HMDB00770 | 744.6410118 | 6150.742611 | 1073.473899 | 1068.153297 | 1077.246764 | HMDB00770 | N-Glycoloylganglioside GM2 | Mass, RT and Spectra | 14690.1123 | 607.3369578 | 1023.914445 | 10528.95028 | 6287.669787 | 10685.01067 |
| 662 | C16155 | 533.0175031 | 29971.56877 | 291.7795038 | 287.4101945 | 300.2381625 | C16155 | UDP-L-Ara4O | Mass, RT and Spectra | 1660.395512 | 66757.95417 | 45996.81542 | 2024.152479 | 18082.13515 | 28941.79364 |
| 663 | C04540 | 336.1435053 | 85310.46905 | 679.3524502 | 666.9489247 | 685.0638018 | C04540 | Aspartylglycosamine | Mass, RT and Spectra | 123283.6554 | 44965.06731 | 40875.30677 | 126914.7257 | 118640.4385 | 128399.828 |
| 664 | C04555 | 367.1238141 | 12145.56313 | 147.9549271 | 138.962748 | 156.1544158 | C04555 | Dehydroepiandrosterone sulfate | Mass, RT and Spectra | 215.7784661 | 25745.46179 | 24123.40582 | 1132.443117 | 2977.233122 | 2809.634915 |
| 665 | C04556 | 218.0325613 | 673876.5539 | 419.2027614 | 412.4687277 | 424.3677209 | C04556 | 4-Amino-2-methyl-5-phosphomethylpyrimidine | Mass, RT and Spectra | 116768.5048 | 1352474.698 | 1235932.188 | 213914.2967 | 244121.8928 | 277254.2414 |
| 666 | C16186 | 257.0074562 | 101521.5528 | 1720.201812 | 1710.415132 | 1735.697223 | C16186 | L-Ascorbate 6-phosphate | Mass, RT and Spectra | 63296.80799 | 149461.6977 | 150182.7555 | 72349.07421 | 74341.3626 | 69454.8996 |
| 667 | C16196 | 239.0031032 | 33677.51226 | 382.4607829 | 371.2266663 | 391.6588645 | C16196 | 1,2-Dihydroxynaphthalene-6-sulfonate | Mass, RT and Spectra | 7979.061062 | 56566.99741 | 69777.53853 | 7857.013393 | 13129.23298 | 16371.019 |
| 668 | C16249 | 236.9383921 | 6003.825759 | 1777.182478 | 1768.853322 | 1792.439032 | C16249 | 2-Bromomaleylacetate | Mass, RT and Spectra | 8410.728487 | 973.6786182 | 492.7586992 | 7911.647422 | 7029.101443 | 7744.358774 |
| 669 | C04582 | 259.004309 | 4479385.947 | 131.2122391 | 125.0613235 | 144.0206862 | C04582 | 5-Methylthioribulose 1-phosphate | Mass, RT and Spectra | 352975.1017 | 321048.663 | 349330.3032 | 309618.0205 | 417997.2771 | 459753.6717 |
| 670 | C16261 | 549.3129415 | 102516.3233 | 909.2073198 | 906.8877678 | 911.4797188 | C16261 | 5beta-scymnol sulfate | Mass, RT and Spectra | 54134.79103 | 158093.4126 | 201671.5666 | 34494.65018 | 45686.50399 | 43278.90034 |
| 671 | C04592 | 127.0751702 | 139226.2482 | 14.53967396 | 11.74657097 | 19.11669617 | C04592 | Toluene-cis-dihydrodiol | Mass, RT and Spectra | 98933.5534 | 183993.8243 | 185183.521 | 101438.4953 | 113166.696 | 111836.3605 |
| 672 | C04599 | 174.1272336 | 14540.35706 | 1165.590749 | 1154.937749 | 1174.314362 | C04599 | 1-Methyl-4-phenyl-1,2,3,6-tetrahydropyridine | Mass, RT and Spectra | 20497.30469 | 1159.864232 | 7966.136403 | 18400.97872 | 4535.385069 | 25382.29208 |
| 673 | C04638 | 717.525105 | 407104.7348 | 1042.396464 | 1010.609205 | 1055.313461 | C04638 | 2,3-Bis-O-(geranylgeranyl)glycerol 1-phosphate | Mass, RT and Spectra | 443338.2173 | 147012.9679 | 318603.9589 | 528566.3901 | 383186.9973 | 389105.9931 |
| 674 | C04640 | 312.0590977 | 7984.33143 | 482.3176417 | 480.6634186 | 484.135008 | C04640 | Phosphoribosylformylglycineamidine | Mass, RT and Spectra | 10332.87338 | 155566.6041 | 200226.2403 | 26506.09829 | 24563.16094 | 15706.84377 |
| 675 | C16265 | 300.0388461 | 9731.72572 | 361.127062 | 350.1437613 | 365.1801226 | C16265 | N-Acetylgalactosamine 4-sulphate | Mass, RT and Spectra | 25420.77102 | 4681.321517 | 5431.180865 | 22102.59728 | 17598.48751 | 20557.44495 |
| 676 | C16308 | 229.1428298 | 82587.50886 | 13.69576111 | 9.523858715 | 16.16691079 | C16308 | Traumatic acid | Mass, RT and Spectra | 2060488.747 | 15689562.26 | 9458515.641 | 5152237.393 | 8359948.391 | 11109416.11 |
| 677 | C16322 | 173.1167989 | 332027.5323 | 14.34511139 | 10.00380892 | 16.9628691 | C16322 | 9-Oxononanoic acid | Mass, RT and Spectra | 284483.9689 | 397844.9229 | 397828.8658 | 285144.2009 | 285226.9121 | 304225.5575 |
| 678 | C04666 | 239.0448558 | 7937.004846 | 593.4229954 | 589.2659521 | 596.2752314 | C04666 | D-Erythro-imidazole-glycerol-phosphate | Mass, RT and Spectra | 14647.21437 | 2683.263607 | 1680.699388 | 9896.719949 | 10968.96795 | 9860.804911 |
| 679 | C04685 | 335.2220221 | 35911.02189 | 101.3745638 | 96.85778288 | 103.8026581 | C04685 | Prostaglandin A1 | Mass, RT and Spectra | 3383.229631 | 70653.65851 | 73769.65735 | 9620.298999 | 9916.470411 | 9881.218231 |
| 680 | C16352 | 298.0946104 | 9450.543668 | 515.9374943 | 512.993699 | 519.0469263 | C16352 | 7-Methylxanthosine | Mass, RT and Spectra | 11.07014666 | 21717.16405 | #DIV/0! | 4457.245083 | 887.2696801 | 1896.794509 |
| 681 | C04692 | 272.1485994 | 26778.65796 | 573.3927626 | 567.7645805 | 577.1909964 | C04692 | 2-(3-Carboxy-3-(methylammonio)propyl)-L-histidine | Mass, RT and Spectra | 43434.31145 | 10975.12702 | 9822.100146 | 39528.87445 | 37689.82599 | 39909.22443 |
| 682 | C16358 | 333.0928926 | 11741.62555 | 234.4493948 | 228.6559857 | 246.381373 | C16358 | 1-Methylxanthine | Mass, RT and Spectra | 105.7420281 | 18982.47838 | 33107.66525 | 91.02247021 | 677.9524438 | 2249.349334 |
| 683 | C04707 | 351.2199853 | 67.40000216 | 1579.274207 | 1579.184927 | 1581.649224 | C04707 | 15-Keto-prostaglandin E2 | Mass, RT and Spectra | 43.81854385 | 69.78657565 | 31.72047625 | 67.01059867 | 78.12041971 | 56.46999466 |
| 684 | C04717 | 311.2224689 | 1087420.57 | 103.5294915 | 94.73091884 | 110.4760107 | C04717 | 13-L-Hydroperoxylinoleic acid | Mass, RT and Spectra | 289575.5944 | 1616027.445 | 1285501.962 | 752281.4865 | 549267.5375 | 274443.9969 |
| 685 | C04742 | 319.2258022 | 36651.92936 | 91.96409477 | 89.19853493 | 94.06054327 | C04742 | 15(S)-HETE | Mass, RT and Spectra | 3503.693127 | 69217.80589 | 63802.46507 | 8049.623662 | 12384.85849 | 7554.782149 |
| 686 | C16366 | 197.0672644 | 7365.715374 | 176.0223833 | 169.4390561 | 185.3582425 | C16366 | 5-Acetylamino-6-amino-3-methyluracil | Mass, RT and Spectra | 27259.1104 | 184015.2719 | 192103.975 | 34703.19787 | 59623.24977 | 59851.67783 |
| 687 | C16396 | 168.0761792 | 436967.7609 | 1554.562519 | 1552.085922 | 1570.530398 | C16396 | 2,4-Diamino-6-nitrotoluene | Mass, RT and Spectra | 495636.8804 | 387401.2409 | 391398.2202 | 446085.4402 | 469063.1574 | 492758.8977 |
| 688 | C16427 | 336.1675395 | 32537.4913 | 1552.091347 | 1546.53191 | 1555.453294 | C16427 | Isopentenyl adenosine | Mass, RT and Spectra | 35862.20182 | 30796.46571 | 31515.46398 | 31662.05035 | 39033.03385 | 29809.18382 |
| 689 | C04744 | 184.0813797 | 16400.47997 | 593.5466723 | 591.668362 | 594.6802982 | C04744 | 2,6-Diamino-4-hydroxy-5-N-methylformamidopyrimidine | Mass, RT and Spectra | 3190.914817 | 29550.95694 | 27227.71498 | 6329.776155 | 11406.86812 | 8117.227857 |
| 690 | C04751 | 340.0571345 | 57115.95684 | 98.03439565 | 95.51586107 | 101.6145689 | C04751 | 5-amino-1-(5-phospho-D-ribosyl)imidazole-4-carboxylate | Mass, RT and Spectra | 2939.100305 | 1350.821113 | 1061.200292 | 3594.677901 | 3284.23068 | 2088.128269 |
| 691 | C04767 | 491.1879112 | 321283.3769 | 681.0422468 | 678.3046424 | 686.1984145 | C04767 | O-(1->4)-alpha-L-Dihydrostreptosyl-streptidine 6-phosphate | Mass, RT and Spectra | 374002.4279 | 280478.1847 | 263779.6152 | 331117.9841 | 336783.9307 | 333141.9181 |
| 692 | C16488 | 307.1504341 | 72871.1871 | 529.177525 | 520.5155793 | 534.9666912 | C16488 | 1-[(5-Amino-5-carboxypentyl)amino]-1-deoxyfructose | Mass, RT and Spectra | 17691.13368 | 154729.7632 | 114312.3122 | 17038.26024 | 34868.54089 | 25927.04597 |
| 693 | C04778 | 357.0839775 | 12172.2426 | 1182.479093 | 1172.73564 | 1194.78455 | C04778 | N1-(5-Phospho-a-D-ribosyl)-5,6-dimethylbenzimidazole | Mass, RT and Spectra | 1407.359784 | 24011.00768 | 21326.50981 | 2436.683077 | 5401.519198 | 7157.693737 |
| 694 | C04834 | 267.194907 | 60945.261 | 12.75965089 | 6.538907486 | 19.43527748 | C04834 | (2E,6E,10R,11S)-10,11-epoxy-3,7,11-trimethyltrideca-2,6-dienoic acid | Mass, RT and Spectra | 13750.50213 | 117048.6271 | 113804.7728 | 18916.52621 | 32700.99939 | 33472.29039 |
| 695 | C16489 | 389.1326222 | 8018.666379 | 425.1288195 | 418.6084722 | 427.6725139 | C16489 | Fructoselysine 6-phosphate | Mass, RT and Spectra | 12335.9605 | 3548.651135 | 2647.211577 | 11505.2168 | 16020.90509 | 20499.36151 |
| 696 | C04874 | 313.0521242 | 6960181.098 | 460.9650064 | 458.1485181 | 473.0083642 | C04874 | 7,8-Dihydroneopterin | Mass, RT and Spectra | 1047970.813 | 12989601.24 | 13517593.76 | 1949765.93 | 2947473.63 | 3146991.472 |
| 697 | C04886 | 675.2442557 | 7096.280434 | 88.54192724 | 86.27029108 | 92.47318101 | C04886 | (N-acetylneuraminosyl(a2-6)lactosamine) | Mass, RT and Spectra | 2753.679125 | 45483.59504 | 42273.53002 | 5171.260405 | 12198.85412 | 13473.36259 |
| 698 | C04895 | 496.0030359 | 68214.99666 | 245.2526258 | 236.918621 | 255.8955896 | C04895 | Dihydroneopterin triphosphate | Mass, RT and Spectra | 120628.7049 | 12772.83165 | 16180.85455 | 103270.9722 | 93070.45042 | 96340.10328 |
| 699 | C05108 | 413.3768421 | 13574.2431 | 15.88003229 | 12.21854924 | 17.85324796 | C05108 | 4,4-Dimethyl-5a-cholesta-8,24-dien-3-b-ol | Mass, RT and Spectra | 324.2836935 | 25496.6361 | 36497.17884 | 729.7140595 | 1463.908173 | 1050.606501 |
| 700 | C05122 | 516.297661 | 1282721.609 | 564.8146716 | 553.2158974 | 571.3381401 | C05122 | Taurocholic acid | Mass, RT and Spectra | 857312.5928 | 1668184.721 | 1309151.69 | 1521350.346 | 2523291.71 | 2044571.664 |
| 701 | C05123 | 124.9910225 | 62316.72093 | 261.8337241 | 247.7613933 | 271.6549512 | C05123 | 2-Hydroxyethanesulfonate | Mass, RT and Spectra | 3536.769199 | 28914.00425 | 23210.56259 | 3664.24723 | 7199.459108 | 8764.834008 |
| 702 | C05127 | 126.1024322 | 9482342.727 | 1274.830872 | 1250.094868 | 1295.937745 | C05127 | 1-Methylhistamine | Mass, RT and Spectra | 8302597.622 | 10102298.55 | 10216296.5 | 8014644.936 | 8541868.149 | 9679604.93 |
| 703 | C05132 | 155.0456329 | 155102.8505 | 293.9865581 | 278.0847032 | 314.8018452 | C05132 | Imidazolelactic acid | Mass, RT and Spectra | 38214.08586 | 257120.3437 | 232250.8013 | 58306.67591 | 98369.12301 | 114072.6649 |
| 704 | C05135 | 152.0826773 | 3588.811868 | 106.9379473 | 105.8523382 | 108.8517931 | C05135 | N-Acetylhistamine | Mass, RT and Spectra | 30.31706766 | 5826.594028 | 9557.617633 | 205.7706885 | 296.891344 | 779.5009098 |
| 705 | C16497 | 447.3095602 | 175002.4096 | 802.378682 | 794.6939511 | 805.0518932 | C16497 | 3-Dehydro-2-deoxyecdysone | Mass, RT and Spectra | 146296.4101 | 247437.448 | 206253.4678 | 111017.2666 | 86132.91545 | 92115.49789 |
| 706 | C05139 | 305.2104312 | 17853.55747 | 9.684396403 | 4.248811164 | 12.34505233 | C05139 | 16a-Hydroxydehydroisoandrosterone | Mass, RT and Spectra | 3168.511684 | 43562.53697 | 30611.24511 | 4143.104111 | 6493.505769 | 6582.884232 |
| 707 | C16526 | 309.2792008 | 495741.0955 | 90.58709399 | 89.09534253 | 91.7997241 | C16526 | Eicosenoic acid | Mass, RT and Spectra | 1301.179261 | 19936.92011 | 16721.20726 | 479.5422288 | 1100.080249 | 741.1366829 |
| 708 | C16537 | 241.216913 | 630197.3316 | 1178.686 | 1172.452768 | 1180.0895 | C16537 | Pentadecanoic acid | Mass, RT | 18.9692467 | 9373.357312 | 8547.861533 | 1466.85576 | 937.583612 | 364.867788 |
| 709 | C16551 | 278.033094 | 17633.70908 | 480.0451488 | 478.5628928 | 481.9486521 | C16551 | Alcophosphamide | Mass, RT | 1420.038818 | 32345.33918 | 22949.66817 | 8506.244728 | 8966.639762 | 13857.38426 |
| 710 | C05141 | 143.0792688 | 4738012.012 | 98.73640911 | 96.70537379 | 105.6989835 | C05141 | Estriol | Mass, RT and Spectra | 142260.9599 | 114911.5647 | 138893.6893 | 158538.2004 | 129093.8215 | 139977.1472 |
| 711 | C05145 | 102.0556865 | 132770.2105 | 396.4550979 | 387.0114894 | 405.917762 | C05145 | 3-Aminoisobutanoic acid | Mass, RT and Spectra | 2372.343768 | 268864.8966 | 259720.2448 | 51275.9132 | 70019.878 | 66069.37264 |

**Supplementary table 3: Baseline metaproteome profile of responders and non-responders.**

| peptide | lca | superkingdom | kingdom | subkingdom | superphylum | phylum | subphylum | superclass | class | subclass | infraclass | superorder | order | suborder | infraorder | parvorder | superfamily | family | subfamily | tribe | subtribe | genus | subgenus | species group | species subgroup | species | subspecies | varietas | superkingdom;phylum;subphylum;superclass;class;order;family;subfamily;genus;species group;speciessubspecies;lca | KO | EC | EC - names | GO (cellular component) | GO (molecular function) | GO (biological process) | GO (cellular component) - names | GO (molecular function) - names | GO (biological process) - names | InterPro | InterPro - names |  |  |  | R average | NR average |
| --- | --- | --- | --- | --- | --- | --- | --- | --- | --- | --- | --- | --- | --- | --- | --- | --- | --- | --- | --- | --- | --- | --- | --- | --- | --- | --- | --- | --- | --- | --- | --- | --- | --- | --- | --- | --- | --- | --- | --- | --- | --- | --- | --- | --- | --- |
| AANAAAFAA | Streptomyces | Bacteria |  |  |  | Actinobacteria |  |  | Actinobacteria |  |  |  | Streptomycetales |  |  |  |  | Streptomycetaceae |  |  |  | Streptomyces |  |  |  |  |  |  | Bacteria;Actinobacteria;;;Actinobacteria;Streptomycetales;Streptomycetaceae;;Streptomyces;;;Streptomyces | [K01462](https://www.genome.jp/dbget-bin/www_bget?ko:K01462) | 3.5.1.88 | Peptide deformylase (100%) |  |  | GO:0042586 (100%);GO:0046872 (100%) | peptide deformylase activity (100%);metal ion binding (100%) | GO:0006412 (100%);GO:0031365 (9%);GO:0043686 (9%) | translation (100%);N-terminal protein amino acid modification (9%);co-translational protein modification (9%) | IPR023635 (100%);IPR036821 (100%) | Peptide deformylase (100%);Peptide deformylase superfamily (100%) |  |  |  | 1 | 1 |
| ADVAEVANTMCDLR | Bacteria | Bacteria |  |  |  |  |  |  |  |  |  |  |  |  |  |  |  |  |  |  |  |  |  |  |  |  |  |  | Bacteria;;;;;;;;;;;Bacteria |  | 2.1.1.- | Methyltransferases (100%) |  |  | GO:0003677 (50%);GO:0008170 (50%);GO:0008233 (50%) | DNA binding (50%);N-methyltransferase activity (50%);peptidase activity (50%) | GO:0006306 (50%) | DNA methylation (50%) | IPR001091 (100%);IPR002052 (100%);IPR002941 (100%) | Restriction/modification DNA-methylase (100%);DNA methylase | N-6 adenine-specific | conserved site (100%);DNA methylase N-4/N-6 (100%) |  | 1 | 1 |
| AEGLLAVTGDGAHAAR | Mycobacterium tuberculosis complex | Bacteria |  |  |  | Actinobacteria |  |  | Actinobacteria |  |  |  | Corynebacteriales |  |  |  |  | Mycobacteriaceae |  |  |  | Mycobacterium |  | Mycobacterium tuberculosis complex |  |  |  |  | Bacteria;Actinobacteria;;;Actinobacteria;Corynebacteriales;Mycobacteriaceae;;Mycobacterium;Mycobacterium tuberculosis complex;;Mycobacterium tuberculosis complex | K11540 | 6.3.5.5 | Carbamoyl-phosphate synthase (glutamine-hydrolyzing) (100%) | GO:0005737 (7%);GO:0005886 (7%) | cytoplasm (7%);plasma membrane (7%) | GO:0004088 (100%);GO:0005524 (100%);GO:0046872 (100%) | carbamoyl-phosphate synthase (glutamine-hydrolyzing) activity (100%);ATP binding (100%);metal ion binding (100%) | GO:0006526 (100%);GO:0044205 (100%);GO:0006541 (7%) | arginine biosynthetic process (100%);'de novo' UMP biosynthetic process (100%);glutamine metabolic process (7%) | IPR005479 (100%);IPR005480 (100%);IPR005483 (100%) | Carbamoyl-phosphate synthetase large subunit-like | ATP-binding domain (100%);Carbamoyl-phosphate synthetase | large subunit oligomerisation domain (100%);Carbamoyl-phosphate synthase large subunit | CPSase domain (100%) | 1 | 1 |
| AEVVAQQGAAGEVLSCSADGLVVACGENALK | Neisseria | Bacteria |  |  |  | Proteobacteria |  |  | Betaproteobacteria |  |  |  | Neisseriales |  |  |  |  | Neisseriaceae |  |  |  | Neisseria |  |  |  |  |  |  | Bacteria;Proteobacteria;;;Betaproteobacteria;Neisseriales;Neisseriaceae;;Neisseria;;;Neisseria | [K00604](https://www.genome.jp/entry/K00604) | 2.1.2.9 | Methionyl-tRNA formyltransferase (100%) | GO:0005829 (3%) | cytosol (3%) | GO:0004479 (100%) | methionyl-tRNA formyltransferase activity (100%) | GO:0071951 (3%) | conversion of methionyl-tRNA to N-formyl-methionyl-tRNA (3%) | IPR001555 (100%);IPR002376 (100%);IPR005793 (100%) | Phosphoribosylglycinamide formyltransferase | active site (100%);Formyl transferase | N-terminal (100%);Formyl transferase | C-terminal (100%) | 1 | 1 |
| AGGNLLTYR | Bacteria | Bacteria |  |  |  |  |  |  |  |  |  |  |  |  |  |  |  |  |  |  |  |  |  |  |  |  |  |  | Bacteria;;;;;;;;;;;Bacteria |  |  |  |  |  | GO:0031177 (94%);GO:0003824 (88%);GO:0016874 (13%) | phosphopantetheine binding (94%);catalytic activity (88%);ligase activity (13%) | GO:0017000 (6%);GO:0030435 (6%) | antibiotic biosynthetic process (6%);sporulation resulting in formation of a cellular spore (6%) | IPR000873 (100%);IPR042099 (100%);IPR001242 (94%) | AMP-dependent synthetase/ligase (100%);AMP-dependent synthetase-like superfamily (100%);Condensation domain (94%) |  |  |  | 1 | 1 |
| APLLELDVQNEEHLASLAGR | Mycobacteriaceae | Bacteria |  |  |  | Actinobacteria |  |  | Actinobacteria |  |  |  | Corynebacteriales |  |  |  |  | Mycobacteriaceae |  |  |  |  |  |  |  |  |  |  | Bacteria;Actinobacteria;;;Actinobacteria;Corynebacteriales;Mycobacteriaceae;;;;;Mycobacteriaceae | [K00208](https://www.genome.jp/entry/K00208+1.3.1.10+R10122) | 1.3.1.9 | Enoyl-[acyl-carrier-protein] reductase (NADH) (100%) | GO:0005618 (2%);GO:0005886 (2%) | cell wall (2%);plasma membrane (2%) | GO:0004318 (100%);GO:0016631 (49%);GO:0005504 (2%) | enoyl-[acyl-carrier-protein] reductase (NADH) activity (100%);enoyl-[acyl-carrier-protein] reductase activity (49%);fatty acid binding (2%) | GO:0006633 (98%);GO:0046677 (5%);GO:0030497 (2%) | fatty acid biosynthetic process (98%);response to antibiotic (5%);fatty acid elongation (2%) | IPR036291 (100%);IPR014358 (94%) | NAD(P)-binding domain superfamily (100%);Enoyl-[acyl-carrier-protein] reductase (NADH) (94%) |  |  |  | 1 | 1 |
| ASLSAVAPGTSLR | Streptomyces | Bacteria |  |  |  | Actinobacteria |  |  | Actinobacteria |  |  |  | Streptomycetales |  |  |  |  | Streptomycetaceae |  |  |  | Streptomyces |  |  |  |  |  |  | Bacteria;Actinobacteria;;;Actinobacteria;Streptomycetales;Streptomycetaceae;;Streptomyces;;;Streptomyces | [K00764](https://www.genome.jp/entry/K00764) | 2.7.7.85 | Diadenylate cyclase (100%) |  |  | GO:0003677 (100%);GO:0005524 (100%);GO:0016779 (100%) | DNA binding (100%);ATP binding (100%);nucleotidyltransferase activity (100%) | GO:0006281 (100%);GO:0019932 (100%) | DNA repair (100%);second-messenger-mediated signaling (100%) | IPR003390 (100%);IPR010994 (100%);IPR018906 (100%) | DNA integrity scanning protein | DisA | N-terminal (100%);RuvA domain 2-like (100%);DNA integrity scanning | DisA | 100.5933 | 1 |
| AVLGLLKPHR | Pseudomonas | Bacteria |  |  |  | Proteobacteria |  |  | Gammaproteobacteria |  |  |  | Pseudomonadales |  |  |  |  | Pseudomonadaceae |  |  |  | Pseudomonas |  |  |  |  |  |  | Bacteria;Proteobacteria;;;Gammaproteobacteria;Pseudomonadales;Pseudomonadaceae;;Pseudomonas;;;Pseudomonas |  | 7.2.2.- | Linked to the hydrolysis of a nucleoside triphosphate (100%) | GO:0005886 (100%) | plasma membrane (100%) | GO:0005524 (100%);GO:0015633 (100%);GO:0016887 (95%) | ATP binding (100%);ATPase-coupled zinc transmembrane transporter activity (100%);ATPase activity (95%) |  |  | IPR003439 (100%);IPR003593 (100%);IPR017871 (100%) | ABC transporter-like (100%);AAA+ ATPase domain (100%);ABC transporter | conserved site (100%) |  |  | 100.6667 | 1 |
| DEFATLGK | Neisseria | Bacteria |  |  |  | Proteobacteria |  |  | Betaproteobacteria |  |  |  | Neisseriales |  |  |  |  | Neisseriaceae |  |  |  | Neisseria |  |  |  |  |  |  | Bacteria;Proteobacteria;;;Betaproteobacteria;Neisseriales;Neisseriaceae;;Neisseria;;;Neisseria | [K10536](https://www.genome.jp/entry/K10536) | 3.4.23.36 | Signal peptidase II (100%) | GO:0005886 (100%);GO:0016021 (100%) | plasma membrane (100%);integral component of membrane (100%) | GO:0004190 (100%) | aspartic-type endopeptidase activity (100%) |  |  | IPR001872 (100%) | Peptidase A8 | signal peptidase II (100%) |  |  | 1 | 1 |
| DFNQNVTFDEVGR | Mycoplasma arthritidis | Bacteria |  |  |  | Tenericutes |  |  | Mollicutes |  |  |  | Mycoplasmatales |  |  |  |  | Mycoplasmataceae |  |  |  | Mycoplasma |  |  |  | Mycoplasma arthritidis |  |  | Bacteria;Tenericutes;;;Mollicutes;Mycoplasmatales;Mycoplasmataceae;;Mycoplasma;;Mycoplasma arthritidis;Mycoplasma arthritidis | [K00762](https://www.genome.jp/entry/K00762) | 7.1.2.2 | H(+)-transporting two-sector ATPase (100%) | GO:0005886 (100%);GO:0045261 (100%) | plasma membrane (100%);proton-transporting ATP synthase complex | catalytic core F(1) (100%) | GO:0005524 (100%);GO:0046933 (100%) | ATP binding (100%);proton-transporting ATP synthase activity | rotational mechanism (100%) | GO:0015986 (100%) | ATP synthesis coupled proton transport (100%) | IPR000194 (100%);IPR000793 (100%);IPR004100 (100%) | ATPase | F1/V1/A1 complex | 100.6 | 1 |
| DLLDPQGCR | Neisseria | Bacteria |  |  |  | Proteobacteria |  |  | Betaproteobacteria |  |  |  | Neisseriales |  |  |  |  | Neisseriaceae |  |  |  | Neisseria |  |  |  |  |  |  | Bacteria;Proteobacteria;;;Betaproteobacteria;Neisseriales;Neisseriaceae;;Neisseria;;;Neisseria | [K01425](https://www.genome.jp/entry/K01425) | 3.5.1.2 ;4.3.2.10 | Glutaminase (100%);Imidazole glycerol-phosphate synthase (100%) | GO:0005737 (100%) | cytoplasm (100%) | GO:0000107 (100%);GO:0004359 (100%);GO:0016829 (100%) | imidazoleglycerol-phosphate synthase activity (100%);glutaminase activity (100%);lyase activity (100%) | GO:0000105 (100%);GO:0006541 (100%) | histidine biosynthetic process (100%);glutamine metabolic process (100%) | IPR010139 (100%);IPR017926 (100%);IPR029062 (100%) | Imidazole glycerol phosphate synthase | subunit H (100%);Glutamine amidotransferase (100%);Class I glutamine amidotransferase-like (100%) |  |  | 100.5933 | 1 |
| DMLHPSGSTK | Shewanella sediminis | Bacteria |  |  |  | Proteobacteria |  |  | Gammaproteobacteria |  |  |  | Alteromonadales |  |  |  |  | Shewanellaceae |  |  |  | Shewanella |  |  |  | Shewanella sediminis |  |  | Bacteria;Proteobacteria;;;Gammaproteobacteria;Alteromonadales;Shewanellaceae;;Shewanella;;Shewanella sediminis;Shewanella sediminis |  |  |  | GO:0005623 (100%);GO:0005887 (100%) | cell (100%);integral component of plasma membrane (100%) | GO:0015386 (100%);GO:0050660 (100%) | potassium:proton antiporter activity (100%);flavin adenine dinucleotide binding (100%) | GO:0006884 (100%) | cell volume homeostasis (100%) | IPR005170 (100%);IPR006037 (100%);IPR006153 (100%) | Transporter-associated domain (100%);Regulator of K+ conductance | C-terminal (100%);Cation/H+ exchanger (100%) |  |  | 100.74 | 1 |
| DVQNDSDAVFTAK | Bacillus | Bacteria |  |  |  | Firmicutes |  |  | Bacilli |  |  |  | Bacillales |  |  |  |  | Bacillaceae |  |  |  | Bacillus |  |  |  |  |  |  | Bacteria;Firmicutes;;;Bacilli;Bacillales;Bacillaceae;;Bacillus;;;Bacillus |  |  |  | GO:0005886 (100%);GO:0016021 (100%) | plasma membrane (100%);integral component of membrane (100%) |  |  | GO:0030435 (100%) | sporulation resulting in formation of a cellular spore (100%) | IPR029046 (100%);IPR025377 (13%) | Lipoprotein localisation LolA/LolB/LppX (100%);Domain of unknown function DUF4367 (13%) |  |  |  | 100.7333 | 1 |
| EAAEQGMGPHGLCVGATGSGK | Mycobacteriaceae | Bacteria |  |  |  | Actinobacteria |  |  | Actinobacteria |  |  |  | Corynebacteriales |  |  |  |  | Mycobacteriaceae |  |  |  |  |  |  |  |  |  |  | Bacteria;Actinobacteria;;;Actinobacteria;Corynebacteriales;Mycobacteriaceae;;;;;Mycobacteriaceae |  |  |  | GO:0016021 (86%);GO:0005886 (9%) | integral component of membrane (86%);plasma membrane (9%) | GO:0003677 (100%);GO:0005524 (100%) | DNA binding (100%);ATP binding (100%) |  |  | IPR002543 (100%);IPR027417 (100%);IPR023837 (95%) | FtsK domain (100%);P-loop containing nucleoside triphosphate hydrolase (100%);EccCb-like | Actinobacteria (95%) |  |  | 100.3267 | 1 |
| EAYSDDVPEGQVVK | Bacillus | Bacteria |  |  |  | Firmicutes |  |  | Bacilli |  |  |  | Bacillales |  |  |  |  | Bacillaceae |  |  |  | Bacillus |  |  |  |  |  |  | Bacteria;Firmicutes;;;Bacilli;Bacillales;Bacillaceae;;Bacillus;;;Bacillus | K14441 | 2.7.11.1 | Non-specific serine/threonine protein kinase (100%) | GO:0016021 (97%);GO:0005886 (3%) | integral component of membrane (97%);plasma membrane (3%) | GO:0005524 (90%);GO:0004674 (47%);GO:0004672 (43%) | ATP binding (90%);protein serine/threonine kinase activity (47%);protein kinase activity (43%) | GO:0006468 (3%);GO:0007165 (3%);GO:0009847 (3%) | protein phosphorylation (3%);signal transduction (3%);spore germination (3%) | IPR000719 (100%);IPR005543 (100%);IPR008271 (100%) | Protein kinase domain (100%);PASTA domain (100%);Serine/threonine-protein kinase | active site (100%) |  |  | 1 | 1 |
| EELAAYLTSYHNK | Bacillus | Bacteria |  |  |  | Firmicutes |  |  | Bacilli |  |  |  | Bacillales |  |  |  |  | Bacillaceae |  |  |  | Bacillus |  |  |  |  |  |  | Bacteria;Firmicutes;;;Bacilli;Bacillales;Bacillaceae;;Bacillus;;;Bacillus |  | 6.-.-.- | Ligases (100%) |  |  | GO:0016874 (100%) | ligase activity (100%) |  |  | IPR011199 (100%) | Bacillithiol biosynthesis BshC (100%) |  |  |  | 1 | 1 |
| EGLTFDDVLLVPAK | Bacteria | Bacteria |  |  |  |  |  |  |  |  |  |  |  |  |  |  |  |  |  |  |  |  |  |  |  |  |  |  | Bacteria;;;;;;;;;;;Bacteria | [K00088](https://www.genome.jp/entry/K00088) | 1.1.1.205 | IMP dehydrogenase (100%) |  |  | GO:0003938 (100%);GO:0046872 (99%);GO:0000166 (98%) | IMP dehydrogenase activity (100%);metal ion binding (99%);nucleotide binding (98%) | GO:0006177 (99%);GO:0006183 (0%);GO:0055114 (0%) | GMP biosynthetic process (99%);GTP biosynthetic process (0%);oxidation-reduction process (0%) | IPR001093 (100%);IPR013785 (100%);IPR000644 (100%) | IMP dehydrogenase/GMP reductase (100%);Aldolase-type TIM barrel (100%);CBS domain (100%) |  |  |  | 100.5267 | 1 |
| ELTDAQVEEYTSQGR | Streptomyces | Bacteria |  |  |  | Actinobacteria |  |  | Actinobacteria |  |  |  | Streptomycetales |  |  |  |  | Streptomycetaceae |  |  |  | Streptomyces |  |  |  |  |  |  | Bacteria;Actinobacteria;;;Actinobacteria;Streptomycetales;Streptomycetaceae;;Streptomyces;;;Streptomyces | [K03149](https://www.genome.jp/entry/K03149) | 6.1.1.17 | Glutamate--tRNA ligase (100%) | GO:0005737 (86%);GO:0005829 (14%) | cytoplasm (86%);cytosol (14%) | GO:0000049 (100%);GO:0004818 (100%);GO:0005524 (100%) | tRNA binding (100%);glutamate-tRNA ligase activity (100%);ATP binding (100%) | GO:0006424 (100%) | glutamyl-tRNA aminoacylation (100%) | IPR000924 (100%);IPR004527 (100%);IPR008925 (100%) | Glutamyl/glutaminyl-tRNA synthetase (100%);Glutamate-tRNA ligase | bacterial/mitochondrial (100%);Aminoacyl-tRNA synthetase | class I | anticodon-binding (100%) | 100.9333 | 1 |
| ELVHDGSLPGVR | Mycobacteriaceae | Bacteria |  |  |  | Actinobacteria |  |  | Actinobacteria |  |  |  | Corynebacteriales |  |  |  |  | Mycobacteriaceae |  |  |  |  |  |  |  |  |  |  | Bacteria;Actinobacteria;;;Actinobacteria;Corynebacteriales;Mycobacteriaceae;;;;;Mycobacteriaceae |  |  |  | GO:0005840 (100%) | ribosome (100%) | GO:0003735 (100%);GO:0019843 (91%) | structural constituent of ribosome (100%);rRNA binding (91%) | GO:0006412 (100%) | translation (100%) | IPR001209 (100%);IPR023036 (91%) | Ribosomal protein S14 (100%);Ribosomal protein S14 | bacterial/plastid (91%) |  |  | 1 | 1 |
| ETDDADYAVK | Bacillus | Bacteria |  |  |  | Firmicutes |  |  | Bacilli |  |  |  | Bacillales |  |  |  |  | Bacillaceae |  |  |  | Bacillus |  |  |  |  |  |  | Bacteria;Firmicutes;;;Bacilli;Bacillales;Bacillaceae;;Bacillus;;;Bacillus |  | 3.1.-.- | Acting on ester bonds (100%) |  |  | GO:0004721 (50%);GO:0016787 (50%) | phosphoprotein phosphatase activity (50%);hydrolase activity (50%) |  |  | IPR011990 (100%);IPR013026 (100%);IPR019734 (100%) | Tetratricopeptide-like helical domain superfamily (100%);Tetratricopeptide repeat-containing domain (100%);Tetratricopeptide repeat (100%) |  |  |  | 1 | 1 |
| EVQELVDVLLSTPNMEQR | Bacteria | Bacteria |  |  |  |  |  |  |  |  |  |  |  |  |  |  |  |  |  |  |  |  |  |  |  |  |  |  | Bacteria;;;;;;;;;;;Bacteria |  | 1.-.-.- (57%);1.12.7.2 (43%) | Oxidoreductases (57%);Ferredoxin hydrogenase (43%) | GO:0016021 (0%) | integral component of membrane (0%) | GO:0048038 (96%);GO:0051287 (96%);GO:0016151 (93%) | quinone binding (96%);NAD binding (96%);nickel cation binding (93%) |  |  | IPR001135 (100%);IPR029014 (100%);IPR038290 (100%) | NADH-quinone oxidoreductase | subunit D (100%);[NiFe]-hydrogenase | large subunit (100%);NADH-quinone oxidoreductase | subunit D superfamily (100%) | 1 | 1 |
| EYLGEELYYLGK | Mycoplasma gallisepticum | Bacteria |  |  |  | Tenericutes |  |  | Mollicutes |  |  |  | Mycoplasmatales |  |  |  |  | Mycoplasmataceae |  |  |  | Mycoplasma |  |  |  | Mycoplasma gallisepticum |  |  | Bacteria;Tenericutes;;;Mollicutes;Mycoplasmatales;Mycoplasmataceae;;Mycoplasma;;Mycoplasma gallisepticum;Mycoplasma gallisepticum | [K03621](https://www.kegg.jp/entry/K03621) | 2.3.1.274 | Phosphate acyltransferase (100%) | GO:0005737 (100%) | cytoplasm (100%) | GO:0043811 (100%) | phosphate:acyl-[acyl carrier protein] acyltransferase activity (100%) | GO:0006633 (100%);GO:0008654 (100%) | fatty acid biosynthetic process (100%);phospholipid biosynthetic process (100%) | IPR003664 (100%);IPR012281 (100%) | Fatty acid synthesis PlsX protein (100%);Phospholipid biosynthesis protein | PlsX-like (100%) |  |  | 100.66 | 1 |
| FCPSPTGNPHVGLVR | Bacteria | Bacteria |  |  |  |  |  |  |  |  |  |  |  |  |  |  |  |  |  |  |  |  |  |  |  |  |  |  | Bacteria;;;;;;;;;;;Bacteria | [K03149](https://www.genome.jp/entry/K03149) | 6.1.1.17 | Glutamate--tRNA ligase (100%) | GO:0005737 (98%);GO:0005829 (0%) | cytoplasm (98%);cytosol (0%) | GO:0004818 (100%);GO:0005524 (98%);GO:0008270 (97%) | glutamate-tRNA ligase activity (100%);ATP binding (98%);zinc ion binding (97%) | GO:0006424 (98%);GO:0006412 (0%);GO:0043039 (0%) | glutamyl-tRNA aminoacylation (98%);translation (0%);tRNA aminoacylation (0%) | IPR000924 (100%);IPR014729 (100%);IPR020058 (100%) | Glutamyl/glutaminyl-tRNA synthetase (100%);Rossmann-like alpha/beta/alpha sandwich fold (100%);Glutamyl/glutaminyl-tRNA synthetase | class Ib | catalytic domain (100%) |  | 100.6667 | 1 |
| FEALDPVVK | Agrobacterium | Bacteria |  |  |  | Proteobacteria |  |  | Alphaproteobacteria |  |  |  | Rhizobiales |  |  |  |  | Rhizobiaceae |  |  |  | Agrobacterium |  |  |  |  |  |  | Bacteria;Proteobacteria;;;Alphaproteobacteria;Rhizobiales;Rhizobiaceae;;Agrobacterium;;;Agrobacterium |  |  |  | GO:0032993 (17%) | protein-DNA complex (17%) | GO:0003677 (83%);GO:0000976 (17%);GO:0001216 (17%) | DNA binding (83%);transcription regulatory region sequence-specific DNA binding (17%);DNA-binding transcription activator activity (17%) | GO:0006355 (83%);GO:0000746 (17%);GO:0009372 (17%) | regulation of transcription | DNA-templated (83%);conjugation (17%);quorum sensing (17%) | IPR000792 (100%);IPR005143 (100%);IPR016032 (100%) | Transcription regulator LuxR | C-terminal (100%);Transcription factor LuxR-like | autoinducer-binding domain (100%);Signal transduction response regulator | 100.6733 | 1 |
| FQGEFADLNAQLAEK | Neisseria meningitidis | Bacteria |  |  |  | Proteobacteria |  |  | Betaproteobacteria |  |  |  | Neisseriales |  |  |  |  | Neisseriaceae |  |  |  | Neisseria |  |  |  | Neisseria meningitidis |  |  | Bacteria;Proteobacteria;;;Betaproteobacteria;Neisseriales;Neisseriaceae;;Neisseria;;Neisseria meningitidis;Neisseria meningitidis |  |  |  | GO:0016021 (92%) | integral component of membrane (92%) |  |  | GO:0006310 (8%) | DNA recombination (8%) | IPR003798 (100%) | DNA recombination RmuC (100%) |  |  |  | 100.6667 | 1 |
| GELGGMDYLSPER | Mycobacteriaceae | Bacteria |  |  |  | Actinobacteria |  |  | Actinobacteria |  |  |  | Corynebacteriales |  |  |  |  | Mycobacteriaceae |  |  |  |  |  |  |  |  |  |  | Bacteria;Actinobacteria;;;Actinobacteria;Corynebacteriales;Mycobacteriaceae;;;;;Mycobacteriaceae |  | 3.6.5.0 ;3.6.5.- (0%) | (100%);Acting on GTP; involved in cellular and subcellular movement (0%) | GO:0005886 (97%);GO:0005618 (0%);GO:0005829 (0%) | plasma membrane (97%);cell wall (0%);cytosol (0%) | GO:0003746 (100%);GO:0003924 (99%);GO:0005525 (99%) | translation elongation factor activity (100%);GTPase activity (99%);GTP binding (99%) | GO:0045727 (97%) | positive regulation of translation (97%) | IPR000640 (100%);IPR035647 (100%);IPR035654 (100%) | Elongation factor EFG | domain V-like (100%);EF-G domain III/V-like (100%);Elongation factor 4 | domain IV (100%) |  | 100.46 | 1 |
| GGAVVWVAR | Caulobacter vibrioides | Bacteria |  |  |  | Proteobacteria |  |  | Alphaproteobacteria |  |  |  | Caulobacterales |  |  |  |  | Caulobacteraceae |  |  |  | Caulobacter |  |  |  | Caulobacter vibrioides |  |  | Bacteria;Proteobacteria;;;Alphaproteobacteria;Caulobacterales;Caulobacteraceae;;Caulobacter;;Caulobacter vibrioides;Caulobacter vibrioides |  |  |  |  |  |  |  | GO:0006281 (100%);GO:0009432 (50%) | DNA repair (100%);SOS response (50%) | IPR017026 (100%);IPR027417 (100%) | Inducible mutagenesis protein A (100%);P-loop containing nucleoside triphosphate hydrolase (100%) |  |  |  | 100.4 | 1 |
| GLNGYLVSPEK | Mycoplasma pneumoniae | Bacteria |  |  |  | Tenericutes |  |  | Mollicutes |  |  |  | Mycoplasmatales |  |  |  |  | Mycoplasmataceae |  |  |  | Mycoplasma |  |  |  | Mycoplasma pneumoniae |  |  | Bacteria;Tenericutes;;;Mollicutes;Mycoplasmatales;Mycoplasmataceae;;Mycoplasma;;Mycoplasma pneumoniae;Mycoplasma pneumoniae |  |  |  | GO:0016021 (100%);GO:0005886 (33%) | integral component of membrane (100%);plasma membrane (33%) |  |  |  |  | IPR030940 (100%) | Uncharacterised protein MG279/MG280 (100%) |  |  |  | 1 | 1 |
| GTQAEQLVNK | Mycoplasma | Bacteria |  |  |  | Tenericutes |  |  | Mollicutes |  |  |  | Mycoplasmatales |  |  |  |  | Mycoplasmataceae |  |  |  | Mycoplasma |  |  |  |  |  |  | Bacteria;Tenericutes;;;Mollicutes;Mycoplasmatales;Mycoplasmataceae;;Mycoplasma;;;Mycoplasma | K03010 | 2.7.4.3 | Adenylate kinase (100%) | GO:0005737 (100%) | cytoplasm (100%) | GO:0004017 (100%);GO:0005524 (100%);GO:0008270 (100%) | adenylate kinase activity (100%);ATP binding (100%);zinc ion binding (100%) | GO:0044209 (100%) | AMP salvage (100%) | IPR000850 (100%);IPR006259 (100%);IPR007862 (100%) | Adenylate kinase/UMP-CMP kinase (100%);Adenylate kinase subfamily (100%);Adenylate kinase | active site lid domain (100%) |  |  | 100.46 | 1 |
| GVAGFLATQGQAR | Mycobacterium | Bacteria |  |  |  | Actinobacteria |  |  | Actinobacteria |  |  |  | Corynebacteriales |  |  |  |  | Mycobacteriaceae |  |  |  | Mycobacterium |  |  |  |  |  |  | Bacteria;Actinobacteria;;;Actinobacteria;Corynebacteriales;Mycobacteriaceae;;Mycobacterium;;;Mycobacterium | [K01447](https://www.genome.jp/dbget-bin/www_bget?ko:K01447) | 3.5.1.28 | N-acetylmuramoyl-L-alanine amidase (100%) | GO:0030288 (2%) | outer membrane-bounded periplasmic space (2%) | GO:0008745 (100%);GO:0046872 (3%) | N-acetylmuramoyl-L-alanine amidase activity (100%);metal ion binding (3%) | GO:0009253 (100%);GO:0016998 (2%);GO:0071555 (2%) | peptidoglycan catabolic process (100%);cell wall macromolecule catabolic process (2%);cell wall organization (2%) | IPR002508 (100%) | N-acetylmuramoyl-L-alanine amidase | catalytic domain (100%) |  |  | 1 | 1 |
| GVQEYGYDFTGFR | Streptomyces | Bacteria |  |  |  | Actinobacteria |  |  | Actinobacteria |  |  |  | Streptomycetales |  |  |  |  | Streptomycetaceae |  |  |  | Streptomyces |  |  |  |  |  |  | Bacteria;Actinobacteria;;;Actinobacteria;Streptomycetales;Streptomycetaceae;;Streptomyces;;;Streptomyces | K00895 | 2.7.1.90 (99%);2.7.1.11 (1%) | Diphosphate--fructose-6-phosphate 1-phosphotransferase (99%);6-phosphofructokinase (1%) | GO:0005737 (97%);GO:0005945 (1%) | cytoplasm (97%);6-phosphofructokinase complex (1%) | GO:0003872 (100%);GO:0046872 (99%);GO:0047334 (99%) | 6-phosphofructokinase activity (100%);metal ion binding (99%);diphosphate-fructose-6-phosphate 1-phosphotransferase activity (99%) | GO:0006002 (90%);GO:0006007 (1%);GO:0030388 (1%) | fructose 6-phosphate metabolic process (90%);glucose catabolic process (1%);fructose 1 | 6-bisphosphate metabolic process (1%) | IPR000023 (100%);IPR012003 (100%);IPR012829 (100%) | Phosphofructokinase domain (100%);ATP-dependent 6-phosphofructokinase | prokaryotic-type (100%);Phosphofructokinase | mixed-substrate PFK group III (100%) | 100.6733 | 1 |
| GWGLSLELPAR | Streptomyces | Bacteria |  |  |  | Actinobacteria |  |  | Actinobacteria |  |  |  | Streptomycetales |  |  |  |  | Streptomycetaceae |  |  |  | Streptomyces |  |  |  |  |  |  | Bacteria;Actinobacteria;;;Actinobacteria;Streptomycetales;Streptomycetaceae;;Streptomyces;;;Streptomyces |  | 1.-.-.- | Oxidoreductases (100%) | GO:0016020 (13%) | membrane (13%) | GO:0016491 (87%);GO:0071949 (73%);GO:0003885 (13%) | oxidoreductase activity (87%);FAD binding (73%);D-arabinono-1 | 4-lactone oxidase activity (13%) |  |  | IPR006094 (87%);IPR016166 (87%);IPR016169 (87%) | FAD linked oxidase | N-terminal (87%);FAD-binding domain | PCMH-type (87%);FAD-binding | 100.7267 | 1 |
| HLVALLQAYGAR | Corynebacterium diphtheriae | Bacteria |  |  |  | Actinobacteria |  |  | Actinobacteria |  |  |  | Corynebacteriales |  |  |  |  | Corynebacteriaceae |  |  |  | Corynebacterium |  |  |  | Corynebacterium diphtheriae |  |  | Bacteria;Actinobacteria;;;Actinobacteria;Corynebacteriales;Corynebacteriaceae;;Corynebacterium;;Corynebacterium diphtheriae;Corynebacterium diphtheriae | K01892 | 6.3.4.19 | tRNA(Ile)-lysidine synthetase (100%) | GO:0005737 (100%) | cytoplasm (100%) | GO:0005524 (100%);GO:0016879 (100%) | ATP binding (100%);ligase activity | forming carbon-nitrogen bonds (100%) | GO:0006400 (100%) | tRNA modification (100%) | IPR011063 (100%);IPR012094 (100%);IPR012795 (100%) | tRNA(Ile)-lysidine/2-thiocytidine synthase | N-terminal (100%);tRNA(Ile)-lysidine synthase (100%);tRNA(Ile)-lysidine synthase | N-terminal (100%) | 1 | 1 |
| HVGVVCNPR | Streptomyces | Bacteria |  |  |  | Actinobacteria |  |  | Actinobacteria |  |  |  | Streptomycetales |  |  |  |  | Streptomycetaceae |  |  |  | Streptomyces |  |  |  |  |  |  | Bacteria;Actinobacteria;;;Actinobacteria;Streptomycetales;Streptomycetaceae;;Streptomyces;;;Streptomyces | [K01462](https://www.genome.jp/dbget-bin/www_bget?ko:K01462) | 3.5.1.88 | Peptide deformylase (100%) |  |  | GO:0042586 (100%);GO:0046872 (97%) | peptide deformylase activity (100%);metal ion binding (97%) | GO:0006412 (97%);GO:0031365 (3%);GO:0043686 (3%) | translation (97%);N-terminal protein amino acid modification (3%);co-translational protein modification (3%) | IPR023635 (100%);IPR036821 (100%) | Peptide deformylase (100%);Peptide deformylase superfamily (100%) |  |  |  | 100.7333 | 1 |
| LEQSSSEEAR | Enterobacteriaceae | Bacteria |  |  |  | Proteobacteria |  |  | Gammaproteobacteria |  |  |  | Enterobacterales |  |  |  |  | Enterobacteriaceae |  |  |  |  |  |  |  |  |  |  | Bacteria;Proteobacteria;;;Gammaproteobacteria;Enterobacterales;Enterobacteriaceae;;;;;Enterobacteriaceae |  |  |  | GO:0005887 (100%) | integral component of plasma membrane (100%) | GO:0015299 (100%);GO:0015503 (100%);GO:0015643 (100%) | solute:proton antiporter activity (100%);glutathione-regulated potassium exporter activity (100%);toxic substance binding (100%) | GO:0051595 (100%) | response to methylglyoxal (100%) | IPR003148 (100%);IPR004771 (100%);IPR006036 (100%) | Regulator of K+ conductance | N-terminal (100%);K+/H+ exchanger (100%);Potassium uptake protein TrkA (100%) |  |  | 1 | 1 |
| LGVALGADPLTFSGLAGLGDLVATCSSPLSR | Actinobacteria | Bacteria |  |  |  | Actinobacteria |  |  | Actinobacteria |  |  |  |  |  |  |  |  |  |  |  |  |  |  |  |  |  |  |  | Bacteria;Actinobacteria;;;Actinobacteria;;;;;;;Actinobacteria | K00057 | 1.1.1.94 | Glycerol-3-phosphate dehydrogenase (NAD(P)(+)) (100%) | GO:0009331 (93%);GO:0005737 (7%);GO:0005829 (1%) | glycerol-3-phosphate dehydrogenase complex (93%);cytoplasm (7%);cytosol (1%) | GO:0004367 (100%);GO:0051287 (93%);GO:0047952 (1%) | glycerol-3-phosphate dehydrogenase [NAD+] activity (100%);NAD binding (93%);glycerol-3-phosphate dehydrogenase [NAD(P)+] activity (1%) | GO:0006650 (100%);GO:0008654 (100%);GO:0046167 (100%) | glycerophospholipid metabolic process (100%);phospholipid biosynthetic process (100%);glycerol-3-phosphate biosynthetic process (100%) | IPR006109 (100%);IPR006168 (100%);IPR008927 (100%) | Glycerol-3-phosphate dehydrogenase | NAD-dependent | C-terminal (100%);Glycerol-3-phosphate dehydrogenase | NAD-dependent (100%);6-phosphogluconate dehydrogenase-like | 100.54 | 1 |
| LHVGGDFQDPR | Petrotoga | Bacteria |  |  |  | Thermotogae |  |  | Thermotogae |  |  |  | Petrotogales |  |  |  |  | Petrotogaceae |  |  |  | Petrotoga |  |  |  |  |  |  | Bacteria;Thermotogae;;;Thermotogae;Petrotogales;Petrotogaceae;;Petrotoga;;;Petrotoga |  |  |  |  |  |  |  |  |  | IPR001296 (100%);IPR007345 (100%);IPR011990 (100%) | Glycosyl transferase | family 1 (100%);Polysaccharide pyruvyl transferase (100%);Tetratricopeptide-like helical domain superfamily (100%) |  |  | 100.8667 | 1 |
| LNEALEAQGDR | Enterobacteriaceae | Bacteria |  |  |  | Proteobacteria |  |  | Gammaproteobacteria |  |  |  | Enterobacterales |  |  |  |  | Enterobacteriaceae |  |  |  |  |  |  |  |  |  |  | Bacteria;Proteobacteria;;;Gammaproteobacteria;Enterobacterales;Enterobacteriaceae;;;;;Enterobacteriaceae |  |  |  | GO:0005829 (100%) | cytosol (100%) |  |  | GO:0033194 (100%) | response to hydroperoxide (100%) | IPR005583 (100%) | Peroxide stress protein YaaA (100%) |  |  |  | 1 | 1 |
| LNNVLNTLK | Bacteria | Bacteria |  |  |  |  |  |  |  |  |  |  |  |  |  |  |  |  |  |  |  |  |  |  |  |  |  |  | Bacteria;;;;;;;;;;;Bacteria | [K03782](https://www.genome.jp/entry/K03782+1.11.1.21+R02670) | 1.11.1.21 (67%);2.7.13.3 (33%) | Catalase peroxidase (67%);Histidine kinase (33%) |  |  | GO:0005524 (73%);GO:0000155 (64%);GO:0004096 (18%) | ATP binding (73%);phosphorelay sensor kinase activity (64%);catalase activity (18%) | GO:0006979 (18%);GO:0042744 (18%) | response to oxidative stress (18%);hydrogen peroxide catabolic process (18%) | IPR003594 (80%);IPR003661 (80%);IPR004358 (80%) | Histidine kinase/HSP90-like ATPase (80%);Signal transduction histidine kinase | dimerisation/phosphoacceptor domain (80%);Signal transduction histidine kinase-related protein | C-terminal (80%) |  | 1 | 1 |
| LSLYMGWLK | Herbinix hemicellulosilytica | Bacteria |  |  |  | Firmicutes |  |  | Clostridia |  |  |  | Clostridiales |  |  |  |  | Lachnospiraceae |  |  |  | Herbinix |  |  |  | Herbinix hemicellulosilytica |  |  | Bacteria;Firmicutes;;;Clostridia;Clostridiales;Lachnospiraceae;;Herbinix;;Herbinix hemicellulosilytica;Herbinix hemicellulosilytica |  |  |  |  |  |  |  |  |  |  |  |  |  |  | 100.9333 | 1 |
| LSVPAGTDDVR | Streptomyces | Bacteria |  |  |  | Actinobacteria |  |  | Actinobacteria |  |  |  | Streptomycetales |  |  |  |  | Streptomycetaceae |  |  |  | Streptomyces |  |  |  |  |  |  | Bacteria;Actinobacteria;;;Actinobacteria;Streptomycetales;Streptomycetaceae;;Streptomyces;;;Streptomyces |  | 2.3.1.- | Transferring groups other than amino-acyl groups (100%) |  |  | GO:0008080 (91%);GO:0016740 (5%);GO:0034069 (5%) | N-acetyltransferase activity (91%);transferase activity (5%);aminoglycoside N-acetyltransferase activity (5%) | GO:0030649 (5%) | aminoglycoside antibiotic catabolic process (5%) | IPR016181 (100%);IPR022902 (100%);IPR025559 (100%) | Acyl-CoA N-acyltransferase (100%);N-acetyltransferase Eis (100%);Enhanced intracellular survival protein domain (100%) |  |  |  | 100.8 | 1 |
| LTGGPVANLK | Nocardiopsaceae | Bacteria |  |  |  | Actinobacteria |  |  | Actinobacteria |  |  |  | Streptosporangiales |  |  |  |  | Nocardiopsaceae |  |  |  |  |  |  |  |  |  |  | Bacteria;Actinobacteria;;;Actinobacteria;Streptosporangiales;Nocardiopsaceae;;;;;Nocardiopsaceae | K14441 | 2.8.4.4 | [Ribosomal protein S12] (aspartate(89)-C(3))-methylthiotransferase (100%) | GO:0005737 (100%);GO:0005840 (93%) | cytoplasm (100%);ribosome (93%) | GO:0046872 (100%);GO:0051539 (100%);GO:0103039 (100%) | metal ion binding (100%);4 iron | 4 sulfur cluster binding (100%);protein methylthiotransferase activity (100%) | GO:0006400 (100%);GO:0018339 (100%) | tRNA modification (100%);peptidyl-L-beta-methylthioaspartic acid biosynthetic process from peptidyl-aspartic acid (100%) | IPR005839 (100%);IPR005840 (100%);IPR006638 (100%) | Methylthiotransferase (100%);Ribosomal protein S12 methylthiotransferase RimO (100%);Elp3/MiaB/NifB (100%) |  |  | 100.6733 | 1 |
| LTPAHLHVLK | Bacillales | Bacteria |  |  |  | Firmicutes |  |  | Bacilli |  |  |  | Bacillales |  |  |  |  |  |  |  |  |  |  |  |  |  |  |  | Bacteria;Firmicutes;;;Bacilli;Bacillales;;;;;;Bacillales |  | 2.3.1.- | Transferring groups other than amino-acyl groups (100%) |  |  | GO:0031177 (92%);GO:0008483 (77%);GO:0030170 (77%) | phosphopantetheine binding (92%);transaminase activity (77%);pyridoxal phosphate binding (77%) | GO:0009058 (8%);GO:0017000 (3%) | biosynthetic process (8%);antibiotic biosynthetic process (3%) | IPR000873 (100%);IPR010071 (99%);IPR020845 (99%) | AMP-dependent synthetase/ligase (100%);Amino acid adenylation domain (99%);AMP-binding | conserved site (99%) |  |  | 1 | 1 |
| LVEGATSFGGVR | Streptomyces | Bacteria |  |  |  | Actinobacteria |  |  | Actinobacteria |  |  |  | Streptomycetales |  |  |  |  | Streptomycetaceae |  |  |  | Streptomyces |  |  |  |  |  |  | Bacteria;Actinobacteria;;;Actinobacteria;Streptomycetales;Streptomycetaceae;;Streptomyces;;;Streptomyces | [K01758](https://www.genome.jp/entry/K01758) | 4.4.1.1 (93%);4.4.1.11 (7%) | Cystathionine gamma-lyase (93%);Methionine gamma-lyase (7%) | GO:0005737 (4%) | cytoplasm (4%) | GO:0030170 (96%);GO:0004123 (52%);GO:0044540 (52%) | pyridoxal phosphate binding (96%);cystathionine gamma-lyase activity (52%);L-cystine L-cysteine-lyase (deaminating) (52%) | GO:0019346 (96%);GO:0019343 (4%) | transsulfuration (96%);cysteine biosynthetic process via cystathionine (4%) | IPR000277 (100%);IPR015421 (100%);IPR015422 (100%) | Cys/Met metabolism | pyridoxal phosphate-dependent enzyme (100%);Pyridoxal phosphate-dependent transferase | major domain (100%);Pyridoxal phosphate-dependent transferase domain 1 (100%) |  | 100.6733 | 1 |
| LVQWGAAYGLK | Bacillus subtilis group | Bacteria |  |  |  | Firmicutes |  |  | Bacilli |  |  |  | Bacillales |  |  |  |  | Bacillaceae |  |  |  | Bacillus |  | Bacillus subtilis group |  |  |  |  | Bacteria;Firmicutes;;;Bacilli;Bacillales;Bacillaceae;;Bacillus;Bacillus subtilis group;;Bacillus subtilis group |  |  |  |  |  |  |  |  |  |  |  |  |  |  | 1 | 1 |
| LYVDTDADLR | Bacteria | Bacteria |  |  |  |  |  |  |  |  |  |  |  |  |  |  |  |  |  |  |  |  |  |  |  |  |  |  | Bacteria;;;;;;;;;;;Bacteria | K00876 | 2.7.1.48 | Uridine kinase (100%) | GO:0005737 (97%);GO:0005874 (0%) | cytoplasm (97%);microtubule (0%) | GO:0004849 (99%);GO:0005524 (98%);GO:0016773 (87%) | uridine kinase activity (99%);ATP binding (98%);phosphotransferase activity | alcohol group as acceptor (87%) | GO:0044206 (98%);GO:0044211 (97%);GO:0007017 (0%) | UMP salvage (98%);CTP salvage (97%);microtubule-based process (0%) | IPR006083 (100%);IPR027417 (100%);IPR000764 (99%) | Phosphoribulokinase/uridine kinase (100%);P-loop containing nucleoside triphosphate hydrolase (100%);Uridine kinase-like (99%) |  |  | 1 | 1 |
| MAQEQTQR | Actinobacteria | Bacteria |  |  |  | Actinobacteria |  |  | Actinobacteria |  |  |  |  |  |  |  |  |  |  |  |  |  |  |  |  |  |  |  | Bacteria;Actinobacteria;;;Actinobacteria;;;;;;;Actinobacteria |  |  |  |  |  | GO:0031386 (100%);GO:0070628 (100%) | protein tag (100%);proteasome binding (100%) | GO:0010498 (100%);GO:0019941 (100%);GO:0070490 (100%) | proteasomal protein catabolic process (100%);modification-dependent protein catabolic process (100%);protein pupylation (100%) | IPR008515 (100%) | Ubiquitin-like | Pup (100%) |  |  | 100.5333 | 1 |
| MGLMLDMSPR | Firmicutes | Bacteria |  |  |  | Firmicutes |  |  |  |  |  |  |  |  |  |  |  |  |  |  |  |  |  |  |  |  |  |  | Bacteria;Firmicutes;;;;;;;;;;Firmicutes | K01390 | 2.7.7.6 | DNA-directed RNA polymerase (100%) |  |  | GO:0003677 (100%);GO:0003899 (100%);GO:0000287 (95%) | DNA binding (100%);DNA-directed 5'-3' RNA polymerase activity (100%);magnesium ion binding (95%) | GO:0006351 (100%) | transcription | DNA-templated (100%) | IPR006592 (100%);IPR007080 (100%);IPR000722 (97%) | RNA polymerase | N-terminal (100%);RNA polymerase Rpb1 | domain 1 (100%);RNA polymerase | 1 | 1 |
| NAAAESAR | Bacteria | Bacteria |  |  |  |  |  |  |  |  |  |  |  |  |  |  |  |  |  |  |  |  |  |  |  |  |  |  | Bacteria;;;;;;;;;;;Bacteria | K03469 | 3.1.26.4 | Ribonuclease H (100%) | GO:0005737 (97%);GO:0016021 (3%);GO:0032299 (3%) | cytoplasm (97%);integral component of membrane (3%);ribonuclease H2 complex (3%) | GO:0000287 (97%);GO:0004523 (97%);GO:0003723 (87%) | magnesium ion binding (97%);RNA-DNA hybrid ribonuclease activity (97%);RNA binding (87%) | GO:0006401 (93%);GO:0006298 (3%);GO:0043137 (3%) | RNA catabolic process (93%);mismatch repair (3%);DNA replication | removal of RNA primer (3%) | IPR001352 (91%);IPR004641 (91%);IPR012295 (91%) | Ribonuclease HII/HIII (91%);Ribonuclease HIII (91%);TBP domain superfamily (91%) |  |  | 1 | 1 |
| NATYLDSSALGMLLLLR | Bacteria | Bacteria |  |  |  |  |  |  |  |  |  |  |  |  |  |  |  |  |  |  |  |  |  |  |  |  |  |  | Bacteria;;;;;;;;;;;Bacteria |  |  |  | GO:0005576 (50%) | extracellular region (50%) | GO:0043856 (50%) | anti-sigma factor antagonist activity (50%) |  |  | IPR002645 (100%);IPR036513 (100%) | STAS domain (100%);STAS domain superfamily (100%) |  |  |  | 100.4 | 1 |
| NAVDAQQK | Bacillus | Bacteria |  |  |  | Firmicutes |  |  | Bacilli |  |  |  | Bacillales |  |  |  |  | Bacillaceae |  |  |  | Bacillus |  |  |  |  |  |  | Bacteria;Firmicutes;;;Bacilli;Bacillales;Bacillaceae;;Bacillus;;;Bacillus |  |  |  |  |  |  |  |  |  | IPR008863 (100%) | Toxic anion resistance (100%) |  |  |  | 1 | 1 |
| NETDQLLFQTDK | Lactobacillus | Bacteria |  |  |  | Firmicutes |  |  | Bacilli |  |  |  | Lactobacillales |  |  |  |  | Lactobacillaceae |  |  |  | Lactobacillus |  |  |  |  |  |  | Bacteria;Firmicutes;;;Bacilli;Lactobacillales;Lactobacillaceae;;Lactobacillus;;;Lactobacillus |  |  |  |  |  | GO:0005524 (100%);GO:0051082 (94%);GO:0016787 (6%) | ATP binding (100%);unfolded protein binding (94%);hydrolase activity (6%) | GO:0006457 (94%) | protein folding (94%) | IPR013126 (100%);IPR029048 (100%);IPR012725 (94%) | Heat shock protein 70 family (100%);Heat shock protein 70kD | C-terminal domain superfamily (100%);Chaperone DnaK (94%) |  |  | 100.46 | 1 |
| NEYVGAVVMK | Corynebacterium | Bacteria |  |  |  | Actinobacteria |  |  | Actinobacteria |  |  |  | Corynebacteriales |  |  |  |  | Corynebacteriaceae |  |  |  | Corynebacterium |  |  |  |  |  |  | Bacteria;Actinobacteria;;;Actinobacteria;Corynebacteriales;Corynebacteriaceae;;Corynebacterium;;;Corynebacterium | [K09759](https://www.genome.jp/entry/K09759) | 6.1.1.23 | Aspartate--tRNA(Asn) ligase (100%) | GO:0005737 (100%) | cytoplasm (100%) | GO:0003676 (100%);GO:0004815 (100%);GO:0005524 (100%) | nucleic acid binding (100%);aspartate-tRNA ligase activity (100%);ATP binding (100%) | GO:0006422 (100%) | aspartyl-tRNA aminoacylation (100%) | IPR002312 (100%);IPR004115 (100%);IPR004364 (100%) | Aspartyl/Asparaginyl-tRNA synthetase | class IIb (100%);GAD-like domain superfamily (100%);Aminoacyl-tRNA synthetase | class II (D/K/N) (100%) |  | 1 | 1 |
| NLGAVAAPTASLHFSETMFENLR | Campylobacter | Bacteria |  |  |  | Proteobacteria | delta/epsilon subdivisions |  | Epsilonproteobacteria |  |  |  | Campylobacterales |  |  |  |  | Campylobacteraceae |  |  |  | Campylobacter |  |  |  |  |  |  | Bacteria;Proteobacteria;delta/epsilon subdivisions;;Epsilonproteobacteria;Campylobacterales;Campylobacteraceae;;Campylobacter;;;Campylobacter | [K07568](https://www.genome.jp/entry/K07568) | 2.4.99.17 | S-adenosylmethionine:tRNA ribosyltransferase-isomerase (100%) | GO:0005737 (100%) | cytoplasm (100%) | GO:0016853 (100%);GO:0016740 (50%);GO:0016757 (50%) | isomerase activity (100%);transferase activity (50%);transferase activity | transferring glycosyl groups (50%) | GO:0008616 (100%) | queuosine biosynthetic process (100%) | IPR003699 (100%);IPR036100 (100%);IPR042118 (100%) | S-adenosylmethionine:tRNA ribosyltransferase-isomerase | QueA (100%);S-adenosylmethionine:tRNA ribosyltransferase-isomerase | QueA superfamily (100%);QueA | 1 | 1 |
| NNGDGSR | Neisseria | Bacteria |  |  |  | Proteobacteria |  |  | Betaproteobacteria |  |  |  | Neisseriales |  |  |  |  | Neisseriaceae |  |  |  | Neisseria |  |  |  |  |  |  | Bacteria;Proteobacteria;;;Betaproteobacteria;Neisseriales;Neisseriaceae;;Neisseria;;;Neisseria |  |  |  | GO:0005829 (4%) | cytosol (4%) | GO:0003690 (96%);GO:0003700 (96%);GO:0003677 (4%) | double-stranded DNA binding (96%);DNA-binding transcription factor activity (96%);DNA binding (4%) | GO:0006355 (4%) | regulation of transcription | DNA-templated (4%) | IPR000944 (100%);IPR010242 (100%);IPR030489 (100%) | Transcription regulator Rrf2-type (100%);Transcription factor HTH | IscR (100%);Transcription regulator Rrf2-type | conserved site (100%) | 100.6667 | 1 |
| NPVWYHNVVK | Mycobacteriaceae | Bacteria |  |  |  | Actinobacteria |  |  | Actinobacteria |  |  |  | Corynebacteriales |  |  |  |  | Mycobacteriaceae |  |  |  |  |  |  |  |  |  |  | Bacteria;Actinobacteria;;;Actinobacteria;Corynebacteriales;Mycobacteriaceae;;;;;Mycobacteriaceae |  | 1.1.98.- (60%);1.-.-.- (40%) | With other | known | acceptors (60%);Oxidoreductases (40%) | GO:0005886 (15%);GO:0005618 (5%) | plasma membrane (15%);cell wall (5%) | GO:0016491 (100%);GO:0048037 (95%);GO:0070967 (5%) | oxidoreductase activity (100%);cofactor binding (95%);coenzyme F420 binding (5%) | GO:0055114 (5%) | oxidation-reduction process (5%) | IPR004378 (100%);IPR012349 (100%) | F420H(2)-dependent quinone reductase (100%);FMN-binding split barrel (100%) |  | 100.9333 | 1 |
| NSCSCSFCGK | canis group | Bacteria |  |  |  | Proteobacteria |  |  | Alphaproteobacteria |  |  |  | Rickettsiales |  |  |  |  | Anaplasmataceae |  |  |  | Ehrlichia |  | canis group |  |  |  |  | Bacteria;Proteobacteria;;;Alphaproteobacteria;Rickettsiales;Anaplasmataceae;;Ehrlichia;canis group;;canis group |  |  |  |  |  | GO:0005524 (100%);GO:0008270 (100%);GO:0046983 (100%) | ATP binding (100%);zinc ion binding (100%);protein dimerization activity (100%) | GO:0006457 (100%) | protein folding (100%) | IPR003593 (100%);IPR003959 (100%);IPR004487 (100%) | AAA+ ATPase domain (100%);ATPase | AAA-type | core (100%);Clp protease | ATP-binding subunit ClpX (100%) | 1 | 1 |
| QCSVAASCADLALATGFLAR | Streptomyces | Bacteria |  |  |  | Actinobacteria |  |  | Actinobacteria |  |  |  | Streptomycetales |  |  |  |  | Streptomycetaceae |  |  |  | Streptomyces |  |  |  |  |  |  | Bacteria;Actinobacteria;;;Actinobacteria;Streptomycetales;Streptomycetaceae;;Streptomyces;;;Streptomyces | [K01425](https://www.genome.jp/entry/K01425) | 3.5.1.2 | Glutaminase (100%) |  |  | GO:0004359 (100%) | glutaminase activity (100%) | GO:0006541 (90%);GO:0006537 (3%);GO:0006543 (3%) | glutamine metabolic process (90%);glutamate biosynthetic process (3%);glutamine catabolic process (3%) | IPR012338 (100%);IPR015868 (100%) | Beta-lactamase/transpeptidase-like (100%);Glutaminase (100%) |  |  |  | 100.6667 | 1 |
| QEGPELLELVEK | Streptomyces | Bacteria |  |  |  | Actinobacteria |  |  | Actinobacteria |  |  |  | Streptomycetales |  |  |  |  | Streptomycetaceae |  |  |  | Streptomyces |  |  |  |  |  |  | Bacteria;Actinobacteria;;;Actinobacteria;Streptomycetales;Streptomycetaceae;;Streptomyces;;;Streptomyces | [K01951](https://www.genome.jp/entry/K01951) | 4.1.1.31 | Phosphoenolpyruvate carboxylase (100%) | GO:0005829 (1%) | cytosol (1%) | GO:0008964 (100%);GO:0000287 (96%);GO:0016301 (1%) | phosphoenolpyruvate carboxylase activity (100%);magnesium ion binding (96%);kinase activity (1%) | GO:0015977 (98%);GO:0006107 (96%);GO:0006099 (93%) | carbon fixation (98%);oxaloacetate metabolic process (96%);tricarboxylic acid cycle (93%) | IPR015813 (100%);IPR021135 (100%);IPR018129 (99%) | Pyruvate/Phosphoenolpyruvate kinase-like domain superfamily (100%);Phosphoenolpyruvate carboxylase (100%);Phosphoenolpyruvate carboxylase | Lys active site (99%) |  |  | 100.9333 | 1 |
| QMQPDMVMY | Mycobacteriaceae | Bacteria |  |  |  | Actinobacteria |  |  | Actinobacteria |  |  |  | Corynebacteriales |  |  |  |  | Mycobacteriaceae |  |  |  |  |  |  |  |  |  |  | Bacteria;Actinobacteria;;;Actinobacteria;Corynebacteriales;Mycobacteriaceae;;;;;Mycobacteriaceae | [K09759](https://www.kegg.jp/entry/K09759+6.1.1.23+R03647) | 2.7.7.- ;2.8.1.- | Nucleotidyltransferases (100%);Sulfurtransferases (100%) | GO:0016021 (42%);GO:0005618 (2%);GO:0005737 (2%) | integral component of membrane (42%);cell wall (2%);cytoplasm (2%) | GO:0008641 (98%);GO:0016779 (67%);GO:0005524 (3%) | ubiquitin-like modifier activating enzyme activity (98%);nucleotidyltransferase activity (67%);ATP binding (3%) | GO:0006535 (2%);GO:0008152 (2%);GO:0019344 (2%) | cysteine biosynthetic process from serine (2%);metabolic process (2%);cysteine biosynthetic process (2%) | IPR001763 (100%);IPR036873 (100%);IPR035985 (98%) | Rhodanese-like domain (100%);Rhodanese-like domain superfamily (100%);Ubiquitin-activating enzyme (98%) |  |  |  | 100.6667 | 1 |
| QSSDSQEQLAEHK | Bacteria | Bacteria |  |  |  |  |  |  |  |  |  |  |  |  |  |  |  |  |  |  |  |  |  |  |  |  |  |  | Bacteria;;;;;;;;;;;Bacteria | K01390 | 3.4.24.13 (92%);3.4.24.- (8%);3.4.21.- (6%) | IgA-specific metalloendopeptidase (92%);Metalloendopeptidases (8%);Serine endopeptidases (6%) | GO:0005576 (99%);GO:0005618 (99%);GO:0016021 (99%) | extracellular region (99%);cell wall (99%);integral component of membrane (99%) | GO:0004222 (89%);GO:0008270 (89%);GO:0008233 (7%) | metalloendopeptidase activity (89%);zinc ion binding (89%);peptidase activity (7%) | GO:0009405 (3%) | pathogenesis (3%) | IPR019931 (97%);IPR019948 (97%);IPR005877 (96%) | LPXTG cell wall anchor domain (97%);Gram-positive LPXTG cell wall anchor (97%);YSIRK Gram-positive signal peptide (96%) |  |  |  | 100.9333 | 1 |
| QVAETDAWEATSGGK | Streptomyces | Bacteria |  |  |  | Actinobacteria |  |  | Actinobacteria |  |  |  | Streptomycetales |  |  |  |  | Streptomycetaceae |  |  |  | Streptomyces |  |  |  |  |  |  | Bacteria;Actinobacteria;;;Actinobacteria;Streptomycetales;Streptomycetaceae;;Streptomyces;;;Streptomyces |  | 3.4.11.- | Aminopeptidases (100%) | GO:0005737 (8%) | cytoplasm (8%) | GO:0008270 (100%);GO:0004177 (92%);GO:0008237 (92%) | zinc ion binding (100%);aminopeptidase activity (92%);metallopeptidase activity (92%) |  |  | IPR001948 (100%);IPR022984 (100%);IPR023358 (100%) | Peptidase M18 (100%);M18 family aminopeptidase 2 | putative (100%);Peptidase M18 | domain 2 (100%) |  | 100.6667 | 1 |
| QVPDNGGMYFVPAFSGLFAPYWR | Actinobacteria | Bacteria |  |  |  | Actinobacteria |  |  | Actinobacteria |  |  |  |  |  |  |  |  |  |  |  |  |  |  |  |  |  |  |  | Bacteria;Actinobacteria;;;Actinobacteria;;;;;;;Actinobacteria | [K03782](https://www.genome.jp/entry/K03782+1.11.1.21+R02670) | 2.7.1.30 | Glycerol kinase (100%) | GO:0005829 (5%) | cytosol (5%) | GO:0004370 (80%);GO:0005524 (75%);GO:0016301 (20%) | glycerol kinase activity (80%);ATP binding (75%);kinase activity (20%) | GO:0006072 (75%);GO:0019563 (75%);GO:0005975 (25%) | glycerol-3-phosphate metabolic process (75%);glycerol catabolic process (75%);carbohydrate metabolic process (25%) | IPR018485 (100%);IPR018483 (95%);IPR018484 (90%) | Carbohydrate kinase | FGGY | C-terminal (100%);Carbohydrate kinase | FGGY | 1 | 1 |
| RPGTPEAR | Actinobacteria | Bacteria |  |  |  | Actinobacteria |  |  | Actinobacteria |  |  |  |  |  |  |  |  |  |  |  |  |  |  |  |  |  |  |  | Bacteria;Actinobacteria;;;Actinobacteria;;;;;;;Actinobacteria | [K04042](https://www.genome.jp/dbget-bin/www_bget?ko+K04042) | 2.3.1.157 ;2.7.7.23 | Glucosamine-1-phosphate N-acetyltransferase (100%);UDP-N-acetylglucosamine diphosphorylase (100%) | GO:0005737 (100%) | cytoplasm (100%) | GO:0000287 (100%);GO:0003977 (100%);GO:0019134 (100%) | magnesium ion binding (100%);UDP-N-acetylglucosamine diphosphorylase activity (100%);glucosamine-1-phosphate N-acetyltransferase activity (100%) | GO:0000902 (100%);GO:0006048 (100%);GO:0008360 (100%) | cell morphogenesis (100%);UDP-N-acetylglucosamine biosynthetic process (100%);regulation of cell shape (100%) | IPR001451 (100%);IPR005882 (100%);IPR011004 (100%) | Hexapeptide repeat (100%);Bifunctional UDP-N-acetylglucosamine pyrophosphorylase/glucosamine-1-phosphate N-acetyltransferase (100%);Trimeric LpxA-like superfamily (100%) |  |  |  | 100.8667 | 1 |
| SDGGYAVSSYR | Bacteria | Bacteria |  |  |  |  |  |  |  |  |  |  |  |  |  |  |  |  |  |  |  |  |  |  |  |  |  |  | Bacteria;;;;;;;;;;;Bacteria | [K05341](https://www.genome.jp/entry/K05341) | 2.4.1.4 | Amylosucrase (100%) | GO:0005576 (5%) | extracellular region (5%) | GO:0003824 (76%);GO:0047669 (24%) | catalytic activity (76%);amylosucrase activity (24%) | GO:0005975 (100%) | carbohydrate metabolic process (100%) | IPR006047 (100%);IPR017853 (100%);IPR013780 (93%) | Glycosyl hydrolase | family 13 | catalytic domain (100%);Glycoside hydrolase superfamily (100%);Glycosyl hydrolase | all-beta (93%) | 100.6667 | 1 |
| SEGAHVR | Bacteria | Bacteria |  |  |  |  |  |  |  |  |  |  |  |  |  |  |  |  |  |  |  |  |  |  |  |  |  |  | Bacteria;;;;;;;;;;;Bacteria | [K01869](https://www.genome.jp/entry/K01869) | 6.1.1.4 (97%);2.10.1.1 (2%);6.3.2.10 (2%) | Leucine--tRNA ligase (97%);Molybdopterin molybdotransferase (2%);UDP-N-acetylmuramoyl-tripeptide--D-alanyl-D-alanine ligase (2%) | GO:0005737 (94%) | cytoplasm (94%) | GO:0005524 (97%);GO:0004823 (95%);GO:0002161 (87%) | ATP binding (97%);leucine-tRNA ligase activity (95%);aminoacyl-tRNA editing activity (87%) | GO:0006429 (95%);GO:0006777 (2%);GO:0007049 (2%) | leucyl-tRNA aminoacylation (95%);Mo-molybdopterin cofactor biosynthetic process (2%);cell cycle (2%) | IPR002302 (92%);IPR009008 (92%);IPR009080 (92%) | Leucine-tRNA ligase (92%);Valyl/Leucyl/Isoleucyl-tRNA synthetase | editing domain (92%);Aminoacyl-tRNA synthetase | class Ia | anticodon-binding (92%) | 100.8 | 1 |
| SGADDPEVMEVGGDELSAYDNLNPR | Mycobacteriaceae | Bacteria |  |  |  | Actinobacteria |  |  | Actinobacteria |  |  |  | Corynebacteriales |  |  |  |  | Mycobacteriaceae |  |  |  |  |  |  |  |  |  |  | Bacteria;Actinobacteria;;;Actinobacteria;Corynebacteriales;Mycobacteriaceae;;;;;Mycobacteriaceae | [K01875](https://www.genome.jp/entry/K01875) | 6.1.1.3 | Threonine--tRNA ligase (100%) | GO:0005737 (100%) | cytoplasm (100%) | GO:0004829 (100%);GO:0005524 (100%);GO:0000049 (96%) | threonine-tRNA ligase activity (100%);ATP binding (100%);tRNA binding (96%) | GO:0006435 (100%) | threonyl-tRNA aminoacylation (100%) | IPR002320 (100%);IPR012947 (100%);IPR018163 (100%) | Threonine-tRNA ligase | class IIa (100%);Threonyl/alanyl tRNA synthetase | SAD (100%);Threonyl/alanyl tRNA synthetase | class II-like | 100.6 | 1 |
| SGLFGLCGDNDMR | Streptomyces | Bacteria |  |  |  | Actinobacteria |  |  | Actinobacteria |  |  |  | Streptomycetales |  |  |  |  | Streptomycetaceae |  |  |  | Streptomyces |  |  |  |  |  |  | Bacteria;Actinobacteria;;;Actinobacteria;Streptomycetales;Streptomycetaceae;;Streptomyces;;;Streptomyces | K01758 | 2.7.2.1 | Acetate kinase (100%) | GO:0005737 (100%) | cytoplasm (100%) | GO:0000287 (100%);GO:0005524 (100%);GO:0008776 (100%) | magnesium ion binding (100%);ATP binding (100%);acetate kinase activity (100%) | GO:0006085 (100%);GO:0006082 (92%) | acetyl-CoA biosynthetic process (100%);organic acid metabolic process (92%) | IPR000890 (100%);IPR004372 (100%);IPR023865 (100%) | Aliphatic acid kinase | short-chain (100%);Acetate/propionate kinase (100%);Aliphatic acid kinase | short-chain | conserved site (100%) | 100.3333 | 1 |
| SMQCESDLSLGR | Enterobacteriaceae | Bacteria |  |  |  | Proteobacteria |  |  | Gammaproteobacteria |  |  |  | Enterobacterales |  |  |  |  | Enterobacteriaceae |  |  |  |  |  |  |  |  |  |  | Bacteria;Proteobacteria;;;Gammaproteobacteria;Enterobacterales;Enterobacteriaceae;;;;;Enterobacteriaceae |  |  |  |  |  |  |  | GO:0051607 (100%) | defense response to virus (100%) | IPR014976 (100%);IPR025382 (100%) | Protein of unknown function DUF1837 (100%);Domain of unknown function DUF4297 (100%) |  |  |  | 1 | 1 |
| SQNNGSLALAHNGNLVNATQLK | Bacteria | Bacteria |  |  |  |  |  |  |  |  |  |  |  |  |  |  |  |  |  |  |  |  |  |  |  |  |  |  | Bacteria;;;;;;;;;;;Bacteria | [K00764](https://www.genome.jp/entry/K00764) | 2.4.2.14 | Amidophosphoribosyltransferase (100%) |  |  | GO:0004044 (96%);GO:0000287 (94%);GO:0051539 (94%) | amidophosphoribosyltransferase activity (96%);magnesium ion binding (94%);4 iron | 4 sulfur cluster binding (94%) | GO:0006189 (94%);GO:0006541 (94%);GO:0009113 (87%) | 'de novo' IMP biosynthetic process (94%);glutamine metabolic process (94%);purine nucleobase biosynthetic process (87%) | IPR017932 (100%);IPR029055 (100%);IPR000836 (99%) | Glutamine amidotransferase type 2 domain (100%);Nucleophile aminohydrolases | N-terminal (100%);Phosphoribosyltransferase domain (99%) |  | 1 | 1 |
| SVADHAR | Bacteria | Bacteria |  |  |  |  |  |  |  |  |  |  |  |  |  |  |  |  |  |  |  |  |  |  |  |  |  |  | Bacteria;;;;;;;;;;;Bacteria | [K01089](https://www.genome.jp/entry/K01089) | 4.2.1.19 (75%);3.1.1.61 (17%);2.10.1.1 (8%) | Imidazoleglycerol-phosphate dehydratase (75%);Protein-glutamate methylesterase (17%);Molybdopterin molybdotransferase (8%) | GO:0005737 (58%);GO:0016021 (5%) | cytoplasm (58%);integral component of membrane (5%) | GO:0004424 (47%);GO:0003677 (21%);GO:0000156 (11%) | imidazoleglycerol-phosphate dehydratase activity (47%);DNA binding (21%);phosphorelay response regulator activity (11%) | GO:0000105 (47%);GO:0000160 (11%);GO:0005975 (11%) | histidine biosynthetic process (47%);phosphorelay signal transduction system (11%);carbohydrate metabolic process (11%) | IPR000807 (39%);IPR020565 (39%);IPR020568 (39%) | Imidazoleglycerol-phosphate dehydratase (39%);Imidazoleglycerol-phosphate dehydratase | conserved site (39%);Ribosomal protein S5 domain 2-type fold (39%) |  |  | 1 | 1 |
| TASFTAFEAEVTEEK | Bacillus subtilis | Bacteria |  |  |  | Firmicutes |  |  | Bacilli |  |  |  | Bacillales |  |  |  |  | Bacillaceae |  |  |  | Bacillus |  | Bacillus subtilis group |  | Bacillus subtilis |  |  | Bacteria;Firmicutes;;;Bacilli;Bacillales;Bacillaceae;;Bacillus;Bacillus subtilis group;Bacillus subtilis;Bacillus subtilis |  |  |  | GO:0016021 (100%);GO:0005886 (7%) | integral component of membrane (100%);plasma membrane (7%) |  |  |  |  | IPR010288 (100%) | Bacterial ABC transporter EcsB (100%) |  |  |  | 100.6 | 1 |
| TGLGVDVLLDK | Streptomyces | Bacteria |  |  |  | Actinobacteria |  |  | Actinobacteria |  |  |  | Streptomycetales |  |  |  |  | Streptomycetaceae |  |  |  | Streptomyces |  |  |  |  |  |  | Bacteria;Actinobacteria;;;Actinobacteria;Streptomycetales;Streptomycetaceae;;Streptomyces;;;Streptomyces | [K01925](https://www.genome.jp/entry/K01925) | 3.6.5.0 | -100% | GO:0005886 (100%) | plasma membrane (100%) | GO:0003746 (100%);GO:0003924 (100%);GO:0005525 (100%) | translation elongation factor activity (100%);GTPase activity (100%);GTP binding (100%) | GO:0045727 (100%) | positive regulation of translation (100%) | IPR000640 (100%);IPR000795 (100%);IPR004161 (100%) | Elongation factor EFG | domain V-like (100%);Transcription factor | GTP-binding domain (100%);Translation elongation factor EFTu-like | domain 2 (100%) | 1 | 1 |
| TMNYFNR | Bacteria | Bacteria |  |  |  |  |  |  |  |  |  |  |  |  |  |  |  |  |  |  |  |  |  |  |  |  |  |  | Bacteria;;;;;;;;;;;Bacteria |  |  |  |  |  |  |  |  |  |  |  |  |  |  | 100.8 | 1 |
| TNEASVPYGFK | Bacillus | Bacteria |  |  |  | Firmicutes |  |  | Bacilli |  |  |  | Bacillales |  |  |  |  | Bacillaceae |  |  |  | Bacillus |  |  |  |  |  |  | Bacteria;Firmicutes;;;Bacilli;Bacillales;Bacillaceae;;Bacillus;;;Bacillus |  |  |  | GO:0016021 (100%);GO:0005886 (6%) | integral component of membrane (100%);plasma membrane (6%) |  |  |  |  | IPR018580 (100%) | Uncharacterised protein family YfhO (100%) |  |  |  | 1 | 1 |
| TPAEYLAEK | Mycoplasma pneumoniae | Bacteria |  |  |  | Tenericutes |  |  | Mollicutes |  |  |  | Mycoplasmatales |  |  |  |  | Mycoplasmataceae |  |  |  | Mycoplasma |  |  |  | Mycoplasma pneumoniae |  |  | Bacteria;Tenericutes;;;Mollicutes;Mycoplasmatales;Mycoplasmataceae;;Mycoplasma;;Mycoplasma pneumoniae;Mycoplasma pneumoniae |  |  |  | GO:0005886 (100%) | plasma membrane (100%) |  |  |  |  | IPR004890 (100%);IPR004984 (100%) | Mycoplasma lipoprotein | C-terminal (100%);Mycoplasma lipoprotein | central domain (100%) |  | 100.8667 | 1 |
| TQDGHVNPVR | Mycobacterium tuberculosis complex | Bacteria |  |  |  | Actinobacteria |  |  | Actinobacteria |  |  |  | Corynebacteriales |  |  |  |  | Mycobacteriaceae |  |  |  | Mycobacterium |  | Mycobacterium tuberculosis complex |  |  |  |  | Bacteria;Actinobacteria;;;Actinobacteria;Corynebacteriales;Mycobacteriaceae;;Mycobacterium;Mycobacterium tuberculosis complex;;Mycobacterium tuberculosis complex |  |  |  | GO:0005618 (100%);GO:0005886 (100%) | cell wall (100%);plasma membrane (100%) | GO:0004222 (100%) | metalloendopeptidase activity (100%) |  |  | IPR011055 (100%);IPR016047 (94%);IPR029044 (94%) | Duplicated hybrid motif (100%);Peptidase M23 (94%);Nucleotide-diphospho-sugar transferases (94%) |  |  |  | 100.6733 | 1 |
| TTDCYNLADNVPASEFMNFEGR | Chlorobium chlorochromatii | Bacteria |  |  |  | Chlorobi |  |  | Chlorobia |  |  |  | Chlorobiales |  |  |  |  | Chlorobiaceae |  |  |  | Chlorobium |  |  |  | Chlorobium chlorochromatii |  |  | Bacteria;Chlorobi;;;Chlorobia;Chlorobiales;Chlorobiaceae;;Chlorobium;;Chlorobium chlorochromatii;Chlorobium chlorochromatii | [K03149](https://www.genome.jp/entry/K03149) | 6.1.1.10 | Methionine--tRNA ligase (100%) | GO:0005737 (100%) | cytoplasm (100%) | GO:0000049 (100%);GO:0004825 (100%);GO:0005524 (100%) | tRNA binding (100%);methionine-tRNA ligase activity (100%);ATP binding (100%) | GO:0006431 (100%) | methionyl-tRNA aminoacylation (100%) | IPR001412 (100%);IPR002547 (100%);IPR004495 (100%) | Aminoacyl-tRNA synthetase | class I | conserved site (100%);tRNA-binding domain (100%);Methionyl-tRNA synthetase | beta subunit | 1 | 1 |
| VAQAVNLVLPHVK | Bacillus | Bacteria |  |  |  | Firmicutes |  |  | Bacilli |  |  |  | Bacillales |  |  |  |  | Bacillaceae |  |  |  | Bacillus |  |  |  |  |  |  | Bacteria;Firmicutes;;;Bacilli;Bacillales;Bacillaceae;;Bacillus;;;Bacillus | [K02319](https://www.genome.jp/entry/K02319) | 4.2.1.126 | N-acetylmuramic acid 6-phosphate etherase (100%) |  |  | GO:0016835 (100%);GO:0097367 (100%) | carbon-oxygen lyase activity (100%);carbohydrate derivative binding (100%) | GO:0005975 (100%);GO:0097173 (100%) | carbohydrate metabolic process (100%);N-acetylmuramic acid catabolic process (100%) | IPR001347 (100%);IPR005486 (100%);IPR005488 (100%) | Sugar isomerase (SIS) (100%);Glucokinase regulatory protein | conserved site (100%);N-acetylmuramic acid 6-phosphate etherase MurQ (100%) |  |  | 1 | 1 |
| VGAGEWPVDDNPLR | Bacteria | Bacteria |  |  |  |  |  |  |  |  |  |  |  |  |  |  |  |  |  |  |  |  |  |  |  |  |  |  | Bacteria;;;;;;;;;;;Bacteria | [K00281](https://www.genome.jp/dbget-bin/www_bget?K00281+K00282+K00283+1.4.4.2+R03425) | 1.4.4.2 | Glycine dehydrogenase (aminomethyl-transferring) (100%) | GO:0005618 (1%);GO:0005829 (1%);GO:0005886 (1%) | cell wall (1%);cytosol (1%);plasma membrane (1%) | GO:0004375 (100%);GO:0016829 (5%);GO:0016594 (1%) | glycine dehydrogenase (decarboxylating) activity (100%);lyase activity (5%);glycine binding (1%) | GO:0019464 (98%);GO:0006546 (2%) | glycine decarboxylation via glycine cleavage system (98%);glycine catabolic process (2%) | IPR015421 (100%);IPR015422 (100%);IPR015424 (100%) | Pyridoxal phosphate-dependent transferase | major domain (100%);Pyridoxal phosphate-dependent transferase domain 1 (100%);Pyridoxal phosphate-dependent transferase (100%) |  |  | 100.4733 | 1 |
| VGAGPFPTELFDEDGEALR | Streptomycetaceae | Bacteria |  |  |  | Actinobacteria |  |  | Actinobacteria |  |  |  | Streptomycetales |  |  |  |  | Streptomycetaceae |  |  |  |  |  |  |  |  |  |  | Bacteria;Actinobacteria;;;Actinobacteria;Streptomycetales;Streptomycetaceae;;;;;Streptomycetaceae | K00372 | 6.3.4.4 | Adenylosuccinate synthase (100%) | GO:0005737 (98%) | cytoplasm (98%) | GO:0004019 (100%);GO:0005525 (99%);GO:0000287 (98%) | adenylosuccinate synthase activity (100%);GTP binding (99%);magnesium ion binding (98%) | GO:0044208 (98%);GO:0006164 (0%);GO:0046040 (0%) | 'de novo' AMP biosynthetic process (98%);purine nucleotide biosynthetic process (0%);IMP metabolic process (0%) | IPR001114 (100%);IPR027417 (100%);IPR042111 (100%) | Adenylosuccinate synthetase (100%);P-loop containing nucleoside triphosphate hydrolase (100%);Adenylosuccinate synthetase | domain 3 (100%) |  |  | 1 | 1 |
| VGALSSPLR | Cyanobacteria | Bacteria |  |  |  | Cyanobacteria |  |  |  |  |  |  |  |  |  |  |  |  |  |  |  |  |  |  |  |  |  |  | Bacteria;Cyanobacteria;;;;;;;;;;Cyanobacteria | [K02209](https://www.genome.jp/entry/K02209) | 3.6.4.12 | DNA helicase (100%) |  |  | GO:0003677 (100%);GO:0005524 (100%);GO:0009378 (100%) | DNA binding (100%);ATP binding (100%);four-way junction helicase activity (100%) | GO:0006281 (100%);GO:0006310 (100%);GO:0009432 (100%) | DNA repair (100%);DNA recombination (100%);SOS response (100%) | IPR003593 (100%);IPR004605 (100%);IPR008823 (100%) | AAA+ ATPase domain (100%);DNA helicase | Holliday junction RuvB-type (100%);RuvB C-terminal winged helix domain (100%) |  |  | 100.6067 | 1 |
| VGEVLDVLVLR | Bacteria | Bacteria |  |  |  |  |  |  |  |  |  |  |  |  |  |  |  |  |  |  |  |  |  |  |  |  |  |  | Bacteria;;;;;;;;;;;Bacteria |  |  |  | GO:0005840 (90%);GO:0022627 (1%) | ribosome (90%);cytosolic small ribosomal subunit (1%) | GO:0003676 (99%);GO:0003729 (1%);GO:0003735 (1%) | nucleic acid binding (99%);mRNA binding (1%);structural constituent of ribosome (1%) | GO:0006412 (1%) | translation (1%) | IPR003029 (100%);IPR012340 (100%);IPR022967 (100%) | S1 domain (100%);Nucleic acid-binding | OB-fold (100%);RNA-binding domain | S1 (100%) |  | 1 | 1 |
| VGLHDDGDGFFSGLVPLK | Streptomyces | Bacteria |  |  |  | Actinobacteria |  |  | Actinobacteria |  |  |  | Streptomycetales |  |  |  |  | Streptomycetaceae |  |  |  | Streptomyces |  |  |  |  |  |  | Bacteria;Actinobacteria;;;Actinobacteria;Streptomycetales;Streptomycetaceae;;Streptomyces;;;Streptomyces | [K00700](https://www.genome.jp/dbget-bin/www_bget?K00700+K16149+2.4.1.18+R02110) | 2.4.1.18 | 1 | 4-alpha-glucan branching enzyme (100%) | GO:0005737 (17%);GO:0005829 (17%) | cytoplasm (17%);cytosol (17%) | GO:0003844 (100%);GO:0004553 (100%);GO:0043169 (100%) | 1 | 4-alpha-glucan branching enzyme activity (100%);hydrolase activity | hydrolyzing O-glycosyl compounds (100%);cation binding (100%) | GO:0005978 (100%);GO:0005975 (17%) | glycogen biosynthetic process (100%);carbohydrate metabolic process (17%) | IPR004193 (100%);IPR006047 (100%);IPR006048 (100%) | Glycoside hydrolase | 1 | 1 |
| VNEAEFEAK | Campylobacter fetus | Bacteria |  |  |  | Proteobacteria | delta/epsilon subdivisions |  | Epsilonproteobacteria |  |  |  | Campylobacterales |  |  |  |  | Campylobacteraceae |  |  |  | Campylobacter |  |  |  | Campylobacter fetus |  |  | Bacteria;Proteobacteria;delta/epsilon subdivisions;;Epsilonproteobacteria;Campylobacterales;Campylobacteraceae;;Campylobacter;;Campylobacter fetus;Campylobacter fetus |  | 3.1.-.- | Acting on ester bonds (100%) | GO:0005886 (100%);GO:0016021 (14%) | plasma membrane (100%);integral component of membrane (14%) | GO:0003723 (100%);GO:0004521 (100%) | RNA binding (100%);endoribonuclease activity (100%) | GO:0006402 (100%) | mRNA catabolic process (100%) | IPR003607 (100%);IPR006674 (100%);IPR006675 (100%) | HD/PDEase domain (100%);HD domain (100%);HDIG domain (100%) |  |  |  | 100.6667 | 1 |
| VSCHSCNGFGAEGGESGLDLCK | Mycoplasma genitalium | Bacteria |  |  |  | Tenericutes |  |  | Mollicutes |  |  |  | Mycoplasmatales |  |  |  |  | Mycoplasmataceae |  |  |  | Mycoplasma |  |  |  | Mycoplasma genitalium |  |  | Bacteria;Tenericutes;;;Mollicutes;Mycoplasmatales;Mycoplasmataceae;;Mycoplasma;;Mycoplasma genitalium;Mycoplasma genitalium |  |  |  | GO:0005737 (100%) | cytoplasm (100%) | GO:0005524 (100%);GO:0008270 (100%);GO:0031072 (100%) | ATP binding (100%);zinc ion binding (100%);heat shock protein binding (100%) | GO:0006260 (100%);GO:0009408 (100%);GO:0042026 (100%) | DNA replication (100%);response to heat (100%);protein refolding (100%) | IPR001305 (100%);IPR001623 (100%);IPR002939 (100%) | Heat shock protein DnaJ | cysteine-rich domain (100%);DnaJ domain (100%);Chaperone DnaJ | C-terminal (100%) |  | 100.5333 | 1 |
| VTAYAPK | Bacteria | Bacteria |  |  |  |  |  |  |  |  |  |  |  |  |  |  |  |  |  |  |  |  |  |  |  |  |  |  | Bacteria;;;;;;;;;;;Bacteria |  |  |  | GO:0005739 (4%) | mitochondrion (4%) | GO:0005525 (100%) | GTP binding (100%) | GO:0032297 (4%) | negative regulation of DNA-dependent DNA replication initiation (4%) | IPR006073 (100%);IPR019988 (100%);IPR027417 (100%) | GTP binding domain (100%);GTP-binding protein | riobosome biogenesis | YqeH (100%);P-loop containing nucleoside triphosphate hydrolase (100%) |  | 100.5333 | 1 |
| VYVTFQAK | Mycoplasma mobile | Bacteria |  |  |  | Tenericutes |  |  | Mollicutes |  |  |  | Mycoplasmatales |  |  |  |  | Mycoplasmataceae |  |  |  | Mycoplasma |  |  |  | Mycoplasma mobile |  |  | Bacteria;Tenericutes;;;Mollicutes;Mycoplasmatales;Mycoplasmataceae;;Mycoplasma;;Mycoplasma mobile;Mycoplasma mobile |  |  |  | GO:0005737 (100%);GO:0005886 (100%) | cytoplasm (100%);plasma membrane (100%) | GO:0005524 (100%) | ATP binding (100%) | GO:0006605 (100%);GO:0017038 (100%);GO:0065002 (100%) | protein targeting (100%);protein import (100%);intracellular protein transmembrane transport (100%) | IPR000185 (100%);IPR011115 (100%);IPR011116 (100%) | Protein translocase subunit SecA (100%);SecA DEAD-like | N-terminal (100%);SecA Wing/Scaffold (100%) |  |  | 1 | 1 |
| WTPTLSFLR | Candidatus Protochlamydia amoebophila | Bacteria |  |  |  | Chlamydiae |  |  | Chlamydiia |  |  |  | Parachlamydiales |  |  |  |  | Parachlamydiaceae |  |  |  | Candidatus Protochlamydia |  |  |  | Candidatus Protochlamydia amoebophila |  |  | Bacteria;Chlamydiae;;;Chlamydiia;Parachlamydiales;Parachlamydiaceae;;Candidatus Protochlamydia;;Candidatus Protochlamydia amoebophila;Candidatus Protochlamydia amoebophila | [K13292](https://www.genome.jp/dbget-bin/www_bget?ko:K13292) | 2.5.1.145 | Phosphatidylglycerol--prolipoprotein diacylglyceryl transferase (100%) | GO:0005887 (100%) | integral component of plasma membrane (100%) | GO:0008961 (100%) | phosphatidylglycerol-prolipoprotein diacylglyceryl transferase activity (100%) | GO:0042158 (100%) | lipoprotein biosynthetic process (100%) | IPR001640 (100%) | Phosphatidylglycerol--prolipoprotein diacylglyceryl transferase Lgt (100%) |  |  |  | 100.8 | 1 |
| YHATGETLTDADLDALK | Streptomyces | Bacteria |  |  |  | Actinobacteria |  |  | Actinobacteria |  |  |  | Streptomycetales |  |  |  |  | Streptomycetaceae |  |  |  | Streptomyces |  |  |  |  |  |  | Bacteria;Actinobacteria;;;Actinobacteria;Streptomycetales;Streptomycetaceae;;Streptomyces;;;Streptomyces | [K00052](https://www.genome.jp/entry/K00052) | 1.1.1.85 | 3-isopropylmalate dehydrogenase (100%) | GO:0005737 (98%) | cytoplasm (98%) | GO:0003862 (100%);GO:0000287 (94%);GO:0051287 (94%) | 3-isopropylmalate dehydrogenase activity (100%);magnesium ion binding (94%);NAD binding (94%) | GO:0009098 (98%) | leucine biosynthetic process (98%) | IPR019818 (100%);IPR023698 (100%);IPR024084 (100%) | Isocitrate/isopropylmalate dehydrogenase | conserved site (100%);3-isopropylmalate dehydrogenase (100%);Isopropylmalate dehydrogenase-like domain (100%) |  |  | 100.74 | 1 |
| YLTVCCLLLTAVFSWAER | Bacteroidetes | Bacteria |  |  |  | Bacteroidetes |  |  |  |  |  |  |  |  |  |  |  |  |  |  |  |  |  |  |  |  |  |  | Bacteria;Bacteroidetes;;;;;;;;;;Bacteroidetes |  |  |  | GO:0016021 (100%) | integral component of membrane (100%) | GO:0004519 (100%);GO:0004527 (100%) | endonuclease activity (100%);exonuclease activity (100%) |  |  | IPR036691 (100%) | Endonuclease/exonuclease/phosphatase superfamily (100%) |  |  |  | 100.9333 | 1 |
| YNLYLGGNK | Gammaproteobacteria | Bacteria |  |  |  | Proteobacteria |  |  | Gammaproteobacteria |  |  |  |  |  |  |  |  |  |  |  |  |  |  |  |  |  |  |  | Bacteria;Proteobacteria;;;Gammaproteobacteria;;;;;;;Gammaproteobacteria | K14085 | 1.8.1.2 | Assimilatory sulfite reductase (NADPH) (100%) | GO:0009337 (92%) | sulfite reductase complex (NADPH) (92%) | GO:0004783 (100%);GO:0020037 (92%);GO:0046872 (92%) | sulfite reductase (NADPH) activity (100%);heme binding (92%);metal ion binding (92%) | GO:0019344 (92%);GO:0070814 (92%);GO:0000103 (83%) | cysteine biosynthetic process (92%);hydrogen sulfide biosynthetic process (92%);sulfate assimilation (83%) | IPR005117 (100%);IPR006066 (100%);IPR006067 (100%) | Nitrite/Sulfite reductase ferredoxin-like domain (100%);Nitrite/sulphite reductase iron-sulphur/sirohaem-binding site (100%);Nitrite/sulphite reductase 4Fe-4S domain (100%) |  |  |  | 100.66 | 1 |
| AADAAQEGGDEFHWHR | Enterobacterales | Bacteria |  |  |  | Proteobacteria |  |  | Gammaproteobacteria |  |  |  | Enterobacterales |  |  |  |  |  |  |  |  |  |  |  |  |  |  |  | Bacteria;Proteobacteria;;;Gammaproteobacteria;Enterobacterales;;;;;;Enterobacterales |  |  |  | GO:0005737 (100%);GO:0009295 (100%) | cytoplasm (100%);nucleoid (100%) | GO:0005509 (94%) | calcium ion binding (94%) | GO:0007049 (100%);GO:0007059 (100%);GO:0030261 (100%) | cell cycle (100%);chromosome segregation (100%);chromosome condensation (100%) | IPR033439 (100%);IPR033440 (100%);IPR033441 (100%) | Chromosome partition protein MukF | winged-helix domain (100%);Chromosome partition protein MukF |  |  | 1 | 100.6923 |
| AAEAEFGTVLR | Streptomyces | Bacteria |  |  |  | Actinobacteria |  |  | Actinobacteria |  |  |  | Streptomycetales |  |  |  |  | Streptomycetaceae |  |  |  | Streptomyces |  |  |  |  |  |  | Bacteria;Actinobacteria;;;Actinobacteria;Streptomycetales;Streptomycetaceae;;Streptomyces;;;Streptomyces | K01488 | 3.5.4.4 (74%);3.5.4.- (13%);3.5.4.40 (10%) | Adenosine deaminase (74%);In cyclic amidines (13%);Aminodeoxyfutalosine deaminase (10%) |  |  | GO:0046872 (98%);GO:0019239 (79%);GO:0004000 (15%) | metal ion binding (98%);deaminase activity (79%);adenosine deaminase activity (15%) | GO:0006146 (1%);GO:0009234 (1%);GO:0043103 (1%) | adenine catabolic process (1%);menaquinone biosynthetic process (1%);hypoxanthine salvage (1%) | IPR001365 (100%);IPR006330 (100%);IPR032466 (100%) | Adenosine/AMP deaminase domain (100%);Adenosine/adenine deaminase (100%);Metal-dependent hydrolase (100%) |  |  |  | 1 | 100.4615 |
| AAFVDPAK | Bacteria | Bacteria |  |  |  |  |  |  |  |  |  |  |  |  |  |  |  |  |  |  |  |  |  |  |  |  |  |  | Bacteria;;;;;;;;;;;Bacteria | [K01200](https://www.genome.jp/dbget-bin/www_bget?ko:K01200) | 3.2.1.41 (93%);3.2.1.1 (7%);3.2.1.- (1%) | Pullulanase (93%);Alpha-amylase (7%);Glycosidases | i.e. enzymes hydrolyzing O- and S-glycosyl compounds (1%) | GO:0005576 (97%);GO:0005618 (94%);GO:0016021 (88%) | extracellular region (97%);cell wall (94%);integral component of membrane (88%) | GO:0030246 (97%);GO:0004553 (51%);GO:0051060 (44%) | carbohydrate binding (97%);hydrolase activity | hydrolyzing O-glycosyl compounds (51%);pullulanase activity (44%) | GO:0005975 (97%);GO:0008152 (1%) | carbohydrate metabolic process (97%);metabolic process (1%) | IPR013783 (100%);IPR004193 (100%);IPR017853 (100%) |  |  | 1 | 100.8462 |
| AALSGHFGSASAAEEALR | Corynebacterium efficiens | Bacteria |  |  |  | Actinobacteria |  |  | Actinobacteria |  |  |  | Corynebacteriales |  |  |  |  | Corynebacteriaceae |  |  |  | Corynebacterium |  |  |  | Corynebacterium efficiens |  |  | Bacteria;Actinobacteria;;;Actinobacteria;Corynebacteriales;Corynebacteriaceae;;Corynebacterium;;Corynebacterium efficiens;Corynebacterium efficiens | [K02528](https://www.genome.jp/entry/K02528) | 2.1.1.182 | 16S rRNA (adenine(1518)-N(6)/adenine(1519)-N(6))-dimethyltransferase (100%) | GO:0005737 (100%) | cytoplasm (100%) | GO:0000179 (100%);GO:0003723 (100%);GO:0052908 (100%) | rRNA (adenine-N6 | N6-)-dimethyltransferase activity (100%);RNA binding (100%);16S rRNA (adenine(1518)-N(6)/adenine(1519)-N(6))-dimethyltransferase activity (100%) |  |  | IPR001737 (100%);IPR011530 (100%);IPR020596 (100%) | Ribosomal RNA adenine methyltransferase KsgA/Erm (100%);Ribosomal RNA adenine dimethylase (100%);Ribosomal RNA adenine methylase transferase |  |  | 1 | 100.6077 |
| AAWASVAR | Bacteria | Bacteria |  |  |  |  |  |  |  |  |  |  |  |  |  |  |  |  |  |  |  |  |  |  |  |  |  |  | Bacteria;;;;;;;;;;;Bacteria | [K10536](https://www.genome.jp/dbget-bin/www_bget?K10536+3.5.3.12+R01416) | 3.5.3.12 | Agmatine deiminase (100%) |  |  | GO:0004668 (93%);GO:0047632 (74%);GO:0009055 (4%) | protein-arginine deiminase activity (93%);agmatine deiminase activity (74%);electron transfer activity (4%) | GO:0009446 (93%) | putrescine biosynthetic process (93%) | IPR007466 (93%);IPR017754 (71%);IPR022655 (6%) | Peptidyl-arginine deiminase | Porphyromonas-type (93%);Agmatine deiminase (71%);Domain of unknown function DUF1553 (6%) |  |  | 1 | 100.6923 |
| ADATVQFGYESGYDR | Mycobacteriaceae | Bacteria |  |  |  | Actinobacteria |  |  | Actinobacteria |  |  |  | Corynebacteriales |  |  |  |  | Mycobacteriaceae |  |  |  |  |  |  |  |  |  |  | Bacteria;Actinobacteria;;;Actinobacteria;Corynebacteriales;Mycobacteriaceae;;;;;Mycobacteriaceae | [K15526](https://www.genome.jp/entry/K15526) | 6.3.1.13 | L-cysteine:1D-myo-inositol 2-amino-2-deoxy-alpha-D-glucopyranosidligase (100%) |  |  | GO:0005524 (100%);GO:0035446 (100%);GO:0008270 (97%) | ATP binding (100%);cysteine-glucosaminylinositol ligase activity (100%);zinc ion binding (97%) | GO:0010125 (100%);GO:0006418 (6%) | mycothiol biosynthetic process (100%);tRNA aminoacylation for protein translation (6%) | IPR014729 (100%);IPR017812 (100%);IPR024909 (100%) | Rossmann-like alpha/beta/alpha sandwich fold (100%);Mycothiol ligase MshC (100%);Cysteinyl-tRNA synthetase/mycothiol ligase (100%) |  |  |  | 1 | 1 |
| ADLTQSAGLVR | Neisseria | Bacteria |  |  |  | Proteobacteria |  |  | Betaproteobacteria |  |  |  | Neisseriales |  |  |  |  | Neisseriaceae |  |  |  | Neisseria |  |  |  |  |  |  | Bacteria;Proteobacteria;;;Betaproteobacteria;Neisseriales;Neisseriaceae;;Neisseria;;;Neisseria | [K06173](https://www.genome.jp/entry/K06173) | 5.4.99.12 | tRNA pseudouridine(38-40) synthase (100%) |  |  | GO:0003723 (100%);GO:0106029 (100%);GO:0016829 (20%) | RNA binding (100%);tRNA pseudouridine synthase activity (100%);lyase activity (20%) | GO:0031119 (100%) | tRNA pseudouridine synthesis (100%) | IPR001406 (100%);IPR020095 (100%);IPR020097 (100%) | Pseudouridine synthase I | TruA (100%);Pseudouridine synthase I |  |  | 1 | 100.6077 |
| ADVEAALANNDLAELK | Bacilli | Bacteria |  |  |  | Firmicutes |  |  | Bacilli |  |  |  |  |  |  |  |  |  |  |  |  |  |  |  |  |  |  |  | Bacteria;Firmicutes;;;Bacilli;;;;;;;Bacilli | K00105 | 1.1.3.21 (95%);6.1.1.13 (4%);1.1.5.3 (2%) | Glycerol-3-phosphate oxidase (95%);D-alanine--poly(phosphoribitol) ligase (4%);Glycerol-3-phosphate dehydrogenase (2%) | GO:0009331 (98%);GO:0016021 (90%);GO:0005886 (1%) | glycerol-3-phosphate dehydrogenase complex (98%);integral component of membrane (90%);plasma membrane (1%) | GO:0004368 (97%);GO:0004369 (53%);GO:0016874 (2%) | glycerol-3-phosphate dehydrogenase (quinone) activity (97%);glycerol-3-phosphate oxidase activity (53%);ligase activity (2%) | GO:0006072 (98%);GO:0006071 (1%);GO:0006650 (1%) | glycerol-3-phosphate metabolic process (98%);glycerol metabolic process (1%);glycerophospholipid metabolic process (1%) | IPR031656 (100%);IPR038299 (100%);IPR000447 (99%) | Alpha-glycerophosphate oxidase | C-terminal (100%);Alpha-glycerophosphate oxidase |  |  | 1 | 100.4538 |
| AEGKPEAALPK | Bacteria | Bacteria |  |  |  |  |  |  |  |  |  |  |  |  |  |  |  |  |  |  |  |  |  |  |  |  |  |  | Bacteria;;;;;;;;;;;Bacteria |  |  |  | GO:0005737 (98%) | cytoplasm (98%) | GO:0003746 (100%) | translation elongation factor activity (100%) | GO:0006414 (0%) | translational elongation (0%) | IPR001816 (100%);IPR014039 (100%);IPR036402 (100%) | Translation elongation factor EFTs/EF1B (100%);Translation elongation factor EFTs/EF1B | dimerisation (100%);Elongation factor Ts |  |  | 1 | 1 |
| AEVEGAVR | Bacteria | Bacteria |  |  |  |  |  |  |  |  |  |  |  |  |  |  |  |  |  |  |  |  |  |  |  |  |  |  | Bacteria;;;;;;;;;;;Bacteria | [K02209](https://www.genome.jp/entry/K02209) | 3.1.-.- (90%);3.6.4.12 (90%);2.3.1.274 (10%) | Acting on ester bonds (90%);DNA helicase (90%);Phosphate acyltransferase (10%) | GO:0005737 (10%) | cytoplasm (10%) | GO:0003678 (90%);GO:0003690 (90%);GO:0005524 (90%) | DNA helicase activity (90%);double-stranded DNA binding (90%);ATP binding (90%) | GO:0000724 (90%);GO:0006633 (10%);GO:0008654 (10%) | double-strand break repair via homologous recombination (90%);fatty acid biosynthetic process (10%);phospholipid biosynthetic process (10%) | IPR000212 (90%);IPR014140 (90%);IPR027417 (90%) | DNA helicase | UvrD/REP type (90%);DNA helicase subunit AddB (90%);P-loop containing nucleoside triphosphate hydrolase (90%) |  |  | 1 | 1 |
| AEYESMQAPAVGGVR | Corynebacteriales | Bacteria |  |  |  | Actinobacteria |  |  | Actinobacteria |  |  |  | Corynebacteriales |  |  |  |  |  |  |  |  |  |  |  |  |  |  |  | Bacteria;Actinobacteria;;;Actinobacteria;Corynebacteriales;;;;;;Corynebacteriales |  |  |  | GO:0005737 (93%) | cytoplasm (93%) | GO:0003743 (100%);GO:0003924 (100%);GO:0005525 (100%) | translation initiation factor activity (100%);GTPase activity (100%);GTP binding (100%) |  |  | IPR000178 (100%);IPR000795 (100%);IPR005225 (100%) | Translation initiation factor aIF-2 | bacterial-like (100%);Transcription factor |  |  | 1 | 100.6923 |
| AGEEAAADGAAPPVAAR | Streptomyces | Bacteria |  |  |  | Actinobacteria |  |  | Actinobacteria |  |  |  | Streptomycetales |  |  |  |  | Streptomycetaceae |  |  |  | Streptomyces |  |  |  |  |  |  | Bacteria;Actinobacteria;;;Actinobacteria;Streptomycetales;Streptomycetaceae;;Streptomyces;;;Streptomyces |  |  |  | GO:0005737 (100%) | cytoplasm (100%) | GO:0003677 (100%);GO:0003700 (100%);GO:0050662 (100%) | DNA binding (100%);DNA-binding transcription factor activity (100%);coenzyme binding (100%) | GO:0045892 (100%);GO:0051775 (100%) | negative regulation of transcription | DNA-templated (100%);response to redox state (100%) | IPR003781 (100%);IPR009718 (100%);IPR022876 (100%) | CoA-binding (100%);Rex DNA-binding |  |  | 1 | 100.3846 |
| AGEPDLWDDPAAAQK | Micrococcaceae | Bacteria |  |  |  | Actinobacteria |  |  | Actinobacteria |  |  |  | Micrococcales |  |  |  |  | Micrococcaceae |  |  |  |  |  |  |  |  |  |  | Bacteria;Actinobacteria;;;Actinobacteria;Micrococcales;Micrococcaceae;;;;;Micrococcaceae |  |  |  | GO:0005737 (94%) | cytoplasm (94%) | GO:0016149 (94%);GO:0003747 (3%) | translation release factor activity | codon specific (94%);translation release factor activity (3%) | GO:0006415 (3%) | translational termination (3%) | IPR005139 (100%);IPR000352 (97%);IPR004374 (94%) | Peptide chain release factor (100%);Peptide chain release factor class I (97%);Peptide chain release factor 2 (94%) |  |  | 1 | 100.7692 |
| AGHGVDLAK | Streptomyces | Bacteria |  |  |  | Actinobacteria |  |  | Actinobacteria |  |  |  | Streptomycetales |  |  |  |  | Streptomycetaceae |  |  |  | Streptomyces |  |  |  |  |  |  | Bacteria;Actinobacteria;;;Actinobacteria;Streptomycetales;Streptomycetaceae;;Streptomyces;;;Streptomyces |  |  |  |  |  | GO:0005524 (100%);GO:0051082 (90%) | ATP binding (100%);unfolded protein binding (90%) | GO:0006457 (90%) | protein folding (90%) | IPR013126 (100%);IPR018181 (100%);IPR029047 (100%) | Heat shock protein 70 family (100%);Heat shock protein 70 | conserved site (100%);Heat shock protein 70kD |  |  | 1 | 100.6923 |
| AGTAFTLSALK | Bacillales | Bacteria |  |  |  | Firmicutes |  |  | Bacilli |  |  |  | Bacillales |  |  |  |  |  |  |  |  |  |  |  |  |  |  |  | Bacteria;Firmicutes;;;Bacilli;Bacillales;;;;;;Bacillales |  | 3.1.-.- | Acting on ester bonds (100%) | GO:0009279 (3%) | cell outer membrane (3%) | GO:0016787 (97%);GO:0052689 (3%) | hydrolase activity (97%);carboxylic ester hydrolase activity (3%) |  |  | IPR008979 (100%);IPR013830 (100%);IPR036514 (100%) | Galactose-binding-like domain superfamily (100%);SGNH hydrolase-type esterase domain (100%);SGNH hydrolase superfamily (100%) |  |  |  | 1 | 100.3923 |
| AGTSGSFNAGVEGGK | Escherichia coli | Bacteria |  |  |  | Proteobacteria |  |  | Gammaproteobacteria |  |  |  | Enterobacterales |  |  |  |  | Enterobacteriaceae |  |  |  | Escherichia |  |  |  | Escherichia coli |  |  | Bacteria;Proteobacteria;;;Gammaproteobacteria;Enterobacterales;Enterobacteriaceae;;Escherichia;;Escherichia coli;Escherichia coli |  | 3.1.-.- | Acting on ester bonds (100%) | GO:0016021 (91%);GO:0005576 (9%) | integral component of membrane (91%);extracellular region (9%) | GO:0004519 (9%);GO:0090729 (9%) | endonuclease activity (9%);toxin activity (9%) | GO:0009405 (9%) | pathogenesis (9%) | IPR006914 (100%);IPR025157 (83%);IPR010069 (78%) | VENN motif-containing domain (100%);Hemagglutinin repeat (83%);Filamentous haemagglutinin | 20-residue x2 repeat (78%) |  |  | 1 | 100.4615 |
| AGVALHELPLK | Bradyrhizobiaceae | Bacteria |  |  |  | Proteobacteria |  |  | Alphaproteobacteria |  |  |  | Rhizobiales |  |  |  |  | Bradyrhizobiaceae |  |  |  |  |  |  |  |  |  |  | Bacteria;Proteobacteria;;;Alphaproteobacteria;Rhizobiales;Bradyrhizobiaceae;;;;;Bradyrhizobiaceae | [K01755](https://www.genome.jp/entry/K01755) | 4.3.2.1 | Argininosuccinate lyase (100%) | GO:0005737 (100%) | cytoplasm (100%) | GO:0004056 (100%) | argininosuccinate lyase activity (100%) | GO:0042450 (80%);GO:0006526 (20%) | arginine biosynthetic process via ornithine (80%);arginine biosynthetic process (20%) | IPR000362 (100%);IPR008948 (100%);IPR009049 (100%) | Fumarate lyase family (100%);L-Aspartase-like (100%);Argininosuccinate lyase (100%) |  |  |  | 1 | 1 |
| AHTALVSLR | Mycobacteriaceae | Bacteria |  |  |  | Actinobacteria |  |  | Actinobacteria |  |  |  | Corynebacteriales |  |  |  |  | Mycobacteriaceae |  |  |  |  |  |  |  |  |  |  | Bacteria;Actinobacteria;;;Actinobacteria;Corynebacteriales;Mycobacteriaceae;;;;;Mycobacteriaceae |  |  |  | GO:0005829 (50%);GO:0005886 (50%) | cytosol (50%);plasma membrane (50%) | GO:0004601 (50%);GO:0020037 (50%) | peroxidase activity (50%);heme binding (50%) | GO:0006979 (50%) | response to oxidative stress (50%) | IPR022812 (100%);IPR027417 (100%);IPR002016 (1%) | Dynamin superfamily (100%);P-loop containing nucleoside triphosphate hydrolase (100%);Haem peroxidase (1%) |  |  |  | 1 | 100.6154 |
| ALENAPLPPK | Thermosynechococcus elongatus | Bacteria |  |  |  | Cyanobacteria |  |  |  |  |  |  | Synechococcales |  |  |  |  | Synechococcaceae |  |  |  | Thermosynechococcus |  |  |  | Thermosynechococcus elongatus |  |  | Bacteria;Cyanobacteria;;;;Synechococcales;Synechococcaceae;;Thermosynechococcus;;Thermosynechococcus elongatus;Thermosynechococcus elongatus | [K01081](https://www.genome.jp/entry/K01081) | 3.1.3.5 | 5'-nucleotidase (100%) | GO:0005737 (100%) | cytoplasm (100%) | GO:0000166 (100%);GO:0008253 (100%);GO:0046872 (100%) | nucleotide binding (100%);5'-nucleotidase activity (100%);metal ion binding (100%) |  |  | IPR002828 (100%);IPR030048 (100%);IPR036523 (100%) | Survival protein SurE-like phosphatase/nucleotidase (100%);Survival protein SurE (100%);SurE-like phosphatase/nucleotidase superfamily (100%) |  |  |  | 1 | 100.6231 |
| ALFDAAADDR | Streptomyces | Bacteria |  |  |  | Actinobacteria |  |  | Actinobacteria |  |  |  | Streptomycetales |  |  |  |  | Streptomycetaceae |  |  |  | Streptomyces |  |  |  |  |  |  | Bacteria;Actinobacteria;;;Actinobacteria;Streptomycetales;Streptomycetaceae;;Streptomyces;;;Streptomyces |  |  |  |  |  | GO:0016793 (90%);GO:0008832 (5%);GO:0016787 (5%) | triphosphoric monoester hydrolase activity (90%);dGTPase activity (5%);hydrolase activity (5%) | GO:0006203 (5%) | dGTP catabolic process (5%) | IPR003607 (100%);IPR006261 (100%);IPR006674 (100%) | HD/PDEase domain (100%);dNTP triphosphohydrolase (100%);HD domain (100%) |  |  |  | 1 | 100.7 |
| ALLGGALEGK | Rhodococcus | Bacteria |  |  |  | Actinobacteria |  |  | Actinobacteria |  |  |  | Corynebacteriales |  |  |  |  | Nocardiaceae |  |  |  | Rhodococcus |  |  |  |  |  |  | Bacteria;Actinobacteria;;;Actinobacteria;Corynebacteriales;Nocardiaceae;;Rhodococcus;;;Rhodococcus |  |  |  |  |  | GO:0003677 (96%);GO:0003700 (19%) | DNA binding (96%);DNA-binding transcription factor activity (19%) | GO:0045892 (96%);GO:0006355 (4%) | negative regulation of transcription | DNA-templated (96%);regulation of transcription | DNA-templated (4%) | IPR002571 (100%);IPR021153 (100%);IPR023120 (100%) |  |  | 1 | 100.9231 |
| ALQAGFDLSNK | Mycoplasma genitalium | Bacteria |  |  |  | Tenericutes |  |  | Mollicutes |  |  |  | Mycoplasmatales |  |  |  |  | Mycoplasmataceae |  |  |  | Mycoplasma |  |  |  | Mycoplasma genitalium |  |  | Bacteria;Tenericutes;;;Mollicutes;Mycoplasmatales;Mycoplasmataceae;;Mycoplasma;;Mycoplasma genitalium;Mycoplasma genitalium |  |  |  | GO:0016021 (100%);GO:0005886 (5%) | integral component of membrane (100%);plasma membrane (5%) |  |  | GO:0020035 (5%) | cytoadherence to microvasculature | mediated by symbiont protein (5%) | IPR004940 (100%);IPR022116 (100%);IPR022400 (100%) | Adhesin P1 domain (100%);Trypsin-sensitive surface-exposed protein (100%);Adhesin P1 (100%) |  |  | 1 | 100.3769 |
| AMMDNLQTETVVNR | Enterobacteriaceae | Bacteria |  |  |  | Proteobacteria |  |  | Gammaproteobacteria |  |  |  | Enterobacterales |  |  |  |  | Enterobacteriaceae |  |  |  |  |  |  |  |  |  |  | Bacteria;Proteobacteria;;;Gammaproteobacteria;Enterobacterales;Enterobacteriaceae;;;;;Enterobacteriaceae | K03010 | 2.7.7.6 | DNA-directed RNA polymerase (100%) |  |  | GO:0003899 (100%);GO:0000287 (90%);GO:0003677 (90%) | DNA-directed 5'-3' RNA polymerase activity (100%);magnesium ion binding (90%);DNA binding (90%) | GO:0006351 (90%) | transcription | DNA-templated (90%) | IPR000722 (100%);IPR006592 (100%);IPR007066 (100%) | RNA polymerase |  |  | 1 | 100.3077 |
| ANNGEVGLK | Mycoplasma gallisepticum | Bacteria |  |  |  | Tenericutes |  |  | Mollicutes |  |  |  | Mycoplasmatales |  |  |  |  | Mycoplasmataceae |  |  |  | Mycoplasma |  |  |  | Mycoplasma gallisepticum |  |  | Bacteria;Tenericutes;;;Mollicutes;Mycoplasmatales;Mycoplasmataceae;;Mycoplasma;;Mycoplasma gallisepticum;Mycoplasma gallisepticum | [K01872](https://www.genome.jp/entry/K01872) | 6.1.1.7 | Alanine--tRNA ligase (100%) | GO:0005737 (100%) | cytoplasm (100%) | GO:0004813 (100%);GO:0005524 (100%);GO:0000049 (75%) | alanine-tRNA ligase activity (100%);ATP binding (100%);tRNA binding (75%) | GO:0006419 (100%) | alanyl-tRNA aminoacylation (100%) | IPR018162 (100%);IPR018164 (100%);IPR018165 (100%) | Alanine-tRNA ligase | class IIc |  |  | 1 | 1 |
| ANPFDAFVSNQYAR | Neisseriaceae | Bacteria |  |  |  | Proteobacteria |  |  | Betaproteobacteria |  |  |  | Neisseriales |  |  |  |  | Neisseriaceae |  |  |  |  |  |  |  |  |  |  | Bacteria;Proteobacteria;;;Betaproteobacteria;Neisseriales;Neisseriaceae;;;;;Neisseriaceae | [K01749](https://www.genome.jp/entry/K01749) | 2.5.1.61 | Hydroxymethylbilane synthase (100%) |  |  | GO:0004418 (100%) | hydroxymethylbilane synthase activity (100%) | GO:0006782 (100%);GO:0018160 (100%) | protoporphyrinogen IX biosynthetic process (100%);peptidyl-pyrromethane cofactor linkage (100%) | IPR000860 (100%);IPR022417 (100%);IPR022418 (100%) | Porphobilinogen deaminase (100%);Porphobilinogen deaminase | N-terminal (100%);Porphobilinogen deaminase |  |  | 1 | 1 |
| ANWALGR | Bacteria | Bacteria |  |  |  |  |  |  |  |  |  |  |  |  |  |  |  |  |  |  |  |  |  |  |  |  |  |  | Bacteria;;;;;;;;;;;Bacteria | [K00861](https://www.genome.jp/entry/K00861) | 2.7.1.26 ;2.7.7.2 | Riboflavin kinase (100%);FAD synthase (100%) | GO:0016021 (2%) | integral component of membrane (2%) | GO:0005524 (98%);GO:0003919 (94%);GO:0008531 (94%) | ATP binding (98%);FMN adenylyltransferase activity (94%);riboflavin kinase activity (94%) | GO:0006747 (94%);GO:0009398 (94%);GO:0009231 (87%) | FAD biosynthetic process (94%);FMN biosynthetic process (94%);riboflavin biosynthetic process (87%) | IPR002606 (94%);IPR014729 (94%);IPR015864 (94%) | Riboflavin kinase | bacterial (94%);Rossmann-like alpha/beta/alpha sandwich fold (94%);FAD synthetase (94%) |  |  | 1 | 100.3077 |
| APAAADVASAADVDADAAAVPEPGEPGPGGAAETP | Clavibacter michiganensis subsp. michiganensis | Bacteria |  |  |  | Actinobacteria |  |  | Actinobacteria |  |  |  | Micrococcales |  |  |  |  | Microbacteriaceae |  |  |  | Clavibacter |  |  |  | Clavibacter michiganensis | Clavibacter michiganensis subsp. michiganensis |  | Bacteria;Actinobacteria;;;Actinobacteria;Micrococcales;Microbacteriaceae;;Clavibacter;;Clavibacter michiganensisClavibacter michiganensis subsp. michiganensis;Clavibacter michiganensis subsp. michiganensis | [K01972](https://www.genome.jp/dbget-bin/www_bget?K01972) | 6.5.1.2 | DNA ligase (NAD(+)) (100%) |  |  | GO:0003911 (100%);GO:0046872 (100%) | DNA ligase (NAD+) activity (100%);metal ion binding (100%) | GO:0006260 (100%);GO:0006281 (100%) | DNA replication (100%);DNA repair (100%) | IPR001357 (100%);IPR001679 (100%);IPR004149 (100%) | BRCT domain (100%);NAD-dependent DNA ligase (100%);Zinc-finger | NAD-dependent DNA ligase C4-type (100%) |  |  | 1 | 100.3769 |
| APSDMSAR | Actinobacteria | Bacteria |  |  |  | Actinobacteria |  |  | Actinobacteria |  |  |  |  |  |  |  |  |  |  |  |  |  |  |  |  |  |  |  | Bacteria;Actinobacteria;;;Actinobacteria;;;;;;;Actinobacteria | [K01950](https://www.genome.jp/entry/K01950) | 6.3.5.1 (99%);6.3.1.5 (1%) | NAD(+) synthase (glutamine-hydrolyzing) (99%);NAD(+) synthase (1%) | GO:0005737 (95%);GO:0005618 (0%);GO:0005886 (0%) | cytoplasm (95%);cell wall (0%);plasma membrane (0%) | GO:0003952 (100%);GO:0005524 (99%);GO:0008795 (98%) | NAD+ synthase (glutamine-hydrolyzing) activity (100%);ATP binding (99%);NAD+ synthase activity (98%) | GO:0009435 (99%) | NAD biosynthetic process (99%) | IPR041856 (100%);IPR003694 (100%);IPR014729 (99%) | Glutamine-dependent NAD+ synthetase | C-terminal (100%);NAD(+) synthetase (100%);Rossmann-like alpha/beta/alpha sandwich fold (99%) |  |  | 1 | 1 |
| AQDDVLEGLK | Staphylococcus | Bacteria |  |  |  | Firmicutes |  |  | Bacilli |  |  |  | Bacillales |  |  |  |  | Staphylococcaceae |  |  |  | Staphylococcus |  |  |  |  |  |  | Bacteria;Firmicutes;;;Bacilli;Bacillales;Staphylococcaceae;;Staphylococcus;;;Staphylococcus |  |  |  |  |  | GO:0003677 (100%) | DNA binding (100%) | GO:0006355 (100%) | regulation of transcription | DNA-templated (100%) | IPR000843 (100%);IPR006377 (100%);IPR010982 (100%) | LacI-type HTH domain (100%);Catabolite control protein A (100%);Lambda repressor-like |  |  | 1 | 100.8462 |
| AQGSNYDVAADLPAAAGLLVEK | Bacteria | Bacteria |  |  |  |  |  |  |  |  |  |  |  |  |  |  |  |  |  |  |  |  |  |  |  |  |  |  | Bacteria;;;;;;;;;;;Bacteria | [K00927](https://www.genome.jp/entry/K00927) | 2.7.2.3 | Phosphoglycerate kinase (100%) | GO:0005737 (99%) | cytoplasm (99%) | GO:0004618 (99%);GO:0005524 (98%);GO:0016301 (1%) | phosphoglycerate kinase activity (99%);ATP binding (98%);kinase activity (1%) | GO:0006096 (99%) | glycolytic process (99%) | IPR001576 (100%);IPR015824 (100%);IPR036043 (100%) | Phosphoglycerate kinase (100%);Phosphoglycerate kinase | N-terminal (100%);Phosphoglycerate kinase superfamily (100%) |  |  | 1 | 100.9231 |
| ASAGPTHTPAVCQR | Mycobacterium leprae | Bacteria |  |  |  | Actinobacteria |  |  | Actinobacteria |  |  |  | Corynebacteriales |  |  |  |  | Mycobacteriaceae |  |  |  | Mycobacterium |  |  |  | Mycobacterium leprae |  |  | Bacteria;Actinobacteria;;;Actinobacteria;Corynebacteriales;Mycobacteriaceae;;Mycobacterium;;Mycobacterium leprae;Mycobacterium leprae | [K00784](https://www.genome.jp/entry/K00784) | 3.1.26.11 | Ribonuclease Z (100%) |  |  | GO:0004519 (100%);GO:0046872 (100%) | endonuclease activity (100%);metal ion binding (100%) | GO:0008033 (100%) | tRNA processing (100%) | IPR001279 (100%);IPR036866 (100%) | Metallo-beta-lactamase (100%);Ribonuclease Z/Hydroxyacylglutathione hydrolase-like (100%) |  |  |  | 1 | 1 |
| ASEAGADFVGGLDK | Mycoplasma genitalium | Bacteria |  |  |  | Tenericutes |  |  | Mollicutes |  |  |  | Mycoplasmatales |  |  |  |  | Mycoplasmataceae |  |  |  | Mycoplasma |  |  |  | Mycoplasma genitalium |  |  | Bacteria;Tenericutes;;;Mollicutes;Mycoplasmatales;Mycoplasmataceae;;Mycoplasma;;Mycoplasma genitalium;Mycoplasma genitalium |  |  |  | GO:0015934 (100%) | large ribosomal subunit (100%) | GO:0000049 (100%);GO:0003735 (100%);GO:0019843 (100%) | tRNA binding (100%);structural constituent of ribosome (100%);rRNA binding (100%) | GO:0006412 (100%);GO:0006417 (100%) | translation (100%);regulation of translation (100%) | IPR002143 (100%);IPR005878 (100%);IPR016095 (100%) | Ribosomal protein L1 (100%);Ribosomal protein L1 | bacterial-type (100%);Ribosomal protein L1 |  |  | 1 | 100.4538 |
| ASELGYSDVHLLLGNDGLR | Bacteria | Bacteria |  |  |  |  |  |  |  |  |  |  |  |  |  |  |  |  |  |  |  |  |  |  |  |  |  |  | Bacteria;;;;;;;;;;;Bacteria | [K14459](https://www.genome.jp/dbget-bin/www_bget?K14459+3.2.1.52+R09323) | 3.2.1.52 | Beta-N-acetylhexosaminidase (100%) | GO:0005576 (97%);GO:0016020 (92%);GO:0005618 (90%) | extracellular region (97%);membrane (92%);cell wall (90%) | GO:0004553 (49%);GO:0004563 (49%);GO:0102148 (49%) | hydrolase activity | hydrolyzing O-glycosyl compounds (49%);beta-N-acetylhexosaminidase activity (49%);N-acetyl-beta-D-galactosaminidase activity (49%) | GO:0005975 (97%);GO:0008152 (2%) | carbohydrate metabolic process (97%);metabolic process (2%) | IPR015883 (100%);IPR017853 (100%);IPR005877 (97%) | Glycoside hydrolase family 20 |  |  | 1 | 100.6923 |
| ASESPEMTPAAAVAK | Streptomyces | Bacteria |  |  |  | Actinobacteria |  |  | Actinobacteria |  |  |  | Streptomycetales |  |  |  |  | Streptomycetaceae |  |  |  | Streptomyces |  |  |  |  |  |  | Bacteria;Actinobacteria;;;Actinobacteria;Streptomycetales;Streptomycetaceae;;Streptomyces;;;Streptomyces |  |  |  | GO:0005886 (100%) | plasma membrane (100%) |  |  |  |  | IPR029046 (100%) | Lipoprotein localisation LolA/LolB/LppX (100%) |  |  |  | 1 | 100.5462 |
| ATAAEVFGLPLDSVTGEQR | Bacteria | Bacteria |  |  |  |  |  |  |  |  |  |  |  |  |  |  |  |  |  |  |  |  |  |  |  |  |  |  | Bacteria;;;;;;;;;;;Bacteria | K02320 | 2.7.7.7 | DNA-directed DNA polymerase (100%) | GO:0005737 (0%) | cytoplasm (0%) | GO:0003887 (100%);GO:0003677 (59%);GO:0008408 (56%) | DNA-directed DNA polymerase activity (100%);DNA binding (59%);3'-5' exonuclease activity (56%) | GO:0006261 (59%);GO:0006281 (57%) | DNA-dependent DNA replication (59%);DNA repair (57%) | IPR001098 (100%);IPR002298 (100%);IPR019760 (100%) | DNA-directed DNA polymerase | family A |  |  | 1 | 100.3692 |
| ATNESAVMAALSR | Campylobacter | Bacteria |  |  |  | Proteobacteria | delta/epsilon subdivisions |  | Epsilonproteobacteria |  |  |  | Campylobacterales |  |  |  |  | Campylobacteraceae |  |  |  | Campylobacter |  |  |  |  |  |  | Bacteria;Proteobacteria;delta/epsilon subdivisions;;Epsilonproteobacteria;Campylobacterales;Campylobacteraceae;;Campylobacter;;;Campylobacter | [K01925](https://www.genome.jp/entry/K01925) | 6.3.2.9 | UDP-N-acetylmuramoyl-L-alanine--D-glutamate ligase (100%) | GO:0005737 (96%) | cytoplasm (96%) | GO:0005524 (100%);GO:0008764 (100%) | ATP binding (100%);UDP-N-acetylmuramoylalanine-D-glutamate ligase activity (100%) | GO:0007049 (96%);GO:0008360 (96%);GO:0009252 (96%) | cell cycle (96%);regulation of cell shape (96%);peptidoglycan biosynthetic process (96%) | IPR036615 (100%);IPR013221 (98%);IPR036565 (98%) | Mur ligase | C-terminal domain superfamily (100%);Mur ligase |  |  | 1 | 100.2308 |
| AVESGSAFGLR | Streptomyces clavuligerus | Bacteria |  |  |  | Actinobacteria |  |  | Actinobacteria |  |  |  | Streptomycetales |  |  |  |  | Streptomycetaceae |  |  |  | Streptomyces |  |  |  | Streptomyces clavuligerus |  |  | Bacteria;Actinobacteria;;;Actinobacteria;Streptomycetales;Streptomycetaceae;;Streptomyces;;Streptomyces clavuligerus;Streptomyces clavuligerus | K10807 | 1.17.4.1 | Ribonucleoside-diphosphate reductase (100%) |  |  | GO:0000166 (100%);GO:0004748 (100%);GO:0031419 (100%) | nucleotide binding (100%);ribonucleoside-diphosphate reductase activity | thioredoxin disulfide as acceptor (100%);cobalamin binding (100%) | GO:0006260 (100%);GO:0071897 (100%) | DNA replication (100%);DNA biosynthetic process (100%) | IPR000788 (100%);IPR013344 (100%);IPR013678 (100%) | Ribonucleotide reductase large subunit |  |  | 1 | 100.7692 |
| AVTTDDFMTAEWAR | Deinococcus | Bacteria |  |  |  | Deinococcus-Thermus |  |  | Deinococci |  |  |  | Deinococcales |  |  |  |  | Deinococcaceae |  |  |  | Deinococcus |  |  |  |  |  |  | Bacteria;Deinococcus-Thermus;;;Deinococci;Deinococcales;Deinococcaceae;;Deinococcus;;;Deinococcus | K01951 | 6.3.5.2 | GMP synthase (glutamine-hydrolyzing) (100%) | GO:0005829 (4%) | cytosol (4%) | GO:0003922 (100%);GO:0005524 (100%);GO:0016462 (96%) | GMP synthase (glutamine-hydrolyzing) activity (100%);ATP binding (100%);pyrophosphatase activity (96%) | GO:0006541 (100%);GO:0006177 (4%) | glutamine metabolic process (100%);GMP biosynthetic process (4%) | IPR001674 (100%);IPR004739 (100%);IPR014729 (100%) | GMP synthase | C-terminal (100%);GMP synthase |  |  | 1 | 100.3769 |
| AVTTEDFMTADWAR | Bacteria | Bacteria |  |  |  |  |  |  |  |  |  |  |  |  |  |  |  |  |  |  |  |  |  |  |  |  |  |  | Bacteria;;;;;;;;;;;Bacteria | 6.3.5.2 | 6.3.5.2 | GMP synthase (glutamine-hydrolyzing) (100%) |  |  | GO:0003922 (100%);GO:0005524 (100%);GO:0016462 (77%) | GMP synthase (glutamine-hydrolyzing) activity (100%);ATP binding (100%);pyrophosphatase activity (77%) | GO:0006541 (95%);GO:0006529 (6%) | glutamine metabolic process (95%);asparagine biosynthetic process (6%) | IPR001674 (100%);IPR014729 (95%);IPR025777 (95%) | GMP synthase | C-terminal (100%);Rossmann-like alpha/beta/alpha sandwich fold (95%);GMP synthetase ATP pyrophosphatase domain (95%) |  |  | 1 | 1 |
| AVVLPQGK | Firmicutes | Bacteria |  |  |  | Firmicutes |  |  |  |  |  |  |  |  |  |  |  |  |  |  |  |  |  |  |  |  |  |  | Bacteria;Firmicutes;;;;;;;;;;Firmicutes | K02320 | 2.7.7.7 (50%);3.1.11.- (50%) | DNA-directed DNA polymerase (50%);Exodeoxyribonucleases producing 5'-phosphomonoesters (50%) |  |  | GO:0004527 (99%);GO:0003887 (1%);GO:0004519 (1%) | exonuclease activity (99%);DNA-directed DNA polymerase activity (1%);endonuclease activity (1%) | GO:0006260 (1%);GO:0006310 (1%) | DNA replication (1%);DNA recombination (1%) | IPR027417 (100%);IPR038729 (85%);IPR027267 (0%) | P-loop containing nucleoside triphosphate hydrolase (100%);Rad50/SbcC-type AAA domain (85%);AH/BAR domain superfamily (0%) |  |  |  | 1 | 100.4462 |
| AVWAAQYGLGGGK | Neisseria | Bacteria |  |  |  | Proteobacteria |  |  | Betaproteobacteria |  |  |  | Neisseriales |  |  |  |  | Neisseriaceae |  |  |  | Neisseria |  |  |  |  |  |  | Bacteria;Proteobacteria;;;Betaproteobacteria;Neisseriales;Neisseriaceae;;Neisseria;;;Neisseria |  |  |  | GO:0009279 (100%);GO:0016021 (100%) | cell outer membrane (100%);integral component of membrane (100%) | GO:0022857 (100%) | transmembrane transporter activity (100%) |  |  | IPR000531 (100%);IPR010917 (100%);IPR010949 (100%) | TonB-dependent receptor-like | beta-barrel (100%);TonB-dependent receptor |  |  | 1 | 100.8462 |
| AWSVEHFVLAEGAR | Frankia casuarinae | Bacteria |  |  |  | Actinobacteria |  |  | Actinobacteria |  |  |  | Frankiales |  |  |  |  | Frankiaceae |  |  |  | Frankia |  |  |  | Frankia casuarinae |  |  | Bacteria;Actinobacteria;;;Actinobacteria;Frankiales;Frankiaceae;;Frankia;;Frankia casuarinae;Frankia casuarinae | 3.5.1.103 | 3.5.1.103 | N-acetyl-1-D-myo-inositol-2-amino-2-deoxy-alpha-D-glucopyranosiddeacetylase (100%) |  |  | GO:0008270 (100%);GO:0035595 (100%) | zinc ion binding (100%);N-acetylglucosaminylinositol deacetylase activity (100%) | GO:0010125 (100%) | mycothiol biosynthetic process (100%) | IPR003737 (100%);IPR017810 (100%);IPR024078 (100%) | N-acetylglucosaminyl phosphatidylinositol deacetylase-related (100%);Mycothiol biosynthesis protein | MshB (100%);Putative deacetylase LmbE-like domain superfamily (100%) |  |  | 1 | 100.6231 |
| CVLDGEVVVPR | Mycobacteriaceae | Bacteria |  |  |  | Actinobacteria |  |  | Actinobacteria |  |  |  | Corynebacteriales |  |  |  |  | Mycobacteriaceae |  |  |  |  |  |  |  |  |  |  | Bacteria;Actinobacteria;;;Actinobacteria;Corynebacteriales;Mycobacteriaceae;;;;;Mycobacteriaceae | [K01971](https://www.genome.jp/entry/K01971) | 6.5.1.1 | DNA ligase (ATP) (100%) |  |  | GO:0003910 (97%);GO:0005524 (92%);GO:0016874 (3%) | DNA ligase (ATP) activity (97%);ATP binding (92%);ligase activity (3%) | GO:0006310 (92%);GO:0051103 (83%);GO:0006281 (10%) | DNA recombination (92%);DNA ligation involved in DNA repair (83%);DNA repair (10%) | IPR012309 (100%);IPR012310 (100%);IPR012340 (100%) | DNA ligase | ATP-dependent |  |  | 1 | 1 |
| DAASLTGVK | Shewanella | Bacteria |  |  |  | Proteobacteria |  |  | Gammaproteobacteria |  |  |  | Alteromonadales |  |  |  |  | Shewanellaceae |  |  |  | Shewanella |  |  |  |  |  |  | Bacteria;Proteobacteria;;;Gammaproteobacteria;Alteromonadales;Shewanellaceae;;Shewanella;;;Shewanella |  |  |  |  |  | GO:0005524 (100%);GO:0016887 (100%);GO:0051082 (100%) | ATP binding (100%);ATPase activity (100%);unfolded protein binding (100%) | GO:0006457 (100%);GO:0016226 (100%) | protein folding (100%);iron-sulfur cluster assembly (100%) | IPR010236 (100%);IPR013126 (100%);IPR018181 (100%) | ISC system FeS cluster assembly | HscA chaperone (100%);Heat shock protein 70 family (100%);Heat shock protein 70 |  |  | 1 | 100.4615 |
| DAAYDFSFSGLK | Actinobacteria | Bacteria |  |  |  | Actinobacteria |  |  | Actinobacteria |  |  |  |  |  |  |  |  |  |  |  |  |  |  |  |  |  |  |  | Bacteria;Actinobacteria;;;Actinobacteria;;;;;;;Actinobacteria | K01409 | 2.3.1.234 | N(6)-L-threonylcarbamoyladenine synthase (100%) | GO:0005737 (98%);GO:0000408 (1%) | cytoplasm (98%);EKC/KEOPS complex (1%) | GO:0061711 (100%);GO:0005506 (98%);GO:0004222 (94%) | N(6)-L-threonylcarbamoyladenine synthase activity (100%);iron ion binding (98%);metalloendopeptidase activity (94%) | GO:0002949 (99%);GO:0006474 (1%) | tRNA threonylcarbamoyladenosine modification (99%);N-terminal protein amino acid acetylation (1%) | IPR000905 (100%);IPR017861 (100%);IPR022450 (100%) | Gcp-like domain (100%);Kae1/TsaD family (100%);tRNA N6-adenosine threonylcarbamoyltransferase | TsaD (100%) |  |  | 1 | 100.8462 |
| DAEDAEGAEDGAEK | Streptomyces | Bacteria |  |  |  | Actinobacteria |  |  | Actinobacteria |  |  |  | Streptomycetales |  |  |  |  | Streptomycetaceae |  |  |  | Streptomyces |  |  |  |  |  |  | Bacteria;Actinobacteria;;;Actinobacteria;Streptomycetales;Streptomycetaceae;;Streptomyces;;;Streptomyces | [K13292](https://www.genome.jp/dbget-bin/www_bget?ko:K13292) | 2.5.1.145 | Phosphatidylglycerol--prolipoprotein diacylglyceryl transferase (100%) | GO:0005887 (100%) | integral component of plasma membrane (100%) | GO:0008961 (100%) | phosphatidylglycerol-prolipoprotein diacylglyceryl transferase activity (100%) | GO:0042158 (100%) | lipoprotein biosynthetic process (100%) | IPR001640 (100%) | Phosphatidylglycerol--prolipoprotein diacylglyceryl transferase Lgt (100%) |  |  |  | 1 | 100.5385 |
| DASPALVAGMAK | Streptomyces | Bacteria |  |  |  | Actinobacteria |  |  | Actinobacteria |  |  |  | Streptomycetales |  |  |  |  | Streptomycetaceae |  |  |  | Streptomyces |  |  |  |  |  |  | Bacteria;Actinobacteria;;;Actinobacteria;Streptomycetales;Streptomycetaceae;;Streptomyces;;;Streptomyces | [K01924](https://www.genome.jp/dbget-bin/www_bget?ko:K01924) | 6.3.2.8 | UDP-N-acetylmuramate--L-alanine ligase (100%) | GO:0005737 (100%) | cytoplasm (100%) | GO:0005524 (100%);GO:0008763 (100%) | ATP binding (100%);UDP-N-acetylmuramate-L-alanine ligase activity (100%) | GO:0007049 (100%);GO:0008360 (100%);GO:0009252 (100%) | cell cycle (100%);regulation of cell shape (100%);peptidoglycan biosynthetic process (100%) | IPR000713 (100%);IPR004101 (100%);IPR005758 (100%) | Mur ligase | N-terminal catalytic domain (100%);Mur ligase |  |  | 1 | 100.0077 |
| DDENDTYTDESSEFLALTK | Mycoplasma penetrans | Bacteria |  |  |  | Tenericutes |  |  | Mollicutes |  |  |  | Mycoplasmatales |  |  |  |  | Mycoplasmataceae |  |  |  | Mycoplasma |  |  |  | Mycoplasma penetrans |  |  | Bacteria;Tenericutes;;;Mollicutes;Mycoplasmatales;Mycoplasmataceae;;Mycoplasma;;Mycoplasma penetrans;Mycoplasma penetrans | K11072 | 7.6.2.11 | ABC-type polyamine transporter (100%) | GO:0005886 (100%) | plasma membrane (100%) | GO:0005524 (100%);GO:0015594 (100%);GO:0015595 (100%) | ATP binding (100%);ATPase-coupled putrescine transmembrane transporter activity (100%);ATPase-coupled spermidine transmembrane transporter activity (100%) |  |  | IPR003439 (100%);IPR003593 (100%);IPR008995 (100%) | ABC transporter-like (100%);AAA+ ATPase domain (100%);Molybdate/tungstate binding | C-terminal (100%) |  |  | 1 | 100.3923 |
| DEQAAANMAR | Campylobacter | Bacteria |  |  |  | Proteobacteria | delta/epsilon subdivisions |  | Epsilonproteobacteria |  |  |  | Campylobacterales |  |  |  |  | Campylobacteraceae |  |  |  | Campylobacter |  |  |  |  |  |  | Bacteria;Proteobacteria;delta/epsilon subdivisions;;Epsilonproteobacteria;Campylobacterales;Campylobacteraceae;;Campylobacter;;;Campylobacter | [K17251](https://www.genome.jp/entry/K17251) | 2.4.99.19 | Undecaprenyl-diphosphooligosaccharide--protein glycotransferase (100%) | GO:0016021 (88%);GO:0016020 (12%);GO:0005886 (3%) | integral component of membrane (88%);membrane (12%);plasma membrane (3%) | GO:0004576 (92%);GO:0016757 (4%);GO:0046872 (3%) | oligosaccharyl transferase activity (92%);transferase activity | transferring glycosyl groups (4%);metal ion binding (3%) | GO:0006486 (90%);GO:0006487 (1%);GO:0018279 (1%) | protein glycosylation (90%);protein N-linked glycosylation (1%);protein N-linked glycosylation via asparagine (1%) | IPR041563 (100%);IPR003674 (99%) | STT3 subunit PglB |  |  | 1 | 100.2385 |
| DFDTYEGYEAR | Alphaproteobacteria | Bacteria |  |  |  | Proteobacteria |  |  | Alphaproteobacteria |  |  |  |  |  |  |  |  |  |  |  |  |  |  |  |  |  |  |  | Bacteria;Proteobacteria;;;Alphaproteobacteria;;;;;;;Alphaproteobacteria |  |  |  | GO:0005737 (100%) | cytoplasm (100%) |  |  | GO:0042274 (100%) | ribosomal small subunit biogenesis (100%) | IPR003728 (100%);IPR028989 (100%);IPR028998 (100%) | Ribosome maturation factor RimP (100%);Ribosome maturation factor RimP | N-terminal (100%);Ribosome maturation factor RimP |  |  | 1 | 1 |
| DFEETVDVLEK | Campylobacter | Bacteria |  |  |  | Proteobacteria | delta/epsilon subdivisions |  | Epsilonproteobacteria |  |  |  | Campylobacterales |  |  |  |  | Campylobacteraceae |  |  |  | Campylobacter |  |  |  |  |  |  | Bacteria;Proteobacteria;delta/epsilon subdivisions;;Epsilonproteobacteria;Campylobacterales;Campylobacteraceae;;Campylobacter;;;Campylobacter | [K06168](https://www.genome.jp/entry/K06168) | 2.8.4.3 | tRNA-2-methylthio-N(6)-dimethylallyladenosine synthase (100%) | GO:0005737 (100%) | cytoplasm (100%) | GO:0035596 (100%);GO:0046872 (100%);GO:0051539 (100%) | methylthiotransferase activity (100%);metal ion binding (100%);4 iron | 4 sulfur cluster binding (100%) | GO:0006400 (100%) | tRNA modification (100%) | IPR002792 (100%);IPR005839 (100%);IPR006463 (100%) | TRAM domain (100%);Methylthiotransferase (100%);tRNA-2-methylthio-N(6)-dimethylallyladenosine synthase MiaB (100%) |  |  | 1 | 100.6923 |
| DGEQMVGPDGK | Actinobacteria | Bacteria |  |  |  | Actinobacteria |  |  | Actinobacteria |  |  |  |  |  |  |  |  |  |  |  |  |  |  |  |  |  |  |  | Bacteria;Actinobacteria;;;Actinobacteria;;;;;;;Actinobacteria | K03010 | 2.7.7.6 | DNA-directed RNA polymerase (100%) |  |  | GO:0003677 (100%);GO:0003899 (100%);GO:0032549 (94%) | DNA binding (100%);DNA-directed 5'-3' RNA polymerase activity (100%);ribonucleoside binding (94%) | GO:0006351 (100%) | transcription | DNA-templated (100%) | IPR007120 (100%);IPR015712 (100%);IPR037033 (100%) | DNA-directed RNA polymerase |  |  | 1 | 100.3846 |
| DGESSPAEYSVSLTR | Bacteria | Bacteria |  |  |  |  |  |  |  |  |  |  |  |  |  |  |  |  |  |  |  |  |  |  |  |  |  |  | Bacteria;;;;;;;;;;;Bacteria |  |  |  |  |  | GO:0003677 (100%);GO:0003700 (90%);GO:0016829 (0%) | DNA binding (100%);DNA-binding transcription factor activity (90%);lyase activity (0%) | GO:0006355 (3%);GO:0015716 (0%) | regulation of transcription | DNA-templated (3%);organic phosphonate transport (0%) | IPR028978 (100%);IPR011663 (99%);IPR012702 (98%) | Chorismate pyruvate-lyase/UbiC transcription regulator-associated domain superfamily (100%);UbiC transcription regulator-associated (99%);Phosphonate C-P lyase system |  |  | 1 | 100.6231 |
| DGTLNVAYNCVDR | Streptomyces | Bacteria |  |  |  | Actinobacteria |  |  | Actinobacteria |  |  |  | Streptomycetales |  |  |  |  | Streptomycetaceae |  |  |  | Streptomyces |  |  |  |  |  |  | Bacteria;Actinobacteria;;;Actinobacteria;Streptomycetales;Streptomycetaceae;;Streptomyces;;;Streptomyces | [K01895](https://www.genome.jp/entry/K01895) | 6.2.1.1 | Acetate--CoA ligase (100%) |  |  | GO:0003987 (100%);GO:0005524 (100%);GO:0046872 (100%) | acetate-CoA ligase activity (100%);ATP binding (100%);metal ion binding (100%) | GO:0019427 (97%) | acetyl-CoA biosynthetic process from acetate (97%) | IPR000873 (100%);IPR011904 (100%);IPR020845 (100%) | AMP-dependent synthetase/ligase (100%);Acetate-CoA ligase (100%);AMP-binding | conserved site (100%) |  |  | 1 | 1 |
| DLADPSYR | Bacteria | Bacteria |  |  |  |  |  |  |  |  |  |  |  |  |  |  |  |  |  |  |  |  |  |  |  |  |  |  | Bacteria;;;;;;;;;;;Bacteria |  |  |  | GO:0016021 (83%);GO:0005886 (11%);GO:0005887 (6%) | integral component of membrane (83%);plasma membrane (11%);integral component of plasma membrane (6%) | GO:0003700 (11%);GO:0043565 (11%) | DNA-binding transcription factor activity (11%);sequence-specific DNA binding (11%) | GO:0006869 (11%);GO:0071555 (11%);GO:0006855 (6%) | lipid transport (11%);cell wall organization (11%);drug transmembrane transport (6%) | IPR004869 (83%);IPR009057 (11%);IPR018060 (11%) | Membrane transport protein MMPL domain (83%);Homeobox-like domain superfamily (11%);DNA binding HTH domain | AraC-type (11%) |  |  | 1 | 100.3846 |
| DLGGAVHTDYEVK | Streptomyces | Bacteria |  |  |  | Actinobacteria |  |  | Actinobacteria |  |  |  | Streptomycetales |  |  |  |  | Streptomycetaceae |  |  |  | Streptomyces |  |  |  |  |  |  | Bacteria;Actinobacteria;;;Actinobacteria;Streptomycetales;Streptomycetaceae;;Streptomyces;;;Streptomyces | K18237 | 5.3.1.29 | Ribose 1 | 5-bisphosphate isomerase (100%) | GO:0016021 (100%) | integral component of membrane (100%) | GO:0043917 (4%) | ribose 1 | 5-bisphosphate isomerase activity (4%) |  |  | IPR036188 (100%) |  |  | 1 | 1 |
| DLNAPVVVVQHMPEGFTASLADR | Bacillus | Bacteria |  |  |  | Firmicutes |  |  | Bacilli |  |  |  | Bacillales |  |  |  |  | Bacillaceae |  |  |  | Bacillus |  |  |  |  |  |  | Bacteria;Firmicutes;;;Bacilli;Bacillales;Bacillaceae;;Bacillus;;;Bacillus | [K03412](https://www.genome.jp/entry/K03412) | 3.1.1.61 ;3.5.1.44 (94%) | Protein-glutamate methylesterase (100%);Protein-glutamine glutaminase (94%) | GO:0005737 (100%) | cytoplasm (100%) | GO:0008984 (100%);GO:0050568 (100%);GO:0000156 (94%) | protein-glutamate methylesterase activity (100%);protein-glutamine glutaminase activity (100%);phosphorelay response regulator activity (94%) | GO:0006482 (100%);GO:0006935 (100%);GO:0018277 (100%) | protein demethylation (100%);chemotaxis (100%);protein deamination (100%) | IPR000673 (100%);IPR001789 (100%);IPR008248 (100%) | Signal transduction response regulator | chemotaxis |  |  | 1 | 100.5308 |
| DNSNVAGFDGQSR | Enterobacteriaceae | Bacteria |  |  |  | Proteobacteria |  |  | Gammaproteobacteria |  |  |  | Enterobacterales |  |  |  |  | Enterobacteriaceae |  |  |  |  |  |  |  |  |  |  | Bacteria;Proteobacteria;;;Gammaproteobacteria;Enterobacterales;Enterobacteriaceae;;;;;Enterobacteriaceae |  |  |  |  |  |  |  |  |  |  |  |  |  |  | 1 | 100.2385 |
| DRPNLQVSLMGLPSSMVMEAVASGR | Agrobacterium tumefaciens | Bacteria |  |  |  | Proteobacteria |  |  | Alphaproteobacteria |  |  |  | Rhizobiales |  |  |  |  | Rhizobiaceae |  |  |  | Agrobacterium |  | Agrobacterium tumefaciens complex |  | Agrobacterium tumefaciens |  |  | Bacteria;Proteobacteria;;;Alphaproteobacteria;Rhizobiales;Rhizobiaceae;;Agrobacterium;Agrobacterium tumefaciens complex;Agrobacterium tumefaciens;Agrobacterium tumefaciens |  |  |  |  |  | GO:0003677 (100%);GO:0003700 (100%) | DNA binding (100%);DNA-binding transcription factor activity (100%) |  |  | IPR000847 (100%);IPR005119 (100%);IPR036388 (100%) | Transcription regulator HTH | LysR (100%);LysR |  |  | 1 | 1 |
| DVSDDALK | Bacteria | Bacteria |  |  |  |  |  |  |  |  |  |  |  |  |  |  |  |  |  |  |  |  |  |  |  |  |  |  | Bacteria;;;;;;;;;;;Bacteria | K07547 | 1.1.1.35 (50%);2.5.1.3 (50%);4.2.1.17 (50%) | 3-hydroxyacyl-CoA dehydrogenase (50%);Thiamine phosphate synthase (50%);Enoyl-CoA hydratase (50%) | GO:0016021 (1%) | integral component of membrane (1%) | GO:0003677 (97%);GO:0005524 (97%);GO:0008270 (97%) | DNA binding (97%);ATP binding (97%);zinc ion binding (97%) | GO:0045892 (97%);GO:0006635 (1%);GO:0009228 (1%) | negative regulation of transcription | DNA-templated (97%);fatty acid beta-oxidation (1%);thiamine biosynthetic process (1%) | IPR003796 (95%);IPR005144 (95%);IPR041311 (2%) | Ribonucleotide reductase regulator NrdR-like (95%);ATP-cone domain (95%);Large polyvalent protein associated domain 29 (2%) |  |  | 1 | 100.3923 |
| DWDQFLK | Bacteria | Bacteria |  |  |  |  |  |  |  |  |  |  |  |  |  |  |  |  |  |  |  |  |  |  |  |  |  |  | Bacteria;;;;;;;;;;;Bacteria |  | 2.1.1.- (50%);4.2.1.20 (50%) | Methyltransferases (50%);Tryptophan synthase (50%) |  |  | GO:0003677 (25%);GO:0004834 (25%);GO:0008080 (25%) | DNA binding (25%);tryptophan synthase activity (25%);N-acetyltransferase activity (25%) | GO:0006306 (25%) | DNA methylation (25%) | IPR000182 (20%);IPR001091 (20%);IPR001926 (20%) | GNAT domain (20%);Restriction/modification DNA-methylase (20%);Pyridoxal-phosphate dependent enzyme (20%) |  |  |  | 1 | 1 |
| EAANAGQLGR | Neisseria | Bacteria |  |  |  | Proteobacteria |  |  | Betaproteobacteria |  |  |  | Neisseriales |  |  |  |  | Neisseriaceae |  |  |  | Neisseria |  |  |  |  |  |  | Bacteria;Proteobacteria;;;Betaproteobacteria;Neisseriales;Neisseriaceae;;Neisseria;;;Neisseria |  | 2.7.-.- | Transferring phosphorus-containing groups (100%) | GO:0005886 (100%);GO:0016021 (100%) | plasma membrane (100%);integral component of membrane (100%) | GO:0004672 (100%);GO:0005524 (100%);GO:0016491 (19%) | protein kinase activity (100%);ATP binding (100%);oxidoreductase activity (19%) | GO:0006744 (100%);GO:0010795 (100%) | ubiquinone biosynthetic process (100%);regulation of ubiquinone biosynthetic process (100%) | IPR004147 (100%);IPR010232 (100%);IPR011009 (100%) | UbiB domain (100%);Probable protein kinase UbiB (100%);Protein kinase-like domain superfamily (100%) |  |  |  | 1 | 1 |
| EAEQQAAAEALK | Bacillales | Bacteria |  |  |  | Firmicutes |  |  | Bacilli |  |  |  | Bacillales |  |  |  |  |  |  |  |  |  |  |  |  |  |  |  | Bacteria;Firmicutes;;;Bacilli;Bacillales;;;;;;Bacillales | [K03685](https://www.genome.jp/entry/K03685) | 3.1.26.3 | Ribonuclease III (100%) | GO:0005737 (99%) | cytoplasm (99%) | GO:0004525 (100%);GO:0019843 (99%);GO:0046872 (99%) | ribonuclease III activity (100%);rRNA binding (99%);metal ion binding (99%) | GO:0006364 (99%);GO:0006397 (99%);GO:0008033 (99%) | rRNA processing (99%);mRNA processing (99%);tRNA processing (99%) | IPR014720 (100%);IPR000999 (100%);IPR036389 (100%) | Double-stranded RNA-binding domain (100%);Ribonuclease III domain (100%);Ribonuclease III | endonuclease domain superfamily (100%) |  |  | 1 | 100.0692 |
| EAGVEADGR | Streptomyces | Bacteria |  |  |  | Actinobacteria |  |  | Actinobacteria |  |  |  | Streptomycetales |  |  |  |  | Streptomycetaceae |  |  |  | Streptomyces |  |  |  |  |  |  | Bacteria;Actinobacteria;;;Actinobacteria;Streptomycetales;Streptomycetaceae;;Streptomyces;;;Streptomyces |  | 1.3.1.- | With NAD(+) or NADP(+) as acceptor (100%) |  |  | GO:0016491 (100%) | oxidoreductase activity (100%) | GO:0017000 (10%) | antibiotic biosynthetic process (10%) | IPR002347 (100%);IPR020904 (100%);IPR036291 (100%) | Short-chain dehydrogenase/reductase SDR (100%);Short-chain dehydrogenase/reductase | conserved site (100%);NAD(P)-binding domain superfamily (100%) |  |  | 1 | 1 |
| EALGTDLVK | Bacteria | Bacteria |  |  |  |  |  |  |  |  |  |  |  |  |  |  |  |  |  |  |  |  |  |  |  |  |  |  | Bacteria;;;;;;;;;;;Bacteria | [K03149](https://www.genome.jp/entry/K03149) | 2.8.1.10 | Thiazole synthase (100%) | GO:0005737 (100%) | cytoplasm (100%) | GO:0016783 (97%);GO:0016740 (3%);GO:0003824 (0%) | sulfurtransferase activity (97%);transferase activity (3%);catalytic activity (0%) | GO:0009228 (100%);GO:0009229 (100%) | thiamine biosynthetic process (100%);thiamine diphosphate biosynthetic process (100%) | IPR008867 (100%);IPR013785 (100%);IPR033983 (100%) | Thiazole synthase (100%);Aldolase-type TIM barrel (100%);Thiazole synthase ThiG (100%) |  |  |  | 1 | 100.0769 |
| EALPARPR | Actinobacteria | Bacteria |  |  |  | Actinobacteria |  |  | Actinobacteria |  |  |  |  |  |  |  |  |  |  |  |  |  |  |  |  |  |  |  | Bacteria;Actinobacteria;;;Actinobacteria;;;;;;;Actinobacteria | K00982 | 2.7.7.42 ;2.7.7.89 | [Glutamine synthetase] adenylyltransferase (100%);[Glutamine synthetase]-adenylyl-L-tyrosine phosphorylase (100%) | GO:0016021 (97%);GO:0005886 (11%);GO:0005737 (1%) | integral component of membrane (97%);plasma membrane (11%);cytoplasm (1%) | GO:0005524 (99%);GO:0003677 (97%);GO:0000287 (1%) | ATP binding (99%);DNA binding (97%);magnesium ion binding (1%) | GO:0051301 (49%);GO:0007049 (11%);GO:0007059 (11%) | cell division (49%);cell cycle (11%);chromosome segregation (11%) | IPR002543 (97%);IPR003593 (97%);IPR018541 (97%) | FtsK domain (97%);AAA+ ATPase domain (97%);FtsK gamma domain (97%) |  |  |  | 1 | 100.8462 |
| EAMAVDVVYDGAAALER | Bacteria | Bacteria |  |  |  |  |  |  |  |  |  |  |  |  |  |  |  |  |  |  |  |  |  |  |  |  |  |  | Bacteria;;;;;;;;;;;Bacteria |  |  |  | GO:0005829 (0%);GO:0032993 (0%) | cytosol (0%);protein-DNA complex (0%) | GO:0003677 (100%);GO:0000156 (0%);GO:0000976 (0%) | DNA binding (100%);phosphorelay response regulator activity (0%);transcription regulatory region sequence-specific DNA binding (0%) | GO:0000160 (98%);GO:0006355 (93%) | phosphorelay signal transduction system (98%);regulation of transcription | DNA-templated (93%) | IPR001789 (100%);IPR001867 (100%);IPR011006 (100%) | Signal transduction response regulator |  |  | 1 | 100.5385 |
| EATEAGADFVGGEDMVNK | Candidatus Solibacter usitatus | Bacteria |  |  |  | Acidobacteria |  |  | Acidobacteriia |  |  |  | Bryobacterales |  |  |  |  | Solibacteraceae |  |  |  | Candidatus Solibacter |  |  |  | Candidatus Solibacter usitatus |  |  | Bacteria;Acidobacteria;;;Acidobacteriia;Bryobacterales;Solibacteraceae;;Candidatus Solibacter;;Candidatus Solibacter usitatus;Candidatus Solibacter usitatus |  |  |  | GO:0015934 (100%) | large ribosomal subunit (100%) | GO:0000049 (100%);GO:0003735 (100%);GO:0019843 (100%) | tRNA binding (100%);structural constituent of ribosome (100%);rRNA binding (100%) | GO:0006412 (100%);GO:0006417 (100%) | translation (100%);regulation of translation (100%) | IPR002143 (100%);IPR005878 (100%);IPR016095 (100%) | Ribosomal protein L1 (100%);Ribosomal protein L1 | bacterial-type (100%);Ribosomal protein L1 |  |  | 1 | 100.6154 |
| EAVVLHK | Bacteria | Bacteria |  |  |  |  |  |  |  |  |  |  |  |  |  |  |  |  |  |  |  |  |  |  |  |  |  |  | Bacteria;;;;;;;;;;;Bacteria | [K00380](https://www.genome.jp/entry/K00380) | 1.8.1.2 | Assimilatory sulfite reductase (NADPH) (100%) | GO:0005829 (33%);GO:0016021 (33%) | cytosol (33%);integral component of membrane (33%) | GO:0000287 (33%);GO:0004783 (33%);GO:0008966 (33%) | magnesium ion binding (33%);sulfite reductase (NADPH) activity (33%);phosphoglucosamine mutase activity (33%) | GO:0000103 (33%);GO:0005975 (33%);GO:0019344 (33%) | sulfate assimilation (33%);carbohydrate metabolic process (33%);cysteine biosynthetic process (33%) | IPR001094 (25%);IPR001433 (25%);IPR001709 (25%) | Flavodoxin-like (25%);Oxidoreductase FAD/NAD(P)-binding (25%);Flavoprotein pyridine nucleotide cytochrome reductase (25%) |  |  |  | 1 | 100.4538 |
| EDMLQLFADSDR | Bacillus | Bacteria |  |  |  | Firmicutes |  |  | Bacilli |  |  |  | Bacillales |  |  |  |  | Bacillaceae |  |  |  | Bacillus |  |  |  |  |  |  | Bacteria;Firmicutes;;;Bacilli;Bacillales;Bacillaceae;;Bacillus;;;Bacillus |  |  |  | GO:0005737 (4%);GO:0032993 (4%) | cytoplasm (4%);protein-DNA complex (4%) | GO:0003677 (100%);GO:0003700 (88%);GO:0000976 (4%) | DNA binding (100%);DNA-binding transcription factor activity (88%);transcription regulatory region sequence-specific DNA binding (4%) | GO:0045892 (4%);GO:1900376 (4%) | negative regulation of transcription | DNA-templated (4%);regulation of secondary metabolite biosynthetic process (4%) | IPR002481 (100%);IPR036388 (100%);IPR036390 (100%) | Ferric-uptake regulator (100%);Winged helix-like DNA-binding domain superfamily (100%);Winged helix DNA-binding domain superfamily (100%) |  |  | 1 | 100.6154 |
| EEAAAHQTALK | Bacillus subtilis | Bacteria |  |  |  | Firmicutes |  |  | Bacilli |  |  |  | Bacillales |  |  |  |  | Bacillaceae |  |  |  | Bacillus |  | Bacillus subtilis group |  | Bacillus subtilis |  |  | Bacteria;Firmicutes;;;Bacilli;Bacillales;Bacillaceae;;Bacillus;Bacillus subtilis group;Bacillus subtilis;Bacillus subtilis |  | 2.3.1.- | Transferring groups other than amino-acyl groups (100%) | GO:0005737 (13%) | cytoplasm (13%) | GO:0031177 (100%);GO:0003824 (88%);GO:0016746 (13%) | phosphopantetheine binding (100%);catalytic activity (88%);transferase activity | transferring acyl groups (13%) | GO:0008152 (13%) | metabolic process (13%) | IPR000873 (100%);IPR001242 (100%);IPR006162 (100%) | AMP-dependent synthetase/ligase (100%);Condensation domain (100%);Phosphopantetheine attachment site (100%) |  |  | 1 | 100.5308 |
| EEGATGRPR | Actinobacteria | Bacteria |  |  |  | Actinobacteria |  |  | Actinobacteria |  |  |  |  |  |  |  |  |  |  |  |  |  |  |  |  |  |  |  | Bacteria;Actinobacteria;;;Actinobacteria;;;;;;;Actinobacteria |  | 4.1.2.- | Aldehyde-lyases (100%) |  |  | GO:0016832 (100%) | aldehyde-lyase activity (100%) | GO:0005975 (95%) | carbohydrate metabolic process (95%) | IPR005593 (100%);IPR009014 (100%);IPR018969 (100%) | Xylulose 5-phosphate/Fructose 6-phosphate phosphoketolase (100%);Transketolase C-terminal/Pyruvate-ferredoxin oxidoreductase domain II (100%);Xylulose 5-phosphate/Fructose 6-phosphate phosphoketolase | C-terminal (100%) |  |  | 1 | 99.99231 |
| EHFELGESLNWLDFMR | Campylobacter | Bacteria |  |  |  | Proteobacteria | delta/epsilon subdivisions |  | Epsilonproteobacteria |  |  |  | Campylobacterales |  |  |  |  | Campylobacteraceae |  |  |  | Campylobacter |  |  |  |  |  |  | Bacteria;Proteobacteria;delta/epsilon subdivisions;;Epsilonproteobacteria;Campylobacterales;Campylobacteraceae;;Campylobacter;;;Campylobacter | [K01875](https://www.genome.jp/dbget-bin/www_bget?ko+K01875) | 6.1.1.11 | Serine--tRNA ligase (100%) | GO:0005737 (93%) | cytoplasm (93%) | GO:0004828 (100%);GO:0005524 (100%) | serine-tRNA ligase activity (100%);ATP binding (100%) | GO:0006434 (100%);GO:0016260 (93%);GO:0097056 (93%) | seryl-tRNA aminoacylation (100%);selenocysteine biosynthetic process (93%);selenocysteinyl-tRNA(Sec) biosynthetic process (93%) | IPR002314 (100%);IPR002317 (100%);IPR006195 (100%) | Aminoacyl-tRNA synthetase | class II (G/ P/ S/T) (100%);Serine-tRNA ligase |  |  | 1 | 100.3846 |
| ELGFGTVDQAR | Firmicutes | Bacteria |  |  |  | Firmicutes |  |  |  |  |  |  |  |  |  |  |  |  |  |  |  |  |  |  |  |  |  |  | Bacteria;Firmicutes;;;;;;;;;;Firmicutes | [K02178](https://www.genome.jp/entry/K02178) | 2.7.11.1 | Non-specific serine/threonine protein kinase (100%) |  |  | GO:0004674 (43%);GO:0016301 (38%);GO:0005524 (23%) | protein serine/threonine kinase activity (43%);kinase activity (38%);ATP binding (23%) |  |  | IPR036890 (100%);IPR003594 (97%) | Histidine kinase/HSP90-like ATPase superfamily (100%);Histidine kinase/HSP90-like ATPase (97%) |  |  |  | 1 | 100.5385 |
| ELQADADAYGDDR | Gammaproteobacteria | Bacteria |  |  |  | Proteobacteria |  |  | Gammaproteobacteria |  |  |  |  |  |  |  |  |  |  |  |  |  |  |  |  |  |  |  | Bacteria;Proteobacteria;;;Gammaproteobacteria;;;;;;;Gammaproteobacteria | K03167 | 5.6.2.2 (99%);5.6.2.3 (1%) | DNA topoisomerase (ATP-hydrolyzing) (99%); (1%) | GO:0019897 (66%);GO:0005694 (65%);GO:0005737 (0%) | extrinsic component of plasma membrane (66%);chromosome (65%);cytoplasm (0%) | GO:0003918 (69%);GO:0003677 (68%);GO:0005524 (68%) | DNA topoisomerase type II (double strand cut | ATP-hydrolyzing) activity (69%);DNA binding (68%);ATP binding (68%) | GO:0006265 (68%);GO:0007059 (66%) | DNA topological change (68%);chromosome segregation (66%) | IPR013757 (100%);IPR002205 (100%);IPR006691 (100%) | Type IIA DNA topoisomerase subunit A |  |  | 1 | 100.4615 |
| EPMTAVR | Bacteria | Bacteria |  |  |  |  |  |  |  |  |  |  |  |  |  |  |  |  |  |  |  |  |  |  |  |  |  |  | Bacteria;;;;;;;;;;;Bacteria |  | 5.-.-.- | Isomerases (100%) | GO:0005887 (40%);GO:0005886 (20%);GO:0016021 (20%) | integral component of plasma membrane (40%);plasma membrane (20%);integral component of membrane (20%) | GO:0015648 (40%);GO:0016853 (40%) | lipid-linked peptidoglycan transporter activity (40%);isomerase activity (40%) | GO:0008360 (60%);GO:0009252 (60%);GO:0071555 (60%) | regulation of cell shape (60%);peptidoglycan biosynthetic process (60%);cell wall organization (60%) | IPR004268 (50%);IPR007400 (50%) | Peptidoglycan biosynthesis protein MurJ (50%);PrpF protein (50%) |  |  |  | 1 | 1 |
| EQEVAQEDSAPASS | Chlamydia pneumoniae | Bacteria |  |  |  | Chlamydiae |  |  | Chlamydiia |  |  |  | Chlamydiales |  |  |  |  | Chlamydiaceae |  |  |  | Chlamydia |  |  |  | Chlamydia pneumoniae |  |  | Bacteria;Chlamydiae;;;Chlamydiia;Chlamydiales;Chlamydiaceae;;Chlamydia;;Chlamydia pneumoniae;Chlamydia pneumoniae |  |  |  | GO:0016021 (100%);GO:0005886 (50%) | integral component of membrane (100%);plasma membrane (50%) | GO:0005471 (100%);GO:0005524 (100%) | ATP:ADP antiporter activity (100%);ATP binding (100%) |  |  | IPR004667 (100%);IPR036259 (100%) | ADP/ATP carrier protein (100%);MFS transporter superfamily (100%) |  |  |  | 1 | 100.6154 |
| ESAAGHDSSTQGLVAALSK | Rhizobiaceae | Bacteria |  |  |  | Proteobacteria |  |  | Alphaproteobacteria |  |  |  | Rhizobiales |  |  |  |  | Rhizobiaceae |  |  |  |  |  |  |  |  |  |  | Bacteria;Proteobacteria;;;Alphaproteobacteria;Rhizobiales;Rhizobiaceae;;;;;Rhizobiaceae | [K01810](https://www.genome.jp/entry/K01810) | 5.3.1.9 | Glucose-6-phosphate isomerase (100%) | GO:0005737 (100%) | cytoplasm (100%) | GO:0004347 (100%) | glucose-6-phosphate isomerase activity (100%) | GO:0006094 (100%);GO:0006096 (100%) | gluconeogenesis (100%);glycolytic process (100%) | IPR001672 (100%);IPR018189 (100%);IPR023096 (100%) | Phosphoglucose isomerase (PGI) (100%);Phosphoglucose isomerase | conserved site (100%);Phosphoglucose isomerase |  |  | 1 | 100.6923 |
| ESMFSSDGTATR | Mycobacterium | Bacteria |  |  |  | Actinobacteria |  |  | Actinobacteria |  |  |  | Corynebacteriales |  |  |  |  | Mycobacteriaceae |  |  |  | Mycobacterium |  |  |  |  |  |  | Bacteria;Actinobacteria;;;Actinobacteria;Corynebacteriales;Mycobacteriaceae;;Mycobacterium;;;Mycobacterium |  |  |  | GO:0016021 (95%);GO:0005886 (10%);GO:0005887 (5%) | integral component of membrane (95%);plasma membrane (10%);integral component of plasma membrane (5%) |  |  | GO:0006869 (10%);GO:0071555 (10%);GO:0006855 (5%) | lipid transport (10%);cell wall organization (10%);drug transmembrane transport (5%) | IPR004869 (100%) | Membrane transport protein MMPL domain (100%) |  |  |  | 1 | 100.8462 |
| ETFASQVEK | Bacillus | Bacteria |  |  |  | Firmicutes |  |  | Bacilli |  |  |  | Bacillales |  |  |  |  | Bacillaceae |  |  |  | Bacillus |  |  |  |  |  |  | Bacteria;Firmicutes;;;Bacilli;Bacillales;Bacillaceae;;Bacillus;;;Bacillus |  |  |  | GO:0016021 (89%);GO:0005623 (85%) | integral component of membrane (89%);cell (85%) | GO:0016491 (4%);GO:0016853 (4%) | oxidoreductase activity (4%);isomerase activity (4%) | GO:0045454 (85%);GO:0030420 (4%) | cell redox homeostasis (85%);establishment of competence for transformation (4%) | IPR012336 (100%);IPR013766 (100%);IPR036249 (100%) | Thioredoxin-like fold (100%);Thioredoxin domain (100%);Thioredoxin-like superfamily (100%) |  |  |  | 1 | 100.6154 |
| EVLGANSR | Actinobacteria | Bacteria |  |  |  | Actinobacteria |  |  | Actinobacteria |  |  |  |  |  |  |  |  |  |  |  |  |  |  |  |  |  |  |  | Bacteria;Actinobacteria;;;Actinobacteria;;;;;;;Actinobacteria |  |  |  |  |  | GO:0016787 (50%) | hydrolase activity (50%) | GO:0017000 (50%) | antibiotic biosynthetic process (50%) | IPR023393 (89%);IPR019587 (78%);IPR000073 (11%) | START-like domain superfamily (89%);Polyketide cyclase/dehydrase (78%);Alpha/beta hydrolase fold-1 (11%) |  |  |  | 1 | 1 |
| EWEPVDGGTLVVVEGPR | Streptomyces | Bacteria |  |  |  | Actinobacteria |  |  | Actinobacteria |  |  |  | Streptomycetales |  |  |  |  | Streptomycetaceae |  |  |  | Streptomyces |  |  |  |  |  |  | Bacteria;Actinobacteria;;;Actinobacteria;Streptomycetales;Streptomycetaceae;;Streptomyces;;;Streptomyces | [K03783](https://www.genome.jp/entry/K03783) | 2.4.2.1 | Purine-nucleoside phosphorylase (100%) | GO:0005737 (6%);GO:0005829 (6%) | cytoplasm (6%);cytosol (6%) | GO:0017061 (88%);GO:0004731 (18%) | S-methyl-5-thioadenosine phosphorylase activity (88%);purine-nucleoside phosphorylase activity (18%) | GO:0006166 (94%);GO:0019509 (6%) | purine ribonucleoside salvage (94%);L-methionine salvage from methylthioadenosine (6%) | IPR000845 (100%);IPR010044 (100%);IPR035994 (100%) | Nucleoside phosphorylase domain (100%);Methylthioadenosine phosphorylase (MTAP) (100%);Nucleoside phosphorylase superfamily (100%) |  |  |  | 1 | 100.5308 |
| FAEYANTLAAMAK | Bacteria | Bacteria |  |  |  |  |  |  |  |  |  |  |  |  |  |  |  |  |  |  |  |  |  |  |  |  |  |  | Bacteria;;;;;;;;;;;Bacteria | [K14459](https://www.genome.jp/dbget-bin/www_bget?K14459+3.2.1.52+R09323) | 3.2.1.52 | Beta-N-acetylhexosaminidase (100%) | GO:0005576 (98%);GO:0005618 (90%);GO:0016020 (87%) | extracellular region (98%);cell wall (90%);membrane (87%) | GO:0004563 (65%);GO:0102148 (65%);GO:0004553 (31%) | beta-N-acetylhexosaminidase activity (65%);N-acetyl-beta-D-galactosaminidase activity (65%);hydrolase activity | hydrolyzing O-glycosyl compounds (31%) | GO:0005975 (93%);GO:0008152 (4%) | carbohydrate metabolic process (93%);metabolic process (4%) | IPR017853 (100%);IPR011098 (98%);IPR015883 (98%) | Glycoside hydrolase superfamily (100%);G5 domain (98%);Glycoside hydrolase family 20 |  |  | 1 | 100.6154 |
| FAPDPELWPR | Streptomycetaceae | Bacteria |  |  |  | Actinobacteria |  |  | Actinobacteria |  |  |  | Streptomycetales |  |  |  |  | Streptomycetaceae |  |  |  |  |  |  |  |  |  |  | Bacteria;Actinobacteria;;;Actinobacteria;Streptomycetales;Streptomycetaceae;;;;;Streptomycetaceae |  | 3.6.3.16 | Transferred entry: 7.3.2.7 (100%) |  |  | GO:0005524 (100%);GO:0016887 (97%) | ATP binding (100%);ATPase activity (97%) |  |  | IPR008978 (100%);IPR040612 (100%);IPR016300 (97%) | HSP20-like chaperone (100%);ArsA | HSP20-like domain (100%);Arsenical pump ATPase |  |  | 1 | 100.7769 |
| FEQEQADFFMR | Neisseria | Bacteria |  |  |  | Proteobacteria |  |  | Betaproteobacteria |  |  |  | Neisseriales |  |  |  |  | Neisseriaceae |  |  |  | Neisseria |  |  |  |  |  |  | Bacteria;Proteobacteria;;;Betaproteobacteria;Neisseriales;Neisseriaceae;;Neisseria;;;Neisseria | [K00943](https://www.genome.jp/entry/K00943) | 2.7.4.9 | dTMP kinase (100%) | GO:0005737 (3%);GO:0005829 (3%) | cytoplasm (3%);cytosol (3%) | GO:0004798 (100%);GO:0005524 (92%);GO:0009041 (3%) | thymidylate kinase activity (100%);ATP binding (92%);uridylate kinase activity (3%) | GO:0006233 (92%);GO:0006235 (92%);GO:0006227 (3%) | dTDP biosynthetic process (92%);dTTP biosynthetic process (92%);dUDP biosynthetic process (3%) | IPR027417 (100%);IPR039430 (100%);IPR018094 (95%) | P-loop containing nucleoside triphosphate hydrolase (100%);Thymidylate kinase-like domain (100%);Thymidylate kinase (95%) |  |  |  | 1 | 100.6154 |
| FFEVGDVK | Bacteria | Bacteria |  |  |  |  |  |  |  |  |  |  |  |  |  |  |  |  |  |  |  |  |  |  |  |  |  |  | Bacteria;;;;;;;;;;;Bacteria | [K00705](https://www.genome.jp/entry/K00705) | 2.4.1.25 | 4-alpha-glucanotransferase (100%) | GO:0005737 (4%) | cytoplasm (4%) | GO:0004134 (100%);GO:0102500 (100%) | 4-alpha-glucanotransferase activity (100%);beta-maltose 4-alpha-glucanotransferase activity (100%) |  |  | IPR003385 (100%);IPR017853 (100%) | Glycoside hydrolase | family 77 (100%);Glycoside hydrolase superfamily (100%) |  |  | 1 | 100.3 |
| FGGDSLAETQR | Bacteria | Bacteria |  |  |  |  |  |  |  |  |  |  |  |  |  |  |  |  |  |  |  |  |  |  |  |  |  |  | Bacteria;;;;;;;;;;;Bacteria | [K01736](https://www.genome.jp/entry/K01736) | 4.2.3.5 | Chorismate synthase (100%) | GO:0005829 (5%) | cytosol (5%) | GO:0004107 (100%);GO:0010181 (5%);GO:0016651 (5%) | chorismate synthase activity (100%);FMN binding (5%);oxidoreductase activity | acting on NAD(P)H (5%) | GO:0009073 (100%);GO:0009423 (85%) | aromatic amino acid family biosynthetic process (100%);chorismate biosynthetic process (85%) | IPR035904 (100%);IPR000453 (95%);IPR020541 (95%) | Chorismate synthase AroC superfamily (100%);Chorismate synthase (95%);Chorismate synthase |  |  | 1 | 100.3769 |
| FLADAENYVYECMK | Bacillus subtilis group | Bacteria |  |  |  | Firmicutes |  |  | Bacilli |  |  |  | Bacillales |  |  |  |  | Bacillaceae |  |  |  | Bacillus |  | Bacillus subtilis group |  |  |  |  | Bacteria;Firmicutes;;;Bacilli;Bacillales;Bacillaceae;;Bacillus;Bacillus subtilis group;;Bacillus subtilis group |  |  |  |  |  | GO:0016301 (50%);GO:0016740 (50%);GO:0004413 (8%) | kinase activity (50%);transferase activity (50%);homoserine kinase activity (8%) | GO:0009088 (8%) | threonine biosynthetic process (8%) | IPR002575 (100%);IPR011009 (100%) | Aminoglycoside phosphotransferase (100%);Protein kinase-like domain superfamily (100%) |  |  |  | 1 | 100.5462 |
| FLLEFWK | Bacteria | Bacteria |  |  |  |  |  |  |  |  |  |  |  |  |  |  |  |  |  |  |  |  |  |  |  |  |  |  | Bacteria;;;;;;;;;;;Bacteria | [K13292](https://www.genome.jp/dbget-bin/www_bget?ko:K13292) | 2.5.1.145 | Phosphatidylglycerol--prolipoprotein diacylglyceryl transferase (100%) | GO:0005887 (100%) | integral component of plasma membrane (100%) | GO:0008961 (100%) | phosphatidylglycerol-prolipoprotein diacylglyceryl transferase activity (100%) | GO:0042158 (100%) | lipoprotein biosynthetic process (100%) | IPR001640 (90%);IPR000160 (10%);IPR001633 (10%) | Phosphatidylglycerol--prolipoprotein diacylglyceryl transferase Lgt (90%);GGDEF domain (10%);EAL domain (10%) |  |  |  | 1 | 100.7 |
| FNAFEAAK | Campylobacter | Bacteria |  |  |  | Proteobacteria | delta/epsilon subdivisions |  | Epsilonproteobacteria |  |  |  | Campylobacterales |  |  |  |  | Campylobacteraceae |  |  |  | Campylobacter |  |  |  |  |  |  | Bacteria;Proteobacteria;delta/epsilon subdivisions;;Epsilonproteobacteria;Campylobacterales;Campylobacteraceae;;Campylobacter;;;Campylobacter | [K15633](https://www.kegg.jp/dbget-bin/www_bget?K15633+K15634+K15635+5.4.2.12+R01518) | 5.4.2.12 | Phosphoglycerate mutase (2 | 3-diphosphoglycerate-independent) (100%) | GO:0005737 (93%) | cytoplasm (93%) | GO:0030145 (99%);GO:0046537 (98%);GO:0004619 (2%) | manganese ion binding (99%);2 | 3-bisphosphoglycerate-independent phosphoglycerate mutase activity (98%);phosphoglycerate mutase activity (2%) | GO:0006096 (98%);GO:0006007 (94%) | glycolytic process (98%);glucose catabolic process (94%) | IPR005995 (100%);IPR006124 (100%);IPR017850 (100%) |  |  | 1 | 100.6923 |
| FNLFNSSR | Bacteria | Bacteria |  |  |  |  |  |  |  |  |  |  |  |  |  |  |  |  |  |  |  |  |  |  |  |  |  |  | Bacteria;;;;;;;;;;;Bacteria | [K00372](https://www.genome.jp/dbget-bin/www_bget?K00372+K00360+1.7.99.4+R00798) | 1.7.99.4 ;1.7.5.1 (0%) | Nitrate reductase (100%);Nitrate reductase (quinone) (0%) | GO:0009325 (71%);GO:0016020 (0%);GO:0005886 (0%) | nitrate reductase complex (71%);membrane (0%);plasma membrane (0%) | GO:0008940 (100%);GO:0009055 (0%);GO:0046872 (0%) | nitrate reductase activity (100%);electron transfer activity (0%);metal ion binding (0%) | GO:0042126 (71%);GO:0009061 (0%);GO:0006974 (0%) | nitrate metabolic process (71%);anaerobic respiration (0%);cellular response to DNA damage stimulus (0%) | IPR029263 (100%);IPR038262 (98%);IPR017896 (98%) | Respiratory nitrate reductase beta | C-terminal (100%);Nitrate reductase beta |  |  | 1 | 100.4462 |
| FPMVVQEVSK | Brucella | Bacteria |  |  |  | Proteobacteria |  |  | Alphaproteobacteria |  |  |  | Rhizobiales |  |  |  |  | Brucellaceae |  |  |  | Brucella |  |  |  |  |  |  | Bacteria;Proteobacteria;;;Alphaproteobacteria;Rhizobiales;Brucellaceae;;Brucella;;;Brucella |  |  |  |  |  |  |  | GO:0006865 (100%) | amino acid transport (100%) | IPR028081 (100%);IPR028082 (100%);IPR000709 (88%) | Leucine-binding protein domain (100%);Periplasmic binding protein-like I (100%);Leu/Ile/Val-binding protein (88%) |  |  |  | 1 | 100.5385 |
| FSETNDDFFAK | Mycoplasma pneumoniae | Bacteria |  |  |  | Tenericutes |  |  | Mollicutes |  |  |  | Mycoplasmatales |  |  |  |  | Mycoplasmataceae |  |  |  | Mycoplasma |  |  |  | Mycoplasma pneumoniae |  |  | Bacteria;Tenericutes;;;Mollicutes;Mycoplasmatales;Mycoplasmataceae;;Mycoplasma;;Mycoplasma pneumoniae;Mycoplasma pneumoniae |  |  |  | GO:0005886 (100%) | plasma membrane (100%) |  |  |  |  | IPR022186 (100%) | Protein of unknown function DUF3713 (100%) |  |  |  | 1 | 100.9231 |
| FSQAALCEVWESLQDR | Chlorobium phaeobacteroides | Bacteria |  |  |  | Chlorobi |  |  | Chlorobia |  |  |  | Chlorobiales |  |  |  |  | Chlorobiaceae |  |  |  | Chlorobium |  |  |  | Chlorobium phaeobacteroides |  |  | Bacteria;Chlorobi;;;Chlorobia;Chlorobiales;Chlorobiaceae;;Chlorobium;;Chlorobium phaeobacteroides;Chlorobium phaeobacteroides |  |  |  |  |  |  |  |  |  | IPR001453 (100%);IPR008135 (100%);IPR008136 (100%) | MoaB/Mog domain (100%);Competence-induced protein CinA (100%);CinA | C-terminal (100%) |  |  | 1 | 1 |
| FSTVAGEAGAADAER | Bacteria | Bacteria |  |  |  |  |  |  |  |  |  |  |  |  |  |  |  |  |  |  |  |  |  |  |  |  |  |  | Bacteria;;;;;;;;;;;Bacteria | [K03781](https://www.genome.jp/dbget-bin/www_bget?ko:K03781) | 1.11.1.6 | Catalase (100%) |  |  | GO:0004096 (97%);GO:0046872 (94%);GO:0020037 (90%) | catalase activity (97%);metal ion binding (94%);heme binding (90%) | GO:0006979 (90%);GO:0042744 (86%) | response to oxidative stress (90%);hydrogen peroxide catabolic process (86%) | IPR011614 (100%);IPR018028 (100%);IPR020835 (100%) | Catalase core domain (100%);Catalase | mono-functional |  |  | 1 | 1 |
| FTTLTGLSVF | Bacteria | Bacteria |  |  |  |  |  |  |  |  |  |  |  |  |  |  |  |  |  |  |  |  |  |  |  |  |  |  | Bacteria;;;;;;;;;;;Bacteria | K01425 | 3.5.1.2 | Glutaminase (100%) |  |  | GO:0004359 (100%) | glutaminase activity (100%) | GO:0006541 (93%);GO:0006537 (0%);GO:0006543 (0%) | glutamine metabolic process (93%);glutamate biosynthetic process (0%);glutamine catabolic process (0%) | IPR012338 (100%);IPR015868 (100%) | Beta-lactamase/transpeptidase-like (100%);Glutaminase (100%) |  |  |  | 1 | 1 |
| GAEDELNR | Firmicutes | Bacteria |  |  |  | Firmicutes |  |  |  |  |  |  |  |  |  |  |  |  |  |  |  |  |  |  |  |  |  |  | Bacteria;Firmicutes;;;;;;;;;;Firmicutes |  |  |  | GO:0005737 (4%) | cytoplasm (4%) | GO:0003677 (93%);GO:0000976 (4%);GO:0003688 (4%) | DNA binding (93%);transcription regulatory region sequence-specific DNA binding (4%);DNA replication origin binding (4%) | GO:0006355 (81%);GO:0006270 (4%);GO:0006275 (4%) | regulation of transcription | DNA-templated (81%);DNA replication initiation (4%);regulation of DNA replication (4%) | IPR000843 (96%);IPR010982 (96%);IPR028082 (96%) | LacI-type HTH domain (96%);Lambda repressor-like |  |  | 1 | 100.9231 |
| GATVLNSTK | Bacteria | Bacteria |  |  |  |  |  |  |  |  |  |  |  |  |  |  |  |  |  |  |  |  |  |  |  |  |  |  | Bacteria;;;;;;;;;;;Bacteria |  |  |  | GO:0005737 (100%) | cytoplasm (100%) | GO:0003746 (100%);GO:0003924 (100%);GO:0005525 (100%) | translation elongation factor activity (100%);GTPase activity (100%);GTP binding (100%) |  |  | IPR000640 (100%);IPR000795 (100%);IPR004161 (100%) | Elongation factor EFG | domain V-like (100%);Transcription factor |  |  | 1 | 100.4462 |
| GAVNDLLAK | Bacteria | Bacteria |  |  |  |  |  |  |  |  |  |  |  |  |  |  |  |  |  |  |  |  |  |  |  |  |  |  | Bacteria;;;;;;;;;;;Bacteria |  |  |  | GO:0005576 (99%);GO:0016021 (99%);GO:0020002 (99%) | extracellular region (99%);integral component of membrane (99%);host cell plasma membrane (99%) | GO:0090729 (99%);GO:0015485 (96%);GO:0008289 (4%) | toxin activity (99%);cholesterol binding (96%);lipid binding (4%) | GO:0044179 (99%);GO:0009405 (96%) | hemolysis in other organism (99%);pathogenesis (96%) | IPR001869 (100%);IPR036359 (100%);IPR036363 (99%) | Thiol-activated cytolysin (100%);Thiol-activated cytolysin superfamily (100%);Thiol-activated cytolysin | alpha-beta domain superfamily (99%) |  |  | 1 | 100.9231 |
| GDGGNSWGNQMR | Bacteria | Bacteria |  |  |  |  |  |  |  |  |  |  |  |  |  |  |  |  |  |  |  |  |  |  |  |  |  |  | Bacteria;;;;;;;;;;;Bacteria |  |  |  | GO:0005737 (100%) | cytoplasm (100%) | GO:0016149 (100%);GO:0003747 (0%);GO:0004674 (0%) | translation release factor activity | codon specific (100%);translation release factor activity (0%);protein serine/threonine kinase activity (0%) |  |  | IPR000352 (100%);IPR004374 (100%);IPR005139 (100%) | Peptide chain release factor class I (100%);Peptide chain release factor 2 (100%);Peptide chain release factor (100%) |  |  | 1 | 100.4615 |
| GEGDDSEGK | Thermotoga | Bacteria |  |  |  | Thermotogae |  |  | Thermotogae |  |  |  | Thermotogales |  |  |  |  | Thermotogaceae |  |  |  | Thermotoga |  |  |  |  |  |  | Bacteria;Thermotogae;;;Thermotogae;Thermotogales;Thermotogaceae;;Thermotoga;;;Thermotoga | [K01802](https://www.genome.jp/entry/K01802) | 5.2.1.8 | Peptidylprolyl isomerase (100%) | GO:0005737 (100%) | cytoplasm (100%) | GO:0003677 (100%);GO:0003755 (100%);GO:0043022 (13%) | DNA binding (100%);peptidyl-prolyl cis-trans isomerase activity (100%);ribosome binding (13%) | GO:0007049 (100%);GO:0015031 (100%);GO:0032784 (100%) | cell cycle (100%);protein transport (100%);regulation of DNA-templated transcription | elongation (100%) | IPR005215 (100%);IPR008880 (100%);IPR008881 (100%) | Trigger factor (100%);Trigger factor |  |  | 1 | 100.7692 |
| GENVAVR | Bacteria | Bacteria |  |  |  |  |  |  |  |  |  |  |  |  |  |  |  |  |  |  |  |  |  |  |  |  |  |  | Bacteria;;;;;;;;;;;Bacteria |  | 1.3.1.- (99%);1.-.-.- (0%);6.2.1.- (0%) | With NAD(+) or NADP(+) as acceptor (99%);Oxidoreductases (0%);Acid--thiol ligases (0%) |  |  | GO:0017150 (88%);GO:0050660 (88%);GO:0016491 (7%) | tRNA dihydrouridine synthase activity (88%);flavin adenine dinucleotide binding (88%);oxidoreductase activity (7%) | GO:0008033 (5%);GO:0006355 (0%) | tRNA processing (5%);regulation of transcription | DNA-templated (0%) | IPR024036 (94%);IPR001269 (93%);IPR013785 (92%) | tRNA-dihydrouridine synthase |  |  | 1 | 1 |
| GEVYFPMEK | Actinobacteria | Bacteria |  |  |  | Actinobacteria |  |  | Actinobacteria |  |  |  |  |  |  |  |  |  |  |  |  |  |  |  |  |  |  |  | Bacteria;Actinobacteria;;;Actinobacteria;;;;;;;Actinobacteria | [K01972](https://www.genome.jp/dbget-bin/www_bget?K01972) | 6.5.1.2 | DNA ligase (NAD(+)) (100%) | GO:0005829 (0%) | cytosol (0%) | GO:0003911 (100%);GO:0046872 (98%);GO:0051213 (9%) | DNA ligase (NAD+) activity (100%);metal ion binding (98%);dioxygenase activity (9%) | GO:0006260 (98%);GO:0006281 (98%);GO:0006288 (0%) | DNA replication (98%);DNA repair (98%);base-excision repair | DNA ligation (0%) | IPR013839 (100%);IPR013840 (100%);IPR018239 (100%) | NAD-dependent DNA ligase |  |  | 1 | 100.8462 |
| GFCAVENLK | Campylobacter | Bacteria |  |  |  | Proteobacteria | delta/epsilon subdivisions |  | Epsilonproteobacteria |  |  |  | Campylobacterales |  |  |  |  | Campylobacteraceae |  |  |  | Campylobacter |  |  |  |  |  |  | Bacteria;Proteobacteria;delta/epsilon subdivisions;;Epsilonproteobacteria;Campylobacterales;Campylobacteraceae;;Campylobacter;;;Campylobacter | [K00762](https://www.genome.jp/entry/K00762) | 2.4.2.10 | Orotate phosphoribosyltransferase (100%) |  |  | GO:0004588 (98%);GO:0000287 (96%);GO:0016757 (2%) | orotate phosphoribosyltransferase activity (98%);magnesium ion binding (96%);transferase activity | transferring glycosyl groups (2%) | GO:0044205 (98%);GO:0009116 (91%);GO:0019856 (91%) | 'de novo' UMP biosynthetic process (98%);nucleoside metabolic process (91%);pyrimidine nucleobase biosynthetic process (91%) | IPR029057 (100%);IPR000836 (98%);IPR006273 (98%) | Phosphoribosyltransferase-like (100%);Phosphoribosyltransferase domain (98%);Orotate phosphoribosyltransferase |  |  | 1 | 1 |
| GFDVAAMFADAVAAWSVR | Neisseria | Bacteria |  |  |  | Proteobacteria |  |  | Betaproteobacteria |  |  |  | Neisseriales |  |  |  |  | Neisseriaceae |  |  |  | Neisseria |  |  |  |  |  |  | Bacteria;Proteobacteria;;;Betaproteobacteria;Neisseriales;Neisseriaceae;;Neisseria;;;Neisseria | [K01920](https://www.genome.jp/entry/K01920) | 6.3.2.3 | Glutathione synthase (100%) | GO:0005737 (3%) | cytoplasm (3%) | GO:0004363 (100%);GO:0005524 (100%);GO:0046872 (94%) | glutathione synthase activity (100%);ATP binding (100%);metal ion binding (94%) |  |  | IPR004218 (100%);IPR006284 (100%);IPR004215 (97%) | Prokaryotic glutathione synthetase | ATP-binding (100%);Glutathione synthetase |  |  | 1 | 100.4769 |
| GFEALDDTGK | Mycoplasma pneumoniae | Bacteria |  |  |  | Tenericutes |  |  | Mollicutes |  |  |  | Mycoplasmatales |  |  |  |  | Mycoplasmataceae |  |  |  | Mycoplasma |  |  |  | Mycoplasma pneumoniae |  |  | Bacteria;Tenericutes;;;Mollicutes;Mycoplasmatales;Mycoplasmataceae;;Mycoplasma;;Mycoplasma pneumoniae;Mycoplasma pneumoniae |  |  |  | GO:0016021 (100%);GO:0005886 (33%) | integral component of membrane (100%);plasma membrane (33%) |  |  |  |  |  |  |  |  |  | 1 | 100.6154 |
| GGANTDHHAAPR | Mycobacterium tuberculosis | Bacteria |  |  |  | Actinobacteria |  |  | Actinobacteria |  |  |  | Corynebacteriales |  |  |  |  | Mycobacteriaceae |  |  |  | Mycobacterium |  | Mycobacterium tuberculosis complex |  | Mycobacterium tuberculosis |  |  | Bacteria;Actinobacteria;;;Actinobacteria;Corynebacteriales;Mycobacteriaceae;;Mycobacterium;Mycobacterium tuberculosis complex;Mycobacterium tuberculosis;Mycobacterium tuberculosis | [K00936](https://www.genome.jp/entry/K00936) | 2.7.13.3 | Histidine kinase (100%) |  |  | GO:0004673 (100%);GO:0005524 (100%) | protein histidine kinase activity (100%);ATP binding (100%) | GO:0000160 (100%) | phosphorelay signal transduction system (100%) | IPR003594 (100%);IPR004358 (100%);IPR005467 (100%) | Histidine kinase/HSP90-like ATPase (100%);Signal transduction histidine kinase-related protein | C-terminal (100%);Histidine kinase domain (100%) |  |  | 1 | 100.9231 |
| GGEADEN | Firmicutes | Bacteria |  |  |  | Firmicutes |  |  |  |  |  |  |  |  |  |  |  |  |  |  |  |  |  |  |  |  |  |  | Bacteria;Firmicutes;;;;;;;;;;Firmicutes |  |  |  | GO:0005623 (50%);GO:0005840 (50%);GO:0016021 (50%) | cell (50%);ribosome (50%);integral component of membrane (50%) | GO:0000049 (50%);GO:0003735 (50%);GO:0008137 (50%) | tRNA binding (50%);structural constituent of ribosome (50%);NADH dehydrogenase (ubiquinone) activity (50%) | GO:0006412 (50%);GO:0030004 (50%);GO:0042773 (50%) | translation (50%);cellular monovalent inorganic cation homeostasis (50%);ATP synthesis coupled electron transport (50%) | IPR000114 (50%);IPR001516 (50%);IPR001750 (50%) | Ribosomal protein L16 (50%);NADH-Ubiquinone oxidoreductase (complex I) | chain 5 N-terminal (50%);NADH:quinone oxidoreductase/Mrp antiporter |  |  | 1 | 100.4615 |
| GGHGSAPEK | Proteobacteria | Bacteria |  |  |  | Proteobacteria |  |  |  |  |  |  |  |  |  |  |  |  |  |  |  |  |  |  |  |  |  |  | Bacteria;Proteobacteria;;;;;;;;;;Proteobacteria | [K01451](https://www.genome.jp/entry/K01451) | 3.-.-.- (63%);3.5.1.32 (38%) | Hydrolases (63%);Hippurate hydrolase (38%) |  |  | GO:0016787 (96%);GO:0047980 (4%) | hydrolase activity (96%);hippurate hydrolase activity (4%) |  |  | IPR011650 (92%);IPR036264 (92%);IPR002933 (88%) | Peptidase M20 | dimerisation domain (92%);Bacterial exopeptidase dimerisation domain (92%);Peptidase M20 (88%) |  |  | 1 | 100.2231 |
| GGQEMKPR | Proteobacteria | Bacteria |  |  |  | Proteobacteria |  |  |  |  |  |  |  |  |  |  |  |  |  |  |  |  |  |  |  |  |  |  | Bacteria;Proteobacteria;;;;;;;;;;Proteobacteria |  |  |  | GO:0016021 (98%) | integral component of membrane (98%) |  |  | GO:0000746 (2%) | conjugation (2%) | IPR020065 (100%);IPR024475 (100%) | Conjugal transfer protein TrbK (100%);Type IV conjugative transfer protein TrbJ/K | C-terminal (100%) |  |  | 1 | 100.4615 |
| GGSPFGGFEDAFGGGGGGGAR | Chlorobium chlorochromatii | Bacteria |  |  |  | Chlorobi |  |  | Chlorobia |  |  |  | Chlorobiales |  |  |  |  | Chlorobiaceae |  |  |  | Chlorobium |  |  |  | Chlorobium chlorochromatii |  |  | Bacteria;Chlorobi;;;Chlorobia;Chlorobiales;Chlorobiaceae;;Chlorobium;;Chlorobium chlorochromatii;Chlorobium chlorochromatii |  |  |  | GO:0005737 (100%) | cytoplasm (100%) | GO:0005524 (100%);GO:0008270 (100%);GO:0031072 (100%) | ATP binding (100%);zinc ion binding (100%);heat shock protein binding (100%) | GO:0006260 (100%);GO:0006457 (100%);GO:0009408 (100%) | DNA replication (100%);protein folding (100%);response to heat (100%) | IPR001305 (100%);IPR001623 (100%);IPR002939 (100%) | Heat shock protein DnaJ | cysteine-rich domain (100%);DnaJ domain (100%);Chaperone DnaJ |  |  | 1 | 100.7692 |
| GGSYAPGTPR | Bacteria | Bacteria |  |  |  |  |  |  |  |  |  |  |  |  |  |  |  |  |  |  |  |  |  |  |  |  |  |  | Bacteria;;;;;;;;;;;Bacteria | [K01118](https://www.genome.jp/entry/K01118) | 1.7.1.17 | FMN-dependent NADH-azoreductase (100%) |  |  | GO:0009055 (100%);GO:0010181 (100%);GO:0016652 (100%) | electron transfer activity (100%);FMN binding (100%);oxidoreductase activity | acting on NAD(P)H | NAD(P) as acceptor (100%) |  |  | IPR003680 (100%);IPR029039 (100%);IPR023048 (100%) |  |  | 1 | 100.8462 |
| GHATKPR | Mycobacterium tuberculosis complex | Bacteria |  |  |  | Actinobacteria |  |  | Actinobacteria |  |  |  | Corynebacteriales |  |  |  |  | Mycobacteriaceae |  |  |  | Mycobacterium |  | Mycobacterium tuberculosis complex |  |  |  |  | Bacteria;Actinobacteria;;;Actinobacteria;Corynebacteriales;Mycobacteriaceae;;Mycobacterium;Mycobacterium tuberculosis complex;;Mycobacterium tuberculosis complex |  |  |  | GO:0005737 (100%);GO:0005618 (7%);GO:0005886 (7%) | cytoplasm (100%);cell wall (7%);plasma membrane (7%) | GO:0003677 (100%);GO:0003700 (100%);GO:0016987 (100%) | DNA binding (100%);DNA-binding transcription factor activity (100%);sigma factor activity (100%) | GO:0001123 (100%);GO:0009405 (7%);GO:0009415 (7%) | transcription initiation from bacterial-type RNA polymerase promoter (100%);pathogenesis (7%);response to water (7%) | IPR000943 (100%);IPR007624 (100%);IPR007627 (100%) | RNA polymerase sigma-70 (100%);RNA polymerase sigma-70 region 3 (100%);RNA polymerase sigma-70 region 2 (100%) |  |  |  | 1 | 1 |
| GKPVAVAGNVK | Bacillus | Bacteria |  |  |  | Firmicutes |  |  | Bacilli |  |  |  | Bacillales |  |  |  |  | Bacillaceae |  |  |  | Bacillus |  |  |  |  |  |  | Bacteria;Firmicutes;;;Bacilli;Bacillales;Bacillaceae;;Bacillus;;;Bacillus | K02320 | 2.7.7.7 | DNA-directed DNA polymerase (100%) | GO:0005737 (100%) | cytoplasm (100%) | GO:0000287 (100%);GO:0003887 (100%);GO:0003684 (90%) | magnesium ion binding (100%);DNA-directed DNA polymerase activity (100%);damaged DNA binding (90%) | GO:0006261 (100%);GO:0006281 (97%);GO:0009432 (3%) | DNA-dependent DNA replication (100%);DNA repair (97%);SOS response (3%) | IPR001126 (100%);IPR017961 (100%);IPR022880 (100%) | UmuC domain (100%);DNA polymerase | Y-family |  |  | 1 | 100.6231 |
| GLDAAVEDVPEDR | Enterobacteriaceae | Bacteria |  |  |  | Proteobacteria |  |  | Gammaproteobacteria |  |  |  | Enterobacterales |  |  |  |  | Enterobacteriaceae |  |  |  |  |  |  |  |  |  |  | Bacteria;Proteobacteria;;;Gammaproteobacteria;Enterobacterales;Enterobacteriaceae;;;;;Enterobacteriaceae |  |  |  |  |  |  |  |  |  | IPR011646 (100%);IPR027417 (100%) | KAP family P-loop domain (100%);P-loop containing nucleoside triphosphate hydrolase (100%) |  |  |  | 1 | 100.2308 |
| GLTDGQELTDGPALK | Streptomyces | Bacteria |  |  |  | Actinobacteria |  |  | Actinobacteria |  |  |  | Streptomycetales |  |  |  |  | Streptomycetaceae |  |  |  | Streptomyces |  |  |  |  |  |  | Bacteria;Actinobacteria;;;Actinobacteria;Streptomycetales;Streptomycetaceae;;Streptomyces;;;Streptomyces |  |  |  | GO:0005737 (100%);GO:0009380 (100%) | cytoplasm (100%);excinuclease repair complex (100%) | GO:0003677 (100%);GO:0009381 (100%) | DNA binding (100%);excinuclease ABC activity (100%) | GO:0006289 (100%);GO:0009432 (100%);GO:0006974 (14%) | nucleotide-excision repair (100%);SOS response (100%);cellular response to DNA damage stimulus (14%) | IPR000305 (100%);IPR001162 (100%);IPR001943 (100%) | GIY-YIG endonuclease (100%);UvrC family homology region (100%);UVR domain (100%) |  |  |  | 1 | 100.1615 |
| GNLPEGKPGFK | Bacillus | Bacteria |  |  |  | Firmicutes |  |  | Bacilli |  |  |  | Bacillales |  |  |  |  | Bacillaceae |  |  |  | Bacillus |  |  |  |  |  |  | Bacteria;Firmicutes;;;Bacilli;Bacillales;Bacillaceae;;Bacillus;;;Bacillus |  |  |  | GO:0016020 (50%);GO:0016021 (50%);GO:0005886 (5%) | membrane (50%);integral component of membrane (50%);plasma membrane (5%) | GO:0022857 (95%);GO:0042802 (5%) | transmembrane transporter activity (95%);identical protein binding (5%) |  |  | IPR006143 (100%);IPR032317 (100%) | RND efflux pump | membrane fusion protein (100%);RND efflux pump |  |  | 1 | 100.4538 |
| GPLPEENDALAGNR | Streptomyces | Bacteria |  |  |  | Actinobacteria |  |  | Actinobacteria |  |  |  | Streptomycetales |  |  |  |  | Streptomycetaceae |  |  |  | Streptomyces |  |  |  |  |  |  | Bacteria;Actinobacteria;;;Actinobacteria;Streptomycetales;Streptomycetaceae;;Streptomyces;;;Streptomyces |  | 2.1.1.- | Methyltransferases (100%) |  |  | GO:0008168 (100%) | methyltransferase activity (100%) | GO:0032259 (100%) | methylation (100%) | IPR007213 (100%);IPR011610 (100%);IPR029063 (100%) | Methyltransferase Ppm1/Ppm2/Tcmp (100%);S-adenosyl-L-methionine-dependent methyltransferase | putative (100%);S-adenosyl-L-methionine-dependent methyltransferase (100%) |  |  | 1 | 100.3077 |
| GTHFHPGAGVGR | Actinobacteria | Bacteria |  |  |  | Actinobacteria |  |  | Actinobacteria |  |  |  |  |  |  |  |  |  |  |  |  |  |  |  |  |  |  |  | Bacteria;Actinobacteria;;;Actinobacteria;;;;;;;Actinobacteria |  |  |  | GO:0005840 (100%);GO:0022625 (0%) | ribosome (100%);cytosolic large ribosomal subunit (0%) | GO:0003735 (93%) | structural constituent of ribosome (93%) | GO:0006412 (99%) | translation (99%) | IPR001684 (100%);IPR018261 (100%) | Ribosomal protein L27 (100%);Ribosomal protein L27 | conserved site (100%) |  |  | 1 | 100.4615 |
| GVGSEQAYAK | Corynebacteriales | Bacteria |  |  |  | Actinobacteria |  |  | Actinobacteria |  |  |  | Corynebacteriales |  |  |  |  |  |  |  |  |  |  |  |  |  |  |  | Bacteria;Actinobacteria;;;Actinobacteria;Corynebacteriales;;;;;;Corynebacteriales | [K00248](https://www.genome.jp/entry/K00248+1.3.8.1+R04751) | 1.3.8.1 (50%);1.3.99.- (27%);1.3.8.- (23%) | Short-chain acyl-CoA dehydrogenase (50%);With other acceptors (27%);With a flavin as acceptor (23%) | GO:0005576 (0%);GO:0005886 (0%) | extracellular region (0%);plasma membrane (0%) | GO:0050660 (99%);GO:0016627 (93%);GO:0004085 (4%) | flavin adenine dinucleotide binding (99%);oxidoreductase activity | acting on the CH-CH group of donors (93%);butyryl-CoA dehydrogenase activity (4%) | GO:0052572 (0%) | response to host immune response (0%) | IPR009075 (100%);IPR036250 (100%);IPR006091 (99%) | Acyl-CoA dehydrogenase/oxidase C-terminal (100%);Acyl-CoA dehydrogenase-like |  |  | 1 | 100.7692 |
| GVLLLGRPGTGK | Bacteria | Bacteria |  |  |  |  |  |  |  |  |  |  |  |  |  |  |  |  |  |  |  |  |  |  |  |  |  |  | Bacteria;;;;;;;;;;;Bacteria |  | 3.4.24.- | Metalloendopeptidases (100%) | GO:0005886 (100%);GO:0016021 (100%) | plasma membrane (100%);integral component of membrane (100%) | GO:0005524 (100%);GO:0008270 (100%);GO:0016887 (100%) | ATP binding (100%);zinc ion binding (100%);ATPase activity (100%) | GO:0030163 (100%);GO:0051301 (23%) | protein catabolic process (100%);cell division (23%) | IPR000642 (100%);IPR003593 (100%);IPR003959 (100%) | Peptidase M41 (100%);AAA+ ATPase domain (100%);ATPase | AAA-type |  |  | 1 | 100.5308 |
| GVPVEADAR | Bacteria | Bacteria |  |  |  |  |  |  |  |  |  |  |  |  |  |  |  |  |  |  |  |  |  |  |  |  |  |  | Bacteria;;;;;;;;;;;Bacteria | [K01974](https://www.genome.jp/entry/K01974) | 6.5.1.- (97%);6.5.1.4 (1%);6.5.1.8 (1%) | Forming phosphoric ester bonds (97%);RNA 3'-terminal-phosphate cyclase (ATP) (1%);3'-phosphate/5'-hydroxy nucleic acid ligase (1%) |  |  | GO:0008452 (95%);GO:0046872 (95%);GO:0016874 (4%) | RNA ligase activity (95%);metal ion binding (95%);ligase activity (4%) | GO:0006396 (95%);GO:0006281 (0%);GO:0006974 (0%) | RNA processing (95%);DNA repair (0%);cellular response to DNA damage stimulus (0%) | IPR001233 (100%);IPR036025 (100%);IPR000228 (0%) | RNA-splicing ligase | RtcB (100%);tRNA-splicing ligase RtcB-like superfamily (100%);RNA 3'-terminal phosphate cyclase (0%) |  |  | 1 | 100.9231 |
| GYLNFSSEQDSD | Corynebacterium efficiens | Bacteria |  |  |  | Actinobacteria |  |  | Actinobacteria |  |  |  | Corynebacteriales |  |  |  |  | Corynebacteriaceae |  |  |  | Corynebacterium |  |  |  | Corynebacterium efficiens |  |  | Bacteria;Actinobacteria;;;Actinobacteria;Corynebacteriales;Corynebacteriaceae;;Corynebacterium;;Corynebacterium efficiens;Corynebacterium efficiens | [K09887](https://www.kegg.jp/entry/K09887+3.5.4.30+R07307) | 3.5.4.30 | dCTP deaminase (dUMP-forming) (100%) |  |  | GO:0000166 (100%);GO:0008829 (100%);GO:0033973 (100%) | nucleotide binding (100%);dCTP deaminase activity (100%);dCTP deaminase (dUMP-forming) activity (100%) | GO:0006226 (100%);GO:0006229 (100%) | dUMP biosynthetic process (100%);dUTP biosynthetic process (100%) | IPR011962 (100%);IPR029054 (100%);IPR033704 (100%) | dCTP deaminase (100%);dUTPase-like (100%);dUTPase | trimeric (100%) |  |  | 1 | 100.9231 |
| HAGSTGMR | Bacteria | Bacteria |  |  |  |  |  |  |  |  |  |  |  |  |  |  |  |  |  |  |  |  |  |  |  |  |  |  | Bacteria;;;;;;;;;;;Bacteria | [K00164](https://www.genome.jp/dbget-bin/www_bget?K00164+1.2.4.2+R03316) | 1.2.4.2 | Oxoglutarate dehydrogenase (succinyl-transferring) (100%) | GO:0005840 (57%);GO:0005829 (3%);GO:0022625 (3%) | ribosome (57%);cytosol (3%);cytosolic large ribosomal subunit (3%) | GO:0003735 (60%);GO:0019843 (57%);GO:0004591 (40%) | structural constituent of ribosome (60%);rRNA binding (57%);oxoglutarate dehydrogenase (succinyl-transferring) activity (40%) | GO:0006412 (60%);GO:0006099 (40%) | translation (60%);tricarboxylic acid cycle (40%) | IPR000597 (60%);IPR009000 (60%);IPR019927 (60%) | Ribosomal protein L3 (60%);Translation protein | beta-barrel domain superfamily (60%);Ribosomal protein L3 |  |  | 1 | 1 |
| HDHQEDVTR | Bacillus | Bacteria |  |  |  | Firmicutes |  |  | Bacilli |  |  |  | Bacillales |  |  |  |  | Bacillaceae |  |  |  | Bacillus |  |  |  |  |  |  | Bacteria;Firmicutes;;;Bacilli;Bacillales;Bacillaceae;;Bacillus;;;Bacillus |  |  |  |  |  |  |  |  |  |  |  |  |  |  | 1 | 100.7 |
| HEGPFLAAVEDGPLSATQFHPEK | Mycobacterium avium complex (MAC) | Bacteria |  |  |  | Actinobacteria |  |  | Actinobacteria |  |  |  | Corynebacteriales |  |  |  |  | Mycobacteriaceae |  |  |  | Mycobacterium |  | Mycobacterium avium complex (MAC) |  |  |  |  | Bacteria;Actinobacteria;;;Actinobacteria;Corynebacteriales;Mycobacteriaceae;;Mycobacterium;Mycobacterium avium complex (MAC);;Mycobacterium avium complex (MAC) | [K01663](https://www.genome.jp/entry/K01663) | 4.3.2.10 ;3.5.1.2 (94%) | Imidazole glycerol-phosphate synthase (100%);Glutaminase (94%) | GO:0005737 (100%) | cytoplasm (100%) | GO:0004359 (100%);GO:0016829 (100%);GO:0000107 (94%) | glutaminase activity (100%);lyase activity (100%);imidazoleglycerol-phosphate synthase activity (94%) | GO:0000105 (100%);GO:0006541 (100%) | histidine biosynthetic process (100%);glutamine metabolic process (100%) | IPR010139 (100%);IPR017926 (100%);IPR029062 (100%) | Imidazole glycerol phosphate synthase | subunit H (100%);Glutamine amidotransferase (100%);Class I glutamine amidotransferase-like (100%) |  |  | 1 | 1 |
| HHLTATGAVAR | Clostridiales | Bacteria |  |  |  | Firmicutes |  |  | Clostridia |  |  |  | Clostridiales |  |  |  |  |  |  |  |  |  |  |  |  |  |  |  | Bacteria;Firmicutes;;;Clostridia;Clostridiales;;;;;;Clostridiales |  |  |  | GO:0016021 (3%) | integral component of membrane (3%) | GO:0003677 (100%);GO:0004803 (100%) | DNA binding (100%);transposase activity (100%) | GO:0006313 (100%) | transposition | DNA-mediated (100%) | IPR003346 (100%);IPR002525 (78%) | Transposase |  |  | 1 | 100.2231 |
| HLFLGSLR | Bacillaceae | Bacteria |  |  |  | Firmicutes |  |  | Bacilli |  |  |  | Bacillales |  |  |  |  | Bacillaceae |  |  |  |  |  |  |  |  |  |  | Bacteria;Firmicutes;;;Bacilli;Bacillales;Bacillaceae;;;;;Bacillaceae |  |  |  | GO:0016021 (100%);GO:0019867 (100%) | integral component of membrane (100%);outer membrane (100%) | GO:0004553 (100%) | hydrolase activity | hydrolyzing O-glycosyl compounds (100%) | GO:0009254 (100%) | peptidoglycan turnover (100%) | IPR012543 (98%);IPR029064 (98%);IPR007137 (2%) | Protein of unknown function DUF1694 (98%);50S ribosomal protein L30e-like (98%);Resuscitation-promoting factor |  |  | 1 | 1 |
| HPYCALPQSAR | Mycobacterium | Bacteria |  |  |  | Actinobacteria |  |  | Actinobacteria |  |  |  | Corynebacteriales |  |  |  |  | Mycobacteriaceae |  |  |  | Mycobacterium |  |  |  |  |  |  | Bacteria;Actinobacteria;;;Actinobacteria;Corynebacteriales;Mycobacteriaceae;;Mycobacterium;;;Mycobacterium |  |  |  | GO:0005887 (96%);GO:0033281 (96%);GO:0016021 (4%) | integral component of plasma membrane (96%);TAT protein transport complex (96%);integral component of membrane (4%) | GO:0008320 (94%);GO:0009977 (2%) | protein transmembrane transporter activity (94%);proton motive force dependent protein transmembrane transporter activity (2%) | GO:0043953 (96%);GO:0065002 (2%) | protein transport by the Tat complex (96%);intracellular protein transmembrane transport (2%) | IPR002033 (100%);IPR019820 (100%);IPR003369 (2%) | Sec-independent periplasmic protein translocase TatC (100%);Sec-independent periplasmic protein translocase | conserved site (100%);Sec-independent protein translocase protein TatA/B/E (2%) |  |  | 1 | 1 |
| HTPVDGFYAR | Streptomyces | Bacteria |  |  |  | Actinobacteria |  |  | Actinobacteria |  |  |  | Streptomycetales |  |  |  |  | Streptomycetaceae |  |  |  | Streptomyces |  |  |  |  |  |  | Bacteria;Actinobacteria;;;Actinobacteria;Streptomycetales;Streptomycetaceae;;Streptomyces;;;Streptomyces |  | 3.4.21.- | Serine endopeptidases (100%) | GO:0005737 (95%) | cytoplasm (95%) | GO:0008236 (100%) | serine-type peptidase activity (100%) | GO:0006508 (2%) | proteolysis (2%) | IPR011659 (100%);IPR012393 (100%);IPR005151 (98%) | WD40-like Beta Propeller (100%);Tricorn protease (100%);Tail specific protease (98%) |  |  |  | 1 | 100.3846 |
| HVDEGLDGYR | Enterobacteriaceae | Bacteria |  |  |  | Proteobacteria |  |  | Gammaproteobacteria |  |  |  | Enterobacterales |  |  |  |  | Enterobacteriaceae |  |  |  |  |  |  |  |  |  |  | Bacteria;Proteobacteria;;;Gammaproteobacteria;Enterobacterales;Enterobacteriaceae;;;;;Enterobacteriaceae |  |  |  | GO:0016021 (100%);GO:0005886 (95%) | integral component of membrane (100%);plasma membrane (95%) |  |  | GO:0009306 (99%);GO:0009405 (1%);GO:0030254 (1%) | protein secretion (99%);pathogenesis (1%);protein secretion by the type III secretion system (1%) | IPR005838 (100%);IPR005773 (97%);IPR001543 (0%) | Type III secretion system inner membrane P protein (100%);Yop virulence translocation protein R (97%);Surface presentation of antigen (SpoA) (0%) |  |  |  | 1 | 100.9231 |
| LAMMTPQNAR | Bacteria | Bacteria |  |  |  |  |  |  |  |  |  |  |  |  |  |  |  |  |  |  |  |  |  |  |  |  |  |  | Bacteria;;;;;;;;;;;Bacteria | [K01407](https://www.genome.jp/entry/K01407) | 3.4.24.55 (99%);3.4.24.- (1%) | Pitrilysin (99%);Metalloendopeptidases (1%) | GO:0042597 (0%);GO:0005737 (0%);GO:0030288 (0%) | periplasmic space (0%);cytoplasm (0%);outer membrane-bounded periplasmic space (0%) | GO:0046872 (79%);GO:0004222 (76%);GO:0016787 (20%) | metal ion binding (79%);metalloendopeptidase activity (76%);hydrolase activity (20%) | GO:0006508 (0%) | proteolysis (0%) | IPR011249 (100%);IPR032632 (100%);IPR007863 (99%) | Metalloenzyme | LuxS/M16 peptidase-like (100%);Peptidase M16 |  |  | 1 | 100.5308 |
| LDNDDLEEGGMK | Cyanothece | Bacteria |  |  |  | Cyanobacteria |  |  |  | Oscillatoriophycideae |  |  | Oscillatoriales |  |  |  |  | Cyanothecaceae |  |  |  | Cyanothece |  |  |  |  |  |  | Bacteria;Cyanobacteria;;;;Oscillatoriales;Cyanothecaceae;;Cyanothece;;;Cyanothece | K03010 | 2.7.7.6 | DNA-directed RNA polymerase (100%) |  |  | GO:0003677 (100%);GO:0003899 (100%) | DNA binding (100%);DNA-directed 5'-3' RNA polymerase activity (100%) | GO:0006351 (100%) | transcription | DNA-templated (100%) | IPR003716 (100%);IPR006110 (100%);IPR036161 (100%) | DNA-directed RNA polymerase |  |  | 1 | 100.5385 |
| LDTENFDFHR | Caldicellulosiruptor | Bacteria |  |  |  | Firmicutes |  |  | Clostridia |  |  |  | Thermoanaerobacterales |  |  |  |  | Thermoanaerobacterales Family III. Incertae Sedis |  |  |  | Caldicellulosiruptor |  |  |  |  |  |  | Bacteria;Firmicutes;;;Clostridia;Thermoanaerobacterales;Thermoanaerobacterales Family III. Incertae Sedis;;Caldicellulosiruptor;;;Caldicellulosiruptor | [K00943](https://www.genome.jp/entry/K00943) | 2.7.4.9 | dTMP kinase (100%) |  |  | GO:0004798 (100%);GO:0005524 (100%) | thymidylate kinase activity (100%);ATP binding (100%) | GO:0006233 (100%);GO:0006235 (100%) | dTDP biosynthetic process (100%);dTTP biosynthetic process (100%) | IPR018094 (100%);IPR018095 (100%);IPR027417 (100%) | Thymidylate kinase (100%);Thymidylate kinase | conserved site (100%);P-loop containing nucleoside triphosphate hydrolase (100%) |  |  | 1 | 100.6231 |
| LETPDLTEWR | Bacteroidetes | Bacteria |  |  |  | Bacteroidetes |  |  |  |  |  |  |  |  |  |  |  |  |  |  |  |  |  |  |  |  |  |  | Bacteria;Bacteroidetes;;;;;;;;;;Bacteroidetes | [K01811](https://www.genome.jp/dbget-bin/www_bget?ko:K01811) | 3.2.1.177 | Alpha-D-xyloside xylohydrolase (100%) |  |  | GO:0061634 (100%) | alpha-D-xyloside xylohydrolase (100%) | GO:0005975 (100%) | carbohydrate metabolic process (100%) | IPR000322 (100%);IPR011658 (100%);IPR013780 (100%) | Glycoside hydrolase family 31 (100%);PA14 domain (100%);Glycosyl hydrolase | all-beta (100%) |  |  | 1 | 100.5385 |
| LETVEQVR | Bacteria | Bacteria |  |  |  |  |  |  |  |  |  |  |  |  |  |  |  |  |  |  |  |  |  |  |  |  |  |  | Bacteria;;;;;;;;;;;Bacteria | [K01012](https://www.genome.jp/entry/K01012) | 2.8.1.6 | Biotin synthase (100%) |  |  | GO:0004076 (86%);GO:0005506 (86%);GO:0051537 (86%) | biotin synthase activity (86%);iron ion binding (86%);2 iron | 2 sulfur cluster binding (86%) | GO:0009102 (86%);GO:0006352 (14%) | biotin biosynthetic process (86%);DNA-templated transcription | initiation (14%) | IPR002684 (86%);IPR006638 (86%);IPR007197 (86%) |  |  | 1 | 1 |
| LEVELVVER | Actinobacteria | Bacteria |  |  |  | Actinobacteria |  |  | Actinobacteria |  |  |  |  |  |  |  |  |  |  |  |  |  |  |  |  |  |  |  | Bacteria;Actinobacteria;;;Actinobacteria;;;;;;;Actinobacteria | K03010 | 2.7.7.6 | DNA-directed RNA polymerase (100%) | GO:0005618 (0%);GO:0005737 (0%);GO:0005829 (0%) | cell wall (0%);cytoplasm (0%);cytosol (0%) | GO:0003899 (100%);GO:0003677 (100%);GO:0046983 (95%) | DNA-directed 5'-3' RNA polymerase activity (100%);DNA binding (100%);protein dimerization activity (95%) | GO:0006351 (100%) | transcription | DNA-templated (100%) | IPR036643 (100%);IPR011262 (100%);IPR011263 (100%) | DNA-directed RNA polymerase |  |  | 1 | 100.9231 |
| LGDGVDWSR | Bacteria | Bacteria |  |  |  |  |  |  |  |  |  |  |  |  |  |  |  |  |  |  |  |  |  |  |  |  |  |  | Bacteria;;;;;;;;;;;Bacteria | [K01873](https://www.genome.jp/entry/K01873+6.1.1.9+R03665) | 6.1.1.9 | Valine--tRNA ligase (100%) | GO:0005737 (98%);GO:0005618 (0%);GO:0005829 (0%) | cytoplasm (98%);cell wall (0%);cytosol (0%) | GO:0005524 (100%);GO:0004832 (100%);GO:0002161 (94%) | ATP binding (100%);valine-tRNA ligase activity (100%);aminoacyl-tRNA editing activity (94%) | GO:0006438 (99%);GO:0006418 (1%);GO:0006412 (0%) | valyl-tRNA aminoacylation (99%);tRNA aminoacylation for protein translation (1%);translation (0%) | IPR002300 (100%);IPR014729 (100%);IPR002303 (99%) | Aminoacyl-tRNA synthetase | class Ia (100%);Rossmann-like alpha/beta/alpha sandwich fold (100%);Valine-tRNA ligase (99%) |  |  | 1 | 100.6923 |
| LGLAFNEMVK | Bacteria | Bacteria |  |  |  |  |  |  |  |  |  |  |  |  |  |  |  |  |  |  |  |  |  |  |  |  |  |  | Bacteria;;;;;;;;;;;Bacteria | K01712 | 4.2.1.49 | Urocanate hydratase (100%) | GO:0005737 (99%) | cytoplasm (99%) | GO:0016153 (100%);GO:0016787 (3%) | urocanate hydratase activity (100%);hydrolase activity (3%) | GO:0019556 (100%);GO:0019557 (100%);GO:0006548 (0%) | histidine catabolic process to glutamate and formamide (100%);histidine catabolic process to glutamate and formate (100%);histidine catabolic process (0%) | IPR023636 (100%);IPR023637 (100%);IPR035085 (100%) | Urocanase conserved site (100%);Urocanase (100%);Urocanase | Rossmann-like domain (100%) |  |  | 1 | 100.1538 |
| LGLSDFFPR | Bacillus | Bacteria |  |  |  | Firmicutes |  |  | Bacilli |  |  |  | Bacillales |  |  |  |  | Bacillaceae |  |  |  | Bacillus |  |  |  |  |  |  | Bacteria;Firmicutes;;;Bacilli;Bacillales;Bacillaceae;;Bacillus;;;Bacillus |  | 3.1.-.- | Acting on ester bonds (100%) |  |  | GO:0005524 (100%);GO:0008408 (100%);GO:0003677 (95%) | ATP binding (100%);3'-5' exonuclease activity (100%);DNA binding (95%) | GO:0006260 (95%) | DNA replication (95%) | IPR001650 (100%);IPR006054 (100%);IPR006310 (100%) | Helicase | C-terminal (100%);DNA polymerase III epsilon subunit |  |  | 1 | 100.5308 |
| LGTAGDQLAK | Mycobacterium tuberculosis complex | Bacteria |  |  |  | Actinobacteria |  |  | Actinobacteria |  |  |  | Corynebacteriales |  |  |  |  | Mycobacteriaceae |  |  |  | Mycobacterium |  | Mycobacterium tuberculosis complex |  |  |  |  | Bacteria;Actinobacteria;;;Actinobacteria;Corynebacteriales;Mycobacteriaceae;;Mycobacterium;Mycobacterium tuberculosis complex;;Mycobacterium tuberculosis complex |  |  |  | GO:0005618 (100%);GO:0005829 (100%);GO:0005886 (100%) | cell wall (100%);cytosol (100%);plasma membrane (100%) |  |  |  |  | IPR036188 (100%) | FAD/NAD(P)-binding domain superfamily (100%) |  |  |  | 1 | 100.3846 |
| LLDYHAAHPELLR | Streptomyces | Bacteria |  |  |  | Actinobacteria |  |  | Actinobacteria |  |  |  | Streptomycetales |  |  |  |  | Streptomycetaceae |  |  |  | Streptomyces |  |  |  |  |  |  | Bacteria;Actinobacteria;;;Actinobacteria;Streptomycetales;Streptomycetaceae;;Streptomyces;;;Streptomyces |  |  |  |  |  | GO:0003677 (100%) | DNA binding (100%) |  |  | IPR001647 (100%);IPR009057 (100%);IPR036271 (100%) | DNA-binding HTH domain | TetR-type (100%);Homeobox-like domain superfamily (100%);Tetracyclin repressor-like |  |  | 1 | 100.3769 |
| LLLDEGAGPGR | Streptomyces niveus | Bacteria |  |  |  | Actinobacteria |  |  | Actinobacteria |  |  |  | Streptomycetales |  |  |  |  | Streptomycetaceae |  |  |  | Streptomyces |  |  |  | Streptomyces niveus |  |  | Bacteria;Actinobacteria;;;Actinobacteria;Streptomycetales;Streptomycetaceae;;Streptomyces;;Streptomyces niveus;Streptomyces niveus |  | 6.-.-.- | Ligases (100%) |  |  | GO:0003824 (67%);GO:0016874 (33%) | catalytic activity (67%);ligase activity (33%) | GO:0043642 (33%) | novobiocin biosynthetic process (33%) | IPR000873 (100%);IPR006162 (100%);IPR009081 (100%) | AMP-dependent synthetase/ligase (100%);Phosphopantetheine attachment site (100%);Phosphopantetheine binding ACP domain (100%) |  |  |  | 1 | 100.8462 |
| LLLVTNLYR | Bacteria | Bacteria |  |  |  |  |  |  |  |  |  |  |  |  |  |  |  |  |  |  |  |  |  |  |  |  |  |  | Bacteria;;;;;;;;;;;Bacteria |  |  |  |  |  | GO:0008483 (100%) | transaminase activity (100%) |  |  | IPR007737 (100%);IPR013236 (100%);IPR036388 (100%) | Mga helix-turn-helix domain (100%);M protein trans-acting positive regulator (MGA) | PRD domain (100%);Winged helix-like DNA-binding domain superfamily (100%) |  |  | 1 | 100.3 |
| LNLVTNFGK | Bacillales | Bacteria |  |  |  | Firmicutes |  |  | Bacilli |  |  |  | Bacillales |  |  |  |  |  |  |  |  |  |  |  |  |  |  |  | Bacteria;Firmicutes;;;Bacilli;Bacillales;;;;;;Bacillales | [K00995](https://www.genome.jp/dbget-bin/www_bget?ko:K00995) | 2.7.8.5 | CDP-diacylglycerol--glycerol-3-phosphate 1-phosphatidyltransferase (100%) | GO:0016021 (97%);GO:0005886 (1%) | integral component of membrane (97%);plasma membrane (1%) | GO:0008444 (98%);GO:0016740 (2%) | CDP-diacylglycerol-glycerol-3-phosphate 3-phosphatidyltransferase activity (98%);transferase activity (2%) | GO:0008654 (96%);GO:0006655 (1%);GO:0046474 (1%) | phospholipid biosynthetic process (96%);phosphatidylglycerol biosynthetic process (1%);glycerophospholipid biosynthetic process (1%) | IPR000462 (100%);IPR004570 (100%) | CDP-alcohol phosphatidyltransferase (100%);CDP-diacylglycerol--glycerol-3-phosphate 3-phosphatidyltransferase (100%) |  |  |  | 1 | 100.4615 |
| LPGLDSFTPQLSAVPGSDAER | Mycobacterium tuberculosis | Bacteria |  |  |  | Actinobacteria |  |  | Actinobacteria |  |  |  | Corynebacteriales |  |  |  |  | Mycobacteriaceae |  |  |  | Mycobacterium |  | Mycobacterium tuberculosis complex |  | Mycobacterium tuberculosis |  |  | Bacteria;Actinobacteria;;;Actinobacteria;Corynebacteriales;Mycobacteriaceae;;Mycobacterium;Mycobacterium tuberculosis complex;Mycobacterium tuberculosis;Mycobacterium tuberculosis |  | 7.1.1.- | Hydron translocation or charge separation linked to oxidoreductase reactions (100%) | GO:0005886 (100%);GO:0016021 (100%) | plasma membrane (100%);integral component of membrane (100%) | GO:0008137 (100%);GO:0048038 (100%) | NADH dehydrogenase (ubiquinone) activity (100%);quinone binding (100%) | GO:0042773 (100%) | ATP synthesis coupled electron transport (100%) | IPR001750 (100%);IPR010096 (100%) | NADH:quinone oxidoreductase/Mrp antiporter | membrane subunit (100%);NAD(P)H-quinone oxidoreductase |  |  | 1 | 100.6154 |
| LQGLDVGEVR | Streptomyces | Bacteria |  |  |  | Actinobacteria |  |  | Actinobacteria |  |  |  | Streptomycetales |  |  |  |  | Streptomycetaceae |  |  |  | Streptomyces |  |  |  |  |  |  | Bacteria;Actinobacteria;;;Actinobacteria;Streptomycetales;Streptomycetaceae;;Streptomyces;;;Streptomyces | K01707 | 4.2.1.41 | 5-dehydro-4-deoxyglucarate dehydratase (100%) |  |  | GO:0047448 (98%);GO:0016829 (2%) | 5-dehydro-4-deoxyglucarate dehydratase activity (98%);lyase activity (2%) | GO:0042838 (98%) | D-glucarate catabolic process (98%) | IPR002220 (100%);IPR013785 (100%);IPR017655 (98%) | DapA-like (100%);Aldolase-type TIM barrel (100%);5-dehydro-4-deoxyglucarate dehydratase (98%) |  |  |  | 1 | 100.3 |
| LSDNGMVTVNMGKPK | Neisseria | Bacteria |  |  |  | Proteobacteria |  |  | Betaproteobacteria |  |  |  | Neisseriales |  |  |  |  | Neisseriaceae |  |  |  | Neisseria |  |  |  |  |  |  | Bacteria;Proteobacteria;;;Betaproteobacteria;Neisseriales;Neisseriaceae;;Neisseria;;;Neisseria | [K00370](https://www.genome.jp/dbget-bin/www_bget?K00370+K00371+K00374+1.7.5.1+R09497) | 5.1.1.7 | Diaminopimelate epimerase (100%) | GO:0005737 (93%);GO:0005829 (4%) | cytoplasm (93%);cytosol (4%) | GO:0008837 (100%) | diaminopimelate epimerase activity (100%) | GO:0009089 (100%) | lysine biosynthetic process via diaminopimelate (100%) | IPR001653 (100%);IPR018510 (96%) | Diaminopimelate epimerase | DapF (100%);Diaminopimelate epimerase |  |  | 1 | 100.6923 |
| LSGLSLPTLAVDLPGGGGK | Treponema | Bacteria |  |  |  | Spirochaetes |  |  | Spirochaetia |  |  |  | Spirochaetales |  |  |  |  | Spirochaetaceae |  |  |  | Treponema |  |  |  |  |  |  | Bacteria;Spirochaetes;;;Spirochaetia;Spirochaetales;Spirochaetaceae;;Treponema;;;Treponema | [K01843](https://www.genome.jp/dbget-bin/www_bget?K01843+5.4.3.2+R00461) | 5.4.3.2 (86%);5.4.3.- (14%) | Lysine 2 | 3-aminomutase (86%);Transferring amino groups (14%) |  |  | GO:0046872 (100%);GO:0051539 (100%);GO:0050066 (75%) | metal ion binding (100%);4 iron | 4 sulfur cluster binding (100%);lysine 2 | 3-aminomutase activity (75%) |  |  |  |  | 1 | 100.6077 |
| LSSPAWDGKPR | Tropheryma whipplei | Bacteria |  |  |  | Actinobacteria |  |  | Actinobacteria |  |  |  | Micrococcales |  |  |  |  |  |  |  |  | Tropheryma |  |  |  | Tropheryma whipplei |  |  | Bacteria;Actinobacteria;;;Actinobacteria;Micrococcales;;;Tropheryma;;Tropheryma whipplei;Tropheryma whipplei | [K00566](https://www.genome.jp/entry/K00566) | 2.8.1.13 | tRNA-uridine 2-sulfurtransferase (100%) | GO:0005737 (100%) | cytoplasm (100%) | GO:0000049 (100%);GO:0005524 (100%);GO:0016783 (100%) | tRNA binding (100%);ATP binding (100%);sulfurtransferase activity (100%) | GO:0006400 (100%) | tRNA modification (100%) | IPR004506 (100%);IPR014729 (100%);IPR023382 (100%) | tRNA-specific 2-thiouridylase (100%);Rossmann-like alpha/beta/alpha sandwich fold (100%);Adenine nucleotide alpha hydrolase-like domains (100%) |  |  |  | 1 | 1 |
| LSVGFNR | Bacteria | Bacteria |  |  |  |  |  |  |  |  |  |  |  |  |  |  |  |  |  |  |  |  |  |  |  |  |  |  | Bacteria;;;;;;;;;;;Bacteria |  |  |  | GO:0016021 (92%);GO:0005886 (1%) | integral component of membrane (92%);plasma membrane (1%) | GO:0003677 (98%);GO:0005524 (94%);GO:0003700 (4%) | DNA binding (98%);ATP binding (94%);DNA-binding transcription factor activity (4%) | GO:0051301 (41%);GO:0007049 (1%);GO:0007059 (1%) | cell division (41%);cell cycle (1%);chromosome segregation (1%) | IPR036388 (99%);IPR036390 (99%);IPR018541 (94%) | Winged helix-like DNA-binding domain superfamily (99%);Winged helix DNA-binding domain superfamily (99%);FtsK gamma domain (94%) |  |  |  | 1 | 100.3077 |
| LTLVEYGFR | Bacteria | Bacteria |  |  |  |  |  |  |  |  |  |  |  |  |  |  |  |  |  |  |  |  |  |  |  |  |  |  | Bacteria;;;;;;;;;;;Bacteria |  |  |  | GO:0005737 (98%);GO:0009380 (93%) | cytoplasm (98%);excinuclease repair complex (93%) | GO:0003677 (100%);GO:0005524 (100%);GO:0016887 (93%) | DNA binding (100%);ATP binding (100%);ATPase activity (93%) | GO:0006289 (100%);GO:0009432 (98%) | nucleotide-excision repair (100%);SOS response (98%) | IPR004807 (100%);IPR027417 (100%);IPR001650 (98%) | UvrABC system | subunit B (100%);P-loop containing nucleoside triphosphate hydrolase (100%);Helicase |  |  | 1 | 100.7692 |
| LTVLSGPSGVGK | Bacteria | Bacteria |  |  |  |  |  |  |  |  |  |  |  |  |  |  |  |  |  |  |  |  |  |  |  |  |  |  | Bacteria;;;;;;;;;;;Bacteria | [K00942](https://www.genome.jp/entry/K00942) | 2.7.4.8 (99%);3.6.1.- (1%) | Guanylate kinase (99%);In phosphorus-containing anhydrides (1%) | GO:0005737 (98%);GO:0016021 (0%);GO:0005829 (0%) | cytoplasm (98%);integral component of membrane (0%);cytosol (0%) | GO:0004385 (99%);GO:0005524 (98%);GO:0003924 (1%) | guanylate kinase activity (99%);ATP binding (98%);GTPase activity (1%) | GO:0042274 (1%);GO:0042883 (0%);GO:0042254 (0%) | ribosomal small subunit biogenesis (1%);cysteine transport (0%);ribosome biogenesis (0%) | IPR027417 (100%);IPR008144 (99%);IPR008145 (99%) | P-loop containing nucleoside triphosphate hydrolase (100%);Guanylate kinase-like domain (99%);Guanylate kinase/L-type calcium channel beta subunit (99%) |  |  |  | 1 | 1 |
| LVGAVADASR | Streptomyces | Bacteria |  |  |  | Actinobacteria |  |  | Actinobacteria |  |  |  | Streptomycetales |  |  |  |  | Streptomycetaceae |  |  |  | Streptomyces |  |  |  |  |  |  | Bacteria;Actinobacteria;;;Actinobacteria;Streptomycetales;Streptomycetaceae;;Streptomyces;;;Streptomyces | [K01514](https://www.genome.jp/entry/K01514) | 3.6.1.11 | Exopolyphosphatase (100%) |  |  | GO:0004309 (50%);GO:0016462 (50%) | exopolyphosphatase activity (50%);pyrophosphatase activity (50%) | GO:0006793 (50%) | phosphorus metabolic process (50%) | IPR003695 (100%) | Ppx/GppA phosphatase (100%) |  |  |  | 1 | 1 |
| LYDEGSSPTR | Rhizobiaceae | Bacteria |  |  |  | Proteobacteria |  |  | Alphaproteobacteria |  |  |  | Rhizobiales |  |  |  |  | Rhizobiaceae |  |  |  |  |  |  |  |  |  |  | Bacteria;Proteobacteria;;;Alphaproteobacteria;Rhizobiales;Rhizobiaceae;;;;;Rhizobiaceae |  |  |  |  |  | GO:0016793 (80%);GO:0016787 (20%) | triphosphoric monoester hydrolase activity (80%);hydrolase activity (20%) |  |  | IPR003607 (100%);IPR006261 (100%);IPR006674 (100%) | HD/PDEase domain (100%);dNTP triphosphohydrolase (100%);HD domain (100%) |  |  |  | 1 | 100.6154 |
| MEGGYNFYSK | Mycoplasma mobile | Bacteria |  |  |  | Tenericutes |  |  | Mollicutes |  |  |  | Mycoplasmatales |  |  |  |  | Mycoplasmataceae |  |  |  | Mycoplasma |  |  |  | Mycoplasma mobile |  |  | Bacteria;Tenericutes;;;Mollicutes;Mycoplasmatales;Mycoplasmataceae;;Mycoplasma;;Mycoplasma mobile;Mycoplasma mobile | [K10563](https://www.genome.jp/entry/K10563) | 3.2.2.23 ;4.2.99.18 | DNA-formamidopyrimidine glycosylase (100%);DNA-(apurinic or apyrimidinic site) lyase (100%) |  |  | GO:0003684 (100%);GO:0008270 (100%);GO:0008534 (100%) | damaged DNA binding (100%);zinc ion binding (100%);oxidized purine nucleobase lesion DNA N-glycosylase activity (100%) | GO:0006284 (100%);GO:0006289 (100%) | base-excision repair (100%);nucleotide-excision repair (100%) | IPR000214 (100%);IPR010663 (100%);IPR010979 (100%) | Zinc finger | DNA glycosylase/AP lyase-type (100%);Zinc finger |  |  | 1 | 100.9231 |
| MTPTQAK | Bacteria | Bacteria |  |  |  |  |  |  |  |  |  |  |  |  |  |  |  |  |  |  |  |  |  |  |  |  |  |  | Bacteria;;;;;;;;;;;Bacteria | [K08483](https://www.genome.jp/entry/K08483) | 2.7.3.9 (50%);3.3.1.1 (50%) | Phosphoenolpyruvate--protein phosphotransferase (50%);Adenosylhomocysteinase (50%) | GO:0005737 (50%) | cytoplasm (50%) | GO:0004013 (50%);GO:0008965 (50%);GO:0016301 (50%) | adenosylhomocysteinase activity (50%);phosphoenolpyruvate-protein phosphotransferase activity (50%);kinase activity (50%) | GO:0006730 (50%);GO:0009401 (50%) | one-carbon metabolic process (50%);phosphoenolpyruvate-dependent sugar phosphotransferase system (50%) | IPR000043 (50%);IPR000121 (50%);IPR006318 (50%) | Adenosylhomocysteinase-like (50%);PEP-utilising enzyme | C-terminal (50%);Phosphotransferase system |  |  | 1 | 1 |
| NAELVYTEYPVYR | Candidatus Endomicrobium trichonymphae | Bacteria |  |  |  | Elusimicrobia |  |  | Endomicrobia |  |  |  | Endomicrobiales |  |  |  |  | Endomicrobiaceae |  |  |  | Endomicrobium |  |  |  | Candidatus Endomicrobium trichonymphae |  |  | Bacteria;Elusimicrobia;;;Endomicrobia;Endomicrobiales;Endomicrobiaceae;;Endomicrobium;;Candidatus Endomicrobium trichonymphae;Candidatus Endomicrobium trichonymphae | K03010 | 2.7.7.6 | DNA-directed RNA polymerase (100%) |  |  | GO:0000287 (100%);GO:0003677 (100%);GO:0003899 (100%) | magnesium ion binding (100%);DNA binding (100%);DNA-directed 5'-3' RNA polymerase activity (100%) | GO:0006351 (100%) | transcription | DNA-templated (100%) | IPR000722 (100%);IPR006592 (100%);IPR007066 (100%) | RNA polymerase |  |  | 1 | 1 |
| NLAHFHAADPEYGK | Streptomyces | Bacteria |  |  |  | Actinobacteria |  |  | Actinobacteria |  |  |  | Streptomycetales |  |  |  |  | Streptomycetaceae |  |  |  | Streptomyces |  |  |  |  |  |  | Bacteria;Actinobacteria;;;Actinobacteria;Streptomycetales;Streptomycetaceae;;Streptomyces;;;Streptomyces | [K03781](https://www.genome.jp/dbget-bin/www_bget?ko:K03781) | 1.11.1.6 | Catalase (100%) | GO:0005737 (1%) | cytoplasm (1%) | GO:0004096 (100%);GO:0046872 (99%);GO:0020037 (93%) | catalase activity (100%);metal ion binding (99%);heme binding (93%) | GO:0042744 (99%);GO:0006979 (92%);GO:0042542 (1%) | hydrogen peroxide catabolic process (99%);response to oxidative stress (92%);response to hydrogen peroxide (1%) | IPR002226 (100%);IPR010582 (100%);IPR011614 (100%) | Catalase haem-binding site (100%);Catalase immune-responsive domain (100%);Catalase core domain (100%) |  |  |  | 1 | 100.3154 |
| NMPVWR | Bacteria | Bacteria |  |  |  |  |  |  |  |  |  |  |  |  |  |  |  |  |  |  |  |  |  |  |  |  |  |  | Bacteria;;;;;;;;;;;Bacteria | K03526 | 1.17.7.3 (93%);1.17.7.1 (7%);3.1.-.- (0%) | (E)-4-hydroxy-3-methylbut-2-enyl-diphosphate synthase (flavodoxin) (93%);(E)-4-hydroxy-3-methylbut-2-enyl-diphosphate synthase (ferredoxin) (7%);Acting on ester bonds (0%) | GO:0005886 (0%);GO:0016021 (0%) | plasma membrane (0%);integral component of membrane (0%) | GO:0046429 (99%);GO:0051539 (96%);GO:0005506 (85%) | 4-hydroxy-3-methylbut-2-en-1-yl diphosphate synthase activity (99%);4 iron | 4 sulfur cluster binding (96%);iron ion binding (85%) | GO:0019288 (96%);GO:0016114 (96%);GO:0006281 (0%) | isopentenyl diphosphate biosynthetic process | methylerythritol 4-phosphate pathway (96%);terpenoid biosynthetic process (96%);DNA repair (0%) | IPR004588 (99%);IPR011005 (98%);IPR016425 (94%) |  |  | 1 | 100.3923 |
| NNAEQGNK | Hungateiclostridium | Bacteria |  |  |  | Firmicutes |  |  | Clostridia |  |  |  | Clostridiales |  |  |  |  | Hungateiclostridiaceae |  |  |  | Hungateiclostridium |  |  |  |  |  |  | Bacteria;Firmicutes;;;Clostridia;Clostridiales;Hungateiclostridiaceae;;Hungateiclostridium;;;Hungateiclostridium |  |  |  | GO:0016021 (100%);GO:0005886 (50%) | integral component of membrane (100%);plasma membrane (50%) | GO:0004888 (50%) | transmembrane signaling receptor activity (50%) | GO:0007165 (100%);GO:0006935 (50%) | signal transduction (100%);chemotaxis (50%) | IPR003660 (100%);IPR004089 (100%);IPR024478 (100%) | HAMP domain (100%);Methyl-accepting chemotaxis protein (MCP) signalling domain (100%);Chemotaxis methyl-accepting receptor HlyB-like | 4HB MCP domain (100%) |  |  | 1 | 100.9231 |
| NSAEHQWFK | Mycoplasma pneumoniae | Bacteria |  |  |  | Tenericutes |  |  | Mollicutes |  |  |  | Mycoplasmatales |  |  |  |  | Mycoplasmataceae |  |  |  | Mycoplasma |  |  |  | Mycoplasma pneumoniae |  |  | Bacteria;Tenericutes;;;Mollicutes;Mycoplasmatales;Mycoplasmataceae;;Mycoplasma;;Mycoplasma pneumoniae;Mycoplasma pneumoniae |  | 2.7.7.- | Nucleotidyltransferases (100%) | GO:1990077 (100%) | primosome complex (100%) | GO:0003677 (100%);GO:0003896 (100%);GO:0008270 (100%) | DNA binding (100%);DNA primase activity (100%);zinc ion binding (100%) |  |  | IPR002694 (100%);IPR006171 (100%);IPR006295 (100%) | Zinc finger | CHC2-type (100%);TOPRIM domain (100%);DNA primase |  |  | 1 | 1 |
| QAFAALDGGK | Neisseria | Bacteria |  |  |  | Proteobacteria |  |  | Betaproteobacteria |  |  |  | Neisseriales |  |  |  |  | Neisseriaceae |  |  |  | Neisseria |  |  |  |  |  |  | Bacteria;Proteobacteria;;;Betaproteobacteria;Neisseriales;Neisseriaceae;;Neisseria;;;Neisseria | K01695 | 4.2.1.20 | Tryptophan synthase (100%) | GO:0009507 (3%) | chloroplast (3%) | GO:0004834 (100%) | tryptophan synthase activity (100%) | GO:0000162 (3%) | tryptophan biosynthetic process (3%) | IPR002028 (100%);IPR011060 (100%);IPR013785 (100%) | Tryptophan synthase | alpha chain (100%);Ribulose-phosphate binding barrel (100%);Aldolase-type TIM barrel (100%) |  |  | 1 | 1 |
| QASYQQHYPVSPAQR | Bacillus subtilis group | Bacteria |  |  |  | Firmicutes |  |  | Bacilli |  |  |  | Bacillales |  |  |  |  | Bacillaceae |  |  |  | Bacillus |  | Bacillus subtilis group |  |  |  |  | Bacteria;Firmicutes;;;Bacilli;Bacillales;Bacillaceae;;Bacillus;Bacillus subtilis group;;Bacillus subtilis group |  |  |  |  |  | GO:0031177 (100%);GO:0003824 (83%);GO:0016874 (17%) | phosphopantetheine binding (100%);catalytic activity (83%);ligase activity (17%) | GO:0017000 (8%);GO:0030435 (8%) | antibiotic biosynthetic process (8%);sporulation resulting in formation of a cellular spore (8%) | IPR000873 (100%);IPR001242 (100%);IPR006162 (100%) | AMP-dependent synthetase/ligase (100%);Condensation domain (100%);Phosphopantetheine attachment site (100%) |  |  |  | 1 | 99.99231 |
| QDGWVEGK | Bacteria | Bacteria |  |  |  |  |  |  |  |  |  |  |  |  |  |  |  |  |  |  |  |  |  |  |  |  |  |  | Bacteria;;;;;;;;;;;Bacteria | [K01155](https://www.genome.jp/entry/K01155) | 3.1.21.4 | Type II site-specific deoxyribonuclease (100%) |  |  | GO:0003677 (100%);GO:0009036 (100%) | DNA binding (100%);type II site-specific deoxyribonuclease activity (100%) | GO:0009307 (100%) | DNA restriction-modification system (100%) | IPR011335 (100%);IPR011336 (100%);IPR022725 (100%) | Restriction endonuclease type II-like (100%);Restriction endonuclease | type II |  |  | 1 | 100.6154 |
| QELNLHEYCVR | Gammaproteobacteria | Bacteria |  |  |  | Proteobacteria |  |  | Gammaproteobacteria |  |  |  |  |  |  |  |  |  |  |  |  |  |  |  |  |  |  |  | Bacteria;Proteobacteria;;;Gammaproteobacteria;;;;;;;Gammaproteobacteria | [K01356](https://www.genome.jp/entry/K01356) | 3.4.21.- (86%);3.4.21.88 (14%) | Serine endopeptidases (86%);Repressor LexA (14%) |  |  | GO:0016787 (95%);GO:0003677 (94%);GO:0003887 (3%) | hydrolase activity (95%);DNA binding (94%);DNA-directed DNA polymerase activity (3%) | GO:0006281 (98%);GO:0009432 (98%);GO:0006355 (94%) | DNA repair (98%);SOS response (98%);regulation of transcription | DNA-templated (94%) | IPR036286 (100%);IPR006197 (99%);IPR015927 (99%) | LexA/Signal peptidase-like superfamily (100%);Peptidase S24 |  |  | 1 | 100.3077 |
| QENEDSPGDFSVDEK | Nitrosospira multiformis | Bacteria |  |  |  | Proteobacteria |  |  | Betaproteobacteria |  |  |  | Nitrosomonadales |  |  |  |  | Nitrosomonadaceae |  |  |  | Nitrosospira |  |  |  | Nitrosospira multiformis |  |  | Bacteria;Proteobacteria;;;Betaproteobacteria;Nitrosomonadales;Nitrosomonadaceae;;Nitrosospira;;Nitrosospira multiformis;Nitrosospira multiformis |  |  |  | GO:0005737 (100%);GO:0005886 (100%) | cytoplasm (100%);plasma membrane (100%) | GO:0005524 (100%);GO:0046872 (100%) | ATP binding (100%);metal ion binding (100%) | GO:0006605 (100%);GO:0017038 (100%);GO:0065002 (100%) | protein targeting (100%);protein import (100%);intracellular protein transmembrane transport (100%) | IPR000185 (100%);IPR004027 (100%);IPR011115 (100%) | Protein translocase subunit SecA (100%);SEC-C motif (100%);SecA DEAD-like | N-terminal (100%) |  |  | 1 | 100.3846 |
| QESPTADR | Streptococcus pneumoniae | Bacteria |  |  |  | Firmicutes |  |  | Bacilli |  |  |  | Lactobacillales |  |  |  |  | Streptococcaceae |  |  |  | Streptococcus |  |  |  | Streptococcus pneumoniae |  |  | Bacteria;Firmicutes;;;Bacilli;Lactobacillales;Streptococcaceae;;Streptococcus;;Streptococcus pneumoniae;Streptococcus pneumoniae | K01390 | 3.4.24.13 (89%);3.4.21.74 (22%);3.4.24.- (11%) | IgA-specific metalloendopeptidase (89%);Venombin A (22%);Metalloendopeptidases (11%) | GO:0005576 (100%);GO:0005618 (100%);GO:0016021 (100%) | extracellular region (100%);cell wall (100%);integral component of membrane (100%) | GO:0004222 (100%);GO:0008270 (100%) | metalloendopeptidase activity (100%);zinc ion binding (100%) | GO:0009405 (10%) | pathogenesis (10%) | IPR008006 (100%);IPR011505 (100%);IPR005877 (91%) | Peptidase M26 | N-terminal domain (100%);Peptidase M26 |  |  | 1 | 1 |
| QGLSGLAEK | Proteobacteria | Bacteria |  |  |  | Proteobacteria |  |  |  |  |  |  |  |  |  |  |  |  |  |  |  |  |  |  |  |  |  |  | Bacteria;Proteobacteria;;;;;;;;;;Proteobacteria | K01523 | 3.6.1.31 ;3.5.4.19 (1%) | Phosphoribosyl-ATP diphosphatase (100%);Phosphoribosyl-AMP cyclohydrolase (1%) | GO:0005737 (97%) | cytoplasm (97%) | GO:0004636 (100%);GO:0005524 (100%);GO:0004635 (1%) | phosphoribosyl-ATP diphosphatase activity (100%);ATP binding (100%);phosphoribosyl-AMP cyclohydrolase activity (1%) | GO:0000105 (100%) | histidine biosynthetic process (100%) | IPR008179 (100%);IPR021130 (100%);IPR002496 (1%) | Phosphoribosyl-ATP pyrophosphohydrolase (100%);Phosphoribosyl-ATP pyrophosphohydrolase-like (100%);Phosphoribosyl-AMP cyclohydrolase domain (1%) |  |  |  | 1 | 100.3077 |
| QGVHDVSFTAK | Rhizobiaceae | Bacteria |  |  |  | Proteobacteria |  |  | Alphaproteobacteria |  |  |  | Rhizobiales |  |  |  |  | Rhizobiaceae |  |  |  |  |  |  |  |  |  |  | Bacteria;Proteobacteria;;;Alphaproteobacteria;Rhizobiales;Rhizobiaceae;;;;;Rhizobiaceae | [K24819](https://www.genome.jp/entry/K24819) | 7.5.2.3 | ABC-type beta-glucan transporter (100%) | GO:0005886 (100%);GO:0016021 (100%) | plasma membrane (100%);integral component of membrane (100%) | GO:0005524 (100%);GO:0015441 (100%);GO:0016887 (91%) | ATP binding (100%);ATPase-coupled beta-glucan transporter activity (100%);ATPase activity (91%) |  |  | IPR003439 (100%);IPR011527 (100%);IPR017871 (100%) | ABC transporter-like (100%);ABC transporter type 1 | transmembrane domain (100%);ABC transporter |  |  | 1 | 100.0077 |
| QLAALHHAGHELVLVSSGAVAAGFGALGFK | Neisseria | Bacteria |  |  |  | Proteobacteria |  |  | Betaproteobacteria |  |  |  | Neisseriales |  |  |  |  | Neisseriaceae |  |  |  | Neisseria |  |  |  |  |  |  | Bacteria;Proteobacteria;;;Betaproteobacteria;Neisseriales;Neisseriaceae;;Neisseria;;;Neisseria | [K00931](https://www.genome.jp/entry/K00931) | 2.7.2.11 | Glutamate 5-kinase (100%) | GO:0005737 (88%) | cytoplasm (88%) | GO:0004349 (100%);GO:0003723 (88%);GO:0005524 (88%) | glutamate 5-kinase activity (100%);RNA binding (88%);ATP binding (88%) | GO:0055129 (88%) | L-proline biosynthetic process (88%) | IPR001048 (100%);IPR001057 (100%);IPR036393 (100%) | Aspartate/glutamate/uridylate kinase (100%);Glutamate/acetylglutamate kinase (100%);Acetylglutamate kinase-like superfamily (100%) |  |  |  | 1 | 100.3846 |
| QMQDQVMALSR | Enterobacteriaceae | Bacteria |  |  |  | Proteobacteria |  |  | Gammaproteobacteria |  |  |  | Enterobacterales |  |  |  |  | Enterobacteriaceae |  |  |  |  |  |  |  |  |  |  | Bacteria;Proteobacteria;;;Gammaproteobacteria;Enterobacterales;Enterobacteriaceae;;;;;Enterobacteriaceae | [K01188](https://www.genome.jp/entry/K01188) | 3.2.1.21 (99%);3.2.1.52 (1%) | Beta-glucosidase (99%);Beta-N-acetylhexosaminidase (1%) | GO:0042597 (0%) | periplasmic space (0%) | GO:0004553 (58%);GO:0008422 (41%);GO:0102483 (41%) | hydrolase activity | hydrolyzing O-glycosyl compounds (58%);beta-glucosidase activity (41%);scopolin beta-glucosidase activity (41%) | GO:0005975 (76%);GO:0008152 (24%);GO:0009254 (0%) | carbohydrate metabolic process (76%);metabolic process (24%);peptidoglycan turnover (0%) | IPR001764 (100%);IPR017853 (100%);IPR036962 (100%) | Glycoside hydrolase |  |  | 1 | 100.3 |
| QNANGTSSSGNK | Mycoplasma genitalium | Bacteria |  |  |  | Tenericutes |  |  | Mollicutes |  |  |  | Mycoplasmatales |  |  |  |  | Mycoplasmataceae |  |  |  | Mycoplasma |  |  |  | Mycoplasma genitalium |  |  | Bacteria;Tenericutes;;;Mollicutes;Mycoplasmatales;Mycoplasmataceae;;Mycoplasma;;Mycoplasma genitalium;Mycoplasma genitalium |  |  |  | GO:0005886 (100%);GO:0016021 (100%) | plasma membrane (100%);integral component of membrane (100%) |  |  |  |  |  |  |  |  |  | 1 | 100.6923 |
| QNFTTSEELVR | Neisseria | Bacteria |  |  |  | Proteobacteria |  |  | Betaproteobacteria |  |  |  | Neisseriales |  |  |  |  | Neisseriaceae |  |  |  | Neisseria |  |  |  |  |  |  | Bacteria;Proteobacteria;;;Betaproteobacteria;Neisseriales;Neisseriaceae;;Neisseria;;;Neisseria |  |  |  | GO:0009279 (100%);GO:0016021 (64%) | cell outer membrane (100%);integral component of membrane (64%) | GO:0015091 (100%) | ferric iron transmembrane transporter activity (100%) | GO:0055072 (4%) | iron ion homeostasis (4%) | IPR000531 (100%);IPR010917 (100%);IPR010948 (100%) | TonB-dependent receptor-like | beta-barrel (100%);TonB-dependent receptor |  |  | 1 | 100.8462 |
| QTTADQENVPSGK | Bacillus | Bacteria |  |  |  | Firmicutes |  |  | Bacilli |  |  |  | Bacillales |  |  |  |  | Bacillaceae |  |  |  | Bacillus |  |  |  |  |  |  | Bacteria;Firmicutes;;;Bacilli;Bacillales;Bacillaceae;;Bacillus;;;Bacillus |  |  |  |  |  |  |  |  |  | IPR024997 (100%) | Protein of unknown function DUF3892 (100%) |  |  |  | 1 | 100.6923 |
| QVLAAGEGTDSMK | Streptomyces | Bacteria |  |  |  | Actinobacteria |  |  | Actinobacteria |  |  |  | Streptomycetales |  |  |  |  | Streptomycetaceae |  |  |  | Streptomyces |  |  |  |  |  |  | Bacteria;Actinobacteria;;;Actinobacteria;Streptomycetales;Streptomycetaceae;;Streptomyces;;;Streptomyces | [K23025](https://www.genome.jp/entry/K23025) | 7.1.3.1 (99%);3.6.1.1 (1%) | H(+)-exporting diphosphatase (99%);Inorganic diphosphatase (1%) | GO:0005887 (98%);GO:0016021 (0%) | integral component of plasma membrane (98%);integral component of membrane (0%) | GO:0004427 (100%);GO:0009678 (99%);GO:0000287 (98%) | inorganic diphosphatase activity (100%);pyrophosphate hydrolysis-driven proton transmembrane transporter activity (99%);magnesium ion binding (98%) |  |  | IPR004131 (100%);IPR036019 (2%) | Pyrophosphate-energised proton pump (100%);Gated mechanosensitive channel (2%) |  |  |  | 1 | 100.1538 |
| QVVPQLTESFNA | Bacteria | Bacteria |  |  |  |  |  |  |  |  |  |  |  |  |  |  |  |  |  |  |  |  |  |  |  |  |  |  | Bacteria;;;;;;;;;;;Bacteria | [K01507](https://www.genome.jp/dbget-bin/www_bget?K01507+K15986+K15987+K11725+K11726+K06019+3.6.1.1) | 3.6.1.1 | Inorganic diphosphatase (100%) | GO:0005737 (100%) | cytoplasm (100%) | GO:0004427 (99%);GO:0030145 (99%);GO:0016462 (1%) | inorganic diphosphatase activity (99%);manganese ion binding (99%);pyrophosphatase activity (1%) | GO:0006798 (0%);GO:0006807 (0%) | polyphosphate catabolic process (0%);nitrogen compound metabolic process (0%) | IPR004097 (100%);IPR038222 (100%);IPR038763 (100%) | DHHA2 domain (100%);DHHA2 domain superfamily (100%);DHH phosphoesterase superfamily (100%) |  |  |  | 1 | 100.4692 |
| RPSDPGALVK | Mycobacterium tuberculosis complex | Bacteria |  |  |  | Actinobacteria |  |  | Actinobacteria |  |  |  | Corynebacteriales |  |  |  |  | Mycobacteriaceae |  |  |  | Mycobacterium |  | Mycobacterium tuberculosis complex |  |  |  |  | Bacteria;Actinobacteria;;;Actinobacteria;Corynebacteriales;Mycobacteriaceae;;Mycobacterium;Mycobacterium tuberculosis complex;;Mycobacterium tuberculosis complex |  |  |  | GO:0005576 (50%);GO:0030288 (50%) | extracellular region (50%);outer membrane-bounded periplasmic space (50%) | GO:1904680 (50%) | peptide transmembrane transporter activity (50%) | GO:0008654 (100%);GO:0009405 (100%);GO:0046488 (100%) | phospholipid biosynthetic process (100%);pathogenesis (100%);phosphatidylinositol metabolic process (100%) | IPR000914 (100%);IPR039424 (100%) | Solute-binding protein family 5 domain (100%);Solute-binding protein family 5 (100%) |  |  |  | 1 | 100.3846 |
| RPSFGWQTEQQLWSGFSK | Bacillus | Bacteria |  |  |  | Firmicutes |  |  | Bacilli |  |  |  | Bacillales |  |  |  |  | Bacillaceae |  |  |  | Bacillus |  |  |  |  |  |  | Bacteria;Firmicutes;;;Bacilli;Bacillales;Bacillaceae;;Bacillus;;;Bacillus |  |  |  | GO:0016021 (100%);GO:0005886 (13%) | integral component of membrane (100%);plasma membrane (13%) |  |  | GO:0030152 (13%);GO:0030153 (13%) | bacteriocin biosynthetic process (13%);bacteriocin immunity (13%) |  |  |  |  |  | 1 | 100.8462 |
| SALAEAEVEYQDK | Bacteria | Bacteria |  |  |  |  |  |  |  |  |  |  |  |  |  |  |  |  |  |  |  |  |  |  |  |  |  |  | Bacteria;;;;;;;;;;;Bacteria | [K01870](https://www.genome.jp/entry/K01870) | 6.1.1.5 | Isoleucine--tRNA ligase (100%) | GO:0005737 (96%) | cytoplasm (96%) | GO:0005524 (100%);GO:0004822 (98%);GO:0008270 (96%) | ATP binding (100%);isoleucine-tRNA ligase activity (98%);zinc ion binding (96%) | GO:0006428 (98%);GO:0006418 (2%) | isoleucyl-tRNA aminoacylation (98%);tRNA aminoacylation for protein translation (2%) | IPR002300 (100%);IPR009008 (100%);IPR014729 (100%) | Aminoacyl-tRNA synthetase | class Ia (100%);Valyl/Leucyl/Isoleucyl-tRNA synthetase |  |  | 1 | 100.5385 |
| SGDQTLLELGK | Mycoplasma pneumoniae | Bacteria |  |  |  | Tenericutes |  |  | Mollicutes |  |  |  | Mycoplasmatales |  |  |  |  | Mycoplasmataceae |  |  |  | Mycoplasma |  |  |  | Mycoplasma pneumoniae |  |  | Bacteria;Tenericutes;;;Mollicutes;Mycoplasmatales;Mycoplasmataceae;;Mycoplasma;;Mycoplasma pneumoniae;Mycoplasma pneumoniae |  |  |  | GO:0005886 (100%) | plasma membrane (100%) |  |  |  |  | IPR022186 (100%) | Protein of unknown function DUF3713 (100%) |  |  |  | 1 | 1 |
| SLECDTLK | Proteobacteria | Bacteria |  |  |  | Proteobacteria |  |  |  |  |  |  |  |  |  |  |  |  |  |  |  |  |  |  |  |  |  |  | Bacteria;Proteobacteria;;;;;;;;;;Proteobacteria |  |  |  | GO:0005840 (100%) | ribosome (100%) | GO:0019843 (97%);GO:0003735 (90%) | rRNA binding (97%);structural constituent of ribosome (90%) | GO:0006412 (98%);GO:0042254 (2%) | translation (98%);ribosome biogenesis (2%) | IPR020069 (100%);IPR036791 (100%);IPR000244 (98%) | Ribosomal protein L9 | C-terminal (100%);Ribosomal protein L9 |  |  | 1 | 100.3923 |
| SQPAADMTHSK | Bacillus subtilis | Bacteria |  |  |  | Firmicutes |  |  | Bacilli |  |  |  | Bacillales |  |  |  |  | Bacillaceae |  |  |  | Bacillus |  | Bacillus subtilis group |  | Bacillus subtilis |  |  | Bacteria;Firmicutes;;;Bacilli;Bacillales;Bacillaceae;;Bacillus;Bacillus subtilis group;Bacillus subtilis;Bacillus subtilis |  |  |  | GO:0016021 (100%);GO:0005886 (14%) | integral component of membrane (100%);plasma membrane (14%) | GO:0022857 (86%);GO:0015171 (14%) | transmembrane transporter activity (86%);amino acid transmembrane transporter activity (14%) | GO:0006865 (86%);GO:0003333 (14%) | amino acid transport (86%);amino acid transmembrane transport (14%) | IPR002293 (100%);IPR004840 (100%);IPR004841 (100%) | Amino acid/polyamine transporter I (100%);Amino acid permease | conserved site (100%);Amino acid permease/ SLC12A domain (100%) |  |  | 1 | 100.1538 |
| SSFDECHYGK | Pasteurella multocida | Bacteria |  |  |  | Proteobacteria |  |  | Gammaproteobacteria |  |  |  | Pasteurellales |  |  |  |  | Pasteurellaceae |  |  |  | Pasteurella |  |  |  | Pasteurella multocida |  |  | Bacteria;Proteobacteria;;;Gammaproteobacteria;Pasteurellales;Pasteurellaceae;;Pasteurella;;Pasteurella multocida;Pasteurella multocida | [K11121](https://www.genome.jp/entry/K11121) | 2.3.1.286 | Protein acetyllysine N-acetyltransferase (100%) | GO:0005737 (86%) | cytoplasm (86%) | GO:0070403 (100%);GO:0034979 (86%);GO:0036054 (86%) | NAD+ binding (100%);NAD-dependent protein deacetylase activity (86%);protein-malonyllysine demalonylase activity (86%) |  |  | IPR003000 (100%);IPR026590 (100%);IPR026591 (100%) | Sirtuin family (100%);Sirtuin family | catalytic core domain (100%);Sirtuin |  |  | 1 | 100.6154 |
| SSPGRPLPMQAFPAQTCAT | Agrobacterium tumefaciens | Bacteria |  |  |  | Proteobacteria |  |  | Alphaproteobacteria |  |  |  | Rhizobiales |  |  |  |  | Rhizobiaceae |  |  |  | Agrobacterium |  | Agrobacterium tumefaciens complex |  | Agrobacterium tumefaciens |  |  | Bacteria;Proteobacteria;;;Alphaproteobacteria;Rhizobiales;Rhizobiaceae;;Agrobacterium;Agrobacterium tumefaciens complex;Agrobacterium tumefaciens;Agrobacterium tumefaciens |  |  |  |  |  |  |  |  |  |  |  |  |  |  | 1 | 100.3846 |
| SYAELGEDVK | Opitutus terrae | Bacteria |  |  |  | Verrucomicrobia |  |  | Opitutae |  |  |  | Opitutales |  |  |  |  | Opitutaceae |  |  |  | Opitutus |  |  |  | Opitutus terrae |  |  | Bacteria;Verrucomicrobia;;;Opitutae;Opitutales;Opitutaceae;;Opitutus;;Opitutus terrae;Opitutus terrae | K02428 | 3.6.1.66 | XTP/dITP diphosphatase (100%) |  |  | GO:0000166 (100%);GO:0017111 (100%);GO:0046872 (100%) | nucleotide binding (100%);nucleoside-triphosphatase activity (100%);metal ion binding (100%) | GO:0009117 (100%);GO:0009146 (100%) | nucleotide metabolic process (100%);purine nucleoside triphosphate catabolic process (100%) | IPR002637 (100%);IPR020922 (100%);IPR029001 (100%) | Ham1-like protein (100%);dITP/XTP pyrophosphatase (100%);Inosine triphosphate pyrophosphatase-like (100%) |  |  |  | 1 | 100.2308 |
| TAESGFDLDQSNTTR | Neisseria meningitidis | Bacteria |  |  |  | Proteobacteria |  |  | Betaproteobacteria |  |  |  | Neisseriales |  |  |  |  | Neisseriaceae |  |  |  | Neisseria |  |  |  | Neisseria meningitidis |  |  | Bacteria;Proteobacteria;;;Betaproteobacteria;Neisseriales;Neisseriaceae;;Neisseria;;Neisseria meningitidis;Neisseria meningitidis |  |  |  | GO:0009279 (100%) | cell outer membrane (100%) |  |  | GO:0071281 (100%) | cellular response to iron ion (100%) | IPR001677 (100%);IPR011250 (100%);IPR035313 (100%) | Transferrin-binding protein | C-lobe/N-lobe beta barrel domain (100%);Outer membrane protein/outer membrane enzyme PagP |  |  | 1 | 100.7769 |
| TANPNGDK | Neisseria meningitidis | Bacteria |  |  |  | Proteobacteria |  |  | Betaproteobacteria |  |  |  | Neisseriales |  |  |  |  | Neisseriaceae |  |  |  | Neisseria |  |  |  | Neisseria meningitidis |  |  | Bacteria;Proteobacteria;;;Betaproteobacteria;Neisseriales;Neisseriaceae;;Neisseria;;Neisseria meningitidis;Neisseria meningitidis |  |  |  | GO:0009279 (100%);GO:0016021 (100%) | cell outer membrane (100%);integral component of membrane (100%) | GO:0015091 (100%) | ferric iron transmembrane transporter activity (100%) |  |  | IPR000531 (100%);IPR010916 (100%);IPR010917 (100%) | TonB-dependent receptor-like | beta-barrel (100%);TonB box |  |  | 1 | 100.3846 |
| TDLDVSENR | Bacteria | Bacteria |  |  |  |  |  |  |  |  |  |  |  |  |  |  |  |  |  |  |  |  |  |  |  |  |  |  | Bacteria;;;;;;;;;;;Bacteria |  |  |  |  |  |  |  |  |  |  |  |  |  |  | 1 | 100.6154 |
| TGFGTGGWQLAAXR | Streptococcus pneumoniae | Bacteria |  |  |  | Firmicutes |  |  | Bacilli |  |  |  | Lactobacillales |  |  |  |  | Streptococcaceae |  |  |  | Streptococcus |  |  |  | Streptococcus pneumoniae |  |  | Bacteria;Firmicutes;;;Bacilli;Lactobacillales;Streptococcaceae;;Streptococcus;;Streptococcus pneumoniae;Streptococcus pneumoniae | [K00364](https://www.genome.jp/entry/K00364) | 1.7.1.7 | GMP reductase (100%) | GO:1902560 (100%) | GMP reductase complex (100%) | GO:0003920 (100%) | GMP reductase activity (100%) | GO:0006163 (100%) | purine nucleotide metabolic process (100%) | IPR001093 (100%);IPR005994 (100%);IPR013785 (100%) | IMP dehydrogenase/GMP reductase (100%);Guanosine monophosphate reductase | type2 (100%);Aldolase-type TIM barrel (100%) |  |  | 1 | 100.3077 |
| TGMFDLDER | Mycobacterium tuberculosis complex | Bacteria |  |  |  | Actinobacteria |  |  | Actinobacteria |  |  |  | Corynebacteriales |  |  |  |  | Mycobacteriaceae |  |  |  | Mycobacterium |  | Mycobacterium tuberculosis complex |  |  |  |  | Bacteria;Actinobacteria;;;Actinobacteria;Corynebacteriales;Mycobacteriaceae;;Mycobacterium;Mycobacterium tuberculosis complex;;Mycobacterium tuberculosis complex | [K00954](https://www.genome.jp/entry/K00954) | 2.7.7.3 | Pantetheine-phosphate adenylyltransferase (100%) | GO:0005737 (100%) | cytoplasm (100%) | GO:0004595 (100%);GO:0005524 (100%) | pantetheine-phosphate adenylyltransferase activity (100%);ATP binding (100%) | GO:0015937 (100%);GO:0034214 (6%) | coenzyme A biosynthetic process (100%);protein hexamerization (6%) | IPR001980 (100%);IPR004821 (100%);IPR014729 (100%) | Phosphopantetheine adenylyltransferase (100%);Cytidyltransferase-like domain (100%);Rossmann-like alpha/beta/alpha sandwich fold (100%) |  |  |  | 1 | 100.2385 |
| TGNLADLVANR | Frankia | Bacteria |  |  |  | Actinobacteria |  |  | Actinobacteria |  |  |  | Frankiales |  |  |  |  | Frankiaceae |  |  |  | Frankia |  |  |  |  |  |  | Bacteria;Actinobacteria;;;Actinobacteria;Frankiales;Frankiaceae;;Frankia;;;Frankia | [K00616](https://www.genome.jp/entry/K00616) | 2.2.1.2 | Transaldolase (100%) | GO:0005737 (100%) | cytoplasm (100%) | GO:0004801 (100%) | sedoheptulose-7-phosphate:D-glyceraldehyde-3-phosphate glyceronetransferase activity (100%) | GO:0005975 (100%);GO:0006098 (100%) | carbohydrate metabolic process (100%);pentose-phosphate shunt (100%) | IPR001585 (100%);IPR004732 (100%);IPR013785 (100%) | Transaldolase/Fructose-6-phosphate aldolase (100%);Transaldolase type 2 (100%);Aldolase-type TIM barrel (100%) |  |  |  | 1 | 100.5308 |
| TVAVSGGSGDSLFDHVR | Streptomyces | Bacteria |  |  |  | Actinobacteria |  |  | Actinobacteria |  |  |  | Streptomycetales |  |  |  |  | Streptomycetaceae |  |  |  | Streptomyces |  |  |  |  |  |  | Bacteria;Actinobacteria;;;Actinobacteria;Streptomycetales;Streptomycetaceae;;Streptomyces;;;Streptomyces |  |  |  | GO:0005737 (100%) | cytoplasm (100%) | GO:0046872 (100%) | metal ion binding (100%) |  |  | IPR002678 (100%);IPR036069 (100%) | GTP cyclohydrolase 1 type 2/Nif3 (100%);GTP cyclohydrolase 1 type 2/Nif3 superfamily (100%) |  |  |  | 1 | 100.4615 |
| TVGEQQVVDEADR | Mycobacteriaceae | Bacteria |  |  |  | Actinobacteria |  |  | Actinobacteria |  |  |  | Corynebacteriales |  |  |  |  | Mycobacteriaceae |  |  |  |  |  |  |  |  |  |  | Bacteria;Actinobacteria;;;Actinobacteria;Corynebacteriales;Mycobacteriaceae;;;;;Mycobacteriaceae | [K15521](https://www.genome.jp/dbget-bin/www_bget?ko:K15521) | 2.4.1.250 | D-inositol-3-phosphate glycosyltransferase (100%) | GO:0005829 (1%) | cytosol (1%) | GO:0008375 (97%);GO:0102710 (96%);GO:0000287 (92%) | acetylglucosaminyltransferase activity (97%);D-inositol-3-phosphate glycosyltransferase activity (96%);magnesium ion binding (92%) | GO:0010125 (97%) | mycothiol biosynthetic process (97%) | IPR028098 (100%);IPR001296 (98%);IPR017814 (98%) | Glycosyltransferase subfamily 4-like | N-terminal domain (100%);Glycosyl transferase |  |  | 1 | 100.5308 |
| TVGETWGLPSPEEAAAAEAEVFATR | Mycobacterium tuberculosis complex | Bacteria |  |  |  | Actinobacteria |  |  | Actinobacteria |  |  |  | Corynebacteriales |  |  |  |  | Mycobacteriaceae |  |  |  | Mycobacterium |  | Mycobacterium tuberculosis complex |  |  |  |  | Bacteria;Actinobacteria;;;Actinobacteria;Corynebacteriales;Mycobacteriaceae;;Mycobacterium;Mycobacterium tuberculosis complex;;Mycobacterium tuberculosis complex |  |  |  | GO:0005576 (100%);GO:0005618 (50%);GO:0005886 (50%) | extracellular region (100%);cell wall (50%);plasma membrane (50%) |  |  | GO:0009405 (100%);GO:0044315 (50%) | pathogenesis (100%);protein secretion by the type VII secretion system (50%) |  |  |  |  |  | 1 | 100.2385 |
| TVLSVTGMK | Streptococcus | Bacteria |  |  |  | Firmicutes |  |  | Bacilli |  |  |  | Lactobacillales |  |  |  |  | Streptococcaceae |  |  |  | Streptococcus |  |  |  |  |  |  | Bacteria;Firmicutes;;;Bacilli;Lactobacillales;Streptococcaceae;;Streptococcus;;;Streptococcus | [K01390](https://www.genome.jp/entry/K01390) | 3.4.24.13 (88%);3.2.1.18 (6%);3.4.24.- (6%) | IgA-specific metalloendopeptidase (88%);Exo-alpha-sialidase (6%);Metalloendopeptidases (6%) | GO:0005576 (94%);GO:0005618 (94%);GO:0016021 (88%) | extracellular region (94%);cell wall (94%);integral component of membrane (88%) | GO:0004222 (94%);GO:0008270 (94%);GO:0008237 (3%) | metalloendopeptidase activity (94%);zinc ion binding (94%);metallopeptidase activity (3%) |  |  | IPR011505 (97%);IPR008006 (91%);IPR011098 (78%) | Peptidase M26 | C-terminal domain (97%);Peptidase M26 |  |  | 1 | 100.5308 |
| TVLSYGTGR | Streptococcus pneumoniae | Bacteria |  |  |  | Firmicutes |  |  | Bacilli |  |  |  | Lactobacillales |  |  |  |  | Streptococcaceae |  |  |  | Streptococcus |  |  |  | Streptococcus pneumoniae |  |  | Bacteria;Firmicutes;;;Bacilli;Lactobacillales;Streptococcaceae;;Streptococcus;;Streptococcus pneumoniae;Streptococcus pneumoniae | [K05366](https://www.genome.jp/entry/K05366+K05365+K05367+K12555+K18770+K21464+2.4.1.129) | 2.4.1.129 ;3.4.16.4 | Peptidoglycan glycosyltransferase (100%);Serine-type D-Ala-D-Ala carboxypeptidase (100%) | GO:0016021 (34%);GO:0005576 (1%) | integral component of membrane (34%);extracellular region (1%) | GO:0008658 (97%);GO:0016740 (72%);GO:0008144 (1%) | penicillin binding (97%);transferase activity (72%);drug binding (1%) | GO:0008360 (1%);GO:0009252 (1%);GO:0046677 (1%) | regulation of cell shape (1%);peptidoglycan biosynthetic process (1%);response to antibiotic (1%) | IPR001460 (100%);IPR012338 (100%);IPR001264 (74%) | Penicillin-binding protein | transpeptidase (100%);Beta-lactamase/transpeptidase-like (100%);Glycosyl transferase |  |  | 1 | 100.9231 |
| VADGAALWR | Actinobacteria | Bacteria |  |  |  | Actinobacteria |  |  | Actinobacteria |  |  |  |  |  |  |  |  |  |  |  |  |  |  |  |  |  |  |  | Bacteria;Actinobacteria;;;Actinobacteria;;;;;;;Actinobacteria | [K06718](https://www.genome.jp/entry/K06718) | 2.3.1.178 | Diaminobutyrate acetyltransferase (100%) |  |  | GO:0033816 (95%);GO:0008080 (92%);GO:0004672 (3%) | diaminobutyrate acetyltransferase activity (95%);N-acetyltransferase activity (92%);protein kinase activity (3%) | GO:0019491 (95%) | ectoine biosynthetic process (95%) | IPR000182 (93%);IPR012772 (93%);IPR016181 (93%) | GNAT domain (93%);L-2 | 4-diaminobutyric acid acetyltransferase (93%);Acyl-CoA N-acyltransferase (93%) |  |  | 1 | 100.6077 |
| VAFLVEAK | Proteobacteria | Bacteria |  |  |  | Proteobacteria |  |  |  |  |  |  |  |  |  |  |  |  |  |  |  |  |  |  |  |  |  |  | Bacteria;Proteobacteria;;;;;;;;;;Proteobacteria |  |  |  | GO:0005737 (100%) | cytoplasm (100%) | GO:0051082 (96%) | unfolded protein binding (96%) | GO:0006457 (100%);GO:0015031 (100%);GO:0051262 (96%) | protein folding (100%);protein transport (100%);protein tetramerization (96%) | IPR003708 (100%);IPR035958 (100%) | Bacterial protein export chaperone SecB (100%);SecB-like superfamily (100%) |  |  |  | 1 | 1 |
| VALWFQR | Bacillus subtilis | Bacteria |  |  |  | Firmicutes |  |  | Bacilli |  |  |  | Bacillales |  |  |  |  | Bacillaceae |  |  |  | Bacillus |  | Bacillus subtilis group |  | Bacillus subtilis |  |  | Bacteria;Firmicutes;;;Bacilli;Bacillales;Bacillaceae;;Bacillus;Bacillus subtilis group;Bacillus subtilis;Bacillus subtilis |  |  |  | GO:0016021 (100%);GO:0005886 (8%) | integral component of membrane (100%);plasma membrane (8%) | GO:0015297 (92%);GO:0015103 (83%);GO:0015104 (8%) | antiporter activity (92%);inorganic anion transmembrane transporter activity (83%);antimonite transmembrane transporter activity (8%) | GO:0015699 (8%);GO:0015700 (8%);GO:0046685 (8%) | antimonite transport (8%);arsenite transport (8%);response to arsenic-containing substance (8%) | IPR002657 (100%);IPR004706 (100%);IPR038770 (100%) | Bile acid:sodium symporter/arsenical resistance protein Acr3 (100%);Arsenical-resistance protein Acr3 (100%);Sodium/solute symporter superfamily (100%) |  |  |  | 1 | 100.5385 |
| VASDEDEASPDVR | Rhodococcus | Bacteria |  |  |  | Actinobacteria |  |  | Actinobacteria |  |  |  | Corynebacteriales |  |  |  |  | Nocardiaceae |  |  |  | Rhodococcus |  |  |  |  |  |  | Bacteria;Actinobacteria;;;Actinobacteria;Corynebacteriales;Nocardiaceae;;Rhodococcus;;;Rhodococcus |  |  |  | GO:0005737 (100%) | cytoplasm (100%) |  |  | GO:0030490 (100%) | maturation of SSU-rRNA (100%) | IPR000238 (100%);IPR015946 (100%);IPR020053 (100%) | Ribosome-binding factor A (100%);K homology domain-like | alpha/beta (100%);Ribosome-binding factor A |  |  | 1 | 100.5308 |
| VAVALNKP | Myxococcus | Bacteria |  |  |  | Proteobacteria | delta/epsilon subdivisions |  | Deltaproteobacteria |  |  |  | Myxococcales | Cystobacterineae |  |  |  | Myxococcaceae |  |  |  | Myxococcus |  |  |  |  |  |  | Bacteria;Proteobacteria;delta/epsilon subdivisions;;Deltaproteobacteria;Myxococcales;Myxococcaceae;;Myxococcus;;;Myxococcus |  |  |  |  |  | GO:0003723 (100%) | RNA binding (100%) | GO:0006353 (100%);GO:0031564 (100%) | DNA-templated transcription | termination (100%);transcription antitermination (100%) | IPR006027 (100%);IPR011605 (100%);IPR035926 (100%) | NusB/RsmB/TIM44 (100%);NusB antitermination factor (100%);NusB-like superfamily (100%) |  |  | 1 | 1 |
| VDDPDAVVHMR | Mycobacterium | Bacteria |  |  |  | Actinobacteria |  |  | Actinobacteria |  |  |  | Corynebacteriales |  |  |  |  | Mycobacteriaceae |  |  |  | Mycobacterium |  |  |  |  |  |  | Bacteria;Actinobacteria;;;Actinobacteria;Corynebacteriales;Mycobacteriaceae;;Mycobacterium;;;Mycobacterium | [K08048](https://www.genome.jp/dbget-bin/www_bget?ko:K08048) | 4.6.1.1 | Adenylate cyclase (100%) |  |  | GO:0016849 (87%);GO:0004016 (13%);GO:0005524 (3%) | phosphorus-oxygen lyase activity (87%);adenylate cyclase activity (13%);ATP binding (3%) | GO:0035556 (99%);GO:0009190 (96%);GO:0006171 (3%) | intracellular signal transduction (99%);cyclic nucleotide biosynthetic process (96%);cAMP biosynthetic process (3%) | IPR032026 (100%);IPR001054 (99%);IPR029787 (99%) | Adenylate cyclase regulatory domain (100%);Adenylyl cyclase class-3/4/guanylyl cyclase (99%);Nucleotide cyclase (99%) |  |  |  | 1 | 100.4 |
| VEALVGLDALR | Nocardiopsaceae | Bacteria |  |  |  | Actinobacteria |  |  | Actinobacteria |  |  |  | Streptosporangiales |  |  |  |  | Nocardiopsaceae |  |  |  |  |  |  |  |  |  |  | Bacteria;Actinobacteria;;;Actinobacteria;Streptosporangiales;Nocardiopsaceae;;;;;Nocardiopsaceae | [K01872](https://www.genome.jp/entry/K01872) | 6.1.1.7 | Alanine--tRNA ligase (100%) | GO:0005737 (100%) | cytoplasm (100%) | GO:0000049 (100%);GO:0004813 (100%);GO:0005524 (100%) | tRNA binding (100%);alanine-tRNA ligase activity (100%);ATP binding (100%) | GO:0006419 (100%) | alanyl-tRNA aminoacylation (100%) | IPR002318 (100%);IPR003156 (100%);IPR009000 (100%) | Alanine-tRNA ligase | class IIc (100%);DHHA1 domain (100%);Translation protein |  |  | 1 | 1 |
| VESEFTGWSETSR | Mycoplasma mycoides group | Bacteria |  |  |  | Tenericutes |  |  | Mollicutes |  |  |  | Mycoplasmatales |  |  |  |  | Mycoplasmataceae |  |  |  | Mycoplasma |  | Mycoplasma mycoides group |  |  |  |  | Bacteria;Tenericutes;;;Mollicutes;Mycoplasmatales;Mycoplasmataceae;;Mycoplasma;Mycoplasma mycoides group;;Mycoplasma mycoides group | [K01872](https://www.genome.jp/entry/K01872) | 6.1.1.7 | Alanine--tRNA ligase (100%) | GO:0005737 (100%) | cytoplasm (100%) | GO:0000049 (100%);GO:0004813 (100%);GO:0005524 (100%) | tRNA binding (100%);alanine-tRNA ligase activity (100%);ATP binding (100%) | GO:0006419 (100%) | alanyl-tRNA aminoacylation (100%) | IPR002318 (100%);IPR009000 (100%);IPR012947 (100%) | Alanine-tRNA ligase | class IIc (100%);Translation protein |  |  | 1 | 100.8462 |
| VEVVNAATEDEAVQVGR | Streptomyces | Bacteria |  |  |  | Actinobacteria |  |  | Actinobacteria |  |  |  | Streptomycetales |  |  |  |  | Streptomycetaceae |  |  |  | Streptomyces |  |  |  |  |  |  | Bacteria;Actinobacteria;;;Actinobacteria;Streptomycetales;Streptomycetaceae;;Streptomyces;;;Streptomyces | [K00618](https://www.genome.jp/entry/K00618) | 2.3.1.1 ;2.3.1.35 | Amino-acid N-acetyltransferase (100%);Glutamate N-acetyltransferase (100%) | GO:0005737 (100%) | cytoplasm (100%) | GO:0004042 (100%);GO:0004358 (100%);GO:0103045 (100%) | acetyl-CoA:L-glutamate N-acetyltransferase activity (100%);glutamate N-acetyltransferase activity (100%);methione N-acyltransferase activity (100%) | GO:0006526 (100%);GO:0006592 (6%) | arginine biosynthetic process (100%);ornithine biosynthetic process (6%) | IPR002813 (100%);IPR016117 (100%);IPR042195 (100%) | Arginine biosynthesis protein ArgJ (100%);ArgJ-like domain superfamily (100%);ArgJ beta chain | C-terminal domain (100%) |  |  | 1 | 100.6231 |
| VFTFFTR | Micrococcales | Bacteria |  |  |  | Actinobacteria |  |  | Actinobacteria |  |  |  | Micrococcales |  |  |  |  |  |  |  |  |  |  |  |  |  |  |  | Bacteria;Actinobacteria;;;Actinobacteria;Micrococcales;;;;;;Micrococcales | K01866 | 6.1.1.1 | Tyrosine--tRNA ligase (100%) | GO:0005737 (100%) | cytoplasm (100%) | GO:0003723 (100%);GO:0004831 (100%);GO:0005524 (100%) | RNA binding (100%);tyrosine-tRNA ligase activity (100%);ATP binding (100%) | GO:0006437 (100%) | tyrosyl-tRNA aminoacylation (100%) | IPR001412 (100%);IPR002305 (100%);IPR002307 (100%) | Aminoacyl-tRNA synthetase | class I |  |  | 1 | 100.8462 |
| VGLVPQEPR | Bacteria | Bacteria |  |  |  |  |  |  |  |  |  |  |  |  |  |  |  |  |  |  |  |  |  |  |  |  |  |  | Bacteria;;;;;;;;;;;Bacteria |  | 3.6.3.- (83%);3.6.3.31 (10%);3.6.3.17 (7%) | Acting on acid anhydrides; catalyzing transmembrane movement of substances (83%);Transferred entry: 7.6.2.11 (10%);Transferred entry: 7.5.2.8 (7%) | GO:0005886 (1%);GO:0016021 (1%) | plasma membrane (1%);integral component of membrane (1%) | GO:0005524 (100%);GO:0016887 (92%);GO:0015407 (1%) | ATP binding (100%);ATPase activity (92%);ATPase-coupled monosaccharide transmembrane transporter activity (1%) | GO:0055085 (0%) | transmembrane transport (0%) | IPR027417 (100%);IPR003439 (99%);IPR003593 (99%) | P-loop containing nucleoside triphosphate hydrolase (100%);ABC transporter-like (99%);AAA+ ATPase domain (99%) |  |  |  | 1 | 100.1538 |
| VGQELSLLAQK | Streptococcus pneumoniae | Bacteria |  |  |  | Firmicutes |  |  | Bacilli |  |  |  | Lactobacillales |  |  |  |  | Streptococcaceae |  |  |  | Streptococcus |  |  |  | Streptococcus pneumoniae |  |  | Bacteria;Firmicutes;;;Bacilli;Lactobacillales;Streptococcaceae;;Streptococcus;;Streptococcus pneumoniae;Streptococcus pneumoniae |  |  |  | GO:0016021 (100%);GO:0005886 (33%) | integral component of membrane (100%);plasma membrane (33%) |  |  | GO:0030420 (33%) | establishment of competence for transformation (33%) | IPR005696 (100%) | Secretion protein MesE/LcnD (100%) |  |  |  | 1 | 1 |
| VGSGAFMPTAEK | Proteobacteria | Bacteria |  |  |  | Proteobacteria |  |  |  |  |  |  |  |  |  |  |  |  |  |  |  |  |  |  |  |  |  |  | Bacteria;Proteobacteria;;;;;;;;;;Proteobacteria | [K00758](https://www.genome.jp/entry/K00758) | 2.4.2.4 | Thymidine phosphorylase (100%) |  |  | GO:0009032 (100%);GO:0004645 (98%);GO:0016154 (98%) | thymidine phosphorylase activity (100%);1 | 4-alpha-oligoglucan phosphorylase activity (98%);pyrimidine-nucleoside phosphorylase activity (98%) | GO:0006206 (98%);GO:0046104 (98%);GO:0006213 (2%) | pyrimidine nucleobase metabolic process (98%);thymidine metabolic process (98%);pyrimidine nucleoside metabolic process (2%) | IPR000053 (100%);IPR000312 (100%);IPR013465 (100%) | Thymidine/pyrimidine-nucleoside phosphorylase (100%);Glycosyl transferase |  |  | 1 | 1 |
| VGVLGPTNAGK | Mycoplasma genitalium | Bacteria |  |  |  | Tenericutes |  |  | Mollicutes |  |  |  | Mycoplasmatales |  |  |  |  | Mycoplasmataceae |  |  |  | Mycoplasma |  |  |  | Mycoplasma genitalium |  |  | Bacteria;Tenericutes;;;Mollicutes;Mycoplasmatales;Mycoplasmataceae;;Mycoplasma;;Mycoplasma genitalium;Mycoplasma genitalium |  |  |  | GO:0005829 (100%);GO:0005886 (100%) | cytosol (100%);plasma membrane (100%) | GO:0003924 (100%);GO:0005525 (100%);GO:0019843 (100%) | GTPase activity (100%);GTP binding (100%);rRNA binding (100%) | GO:0000028 (100%) | ribosomal small subunit assembly (100%) | IPR004044 (100%);IPR005225 (100%);IPR005662 (100%) | K Homology domain | type 2 (100%);Small GTP-binding protein domain (100%);GTP-binding protein Era (100%) |  |  | 1 | 100.7769 |
| VLFHTGK | Bacteria | Bacteria |  |  |  |  |  |  |  |  |  |  |  |  |  |  |  |  |  |  |  |  |  |  |  |  |  |  | Bacteria;;;;;;;;;;;Bacteria | K02320 | 2.7.7.7 (50%);3.5.1.47 (50%) | DNA-directed DNA polymerase (50%);N-acetyldiaminopimelate deacetylase (50%) | GO:0005737 (88%) | cytoplasm (88%) | GO:0003746 (88%);GO:0003924 (88%);GO:0005525 (88%) | translation elongation factor activity (88%);GTPase activity (88%);GTP binding (88%) | GO:0006261 (6%);GO:0006281 (6%) | DNA-dependent DNA replication (6%);DNA repair (6%) | IPR000795 (82%);IPR004161 (82%);IPR004540 (82%) | Transcription factor | GTP-binding domain (82%);Translation elongation factor EFTu-like |  |  | 1 | 100.8462 |
| VLSPEAESVLK | Synechococcus | Bacteria |  |  |  | Cyanobacteria |  |  |  |  |  |  | Synechococcales |  |  |  |  | Synechococcaceae |  |  |  | Synechococcus |  |  |  |  |  |  | Bacteria;Cyanobacteria;;;;Synechococcales;Synechococcaceae;;Synechococcus;;;Synechococcus | [K02111](https://www.genome.jp/entry/K02111) | 7.1.2.2 | H(+)-transporting two-sector ATPase (100%) | GO:0042651 (100%);GO:0045261 (100%) | thylakoid membrane (100%);proton-transporting ATP synthase complex | catalytic core F(1) (100%) | GO:0005524 (100%);GO:0046933 (100%) | ATP binding (100%);proton-transporting ATP synthase activity | rotational mechanism (100%) | GO:0015986 (100%) | ATP synthesis coupled proton transport (100%) | IPR000194 (100%);IPR000793 (100%);IPR004100 (100%) |  |  | 1 | 1 |
| VNPSDYDYTK | Bacillales | Bacteria |  |  |  | Firmicutes |  |  | Bacilli |  |  |  | Bacillales |  |  |  |  |  |  |  |  |  |  |  |  |  |  |  | Bacteria;Firmicutes;;;Bacilli;Bacillales;;;;;;Bacillales |  | 2.7.8.- | Transferases for other substituted phosphate groups (100%) | GO:0005886 (97%);GO:0016021 (97%);GO:0005576 (3%) | plasma membrane (97%);integral component of membrane (97%);extracellular region (3%) | GO:0008484 (84%);GO:0046872 (77%);GO:0003824 (3%) | sulfuric ester hydrolase activity (84%);metal ion binding (77%);catalytic activity (3%) | GO:0070395 (3%);GO:0071555 (3%) | lipoteichoic acid biosynthetic process (3%);cell wall organization (3%) | IPR017850 (100%);IPR000917 (97%);IPR012160 (97%) | Alkaline-phosphatase-like | core domain superfamily (100%);Sulfatase |  |  | 1 | 100.3846 |
| VPVPTGSATDLVVELQR | Streptomyces | Bacteria |  |  |  | Actinobacteria |  |  | Actinobacteria |  |  |  | Streptomycetales |  |  |  |  | Streptomycetaceae |  |  |  | Streptomyces |  |  |  |  |  |  | Bacteria;Actinobacteria;;;Actinobacteria;Streptomycetales;Streptomycetaceae;;Streptomyces;;;Streptomyces | [K00134](https://www.genome.jp/entry/K00134) | 1.2.1.12 | Glyceraldehyde-3-phosphate dehydrogenase (phosphorylating) (100%) | GO:0005737 (2%) | cytoplasm (2%) | GO:0050661 (94%);GO:0051287 (94%);GO:0016620 (82%) | NADP binding (94%);NAD binding (94%);oxidoreductase activity | acting on the aldehyde or oxo group of donors | NAD or NADP as acceptor (82%) | GO:0006006 (94%);GO:0006096 (2%) | glucose metabolic process (94%);glycolytic process (2%) | IPR006424 (100%);IPR020828 (100%);IPR020829 (100%) |  |  | 1 | 100.5308 |
| VSEASTNTSK | Bacillus subtilis | Bacteria |  |  |  | Firmicutes |  |  | Bacilli |  |  |  | Bacillales |  |  |  |  | Bacillaceae |  |  |  | Bacillus |  | Bacillus subtilis group |  | Bacillus subtilis |  |  | Bacteria;Firmicutes;;;Bacilli;Bacillales;Bacillaceae;;Bacillus;Bacillus subtilis group;Bacillus subtilis;Bacillus subtilis |  | 3.1.-.- | Acting on ester bonds (100%) |  |  | GO:0004518 (100%) | nuclease activity (100%) |  |  | IPR006829 (100%);IPR026835 (100%);IPR027797 (100%) | LXG domain (100%);UPF0720 protein YqcG (100%);Pre-toxin TG domain (100%) |  |  |  | 1 | 100.8462 |
| VSFSFPSQESDQTNR | Bacillus | Bacteria |  |  |  | Firmicutes |  |  | Bacilli |  |  |  | Bacillales |  |  |  |  | Bacillaceae |  |  |  | Bacillus |  |  |  |  |  |  | Bacteria;Firmicutes;;;Bacilli;Bacillales;Bacillaceae;;Bacillus;;;Bacillus |  |  |  | GO:0016021 (94%);GO:0005886 (3%) | integral component of membrane (94%);plasma membrane (3%) | GO:0005524 (100%);GO:0003677 (85%) | ATP binding (100%);DNA binding (85%) | GO:0051301 (15%) | cell division (15%) | IPR002543 (100%);IPR022206 (100%);IPR023839 (100%) | FtsK domain (100%);Firmicutes EssC | N-terminal (100%);Firmicutes EssC |  |  | 1 | 100.8462 |
| VSSSGPNSTPLPAAR | Mycobacterium tuberculosis | Bacteria |  |  |  | Actinobacteria |  |  | Actinobacteria |  |  |  | Corynebacteriales |  |  |  |  | Mycobacteriaceae |  |  |  | Mycobacterium |  | Mycobacterium tuberculosis complex |  | Mycobacterium tuberculosis |  |  | Bacteria;Actinobacteria;;;Actinobacteria;Corynebacteriales;Mycobacteriaceae;;Mycobacterium;Mycobacterium tuberculosis complex;Mycobacterium tuberculosis;Mycobacterium tuberculosis |  |  |  | GO:0016021 (67%);GO:0005886 (33%) | integral component of membrane (67%);plasma membrane (33%) |  |  |  |  |  |  |  |  |  | 1 | 100.5385 |
| VVVVEDTSTTGGSPLTAVEAVR | Streptomyces | Bacteria |  |  |  | Actinobacteria |  |  | Actinobacteria |  |  |  | Streptomycetales |  |  |  |  | Streptomycetaceae |  |  |  | Streptomyces |  |  |  |  |  |  | Bacteria;Actinobacteria;;;Actinobacteria;Streptomycetales;Streptomycetaceae;;Streptomyces;;;Streptomyces | [K00762](https://www.genome.jp/entry/K00762) | 2.4.2.10 | Orotate phosphoribosyltransferase (100%) |  |  | GO:0000287 (100%);GO:0004588 (100%) | magnesium ion binding (100%);orotate phosphoribosyltransferase activity (100%) | GO:0044205 (100%);GO:0009116 (85%);GO:0006222 (5%) | 'de novo' UMP biosynthetic process (100%);nucleoside metabolic process (85%);UMP biosynthetic process (5%) | IPR000836 (100%);IPR004467 (100%);IPR023031 (100%) | Phosphoribosyltransferase domain (100%);Orotate phosphoribosyl transferase domain (100%);Orotate phosphoribosyltransferase (100%) |  |  |  | 1 | 1 |
| WEHVMSEDVER | Actinobacteria | Bacteria |  |  |  | Actinobacteria |  |  | Actinobacteria |  |  |  |  |  |  |  |  |  |  |  |  |  |  |  |  |  |  |  | Bacteria;Actinobacteria;;;Actinobacteria;;;;;;;Actinobacteria |  |  |  | GO:0005737 (1%);GO:0005618 (0%);GO:0005886 (0%) | cytoplasm (1%);cell wall (0%);plasma membrane (0%) | GO:0003700 (100%);GO:0046983 (100%);GO:0003677 (100%) | DNA-binding transcription factor activity (100%);protein dimerization activity (100%);DNA binding (100%) | GO:0006355 (0%);GO:0006979 (0%);GO:0019540 (0%) | regulation of transcription | DNA-templated (0%);response to oxidative stress (0%);siderophore biosynthetic process from catechol (0%) | IPR001367 (100%);IPR022689 (100%);IPR036421 (100%) | Iron dependent repressor |  |  | 1 | 100.5462 |
| WNLQGFELHHYAGR | Neisseria | Bacteria |  |  |  | Proteobacteria |  |  | Betaproteobacteria |  |  |  | Neisseriales |  |  |  |  | Neisseriaceae |  |  |  | Neisseria |  |  |  |  |  |  | Bacteria;Proteobacteria;;;Betaproteobacteria;Neisseriales;Neisseriaceae;;Neisseria;;;Neisseria | K04075 | 6.3.4.19 | tRNA(Ile)-lysidine synthetase (100%) | GO:0005737 (100%) | cytoplasm (100%) | GO:0005524 (100%);GO:0016879 (100%) | ATP binding (100%);ligase activity | forming carbon-nitrogen bonds (100%) | GO:0006400 (96%);GO:0008033 (4%) | tRNA modification (96%);tRNA processing (4%) | IPR011063 (100%);IPR012094 (100%);IPR012795 (100%) | tRNA(Ile)-lysidine/2-thiocytidine synthase |  |  | 1 | 100.5385 |
| WQLFAELGTTDK | Mycoplasma genitalium | Bacteria |  |  |  | Tenericutes |  |  | Mollicutes |  |  |  | Mycoplasmatales |  |  |  |  | Mycoplasmataceae |  |  |  | Mycoplasma |  |  |  | Mycoplasma genitalium |  |  | Bacteria;Tenericutes;;;Mollicutes;Mycoplasmatales;Mycoplasmataceae;;Mycoplasma;;Mycoplasma genitalium;Mycoplasma genitalium |  |  |  | GO:0005886 (100%);GO:0016021 (100%) | plasma membrane (100%);integral component of membrane (100%) |  |  |  |  | IPR003838 (100%) | ABC transporter permease protein domain (100%) |  |  |  | 1 | 100.6077 |
| YCGVGCGVMVAAK | Symbiobacterium thermophilum | Bacteria |  |  |  | Firmicutes |  |  | Clostridia |  |  |  | Clostridiales |  |  |  |  | Symbiobacteriaceae |  |  |  | Symbiobacterium |  |  |  | Symbiobacterium thermophilum |  |  | Bacteria;Firmicutes;;;Clostridia;Clostridiales;Symbiobacteriaceae;;Symbiobacterium;;Symbiobacterium thermophilum;Symbiobacterium thermophilum | [K02567](https://www.genome.jp/entry/K02567) | 1.9.6.1 | Nitrate reductase (cytochrome) (100%) | GO:0005576 (100%) | extracellular region (100%) | GO:0005506 (100%);GO:0008940 (100%);GO:0009055 (100%) | iron ion binding (100%);nitrate reductase activity (100%);electron transfer activity (100%) | GO:0006777 (100%);GO:0042128 (100%) | Mo-molybdopterin cofactor biosynthetic process (100%);nitrate assimilation (100%) | IPR006311 (100%);IPR006656 (100%);IPR006657 (100%) | Twin-arginine translocation pathway | signal sequence (100%);Molybdopterin oxidoreductase (100%);Molybdopterin dinucleotide-binding domain (100%) |  |  | 1 | 100.4615 |
| YDTSATLDEMK | Salmonella | Bacteria |  |  |  | Proteobacteria |  |  | Gammaproteobacteria |  |  |  | Enterobacterales |  |  |  |  | Enterobacteriaceae |  |  |  | Salmonella |  |  |  |  |  |  | Bacteria;Proteobacteria;;;Gammaproteobacteria;Enterobacterales;Enterobacteriaceae;;Salmonella;;;Salmonella |  |  |  | GO:0009279 (98%);GO:0046930 (93%);GO:0016021 (3%) | cell outer membrane (98%);pore complex (93%);integral component of membrane (3%) | GO:0015420 (98%);GO:0015288 (93%);GO:0046872 (93%) | ATPase-coupled vitamin B12 transmembrane transporter activity (98%);porin activity (93%);metal ion binding (93%) | GO:0006811 (93%);GO:0015685 (0%);GO:0015889 (0%) | ion transport (93%);ferric-enterobactin import into cell (0%);cobalamin transport (0%) | IPR036942 (100%);IPR039426 (100%);IPR000531 (99%) | TonB-dependent receptor-like | beta-barrel domain superfamily (100%);Vitamin B12 transporter BtuB-like (100%);TonB-dependent receptor-like |  |  | 1 | 100.4615 |
| YEFTGTK | Bacteria | Bacteria |  |  |  |  |  |  |  |  |  |  |  |  |  |  |  |  |  |  |  |  |  |  |  |  |  |  | Bacteria;;;;;;;;;;;Bacteria |  |  |  |  |  | GO:0008168 (100%);GO:0003676 (83%) | methyltransferase activity (100%);nucleic acid binding (83%) | GO:0032259 (100%) | methylation (100%) | IPR029063 (100%);IPR002052 (89%);IPR001525 (9%) | S-adenosyl-L-methionine-dependent methyltransferase (100%);DNA methylase | N-6 adenine-specific |  |  | 1 | 100.6077 |
| YEVELGPGVK | Bacteria | Bacteria |  |  |  |  |  |  |  |  |  |  |  |  |  |  |  |  |  |  |  |  |  |  |  |  |  |  | Bacteria;;;;;;;;;;;Bacteria |  |  |  | GO:0016021 (98%);GO:0005886 (62%);GO:0005576 (0%) | integral component of membrane (98%);plasma membrane (62%);extracellular region (0%) | GO:0003677 (100%);GO:0005524 (100%);GO:0004519 (1%) | DNA binding (100%);ATP binding (100%);endonuclease activity (1%) | GO:0051301 (82%);GO:0007049 (62%);GO:0007059 (62%) | cell division (82%);cell cycle (62%);chromosome segregation (62%) | IPR041027 (100%);IPR002543 (100%);IPR027417 (100%) | FtsK alpha domain (100%);FtsK domain (100%);P-loop containing nucleoside triphosphate hydrolase (100%) |  |  |  | 1 | 1 |
| YGELDVVSR | Bifidobacterium | Bacteria |  |  |  | Actinobacteria |  |  | Actinobacteria |  |  |  | Bifidobacteriales |  |  |  |  | Bifidobacteriaceae |  |  |  | Bifidobacterium |  |  |  |  |  |  | Bacteria;Actinobacteria;;;Actinobacteria;Bifidobacteriales;Bifidobacteriaceae;;Bifidobacterium;;;Bifidobacterium |  |  |  |  |  | GO:0003676 (100%);GO:0004518 (55%);GO:0004519 (45%) | nucleic acid binding (100%);nuclease activity (55%);endonuclease activity (45%) |  |  | IPR003509 (100%);IPR011335 (100%);IPR011856 (100%) | Uncharacterised protein family UPF0102 (100%);Restriction endonuclease type II-like (100%);tRNA endonuclease-like domain superfamily (100%) |  |  |  | 1 | 100.7692 |
| YGETLAAR | Bacteria | Bacteria |  |  |  |  |  |  |  |  |  |  |  |  |  |  |  |  |  |  |  |  |  |  |  |  |  |  | Bacteria;;;;;;;;;;;Bacteria |  |  |  |  |  | GO:0003676 (94%);GO:0004518 (78%);GO:0004519 (20%) | nucleic acid binding (94%);nuclease activity (78%);endonuclease activity (20%) |  |  | IPR003509 (99%);IPR011335 (99%);IPR011856 (99%) | Uncharacterised protein family UPF0102 (99%);Restriction endonuclease type II-like (99%);tRNA endonuclease-like domain superfamily (99%) |  |  |  | 1 | 100.5308 |
| YHNHFNK | Mycoplasma gallisepticum | Bacteria |  |  |  | Tenericutes |  |  | Mollicutes |  |  |  | Mycoplasmatales |  |  |  |  | Mycoplasmataceae |  |  |  | Mycoplasma |  |  |  | Mycoplasma gallisepticum |  |  | Bacteria;Tenericutes;;;Mollicutes;Mycoplasmatales;Mycoplasmataceae;;Mycoplasma;;Mycoplasma gallisepticum;Mycoplasma gallisepticum |  | 2.1.1.199 | 16S rRNA (cytosine(1402)-N(4))-methyltransferase (100%) | GO:0005737 (100%) | cytoplasm (100%) | GO:0071424 (100%) | rRNA (cytosine-N4-)-methyltransferase activity (100%) | GO:0070475 (100%) | rRNA base methylation (100%) | IPR002903 (100%);IPR023397 (100%);IPR029063 (100%) | Ribosomal RNA small subunit methyltransferase H (100%);S-adenosyl-L-methionine-dependent methyltransferase | MraW |  |  | 1 | 100.9231 |
| YKPVALLAVAGGGK | Bacillaceae | Bacteria |  |  |  | Firmicutes |  |  | Bacilli |  |  |  | Bacillales |  |  |  |  | Bacillaceae |  |  |  |  |  |  |  |  |  |  | Bacteria;Firmicutes;;;Bacilli;Bacillales;Bacillaceae;;;;;Bacillaceae | [K03206](https://www.genome.jp/entry/K03206) | 1.7.-.- (50%);1.7.1.6 (50%) | Acting on other nitrogenous compounds as donors (50%);Azobenzene reductase (50%) | GO:0005829 (4%) | cytosol (4%) | GO:0016491 (96%);GO:0010181 (4%);GO:0042802 (4%) | oxidoreductase activity (96%);FMN binding (4%);identical protein binding (4%) |  |  | IPR005025 (100%);IPR029039 (100%) | NADPH-dependent FMN reductase-like (100%);Flavoprotein-like superfamily (100%) |  |  |  | 1 | 100.4692 |
| YLGTEQEFK | Bacteria | Bacteria |  |  |  |  |  |  |  |  |  |  |  |  |  |  |  |  |  |  |  |  |  |  |  |  |  |  | Bacteria;;;;;;;;;;;Bacteria | [K01176](https://www.genome.jp/entry/K01176) | 3.2.1.1 | Alpha-amylase (100%) | GO:0005576 (1%) | extracellular region (1%) | GO:0004556 (98%);GO:0103025 (98%);GO:0043169 (85%) | alpha-amylase activity (98%);alpha-amylase activity (releasing maltohexaose) (98%);cation binding (85%) | GO:0005975 (99%);GO:0008152 (1%) | carbohydrate metabolic process (99%);metabolic process (1%) | IPR017853 (100%);IPR006047 (99%);IPR006046 (97%) | Glycoside hydrolase superfamily (100%);Glycosyl hydrolase | family 13 |  |  | 1 | 100.5385 |
| YPQQASLNLLR | Bacteria | Bacteria |  |  |  |  |  |  |  |  |  |  |  |  |  |  |  |  |  |  |  |  |  |  |  |  |  |  | Bacteria;;;;;;;;;;;Bacteria |  | 3.1.-.- | Acting on ester bonds (100%) |  |  | GO:0004519 (95%);GO:0016787 (5%);GO:0004520 (0%) | endonuclease activity (95%);hydrolase activity (5%);endodeoxyribonuclease activity (0%) |  |  | IPR002625 (100%);IPR036063 (100%) | Smr domain (100%);Smr domain superfamily (100%) |  |  |  | 1 | 1 |
| YQDVVGEVAR | Neisseriaceae | Bacteria |  |  |  | Proteobacteria |  |  | Betaproteobacteria |  |  |  | Neisseriales |  |  |  |  | Neisseriaceae |  |  |  |  |  |  |  |  |  |  | Bacteria;Proteobacteria;;;Betaproteobacteria;Neisseriales;Neisseriaceae;;;;;Neisseriaceae | [K00796](https://www.genome.jp/entry/K00796) | 2.5.1.15 | Dihydropteroate synthase (100%) | GO:0005829 (2%) | cytosol (2%) | GO:0004156 (100%);GO:0046872 (97%) | dihydropteroate synthase activity (100%);metal ion binding (97%) | GO:0046654 (97%);GO:0046656 (97%) | tetrahydrofolate biosynthetic process (97%);folic acid biosynthetic process (97%) | IPR000489 (100%);IPR006390 (100%);IPR011005 (100%) | Pterin-binding domain (100%);Dihydropteroate synthase (100%);Dihydropteroate synthase-like (100%) |  |  |  | 1 | 1 |
| YTGCSPTSYMK | Bacillus | Bacteria |  |  |  | Firmicutes |  |  | Bacilli |  |  |  | Bacillales |  |  |  |  | Bacillaceae |  |  |  | Bacillus |  |  |  |  |  |  | Bacteria;Firmicutes;;;Bacilli;Bacillales;Bacillaceae;;Bacillus;;;Bacillus |  |  |  | GO:0005737 (6%) | cytoplasm (6%) | GO:0003700 (94%);GO:0043565 (94%);GO:0003677 (6%) | DNA-binding transcription factor activity (94%);sequence-specific DNA binding (94%);DNA binding (6%) |  |  | IPR002491 (100%);IPR009057 (100%);IPR018060 (100%) | ABC transporter periplasmic binding domain (100%);Homeobox-like domain superfamily (100%);DNA binding HTH domain | AraC-type (100%) |  |  | 1 | 100.5308 |
| YYQDETVVR | Bacteria | Bacteria |  |  |  |  |  |  |  |  |  |  |  |  |  |  |  |  |  |  |  |  |  |  |  |  |  |  | Bacteria;;;;;;;;;;;Bacteria | [K01727](https://www.genome.jp/entry/K01727) | 4.2.2.1 | Hyaluronate lyase (100%) | GO:0005576 (99%);GO:0005618 (93%);GO:0016021 (81%) | extracellular region (99%);cell wall (93%);integral component of membrane (81%) | GO:0030246 (90%);GO:0030340 (59%);GO:0016829 (41%) | carbohydrate binding (90%);hyaluronate lyase activity (59%);lyase activity (41%) | GO:0005975 (90%) | carbohydrate metabolic process (90%) | IPR008929 (100%);IPR012970 (100%);IPR038970 (100%) | Chondroitin AC/alginate lyase (100%);Polysaccharide lyase 8 | N-terminal alpha-helical (100%);Polysaccharide lyase 8 (100%) |  |  | 1 | 1 |
| AASVLVELAK | Bacteria | Bacteria |  |  |  |  |  |  |  |  |  |  |  |  |  |  |  |  |  |  |  |  |  |  |  |  |  |  | Bacteria;;;;;;;;;;;Bacteria |  |  |  | GO:0005737 (0%);GO:0005829 (0%) | cytoplasm (0%);cytosol (0%) | GO:0003677 (100%);GO:0003700 (95%);GO:0000986 (0%) | DNA binding (100%);DNA-binding transcription factor activity (95%);bacterial-type cis-regulatory region sequence-specific DNA binding (0%) | GO:0070689 (1%);GO:0006355 (0%);GO:0045893 (0%) | L-threonine catabolic process to propionate (1%);regulation of transcription | DNA-templated (0%);positive regulation of transcription | DNA-templated (0%) | IPR005119 (100%);IPR000847 (98%);IPR036388 (98%) | LysR | substrate-binding (100%);Transcription regulator HTH | 1 | 100.0846 |
| AGALVAVR | Bacteria | Bacteria |  |  |  |  |  |  |  |  |  |  |  |  |  |  |  |  |  |  |  |  |  |  |  |  |  |  | Bacteria;;;;;;;;;;;Bacteria | [K01875](https://www.genome.jp/entry/K01875) | 6.1.1.3 | Threonine--tRNA ligase (100%) | GO:0005737 (56%);GO:0005694 (13%);GO:0005886 (6%) | cytoplasm (56%);chromosome (13%);plasma membrane (6%) | GO:0005524 (63%);GO:0046872 (44%);GO:0000049 (38%) | ATP binding (63%);metal ion binding (44%);tRNA binding (38%) | GO:0006435 (38%);GO:0006260 (19%);GO:0032259 (19%) | threonyl-tRNA aminoacylation (38%);DNA replication (19%);methylation (19%) | IPR002314 (32%);IPR002320 (32%);IPR004095 (32%) | Aminoacyl-tRNA synthetase | class II (G/ P/ S/T) (32%);Threonine-tRNA ligase | class IIa (32%);TGS (32%) |  | 1 | 100.3077 |
| DLGALTVGVVTRPFTFEGR | Bacteria | Bacteria |  |  |  |  |  |  |  |  |  |  |  |  |  |  |  |  |  |  |  |  |  |  |  |  |  |  | Bacteria;;;;;;;;;;;Bacteria |  |  |  | GO:0005737 (98%);GO:0032153 (98%);GO:0030428 (0%) | cytoplasm (98%);cell division site (98%);cell septum (0%) | GO:0003924 (99%);GO:0005525 (98%);GO:0042802 (0%) | GTPase activity (99%);GTP binding (98%);identical protein binding (0%) | GO:0000917 (98%);GO:0043093 (98%);GO:0051258 (98%) | division septum assembly (98%);FtsZ-dependent cytokinesis (98%);protein polymerization (98%) | IPR003008 (100%);IPR036525 (100%);IPR000158 (100%) | Tubulin/FtsZ | GTPase domain (100%);Tubulin/FtsZ | GTPase domain superfamily (100%);Cell division protein FtsZ (100%) |  | 1 | 1 |
| EAYETLSDDQK | Bacillales | Bacteria |  |  |  | Firmicutes |  |  | Bacilli |  |  |  | Bacillales |  |  |  |  |  |  |  |  |  |  |  |  |  |  |  | Bacteria;Firmicutes;;;Bacilli;Bacillales;;;;;;Bacillales |  |  |  | GO:0005737 (47%) | cytoplasm (47%) | GO:0051082 (100%);GO:0031072 (97%);GO:0046872 (53%) | unfolded protein binding (100%);heat shock protein binding (97%);metal ion binding (53%) | GO:0006457 (96%);GO:0006260 (47%);GO:0009408 (44%) | protein folding (96%);DNA replication (47%);response to heat (44%) | IPR001305 (100%);IPR001623 (100%);IPR002939 (100%) | Heat shock protein DnaJ | cysteine-rich domain (100%);DnaJ domain (100%);Chaperone DnaJ | C-terminal (100%) |  | 100.7333 | 100.8462 |
| FLQVLEGLK | Candidatus Phytoplasma | Bacteria |  |  |  | Tenericutes |  |  | Mollicutes |  |  |  | Acholeplasmatales |  |  |  |  | Acholeplasmataceae |  |  |  | Candidatus Phytoplasma |  |  |  |  |  |  | Bacteria;Tenericutes;;;Mollicutes;Acholeplasmatales;Acholeplasmataceae;;Candidatus Phytoplasma;;;Candidatus Phytoplasma | [K00380](https://www.genome.jp/entry/K00380) | 6.1.1.21 | Histidine--tRNA ligase (100%) | GO:0005737 (100%) | cytoplasm (100%) | GO:0004821 (100%);GO:0005524 (100%) | histidine-tRNA ligase activity (100%);ATP binding (100%) | GO:0006427 (100%) | histidyl-tRNA aminoacylation (100%) | IPR004154 (100%);IPR004516 (100%);IPR006195 (100%) | Anticodon-binding (100%);Histidine-tRNA ligase/ATP phosphoribosyltransferase regulatory subunit (100%);Aminoacyl-tRNA synthetase | class II (100%) |  |  | 100.3333 | 100.1385 |
| HYDGAEEAMR | Enterobacterales | Bacteria |  |  |  | Proteobacteria |  |  | Gammaproteobacteria |  |  |  | Enterobacterales |  |  |  |  |  |  |  |  |  |  |  |  |  |  |  | Bacteria;Proteobacteria;;;Gammaproteobacteria;Enterobacterales;;;;;;Enterobacterales | K15904 | 4.2.1.75 | Uroporphyrinogen-III synthase (100%) |  |  | GO:0004852 (100%);GO:0008168 (4%);GO:0004418 (0%) | uroporphyrinogen-III synthase activity (100%);methyltransferase activity (4%);hydroxymethylbilane synthase activity (0%) | GO:0006780 (69%);GO:0006782 (69%);GO:0032259 (4%) | uroporphyrinogen III biosynthetic process (69%);protoporphyrinogen IX biosynthetic process (69%);methylation (4%) | IPR003754 (100%);IPR036108 (100%);IPR039793 (100%) | Tetrapyrrole biosynthesis | uroporphyrinogen III synthase (100%);Tetrapyrrole biosynthesis | uroporphyrinogen III synthase superfamily (100%);Uroporphyrinogen-III synthase (100%) |  | 100.7333 | 100.6923 |
| LAAELVGK | Bacteria | Bacteria |  |  |  |  |  |  |  |  |  |  |  |  |  |  |  |  |  |  |  |  |  |  |  |  |  |  | Bacteria;;;;;;;;;;;Bacteria | [K01950](https://www.genome.jp/entry/K01950) | 3.6.1.66 (93%);3.2.1.21 (7%) | XTP/dITP diphosphatase (93%);Beta-glucosidase (7%) | GO:0005737 (6%);GO:0005829 (6%) | cytoplasm (6%);cytosol (6%) | GO:0000166 (72%);GO:0017111 (72%);GO:0046872 (72%) | nucleotide binding (72%);nucleoside-triphosphatase activity (72%);metal ion binding (72%) | GO:0009117 (72%);GO:0009146 (72%);GO:0005975 (11%) | nucleotide metabolic process (72%);purine nucleoside triphosphate catabolic process (72%);carbohydrate metabolic process (11%) | IPR007712 (46%);IPR035093 (46%);IPR002637 (35%) | Toxin-antitoxin system | RelE/ParE toxin family (46%);Toxin-antitoxin system | RelE/ParE toxin domain superfamily (46%);Ham1-like protein (35%) |  | 100.8 | 100.5385 |
| LFPLTADR | Bacteria | Bacteria |  |  |  |  |  |  |  |  |  |  |  |  |  |  |  |  |  |  |  |  |  |  |  |  |  |  | Bacteria;;;;;;;;;;;Bacteria | [K07568](https://www.genome.jp/entry/K07568) | 2.7.7.27 | Glucose-1-phosphate adenylyltransferase (100%) | GO:0016021 (1%) | integral component of membrane (1%) | GO:0008878 (99%);GO:0005524 (97%);GO:0030246 (1%) | glucose-1-phosphate adenylyltransferase activity (99%);ATP binding (97%);carbohydrate binding (1%) | GO:0005978 (99%) | glycogen biosynthetic process (99%) | IPR005835 (99%);IPR005836 (99%);IPR029044 (99%) | Nucleotidyl transferase domain (99%);ADP-glucose pyrophosphorylase | conserved site (99%);Nucleotide-diphospho-sugar transferases (99%) |  |  | 100.5333 | 100.0692 |
| LGFPADAR | Bacteria | Bacteria |  |  |  |  |  |  |  |  |  |  |  |  |  |  |  |  |  |  |  |  |  |  |  |  |  |  | Bacteria;;;;;;;;;;;Bacteria | [K01081](https://www.genome.jp/entry/K01081) | 3.5.4.25 (97%);4.1.99.12 (83%) | GTP cyclohydrolase II (97%);3 | 4-dihydroxy-2-butanone-4-phosphate synthase (83%) | GO:0009349 (3%) | riboflavin synthase complex (3%) | GO:0003935 (97%);GO:0005525 (97%);GO:0008270 (90%) | GTP cyclohydrolase II activity (97%);GTP binding (97%);zinc ion binding (90%) | GO:0009231 (97%) | riboflavin biosynthetic process (97%) | IPR000926 (97%);IPR032677 (97%);IPR036144 (97%) | GTP cyclohydrolase II | RibA (97%);GTP cyclohydrolase II (97%);RibA-like superfamily (97%) |  | 1 | 100.5385 |
| LGVTAMVR | Bacteria | Bacteria |  |  |  |  |  |  |  |  |  |  |  |  |  |  |  |  |  |  |  |  |  |  |  |  |  |  | Bacteria;;;;;;;;;;;Bacteria | K01595 | 2.7.7.3 | Pantetheine-phosphate adenylyltransferase (100%) | GO:0005737 (50%) | cytoplasm (50%) | GO:0004595 (50%);GO:0005344 (50%);GO:0005524 (50%) | pantetheine-phosphate adenylyltransferase activity (50%);oxygen carrier activity (50%);ATP binding (50%) | GO:0015937 (50%) | coenzyme A biosynthetic process (50%) | IPR000971 (50%);IPR001980 (50%);IPR004821 (50%) | Globin (50%);Phosphopantetheine adenylyltransferase (50%);Cytidyltransferase-like domain (50%) |  |  |  | 100.6667 | 100.1 |
| LNAAGTNAR | Neisseriaceae | Bacteria |  |  |  | Proteobacteria |  |  | Betaproteobacteria |  |  |  | Neisseriales |  |  |  |  | Neisseriaceae |  |  |  |  |  |  |  |  |  |  | Bacteria;Proteobacteria;;;Betaproteobacteria;Neisseriales;Neisseriaceae;;;;;Neisseriaceae | K00943 | 6.3.2.8 | UDP-N-acetylmuramate--L-alanine ligase (100%) | GO:0005737 (99%) | cytoplasm (99%) | GO:0005524 (100%);GO:0008763 (100%) | ATP binding (100%);UDP-N-acetylmuramate-L-alanine ligase activity (100%) | GO:0007049 (99%);GO:0008360 (99%);GO:0009252 (99%) | cell cycle (99%);regulation of cell shape (99%);peptidoglycan biosynthetic process (99%) | IPR000713 (100%);IPR005758 (100%);IPR013221 (100%) | Mur ligase | N-terminal catalytic domain (100%);UDP-N-acetylmuramate--L-alanine ligase (100%);Mur ligase | central (100%) |  | 100.66 | 100.1462 |
| LTVAELAELVK | Bacteria | Bacteria |  |  |  |  |  |  |  |  |  |  |  |  |  |  |  |  |  |  |  |  |  |  |  |  |  |  | Bacteria;;;;;;;;;;;Bacteria |  |  |  | GO:0005840 (100%) | ribosome (100%) | GO:0003735 (100%) | structural constituent of ribosome (100%) | GO:0006412 (100%) | translation (100%) | IPR000206 (100%);IPR008932 (100%);IPR013823 (100%) | Ribosomal protein L7/L12 (100%);Ribosomal protein L7/L12 | oligomerisation (100%);Ribosomal protein L7/L12 | C-terminal (100%) |  | 100.6 | 1 |
| NNPALHLAR | Rhizobium | Bacteria |  |  |  | Proteobacteria |  |  | Alphaproteobacteria |  |  |  | Rhizobiales |  |  |  |  | Rhizobiaceae |  |  |  | Rhizobium |  |  |  |  |  |  | Bacteria;Proteobacteria;;;Alphaproteobacteria;Rhizobiales;Rhizobiaceae;;Rhizobium;;;Rhizobium | K00919 | 2.7.1.148 | 4-(cytidine 5'-diphospho)-2-C-methyl-D-erythritol kinase (100%) |  |  | GO:0005524 (100%);GO:0050515 (100%) | ATP binding (100%);4-(cytidine 5'-diphospho)-2-C-methyl-D-erythritol kinase activity (100%) | GO:0016114 (100%);GO:0019288 (100%) | terpenoid biosynthetic process (100%);isopentenyl diphosphate biosynthetic process | methylerythritol 4-phosphate pathway (100%) | IPR004424 (100%);IPR006204 (100%);IPR013750 (100%) | 4-diphosphocytidyl-2C-methyl-D-erythritol kinase (100%);GHMP kinase N-terminal domain (100%);GHMP kinase | C-terminal domain (100%) |  | 100.8 | 100.1538 |
| QEVELVLNWTPK | Bacteria | Bacteria |  |  |  |  |  |  |  |  |  |  |  |  |  |  |  |  |  |  |  |  |  |  |  |  |  |  | Bacteria;;;;;;;;;;;Bacteria |  |  |  |  |  |  |  |  |  | IPR036465 (100%);IPR002035 (93%) | von Willebrand factor A-like domain superfamily (100%);von Willebrand factor | type A (93%) |  |  | 1 | 1 |
| QQESSGPPR | Streptomyces | Bacteria |  |  |  | Actinobacteria |  |  | Actinobacteria |  |  |  | Streptomycetales |  |  |  |  | Streptomycetaceae |  |  |  | Streptomyces |  |  |  |  |  |  | Bacteria;Actinobacteria;;;Actinobacteria;Streptomycetales;Streptomycetaceae;;Streptomyces;;;Streptomyces |  |  |  | GO:0005887 (100%);GO:0032153 (100%) | integral component of plasma membrane (100%);cell division site (100%) |  |  | GO:0043093 (100%);GO:0090529 (96%) | FtsZ-dependent cytokinesis (100%);cell septum assembly (96%) | IPR026579 (100%);IPR034746 (100%);IPR013685 (96%) | Cell division protein FtsQ (100%);POTRA domain (100%);POTRA domain | FtsQ-type (96%) |  |  | 100.3933 | 100.4615 |
| QTVEGVLK | Bacteria | Bacteria |  |  |  |  |  |  |  |  |  |  |  |  |  |  |  |  |  |  |  |  |  |  |  |  |  |  | Bacteria;;;;;;;;;;;Bacteria | [K01889](https://www.genome.jp/dbget-bin/www_bget?ko:K01889) | 6.3.5.- (99%);6.1.1.20 (1%) | Carbon--nitrogen ligases with glutamine as amido-N-donor (99%);Phenylalanine--tRNA ligase (1%) | GO:0005737 (1%) | cytoplasm (1%) | GO:0005524 (96%);GO:0016740 (95%);GO:0050567 (95%) | ATP binding (96%);transferase activity (95%);glutaminyl-tRNA synthase (glutamine-hydrolyzing) activity (95%) | GO:0006412 (95%);GO:0006432 (1%) | translation (95%);phenylalanyl-tRNA aminoacylation (1%) | IPR003789 (99%);IPR006075 (99%);IPR014746 (99%) | Aspartyl/glutamyl-tRNA amidotransferase subunit B-like (99%);Aspartyl/Glutamyl-tRNA(Gln) amidotransferase | subunit B/E | catalytic (99%);Glutamine synthetase/guanido kinase | catalytic domain (99%) | 1 | 100 |
| SLAAQGAELAFTYQGEALGK | Rhizobiales | Bacteria |  |  |  | Proteobacteria |  |  | Alphaproteobacteria |  |  |  | Rhizobiales |  |  |  |  |  |  |  |  |  |  |  |  |  |  |  | Bacteria;Proteobacteria;;;Alphaproteobacteria;Rhizobiales;;;;;;Rhizobiales | [K00208](https://www.genome.jp/entry/K00208+1.3.1.10+R10122) | 1.3.1.9 | Enoyl-[acyl-carrier-protein] reductase (NADH) (100%) | GO:0005886 (9%) | plasma membrane (9%) | GO:0004318 (100%);GO:0016631 (100%) | enoyl-[acyl-carrier-protein] reductase (NADH) activity (100%);enoyl-[acyl-carrier-protein] reductase activity (100%) | GO:0006633 (100%) | fatty acid biosynthetic process (100%) | IPR002347 (100%);IPR014358 (100%);IPR036291 (100%) | Short-chain dehydrogenase/reductase SDR (100%);Enoyl-[acyl-carrier-protein] reductase (NADH) (100%);NAD(P)-binding domain superfamily (100%) |  |  |  | 100.4667 | 99.99231 |
| TMGAFAGK | Streptococcus | Bacteria |  |  |  | Firmicutes |  |  | Bacilli |  |  |  | Lactobacillales |  |  |  |  | Streptococcaceae |  |  |  | Streptococcus |  |  |  |  |  |  | Bacteria;Firmicutes;;;Bacilli;Lactobacillales;Streptococcaceae;;Streptococcus;;;Streptococcus |  |  |  | GO:0005737 (100%);GO:0043590 (100%) | cytoplasm (100%);bacterial nucleoid (100%) | GO:0003677 (100%) | DNA binding (100%) |  |  | IPR004401 (100%);IPR036894 (100%) | Nucleoid-associated protein YbaB/EbfC family (100%);Nucleoid-associated protein YbaB-like domain superfamily (100%) |  |  |  | 100.8 | 100.2308 |
| VAEVTGQAAGK | Streptomyces | Bacteria |  |  |  | Actinobacteria |  |  | Actinobacteria |  |  |  | Streptomycetales |  |  |  |  | Streptomycetaceae |  |  |  | Streptomyces |  |  |  |  |  |  | Bacteria;Actinobacteria;;;Actinobacteria;Streptomycetales;Streptomycetaceae;;Streptomyces;;;Streptomyces | [K01207](https://www.genome.jp/entry/K01207) | 3.6.4.8 | Transferred entry: 5.6.1.5 (100%) | GO:0000502 (100%) | proteasome complex (100%) | GO:0005524 (99%);GO:0016887 (99%);GO:0016853 (2%) | ATP binding (99%);ATPase activity (99%);isomerase activity (2%) | GO:0010498 (93%);GO:0019941 (93%) | proteasomal protein catabolic process (93%);modification-dependent protein catabolic process (93%) | IPR003593 (100%);IPR003959 (100%);IPR003960 (100%) | AAA+ ATPase domain (100%);ATPase | AAA-type | core (100%);ATPase | AAA-type | 100.6667 | 100.0769 |
| VALNLHQQEK | Bacteria | Bacteria |  |  |  |  |  |  |  |  |  |  |  |  |  |  |  |  |  |  |  |  |  |  |  |  |  |  | Bacteria;;;;;;;;;;;Bacteria | K05341 | 2.7.7.7 | DNA-directed DNA polymerase (100%) | GO:0005737 (96%) | cytoplasm (96%) | GO:0003684 (99%);GO:0003887 (99%);GO:0000287 (96%) | damaged DNA binding (99%);DNA-directed DNA polymerase activity (99%);magnesium ion binding (96%) | GO:0006281 (99%);GO:0006261 (96%);GO:0009432 (1%) | DNA repair (99%);DNA-dependent DNA replication (96%);SOS response (1%) | IPR017961 (100%);IPR036775 (100%);IPR001126 (96%) | DNA polymerase | Y-family | little finger domain (100%);DNA polymerase | Y-family | 100.3333 | 100.0615 |
| VEELVLDLK | Mycoplasma | Bacteria |  |  |  | Tenericutes |  |  | Mollicutes |  |  |  | Mycoplasmatales |  |  |  |  | Mycoplasmataceae |  |  |  | Mycoplasma |  |  |  |  |  |  | Bacteria;Tenericutes;;;Mollicutes;Mycoplasmatales;Mycoplasmataceae;;Mycoplasma;;;Mycoplasma | [K02036](https://www.genome.jp/entry/K02036) | 7.3.2.1 | ABC-type phosphate transporter (100%) | GO:0005886 (100%) | plasma membrane (100%) | GO:0005315 (100%);GO:0005524 (100%);GO:0015415 (100%) | inorganic phosphate transmembrane transporter activity (100%);ATP binding (100%);ATPase-coupled phosphate ion transmembrane transporter activity (100%) |  |  | IPR003439 (100%);IPR003593 (100%);IPR005670 (100%) | ABC transporter-like (100%);AAA+ ATPase domain (100%);Phosphate transport system permease protein 1 (100%) |  |  |  | 100.9333 | 100 |
| YGELAAELK | Mycobacterium avium complex (MAC) | Bacteria |  |  |  | Actinobacteria |  |  | Actinobacteria |  |  |  | Corynebacteriales |  |  |  |  | Mycobacteriaceae |  |  |  | Mycobacterium |  | Mycobacterium avium complex (MAC) |  |  |  |  | Bacteria;Actinobacteria;;;Actinobacteria;Corynebacteriales;Mycobacteriaceae;;Mycobacterium;Mycobacterium avium complex (MAC);;Mycobacterium avium complex (MAC) | K00075 | 1.3.1.98 | UDP-N-acetylmuramate dehydrogenase (100%) | GO:0005737 (91%) | cytoplasm (91%) | GO:0008762 (100%);GO:0071949 (100%) | UDP-N-acetylmuramate dehydrogenase activity (100%);FAD binding (100%) | GO:0007049 (91%);GO:0008360 (91%);GO:0009252 (91%) | cell cycle (91%);regulation of cell shape (91%);peptidoglycan biosynthetic process (91%) | IPR003170 (100%);IPR006094 (100%);IPR016166 (100%) | UDP-N-acetylenolpyruvoylglucosamine reductase (100%);FAD linked oxidase | N-terminal (100%);FAD-binding domain | PCMH-type (100%) |  | 100.3933 | 100.1615 |

**Supplementary table 4:**

| **Differentially expressed metabolites** | | | | |
| --- | --- | --- | --- | --- |
| label | NR average | R average | FC | p-value |
| C00003 | 8208.401 | 824.0212 | 9.961396 | 0.004944 |
| C00006 | 3777.029 | 14104.77 | 0.267784 | 0.007392 |
| C00009 | 19340.84 | 8155.378 | 2.371544 | 0.028018 |
| C00042 | 201116.5 | 84227.92 | 2.387765 | 0.023127 |
| C00053 | 2561.935 | 13114.59 | 0.19535 | 0.039116 |
| C00075 | 6475.874 | 18615.08 | 0.347883 | 0.041269 |
| C00078 | 68276.58 | 49847.95 | 1.369697 | 0.001507 |
| C00085 | 4149.334 | 372.7759 | 11.13091 | 0.048749 |
| C00092 | 83.81879 | 24.67107 | 3.397452 | 0.001668 |
| C00097 | 39612.29 | 24845.35 | 1.594355 | 0.029501 |
| C00106 | 3580973 | 2481307 | 1.44318 | 0.006181 |
| C00149 | 16226.82 | 2551.694 | 6.359236 | 0.03942 |
| C00167 | 19773.42 | 68168.44 | 0.290067 | 0.048538 |
| C00169 | 504650.6 | 227611 | 2.217163 | 0.03321 |
| C00170 | 1984.551 | 556.0206 | 3.569205 | 0.005476 |
| C00178 | 128.0018 | 311.2189 | 0.411292 | 0.005252 |
| C00191 | 983597.1 | 441247 | 2.22913 | 0.01402 |
| C00198 | 137558.5 | 55442.44 | 2.481104 | 0.032141 |
| C00214 | 8890.407 | 2298.03 | 3.868709 | 0.018218 |
| C00248 | 17680.92 | 15628.99 | 1.13129 | 0.036133 |
| C00249 | 10478857 | 4948761 | 2.117471 | 0.048385 |
| C00257 | 414670.5 | 115998.5 | 3.574794 | 0.02094 |
| C00299 | 44742.07 | 63006.79 | 0.710115 | 0.032784 |
| C00334 | 2840.215 | 444.8348 | 6.384875 | 0.036295 |
| C00352 | 838.3066 | 19.67521 | 42.60724 | 0.021626 |
| C00398 | 119486.3 | 92760.91 | 1.288111 | 0.036391 |
| C00408 | 33845.91 | 10813.08 | 3.13009 | 0.019061 |
| C00439 | 9743.925 | 2252.225 | 4.326354 | 0.005924 |
| C00463 | 1053363 | 816020 | 1.290854 | 0.02242 |
| C00474 | 13362.43 | 2089.13 | 6.396166 | 0.045834 |
| C00487 | 1279.706 | 401.2472 | 3.18932 | 0.015767 |
| C00499 | 1067.834 | 129.0999 | 8.271379 | 0.036581 |
| C00523 | 2030.986 | 518.5748 | 3.916476 | 0.045008 |
| C00597 | 4816.397 | 654.1482 | 7.362852 | 0.044586 |
| C00606 | 1400.945 | 230.9448 | 6.066149 | 0.020259 |
| C00620 | 4749.408 | 291.8238 | 16.27491 | 0.041331 |
| C00624 | 13878.06 | 6866.082 | 2.021249 | 0.029578 |
| C00651 | 16029.33 | 2428.409 | 6.600754 | 0.048714 |
| C00659 | 4333.377 | 776.3194 | 5.581951 | 0.045564 |
| C00747 | 61955.37 | 16815.24 | 3.684478 | 0.033637 |
| C00785 | 628985.4 | 1389941 | 0.452527 | 0.035362 |
| C00846 | 713.6922 | 16.06213 | 44.43324 | 0.019129 |
| C00864 | 499558.9 | 903501 | 0.552915 | 0.006702 |
| C00882 | 2160.026 | 180.2899 | 11.98085 | 0.033174 |
| C00902 | 2337.057 | 641.2561 | 3.644498 | 0.025043 |
| C00906 | 11145.13 | 1906.884 | 5.844679 | 0.029308 |
| C00931 | 4284.501 | 10.22378 | 419.0721 | 0.031189 |
| C00988 | 3341.56 | 376.3371 | 8.879168 | 0.034529 |
| C00989 | 2047312 | 822717.1 | 2.488476 | 0.047208 |
| C00993 | 13238.03 | 48.81103 | 271.2099 | 0.032062 |
| C01010 | 184590.7 | 49786.49 | 3.707648 | 0.025854 |
| C01026 | 33371.2 | 9197.229 | 3.628397 | 0.039117 |
| C01041 | 897.4756 | 156.3816 | 5.73901 | 0.032632 |
| C01054 | 4189.965 | 306.1165 | 13.68748 | 0.027391 |
| C01059 | 524.5892 | 83.52569 | 6.280573 | 0.028136 |
| C01108 | 24869.07 | 5291.472 | 4.699839 | 0.048637 |
| C01157 | 2793.873 | 183.7545 | 15.20438 | 0.033421 |
| C01192 | 403.2189 | 161.6254 | 2.494775 | 0.019956 |
| C01210 | 36835433 | 29789190 | 1.236537 | 0.042701 |
| C01234 | 13099.14 | 932.7299 | 14.04387 | 0.031532 |
| C01236 | 328.6564 | 1008.852 | 0.325773 | 0.03528 |
| C01239 | 2549.898 | 71.62234 | 35.60199 | 0.036958 |
| C01243 | 4225.602 | 109.1314 | 38.7203 | 0.036383 |
| C01252 | 11454.7 | 4250.674 | 2.694797 | 0.007427 |
| C01419 | 173.6557 | 56.58554 | 3.068905 | 0.0227 |
| C01475 | 325964.3 | 248558.3 | 1.31142 | 0.021796 |
| C01551 | 83042.37 | 22526.06 | 3.686502 | 0.033338 |
| C01595 | 6431340 | 3424571 | 1.877998 | 0.017273 |
| C01838 | 12810.91 | 5175.348 | 2.475371 | 0.003834 |
| C02170 | 16802.31 | 3965.927 | 4.236667 | 0.020815 |
| C02197 | 829.0708 | 336.2199 | 2.465859 | 0.011233 |
| C02217 | 41482.45 | 23368.16 | 1.77517 | 0.019021 |
| C02287 | 43678.33 | 29192.24 | 1.496231 | 0.042572 |
| C02291 | 6455.226 | 895.7161 | 7.206777 | 0.041729 |
| C02355 | 14749.25 | 8539.243 | 1.727231 | 0.004683 |
| C02376 | 4369874 | 5712105 | 0.76502 | 8.35E-05 |
| C02470 | 9382.444 | 0 | #DIV/0! | 0.031826 |
| C02479 | 80827.52 | 52963.29 | 1.526105 | 0.016672 |
| C02571 | 4480.537 | 1353.374 | 3.310642 | 0.023896 |
| C02631 | 841394.2 | 713513.1 | 1.179227 | 0.03892 |
| C02656 | 5400.929 | 116.8116 | 46.23626 | 0.032706 |
| C02678 | 11752.12 | 18992.42 | 0.618779 | 0.045173 |
| C02679 | 221997 | 73309.35 | 3.028222 | 0.038255 |
| C02700 | 3373.34 | 1016.605 | 3.318241 | 0.038077 |
| C02718 | 573409.5 | 486333.4 | 1.179046 | 0.045089 |
| C02771 | 11912.78 | 2994.821 | 3.977795 | 0.033909 |
| C02835 | 365843 | 460433.8 | 0.794561 | 0.008316 |
| C02946 | 20533.32 | 9168.833 | 2.239469 | 0.013099 |
| C03090 | 912.2316 | 375.7084 | 2.428031 | 0.032144 |
| C03242 | 121209.8 | 63569.49 | 1.90673 | 0.030536 |
| C03406 | 3147.184 | 39.79263 | 79.08963 | 0.033987 |
| C03410 | 16453.05 | 4173.838 | 3.941948 | 0.015812 |
| C03413 | 5435.253 | 524.923 | 10.35438 | 0.038693 |
| C03440 | 1779442 | 1420602 | 1.252597 | 0.043571 |
| C03519 | 17335.25 | 3871.838 | 4.477267 | 0.032369 |
| C03564 | 10774.1 | 175.8685 | 61.26224 | 0.032901 |
| C03665 | 24933.83 | 2116.925 | 11.77832 | 0.029134 |
| C03758 | 55825.24 | 39769.31 | 1.403727 | 0.023081 |
| C03862 | 13773.37 | 684.1055 | 20.13339 | 0.039684 |
| C04025 | 47443.06 | 20752.58 | 2.286128 | 0.034529 |
| C04137 | 55045.84 | 26297.8 | 2.093173 | 0.005906 |
| C04411 | 5600.404 | 371.2046 | 15.08711 | 0.000223 |
| C04462 | 8580.274 | 345.3703 | 24.8437 | 0.029213 |
| C04479 | 1445.586 | 152.7923 | 9.461115 | 0.021453 |
| C04555 | 1132.443 | 215.7785 | 5.248175 | 0.022637 |
| C04556 | 213914.3 | 116768.5 | 1.831952 | 0.04228 |
| C04640 | 26506.1 | 10332.87 | 2.56522 | 0.022762 |
| C04685 | 9620.299 | 3383.23 | 2.843525 | 0.01199 |
| C04717 | 752281.5 | 289575.6 | 2.597876 | 0.022419 |
| C04767 | 331118 | 374002.4 | 0.885336 | 0.001352 |
| C05122 | 1521350 | 857312.6 | 1.774557 | 0.040635 |
| C05132 | 58306.68 | 38214.09 | 1.52579 | 0.027023 |
| C05135 | 205.7707 | 30.31707 | 6.787289 | 1E-05 |
| C05145 | 51275.91 | 2372.344 | 21.61403 | 0.032353 |
| C05239 | 464.2901 | 97.55529 | 4.759251 | 0.044508 |
| C05400 | 4113.707 | 43.21641 | 95.18855 | 0.03361 |
| C05462 | 6456817 | 3711889 | 1.739496 | 0.001411 |
| C05463 | 3545.056 | 433.9208 | 8.169823 | 0.028107 |
| C05465 | 3480.745 | 82.75733 | 42.05966 | 0.030516 |
| C05466 | 1379.422 | 550.7688 | 2.50454 | 0.025676 |
| C05512 | 112965.6 | 13407.61 | 8.425488 | 0.0022 |
| C05565 | 183776.3 | 74685.43 | 2.460671 | 0.007271 |
| C05579 | 6734.335 | 933.5077 | 7.214011 | 0.034075 |
| C05636 | 122116.8 | 19409.69 | 6.291541 | 0.01804 |
| C05645 | 198.9546 | 71.83016 | 2.769791 | 0.03428 |
| C05656 | 4577.477 | 1178.506 | 3.884135 | 0.032432 |
| C05659 | 1743.846 | 586.9804 | 2.970876 | 0.047587 |
| C05787 | 114946.5 | 68144.07 | 1.686816 | 0.018758 |
| C05791 | 459196.5 | 126371.9 | 3.633693 | 0.014356 |
| C05795 | 34508.28 | 50788.54 | 0.67945 | 0.023078 |
| C05832 | 24522.1 | 18260.88 | 1.342876 | 0.029908 |
| C05849 | 2948.175 | 273.2912 | 10.78767 | 0.023691 |
| C05901 | 184.9367 | 52.93288 | 3.493797 | 0.012148 |
| C05933 | 972.8422 | 177.1753 | 5.490845 | 0.035842 |
| C05944 | 470985.4 | 585336 | 0.804641 | 0.0247 |
| C06451 | 49112.77 | 2118.822 | 23.17928 | 0.032303 |
| C06645 | 27545.17 | 46197.05 | 0.596254 | 0.020441 |
| C06670 | 34263.37 | 21770.29 | 1.573859 | 0.013858 |
| C06677 | 86.87612 | 14.86414 | 5.844679 | 1.69E-07 |
| C08302 | 390619.1 | 303145.2 | 1.288554 | 0.041556 |
| C08317 | 126374.8 | 205175.3 | 0.615936 | 0.032071 |
| C08320 | 721.3429 | 27.2306 | 26.49016 | 0.0082 |
| C08348 | 52946.54 | 20988.32 | 2.522667 | 0.044678 |
| C08375 | 16932 | 37473.08 | 0.451844 | 0.028664 |
| C08390 | 1582.158 | 161.0646 | 9.823126 | 0.04765 |
| C08486 | 56455.28 | 29335 | 1.924503 | 0.036516 |
| C10264 | 38059.83 | 22963.92 | 1.657375 | 0.00042 |
| C10998 | 6083.823 | 4792.311 | 1.269497 | 0.010599 |
| C11150 | 4438.553 | 53.53804 | 82.90466 | 0.032764 |
| C11457 | 6399.224 | 1267.874 | 5.04721 | 0.04054 |
| C11811 | 217072.9 | 310935.4 | 0.698129 | 0.042933 |
| C11849 | 4929.796 | 1249.198 | 3.946367 | 0.025316 |
| C11896 | 98646.97 | 62428.41 | 1.580161 | 0.040944 |
| C11924 | 226296.3 | 125527.3 | 1.802765 | 0.04382 |
| C11992 | 108469.3 | 130613.5 | 0.83046 | 0.000973 |
| C12002 | 54688.33 | 73101.96 | 0.74811 | 2.46E-05 |
| C12106 | 4405.529 | 186.9038 | 23.5711 | 0.035206 |
| C12294 | 43461.12 | 38053.76 | 1.142098 | 0.013504 |
| C12452 | 40916.3 | 31049.61 | 1.317772 | 0.01899 |
| C12625 | 1312.636 | 302.0904 | 4.345177 | 0.006732 |
| C12889 | 4135.224 | 62.53635 | 66.12513 | 0.034424 |
| C13358 | 32894.55 | 22287.73 | 1.475904 | 0.016964 |
| C13790 | 6725.931 | 1370.945 | 4.906055 | 0.040842 |
| C13792 | 29151.63 | 49996.77 | 0.58307 | 0.015683 |
| C13846 | 96078.1 | 38514.31 | 2.494608 | 0.03528 |
| C13864 | 32876.56 | 81.94713 | 401.1923 | 0.031773 |
| C13914 | 195836.8 | 76137.94 | 2.572131 | 0.048713 |
| C14218 | 1156.649 | 166.5279 | 6.945676 | 0.01488 |
| C14772 | 3058.233 | 1217.202 | 2.51251 | 0.040302 |
| C14820 | 92169.62 | 38341.8 | 2.403894 | 0.018574 |
| C14872 | 6671.843 | 403.8929 | 16.51884 | 0.027293 |
| C14877 | 40365.41 | 25017 | 1.613519 | 0.014265 |
| C15532 | 47121.4 | 16258.02 | 2.898348 | 0.042726 |
| C15559 | 458403.8 | 251328 | 1.823926 | 0.041714 |
| C15974 | 2183.601 | 12916.67 | 0.169053 | 0.037065 |
| C16038 | 3145.196 | 639.5058 | 4.918166 | 0.007558 |
| C16138 | 3156.229 | 28.7723 | 109.6968 | 0.031807 |
| C16308 | 5152237 | 2060489 | 2.500493 | 0.000505 |
| C16352 | 4457.245 | 11.07015 | 402.6365 | 0.031026 |
| C16537 | 1466.856 | 18.96925 | 77.3281 | 0.031062 |
| C16551 | 8506.245 | 1420.039 | 5.990149 | 0.0325 |
| C16682 | 6434.724 | 24677.27 | 0.260755 | 0.043361 |
| C16939 | 16051.05 | 108.1129 | 148.4656 | 0.032159 |
| C17076 | 4095443 | 3400565 | 1.204342 | 0.038622 |
| C18043 | 619.9721 | 97.08279 | 6.386014 | 0.00224 |
| C19615 | 30242.62 | 16457.48 | 1.837622 | 0.026809 |
| D06890 | 18794.68 | 680.6118 | 27.61439 | 0.029559 |
| HMDB0000622 | 3822.764 | 151.806 | 25.18191 | 0.035161 |
| HMDB0000695 | 19375.53 | 5329.269 | 3.635682 | 0.013421 |
| HMDB0000701 | 43179.88 | 97049.46 | 0.444926 | 0.018328 |
| HMDB0001863 | 62271.59 | 22318.64 | 2.790115 | 0.048088 |
| HMDB00044 | 53611.56 | 17697.7 | 3.029295 | 0.007143 |
| HMDB00079 | 7236.925 | 250.3797 | 28.9038 | 0.035428 |

**Supplementary table 5**

| **Temporal Change in Expression of metabolites** | | | | | | | | | | | | |
| --- | --- | --- | --- | --- | --- | --- | --- | --- | --- | --- | --- | --- |
| **day 0** | | |  |  | **day 3** | | |  |  | **day 7** | | |
| label | FC(NR/R) | T test |  |  | label | FC(NR/R) | P VALUE |  |  | label | FC(NR/R) | PVALUE |
| C00003 | 9.961396 | 0.004944 |  |  | C00003 | 51.40571 | 0.001770681 |  |  | C00006 | 0.50153 | 0.026208737 |
| C00006 | 0.267784 | 0.007392 |  |  | C00006 | 0.506092 | 0.021131481 |  |  | C00009 | 2.718549 | 0.04437021 |
| C00009 | 2.371544 | 0.028018 |  |  | C00020 | 0.104955 | 0.019653572 |  |  | C00020 | 0.190065 | 4.51891E-10 |
| C00042 | 2.387765 | 0.023127 |  |  | C00022 | 0.157215 | 0.011854973 |  |  | C00022 | 0.156883 | 0.002479178 |
| C00053 | 0.19535 | 0.039116 |  |  | C00025 | 0.21483 | 0.003730538 |  |  | C00025 | 0.083987 | 4.23615E-11 |
| C00075 | 0.347883 | 0.041269 |  |  | C00037 | 0.620013 | 4.19918E-07 |  |  | C00037 | 0.502125 | 1.12828E-08 |
| C00078 | 1.369697 | 0.001507 |  |  | C00042 | 0.238359 | 1.14541E-06 |  |  | C00041 | 0.574063 | 5.96935E-07 |
| C00085 | 11.13091 | 0.048749 |  |  | C00047 | 0.221267 | 0.000781869 |  |  | C00042 | 0.221808 | 7.51822E-06 |
| C00092 | 3.397452 | 0.001668 |  |  | C00049 | 0.233302 | 0.003781516 |  |  | C00047 | 0.220848 | 3.07313E-12 |
| C00097 | 1.594355 | 0.029501 |  |  | C00053 | 0.187215 | 0.010546098 |  |  | C00049 | 0.212092 | 5.14778E-10 |
| C00106 | 1.44318 | 0.006181 |  |  | C00062 | 0.165513 | 0.001462942 |  |  | C00053 | 0.229113 | 0.000221931 |
| C00149 | 6.359236 | 0.03942 |  |  | C00063 | 0.127908 | 0.00141521 |  |  | C00062 | 0.121659 | 1.46996E-06 |
| C00167 | 0.290067 | 0.048538 |  |  | C00065 | 0.177983 | 0.001306316 |  |  | C00063 | 0.151846 | 1.719E-07 |
| C00169 | 2.217163 | 0.03321 |  |  | C00072 | 1.547766 | 3.64474E-07 |  |  | C00065 | 0.176581 | 7.91455E-22 |
| C00170 | 3.569205 | 0.005476 |  |  | C00073 | 0.268628 | 0.000771455 |  |  | C00072 | 1.59208 | 7.79645E-07 |
| C00178 | 0.411292 | 0.005252 |  |  | C00075 | 0.326606 | 0.005103032 |  |  | C00073 | 0.498265 | 0.040714495 |
| C00191 | 2.22913 | 0.01402 |  |  | C00079 | 0.195333 | 2.62407E-10 |  |  | C00075 | 0.280265 | 0.000158701 |
| C00198 | 2.481104 | 0.032141 |  |  | C00082 | 0.136341 | 3.01903E-10 |  |  | C00079 | 0.195972 | 6.96941E-17 |
| C00214 | 3.868709 | 0.018218 |  |  | C00092 | 0.032025 | 0.001973245 |  |  | C00082 | 0.129109 | 1.89577E-13 |
| C00248 | 1.13129 | 0.036133 |  |  | C00104 | 0.189579 | 0.006046079 |  |  | C00085 | 0.132328 | 0.007609296 |
| C00249 | 2.117471 | 0.048385 |  |  | C00106 | 0.597639 | 0.000106007 |  |  | C00089 | 0.284183 | 0.011879538 |
| C00257 | 3.574794 | 0.02094 |  |  | C00109 | 0.191965 | 4.03494E-06 |  |  | C00092 | 0.064802 | 0.012888854 |
| C00299 | 0.710115 | 0.032784 |  |  | C00110 | 0.140903 | 0.017078111 |  |  | C00103 | 0.057228 | 0.000745653 |
| C00334 | 6.384875 | 0.036295 |  |  | C00111 | 0.259381 | 0.000643365 |  |  | C00104 | 0.186904 | 1.2099E-07 |
| C00352 | 42.60724 | 0.021626 |  |  | C00118 | 0.202489 | 0.019241989 |  |  | C00106 | 0.526453 | 1.55909E-05 |
| C00398 | 1.288111 | 0.036391 |  |  | C00121 | 0.189465 | 0.033992966 |  |  | C00109 | 0.194381 | 1.52274E-09 |
| C00408 | 3.13009 | 0.019061 |  |  | C00122 | 0.292937 | 7.79483E-07 |  |  | C00111 | 0.282373 | 1.21324E-13 |
| C00439 | 4.326354 | 0.005924 |  |  | C00123 | 0.161833 | 6.98369E-13 |  |  | C00118 | 0.175594 | 0.024346228 |
| C00463 | 1.290854 | 0.02242 |  |  | C00129 | 1.612338 | 0.001680503 |  |  | C00121 | 0.237381 | 4.42659E-05 |
| C00474 | 6.396166 | 0.045834 |  |  | C00131 | 0.102525 | 2.72536E-05 |  |  | C00122 | 0.234683 | 1.78682E-12 |
| C00487 | 3.18932 | 0.015767 |  |  | C00135 | 0.232596 | 7.24437E-16 |  |  | C00123 | 0.134012 | 1.55624E-14 |
| C00499 | 8.271379 | 0.036581 |  |  | C00140 | 0.244473 | 2.39772E-08 |  |  | C00131 | 0.103567 | 1.49021E-06 |
| C00523 | 3.916476 | 0.045008 |  |  | C00141 | 0.198863 | 3.46933E-13 |  |  | C00135 | 0.218502 | 3.59191E-20 |
| C00597 | 7.362852 | 0.044586 |  |  | C00148 | 0.134916 | 0.034380038 |  |  | C00140 | 0.158345 | 3.47207E-09 |
| C00606 | 6.066149 | 0.020259 |  |  | C00152 | 1.598947 | 1.07603E-06 |  |  | C00141 | 0.251221 | 6.51703E-14 |
| C00620 | 16.27491 | 0.041331 |  |  | C00157 | 2.225131 | 4.62923E-08 |  |  | C00148 | 0.127979 | 1.82892E-16 |
| C00624 | 2.021249 | 0.029578 |  |  | C00158 | 0.256086 | 0.000189312 |  |  | C00149 | 0.290438 | 3.01189E-06 |
| C00651 | 6.600754 | 0.048714 |  |  | C00160 | 0.154887 | 1.28471E-08 |  |  | C00157 | 2.117607 | 1.39116E-06 |
| C00659 | 5.581951 | 0.045564 |  |  | C00166 | 0.104193 | 4.18978E-07 |  |  | C00158 | 0.257664 | 2.47685E-07 |
| C00747 | 3.684478 | 0.033637 |  |  | C00169 | 0.207846 | 0.016767476 |  |  | C00160 | 0.214851 | 1.07402E-09 |
| C00785 | 0.452527 | 0.035362 |  |  | C00178 | 0.11113 | 0.000308843 |  |  | C00166 | 0.125315 | 0.000174154 |
| C00846 | 44.43324 | 0.019129 |  |  | C00183 | 0.191802 | 1.6709E-12 |  |  | C00167 | 0.046423 | 0.006679726 |
| C00864 | 0.552915 | 0.006702 |  |  | C00187 | 0.240353 | 0.003740088 |  |  | C00169 | 0.294486 | 3.31313E-15 |
| C00882 | 11.98085 | 0.033174 |  |  | C00191 | 0.444698 | 0.001193392 |  |  | C00178 | 0.11617 | 8.68085E-08 |
| C00902 | 3.644498 | 0.025043 |  |  | C00198 | 0.646933 | 0.024166223 |  |  | C00183 | 0.208491 | 7.22278E-15 |
| C00906 | 5.844679 | 0.029308 |  |  | C00209 | 0.138507 | 0.008599273 |  |  | C00187 | 0.176652 | 0.00101327 |
| C00931 | 419.0721 | 0.031189 |  |  | C00212 | 2.343615 | 0.00132836 |  |  | C00198 | 0.626827 | 0.017895026 |
| C00988 | 8.879168 | 0.034529 |  |  | C00216 | 0.366324 | 0.000309191 |  |  | C00209 | 0.158336 | 5.6556E-10 |
| C00989 | 2.488476 | 0.047208 |  |  | C00219 | 0.252102 | 5.48749E-05 |  |  | C00213 | 0.149588 | 7.00217E-18 |
| C00993 | 271.2099 | 0.032062 |  |  | C00245 | 0.373654 | 0.001155787 |  |  | C00216 | 0.268137 | 0.000109381 |
| C01010 | 3.707648 | 0.025854 |  |  | C00249 | 0.19803 | 2.94052E-06 |  |  | C00219 | 0.201239 | 3.60587E-10 |
| C01026 | 3.628397 | 0.039117 |  |  | C00250 | 7.253846 | 0.000180059 |  |  | C00245 | 0.219116 | 0.001635922 |
| C01041 | 5.73901 | 0.032632 |  |  | C00256 | 0.161668 | 1.85749E-10 |  |  | C00249 | 0.22818 | 3.31848E-14 |
| C01054 | 13.68748 | 0.027391 |  |  | C00257 | 0.434466 | 0.004506408 |  |  | C00256 | 0.177629 | 6.27417E-09 |
| C01059 | 6.280573 | 0.028136 |  |  | C00262 | 0.143907 | 0.002264973 |  |  | C00257 | 0.553464 | 0.008220415 |
| C01108 | 4.699839 | 0.048637 |  |  | C00263 | 0.158426 | 2.90825E-09 |  |  | C00258 | 0.199607 | 2.03655E-10 |
| C01157 | 15.20438 | 0.033421 |  |  | C00281 | 42.04595 | 1.80963E-12 |  |  | C00262 | 0.125531 | 0.00344952 |
| C01192 | 2.494775 | 0.019956 |  |  | C00294 | 0.042948 | 0.033894353 |  |  | C00263 | 0.189121 | 1.23113E-13 |
| C01210 | 1.236537 | 0.042701 |  |  | C00295 | 0.067168 | 0.00971338 |  |  | C00281 | 142.4615 | 5.36681E-07 |
| C01234 | 14.04387 | 0.031532 |  |  | C00296 | 0.391625 | 0.001745557 |  |  | C00295 | 0.305786 | 4.20716E-05 |
| C01236 | 0.325773 | 0.03528 |  |  | C00299 | 2.049338 | 8.61832E-09 |  |  | C00299 | 2.621935 | 1.21871E-13 |
| C01239 | 35.60199 | 0.036958 |  |  | C00300 | 4.264895 | 0.000773249 |  |  | C00300 | 7.596065 | 0.000150956 |
| C01243 | 38.7203 | 0.036383 |  |  | C00311 | 0.163181 | 0.000404384 |  |  | C00311 | 0.171061 | 7.32681E-09 |
| C01252 | 2.694797 | 0.007427 |  |  | C00318 | 0.22542 | 0.00135454 |  |  | C00318 | 0.185982 | 7.07807E-06 |
| C01419 | 3.068905 | 0.0227 |  |  | C00319 | 1.595455 | 8.20096E-05 |  |  | C00327 | 0.120148 | 8.42014E-10 |
| C01475 | 1.31142 | 0.021796 |  |  | C00327 | 0.140033 | 0.000102738 |  |  | C00334 | 0.189604 | 0.000155521 |
| C01551 | 3.686502 | 0.033338 |  |  | C00328 | 0.155557 | 4.77707E-05 |  |  | C00345 | 0.259817 | 6.26704E-10 |
| C01595 | 1.877998 | 0.017273 |  |  | C00345 | 0.246259 | 0.001671679 |  |  | C00346 | 1.881096 | 0.017793693 |
| C01838 | 2.475371 | 0.003834 |  |  | C00346 | 3.417696 | 1.13093E-07 |  |  | C00362 | 1.718495 | 0.004563523 |
| C02170 | 4.236667 | 0.020815 |  |  | C00362 | 1.74534 | 0.00011587 |  |  | C00366 | 0.261139 | 9.01001E-13 |
| C02197 | 2.465859 | 0.011233 |  |  | C00366 | 0.300212 | 7.82356E-11 |  |  | C00378 | 6.227835 | 2.83253E-05 |
| C02217 | 1.77517 | 0.019021 |  |  | C00378 | 2.479086 | 1.69621E-05 |  |  | C00385 | 0.094151 | 5.735E-05 |
| C02287 | 1.496231 | 0.042572 |  |  | C00380 | 0.556073 | 0.00966187 |  |  | C00386 | 0.143931 | 0.009524451 |
| C02291 | 7.206777 | 0.041729 |  |  | C00385 | 0.157068 | 1.93616E-05 |  |  | C00392 | 19.31634 | 1.50535E-08 |
| C02355 | 1.727231 | 0.004683 |  |  | C00386 | 0.258889 | 0.024370817 |  |  | C00396 | 0.155684 | 3.18801E-05 |
| C02376 | 0.76502 | 8.35E-05 |  |  | C00392 | 12.49012 | 2.70214E-05 |  |  | C00398 | 0.462622 | 3.56836E-06 |
| C02470 | 9382 | 0.031826 |  |  | C00396 | 0.190531 | 0.001937972 |  |  | C00408 | 0.233484 | 8.27338E-06 |
| C02479 | 1.526105 | 0.016672 |  |  | C00398 | 0.544303 | 2.72227E-05 |  |  | C00418 | 0.232841 | 2.16294E-06 |
| C02571 | 3.310642 | 0.023896 |  |  | C00408 | 0.150993 | 5.53879E-05 |  |  | C00429 | 0.24473 | 8.04595E-12 |
| C02631 | 1.179227 | 0.03892 |  |  | C00418 | 0.150539 | 0.000576223 |  |  | C00433 | 0.26981 | 8.66884E-07 |
| C02656 | 46.23626 | 0.032706 |  |  | C00429 | 0.25887 | 6.74526E-07 |  |  | C00437 | 0.265642 | 1.45806E-05 |
| C02678 | 0.618779 | 0.045173 |  |  | C00433 | 0.272563 | 9.89851E-05 |  |  | C00449 | 0.517006 | 6.74506E-08 |
| C02679 | 3.028222 | 0.038255 |  |  | C00437 | 0.356372 | 0.000892507 |  |  | C00450 | 0.215808 | 0.000202158 |
| C02700 | 3.318241 | 0.038077 |  |  | C00445 | 1.622485 | 1.70471E-07 |  |  | C00474 | 0.37051 | 0.000853885 |
| C02718 | 1.179046 | 0.045089 |  |  | C00449 | 0.651808 | 0.000274095 |  |  | C00487 | 0.134826 | 2.32992E-08 |
| C02771 | 3.977795 | 0.033909 |  |  | C00450 | 0.20174 | 0.046002285 |  |  | C00491 | 0.112013 | 9.49054E-07 |
| C02835 | 0.794561 | 0.008316 |  |  | C00474 | 0.299872 | 0.006678739 |  |  | C00493 | 3.357896 | 3.77916E-10 |
| C02946 | 2.239469 | 0.013099 |  |  | C00487 | 0.055504 | 6.18515E-08 |  |  | C00499 | 0.244532 | 0.002622761 |
| C03090 | 2.428031 | 0.032144 |  |  | C00491 | 0.074211 | 0.007495799 |  |  | C00502 | 0.361319 | 4.01812E-06 |
| C03242 | 1.90673 | 0.030536 |  |  | C00493 | 2.599963 | 1.3507E-16 |  |  | C00503 | 0.276848 | 5.47755E-06 |
| C03406 | 79.08963 | 0.033987 |  |  | C00502 | 0.340434 | 0.001953349 |  |  | C00504 | 0.063681 | 0.003839538 |
| C03410 | 3.941948 | 0.015812 |  |  | C00503 | 0.219268 | 0.000410471 |  |  | C00515 | 6596.498 | 3.60126E-06 |
| C03413 | 10.35438 | 0.038693 |  |  | C00506 | 0.492902 | 0.042894456 |  |  | C00523 | 0.221755 | 0.001424104 |
| C03440 | 1.252597 | 0.043571 |  |  | C00515 | 442.9772 | 0.000802785 |  |  | C00525 | 0.13639 | 5.56015E-15 |
| C03519 | 4.477267 | 0.032369 |  |  | C00525 | 0.133545 | 1.04064E-12 |  |  | C00532 | 2.054606 | 3.35397E-07 |
| C03564 | 61.26224 | 0.032901 |  |  | C00555 | 0.45739 | 1.82379E-08 |  |  | C00555 | 0.433567 | 1.86648E-09 |
| C03665 | 11.77832 | 0.029134 |  |  | C00575 | 0.169201 | 0.023489979 |  |  | C00575 | 0.33643 | 4.70545E-08 |
| C03758 | 1.403727 | 0.023081 |  |  | C00581 | 0.150166 | 0.005261448 |  |  | C00581 | 0.11349 | 0.000739577 |
| C03862 | 20.13339 | 0.039684 |  |  | C00587 | 2.202542 | 5.14294E-08 |  |  | C00587 | 2.149076 | 4.34012E-10 |
| C04025 | 2.286128 | 0.034529 |  |  | C00596 | 0.190346 | 0.002867642 |  |  | C00596 | 0.233237 | 2.42698E-13 |
| C04137 | 2.093173 | 0.005906 |  |  | C00597 | 0.068406 | 0.004416312 |  |  | C00597 | 0.174974 | 0.002085284 |
| C04411 | 15.08711 | 0.000223 |  |  | C00599 | 0.395522 | 9.43897E-05 |  |  | C00599 | 0.228538 | 0.000122538 |
| C04462 | 24.8437 | 0.029213 |  |  | C00606 | 0.057932 | 0.047292753 |  |  | C00606 | 0.09944 | 6.4949E-12 |
| C04479 | 9.461115 | 0.021453 |  |  | C00620 | 0.19286 | 0.039842555 |  |  | C00620 | 0.268234 | 0.023513703 |
| C04555 | 5.248175 | 0.022637 |  |  | C00631 | 0.23971 | 0.005486005 |  |  | C00631 | 0.251069 | 4.58873E-11 |
| C04556 | 1.831952 | 0.04228 |  |  | C00633 | 0.600688 | 2.0422E-06 |  |  | C00632 | 1.569743 | 3.58549E-06 |
| C04640 | 2.56522 | 0.022762 |  |  | C00651 | 0.097316 | 0.031708413 |  |  | C00633 | 0.585968 | 3.92697E-07 |
| C04685 | 2.843525 | 0.01199 |  |  | C00655 | 2.425966 | 6.75524E-11 |  |  | C00651 | 0.150911 | 0.000114479 |
| C04717 | 2.597876 | 0.022419 |  |  | C00659 | 0.155355 | 4.74572E-06 |  |  | C00655 | 3.327176 | 5.50239E-14 |
| C04767 | 0.885336 | 0.001352 |  |  | C00666 | 1.803901 | 1.62613E-06 |  |  | C00659 | 0.326962 | 9.36952E-05 |
| C05122 | 1.774557 | 0.040635 |  |  | C00669 | 0.08337 | 0.025473192 |  |  | C00666 | 2.114981 | 1.09859E-06 |
| C05132 | 1.52579 | 0.027023 |  |  | C00672 | 0.152122 | 0.001490901 |  |  | C00669 | 0.123486 | 0.000613905 |
| C05135 | 6.787289 | 1E-05 |  |  | C00682 | 0.513901 | 0.013072413 |  |  | C00672 | 0.179793 | 6.98392E-14 |
| C05145 | 21.61403 | 0.032353 |  |  | C00700 | 0.151544 | 0.015046229 |  |  | C00682 | 0.263996 | 0.000285957 |
| C05239 | 4.759251 | 0.044508 |  |  | C00718 | 0.194844 | 0.006149225 |  |  | C00695 | 0.128321 | 0.003212081 |
| C05400 | 95.18855 | 0.03361 |  |  | C00719 | 0.253825 | 7.39181E-05 |  |  | C00700 | 0.31385 | 0.008093317 |
| C05462 | 1.739496 | 0.001411 |  |  | C00735 | 4.123417 | 1.64936E-13 |  |  | C00718 | 0.252216 | 6.56287E-22 |
| C05463 | 8.169823 | 0.028107 |  |  | C00777 | 1.77201 | 6.50336E-09 |  |  | C00719 | 0.506304 | 0.023937386 |
| C05465 | 42.05966 | 0.030516 |  |  | C00785 | 1.904663 | 8.55749E-07 |  |  | C00735 | 2.130247 | 4.30318E-05 |
| C05466 | 2.50454 | 0.025676 |  |  | C00791 | 0.276501 | 9.13821E-12 |  |  | C00747 | 0.370497 | 0.0010913 |
| C05512 | 8.425488 | 0.0022 |  |  | C00794 | 0.097451 | 0.000260818 |  |  | C00777 | 1.500635 | 0.000208078 |
| C05565 | 2.460671 | 0.007271 |  |  | C00798 | 0.08403 | 0.026493155 |  |  | C00788 | 0.557044 | 0.028027445 |
| C05579 | 7.214011 | 0.034075 |  |  | C00803 | 0.134916 | 1.93393E-07 |  |  | C00791 | 0.294276 | 7.41458E-13 |
| C05636 | 6.291541 | 0.01804 |  |  | C00818 | 0.237389 | 0.004430171 |  |  | C00794 | 0.097174 | 0.004193387 |
| C05645 | 2.769791 | 0.03428 |  |  | C00819 | 0.203134 | 0.027987549 |  |  | C00798 | 0.131788 | 4.0147E-07 |
| C05656 | 3.884135 | 0.032432 |  |  | C00828 | 1.881494 | 0.00022541 |  |  | C00803 | 0.118632 | 6.46084E-09 |
| C05659 | 2.970876 | 0.047587 |  |  | C00846 | 0.054309 | 0.000392204 |  |  | C00818 | 0.263675 | 0.001608418 |
| C05787 | 1.686816 | 0.018758 |  |  | C00857 | 106.304 | 1.00516E-06 |  |  | C00819 | 0.190955 | 6.8661E-13 |
| C05791 | 3.633693 | 0.014356 |  |  | C00864 | 2.077157 | 5.53919E-06 |  |  | C00836 | 0.141099 | 0.000278989 |
| C05795 | 0.67945 | 0.023078 |  |  | C00869 | 0.172803 | 0.000350428 |  |  | C00846 | 0.04517 | 0.002770144 |
| C05832 | 1.342876 | 0.029908 |  |  | C00879 | 0.584986 | 0.012997213 |  |  | C00847 | 0.096061 | 0.000714471 |
| C05849 | 10.78767 | 0.023691 |  |  | C00902 | 0.413201 | 0.003080925 |  |  | C00864 | 1.997426 | 0.000112157 |
| C05901 | 3.493797 | 0.012148 |  |  | C00906 | 0.127933 | 4.84963E-06 |  |  | C00869 | 0.234058 | 1.11689E-13 |
| C05933 | 5.490845 | 0.035842 |  |  | C00921 | 14.87305 | 3.96402E-20 |  |  | C00870 | 0.138081 | 0.009680529 |
| C05944 | 0.804641 | 0.0247 |  |  | C00936 | 0.185689 | 0.000858876 |  |  | C00882 | 0.132625 | 1.22888E-05 |
| C06451 | 23.17928 | 0.032303 |  |  | C00940 | 0.334101 | 4.53219E-06 |  |  | C00902 | 0.385457 | 0.01090324 |
| C06645 | 0.596254 | 0.020441 |  |  | C00944 | 0.271222 | 4.60533E-07 |  |  | C00906 | 0.107352 | 4.51832E-07 |
| C06670 | 1.573859 | 0.013858 |  |  | C00954 | 0.20058 | 6.11793E-06 |  |  | C00921 | 19.12368 | 3.96971E-16 |
| C06677 | 86.87612 | 1.69E-07 |  |  | C00955 | 0.096345 | 6.6407E-06 |  |  | C00931 | 0.073073 | 0.018172725 |
| C08302 | 1.288554 | 0.041556 |  |  | C00956 | 0.347436 | 1.93062E-05 |  |  | C00936 | 0.200744 | 8.57039E-16 |
| C08317 | 0.615936 | 0.032071 |  |  | C00969 | 0.150198 | 0.001776364 |  |  | C00940 | 0.244242 | 8.12471E-08 |
| C08320 | 26.49016 | 0.0082 |  |  | C00979 | 0.380919 | 3.05212E-07 |  |  | C00944 | 0.334031 | 1.96896E-08 |
| C08348 | 2.522667 | 0.044678 |  |  | C00984 | 0.200103 | 0.001702654 |  |  | C00954 | 0.257389 | 2.52217E-06 |
| C08375 | 0.451844 | 0.028664 |  |  | C00989 | 0.22471 | 1.13158E-08 |  |  | C00955 | 0.096504 | 5.4187E-06 |
| C08390 | 9.823126 | 0.04765 |  |  | C00990 | 0.30712 | 1.50292E-07 |  |  | C00956 | 0.330854 | 3.14919E-06 |
| C08486 | 1.924503 | 0.036516 |  |  | C01010 | 0.288888 | 2.86393E-07 |  |  | C00979 | 0.35177 | 1.31603E-08 |
| C10264 | 1.657375 | 0.00042 |  |  | C01017 | 1.778746 | 3.33236E-10 |  |  | C00984 | 0.225282 | 4.62551E-16 |
| C10998 | 1.269497 | 0.010599 |  |  | C01026 | 0.132764 | 0.004869675 |  |  | C00988 | 0.29691 | 1.77915E-05 |
| C11150 | 82.90466 | 0.032764 |  |  | C01029 | 1.920513 | 9.45296E-12 |  |  | C00989 | 0.2618 | 1.76854E-09 |
| C11457 | 5.04721 | 0.04054 |  |  | C01041 | 0.175182 | 0.000048291 |  |  | C00990 | 0.285936 | 1.16857E-07 |
| C11811 | 0.698129 | 0.042933 |  |  | C01042 | 0.13849 | 0.005957729 |  |  | C00993 | 0.075545 | 9.02657E-05 |
| C11849 | 3.946367 | 0.025316 |  |  | C01044 | 0.181195 | 0.003228923 |  |  | C01010 | 0.366929 | 5.0836E-06 |
| C11896 | 1.580161 | 0.040944 |  |  | C01054 | 0.134701 | 8.57108E-05 |  |  | C01017 | 1.864513 | 1.10507E-10 |
| C11924 | 1.802765 | 0.04382 |  |  | C01061 | 4.671541 | 5.18682E-21 |  |  | C01026 | 0.212417 | 2.46571E-06 |
| C11992 | 0.83046 | 0.000973 |  |  | C01074 | 1.902121 | 0.000253485 |  |  | C01029 | 1.65082 | 2.09746E-05 |
| C12002 | 0.74811 | 2.46E-05 |  |  | C01103 | 20.15653 | 2.35887E-08 |  |  | C01041 | 0.286174 | 0.0029161 |
| C12106 | 23.5711 | 0.035206 |  |  | C01127 | 0.22554 | 0.002262528 |  |  | C01042 | 0.166821 | 2.80599E-12 |
| C12294 | 1.142098 | 0.013504 |  |  | C01137 | 0.048758 | 0.040535314 |  |  | C01044 | 0.147846 | 0.000262234 |
| C12452 | 1.317772 | 0.01899 |  |  | C01157 | 0.315432 | 0.023327871 |  |  | C01045 | 1.757889 | 1.55874E-10 |
| C12625 | 4.345177 | 0.006732 |  |  | C01176 | 0.239989 | 0.049381527 |  |  | C01054 | 0.107444 | 0.000155324 |
| C12889 | 66.12513 | 0.034424 |  |  | C01187 | 5.841214 | 0.000359012 |  |  | C01056 | 4.64989 | 6.81681E-05 |
| C13358 | 1.475904 | 0.016964 |  |  | C01190 | 2.439986 | 2.45737E-07 |  |  | C01059 | 0.210965 | 0.000185367 |
| C13790 | 4.906055 | 0.040842 |  |  | C01192 | 0.020328 | 1.7287E-06 |  |  | C01061 | 4.143874 | 8.18349E-22 |
| C13792 | 0.58307 | 0.015683 |  |  | C01225 | 3.172724 | 3.03045E-12 |  |  | C01074 | 3.079832 | 2.08791E-11 |
| C13846 | 2.494608 | 0.03528 |  |  | C01226 | 0.210837 | 6.89847E-06 |  |  | C01102 | 0.27492 | 0.002521541 |
| C13864 | 401.1923 | 0.031773 |  |  | C01227 | 0.240358 | 0.039298457 |  |  | C01103 | 34.76459 | 1.09386E-18 |
| C13914 | 2.572131 | 0.048713 |  |  | C01230 | 2.277843 | 2.4998E-14 |  |  | C01108 | 0.176352 | 4.22306E-09 |
| C14218 | 6.945676 | 0.01488 |  |  | C01233 | 0.193748 | 0.008193436 |  |  | C01118 | 9.817419 | 3.72712E-05 |
| C14772 | 2.51251 | 0.040302 |  |  | C01236 | 0.019122 | 1.86011E-05 |  |  | C01127 | 0.143368 | 0.000363753 |
| C14820 | 2.403894 | 0.018574 |  |  | C01252 | 0.295448 | 0.000559826 |  |  | C01137 | 0.043386 | 0.00026564 |
| C14872 | 16.51884 | 0.027293 |  |  | C01269 | 4.159748 | 3.34576E-10 |  |  | C01176 | 0.341974 | 0.036873142 |
| C14877 | 1.613519 | 0.014265 |  |  | C01273 | 0.290548 | 2.25838E-05 |  |  | C01187 | 19.71723 | 5.56867E-05 |
| C15532 | 2.898348 | 0.042726 |  |  | C01290 | 2.637001 | 4.25603E-07 |  |  | C01190 | 3.148429 | 1.26017E-05 |
| C15559 | 1.823926 | 0.041714 |  |  | C01344 | 2.801342 | 8.85412E-10 |  |  | C01192 | 0.025602 | 1.18931E-06 |
| C15974 | 0.169053 | 0.037065 |  |  | C01345 | 0.151674 | 1.49599E-06 |  |  | C01210 | 1.599241 | 4.91772E-05 |
| C16038 | 4.918166 | 0.007558 |  |  | C01384 | 0.303437 | 3.0582E-06 |  |  | C01225 | 2.604692 | 0.000258409 |
| C16138 | 109.6968 | 0.031807 |  |  | C01475 | 0.50388 | 8.1507E-07 |  |  | C01226 | 0.247432 | 2.84979E-12 |
| C16308 | 2.500493 | 0.000505 |  |  | C01479 | 0.206358 | 0.000413851 |  |  | C01227 | 0.280092 | 0.00806008 |
| C16352 | 402.6365 | 0.031026 |  |  | C01530 | 0.21134 | 2.12067E-09 |  |  | C01230 | 2.469021 | 1.16131E-13 |
| C16537 | 77.3281 | 0.031062 |  |  | C01545 | 0.571189 | 3.20845E-05 |  |  | C01233 | 0.229302 | 0.003918751 |
| C16551 | 5.990149 | 0.0325 |  |  | C01551 | 0.203868 | 0.031603844 |  |  | C01234 | 0.131576 | 3.80958E-07 |
| C16682 | 0.260755 | 0.043361 |  |  | C01571 | 0.278323 | 0.004715814 |  |  | C01236 | 0.043053 | 1.65368E-06 |
| C16939 | 148.4656 | 0.032159 |  |  | C01585 | 0.171557 | 2.58479E-11 |  |  | C01269 | 6.676409 | 2.86553E-07 |
| C17076 | 1.204342 | 0.038622 |  |  | C01595 | 0.180761 | 0.001395667 |  |  | C01273 | 0.375234 | 0.000638817 |
| C18043 | 6.386014 | 0.00224 |  |  | C01596 | 0.192525 | 7.68443E-08 |  |  | C01344 | 4.913112 | 5.01307E-12 |
| C19615 | 1.837622 | 0.026809 |  |  | C01606 | 0.137991 | 1.69996E-07 |  |  | C01345 | 0.264118 | 3.1281E-05 |
| D06890 | 27.61439 | 0.029559 |  |  | C01674 | 1.914913 | 2.26008E-15 |  |  | C01367 | 0.127639 | 0.000327967 |
| HMDB0000622 | 25.18191 | 0.035161 |  |  | C01801 | 0.181382 | 1.44153E-09 |  |  | C01384 | 0.238978 | 9.73162E-12 |
| HMDB0000695 | 3.635682 | 0.013421 |  |  | C01808 | 0.125119 | 0.005155351 |  |  | C01419 | 0.154671 | 0.007603278 |
| HMDB0000701 | 0.444926 | 0.018328 |  |  | C01921 | 0.211836 | 0.000317097 |  |  | C01475 | 0.582872 | 3.46893E-07 |
| HMDB0001863 | 2.790115 | 0.048088 |  |  | C01924 | 0.079075 | 0.010424897 |  |  | C01479 | 0.154624 | 0.013562439 |
| HMDB00044 | 3.029295 | 0.007143 |  |  | C01933 | 0.377311 | 9.63529E-06 |  |  | C01530 | 0.255518 | 2.2146E-19 |
| HMDB00079 | 28.9038 | 0.035428 |  |  | C01957 | 0.056166 | 0.035632731 |  |  | C01545 | 0.558163 | 9.78902E-07 |
|  |  |  |  |  | C01991 | 0.577683 | 0.001991881 |  |  | C01551 | 0.240894 | 2.22985E-14 |
|  |  |  |  |  | C02037 | 0.206108 | 0.008129272 |  |  | C01571 | 0.129356 | 1.43503E-06 |
|  |  |  |  |  | C02043 | 0.338458 | 2.40133E-05 |  |  | C01585 | 0.127115 | 4.60328E-14 |
|  |  |  |  |  | C02067 | 0.3841 | 3.78837E-06 |  |  | C01595 | 0.454065 | 3.10727E-05 |
|  |  |  |  |  | C02076 | 0.50653 | 7.22894E-10 |  |  | C01596 | 0.264724 | 0.000108168 |
|  |  |  |  |  | C02147 | 0.111088 | 0.006596106 |  |  | C01606 | 0.198659 | 4.91213E-16 |
|  |  |  |  |  | C02170 | 0.347559 | 0.005347011 |  |  | C01613 | 2.355464 | 0.02108637 |
|  |  |  |  |  | C02197 | 0.128205 | 4.64797E-07 |  |  | C01674 | 2.12368 | 2.61832E-19 |
|  |  |  |  |  | C02217 | 0.248044 | 0.001911472 |  |  | C01801 | 0.211013 | 1.33708E-07 |
|  |  |  |  |  | C02237 | 0.207842 | 0.00020932 |  |  | C01808 | 0.134729 | 0.004274462 |
|  |  |  |  |  | C02242 | 1.544216 | 0.007158013 |  |  | C01838 | 0.335708 | 0.00012291 |
|  |  |  |  |  | C02287 | 0.136782 | 0.008427415 |  |  | C01921 | 0.318598 | 0.000437545 |
|  |  |  |  |  | C02291 | 0.247896 | 0.008388297 |  |  | C01924 | 0.014714 | 5.64475E-05 |
|  |  |  |  |  | C02355 | 0.172643 | 0.000914467 |  |  | C01933 | 0.316954 | 0.000316797 |
|  |  |  |  |  | C02356 | 0.32368 | 0.015093125 |  |  | C01957 | 0.010651 | 0.006234561 |
|  |  |  |  |  | C02367 | 0.187291 | 4.26372E-05 |  |  | C01991 | 0.511088 | 0.001560398 |
|  |  |  |  |  | C02427 | 0.28487 | 0.042607908 |  |  | C02037 | 0.22171 | 2.62793E-15 |
|  |  |  |  |  | C02479 | 0.254243 | 3.59309E-11 |  |  | C02043 | 0.382676 | 6.68082E-06 |
|  |  |  |  |  | C02494 | 0.160808 | 0.00021195 |  |  | C02067 | 0.355942 | 2.784E-08 |
|  |  |  |  |  | C02515 | 0 | 0.024822473 |  |  | C02076 | 0.461574 | 6.72E-12 |
|  |  |  |  |  | C02592 | 0.477751 | 0.025372229 |  |  | C02140 | 0.245117 | 0.004568521 |
|  |  |  |  |  | C02656 | 0.241178 | 0.025682124 |  |  | C02147 | 0.109335 | 0.022176723 |
|  |  |  |  |  | C02679 | 0.353549 | 0.003308602 |  |  | C02166 | 0.626216 | 0.018555848 |
|  |  |  |  |  | C02714 | 3.028626 | 5.60961E-20 |  |  | C02170 | 0.225864 | 2.64704E-05 |
|  |  |  |  |  | C02718 | 0.569225 | 1.11258E-09 |  |  | C02197 | 0.145573 | 1.62022E-05 |
|  |  |  |  |  | C02737 | 1.796538 | 0.002363245 |  |  | C02217 | 0.261639 | 1.7062E-05 |
|  |  |  |  |  | C02739 | 7.137497 | 1.98138E-17 |  |  | C02237 | 0.136411 | 6.51761E-13 |
|  |  |  |  |  | C02771 | 0.33887 | 0.006776828 |  |  | C02242 | 2.345902 | 3.87515E-05 |
|  |  |  |  |  | C02774 | 0.249795 | 9.37112E-06 |  |  | C02287 | 0.141113 | 1.81567E-06 |
|  |  |  |  |  | C02794 | 0.608583 | 2.43063E-08 |  |  | C02291 | 0.215643 | 0.000129675 |
|  |  |  |  |  | C02915 | 0.170721 | 0.044419052 |  |  | C02355 | 0.366307 | 1.67839E-05 |
|  |  |  |  |  | C02979 | 0.162443 | 3.01577E-05 |  |  | C02356 | 0.392677 | 1.69365E-05 |
|  |  |  |  |  | C02990 | 1.928766 | 0.001886064 |  |  | C02367 | 0.195712 | 2.23001E-10 |
|  |  |  |  |  | C02997 | 0.239667 | 0.000424426 |  |  | C02376 | 1.512061 | 8.18727E-10 |
|  |  |  |  |  | C03017 | 2.268896 | 0.000170849 |  |  | C02378 | 0.637754 | 0.002077275 |
|  |  |  |  |  | C03042 | 0.273701 | 0.000017363 |  |  | C02479 | 0.275478 | 1.53447E-17 |
|  |  |  |  |  | C03044 | 0.048758 | 0.007786902 |  |  | C02494 | 0.097334 | 3.49062E-05 |
|  |  |  |  |  | C03087 | 1.8813 | 0.023701212 |  |  | C02538 | 0.218814 | 0.00368928 |
|  |  |  |  |  | C03089 | 4.499246 | 8.75194E-15 |  |  | C02571 | 0.35064 | 0.005322833 |
|  |  |  |  |  | C03090 | 0.090351 | 4.76025E-09 |  |  | C02656 | 0.228181 | 0.017513949 |
|  |  |  |  |  | C03139 | 0.423872 | 0.000951246 |  |  | C02679 | 0.296749 | 6.1918E-06 |
|  |  |  |  |  | C03145 | 0.238882 | 1.4399E-07 |  |  | C02700 | 0.012341 | 0.01501006 |
|  |  |  |  |  | C03187 | 3.994376 | 0.000288978 |  |  | C02713 | 0.105243 | 0.003451996 |
|  |  |  |  |  | C03197 | 0.222512 | 1.31672E-07 |  |  | C02714 | 3.040454 | 9.44166E-11 |
|  |  |  |  |  | C03232 | 0.635289 | 0.008375928 |  |  | C02718 | 0.49397 | 3.95636E-11 |
|  |  |  |  |  | C03242 | 0.176154 | 0.000687348 |  |  | C02737 | 1.741584 | 0.003847261 |
|  |  |  |  |  | C03277 | 0.36151 | 0.00502906 |  |  | C02739 | 7.616567 | 4.61618E-17 |
|  |  |  |  |  | C03283 | 0.541527 | 2.76087E-08 |  |  | C02774 | 0.177379 | 2.66676E-05 |
|  |  |  |  |  | C03401 | 2.29394 | 1.38353E-07 |  |  | C02835 | 1.522725 | 7.14302E-06 |
|  |  |  |  |  | C03413 | 0.32484 | 0.008206417 |  |  | C02979 | 0.19646 | 2.99483E-05 |
|  |  |  |  |  | C03428 | 2.455423 | 2.6582E-16 |  |  | C02990 | 1.952446 | 0.009590102 |
|  |  |  |  |  | C03451 | 0.035635 | 0.011600577 |  |  | C02997 | 0.197969 | 1.44052E-06 |
|  |  |  |  |  | C03458 | 0.46405 | 0.002749558 |  |  | C03017 | 2.125684 | 0.010161011 |
|  |  |  |  |  | C03519 | 0.248437 | 0.00012597 |  |  | C03033 | 0.389039 | 0.033610873 |
|  |  |  |  |  | C03557 | 0.336455 | 2.02717E-05 |  |  | C03042 | 0.209986 | 3.38897E-07 |
|  |  |  |  |  | C03577 | 1.666474 | 3.81608E-15 |  |  | C03044 | 0.096564 | 0.023895624 |
|  |  |  |  |  | C03626 | 0.158363 | 0.00074255 |  |  | C03089 | 8.232907 | 4.28023E-11 |
|  |  |  |  |  | C03664 | 2.731173 | 3.11456E-12 |  |  | C03090 | 0.075859 | 4.325E-07 |
|  |  |  |  |  | C03690 | 0.080669 | 0.000101155 |  |  | C03139 | 2.198477 | 0.015093705 |
|  |  |  |  |  | C03740 | 12.45336 | 5.85565E-18 |  |  | C03145 | 0.152073 | 3.45634E-07 |
|  |  |  |  |  | C03758 | 0.540788 | 2.13603E-05 |  |  | C03187 | 2.769083 | 0.021046695 |
|  |  |  |  |  | C03761 | 0.147139 | 0.025601198 |  |  | C03197 | 0.247875 | 8.73377E-09 |
|  |  |  |  |  | C03772 | 0.428595 | 4.04647E-07 |  |  | C03242 | 0.23659 | 2.34475E-10 |
|  |  |  |  |  | C03793 | 1.661173 | 0.039979377 |  |  | C03277 | 0.263318 | 0.000839355 |
|  |  |  |  |  | C03794 | 2.140768 | 4.28821E-06 |  |  | C03283 | 0.480887 | 4.59055E-11 |
|  |  |  |  |  | C03805 | 0.060173 | 1.15464E-07 |  |  | C03366 | 0.037419 | 0.037605748 |
|  |  |  |  |  | C03838 | 4.51496 | 2.95971E-18 |  |  | C03401 | 2.370532 | 1.97396E-05 |
|  |  |  |  |  | C03872 | 0.141126 | 0.042110027 |  |  | C03413 | 0.255814 | 0.002746334 |
|  |  |  |  |  | C03925 | 42.52856 | 2.59277E-12 |  |  | C03428 | 2.419038 | 8.68955E-17 |
|  |  |  |  |  | C03955 | 0.139359 | 0.007901034 |  |  | C03440 | 1.928602 | 5.59491E-06 |
|  |  |  |  |  | C04025 | 0.204661 | 0.000160099 |  |  | C03451 | 0.050561 | 3.24481E-05 |
|  |  |  |  |  | C04148 | 0.276225 | 0.021583874 |  |  | C03458 | 0.301884 | 4.0637E-05 |
|  |  |  |  |  | C04188 | 0.564438 | 0.007932928 |  |  | C03519 | 0.340848 | 0.001439642 |
|  |  |  |  |  | C04218 | 0.171861 | 2.06983E-06 |  |  | C03557 | 0.258155 | 9.66558E-11 |
|  |  |  |  |  | C04271 | 2.582156 | 7.58969E-14 |  |  | C03564 | 0.141965 | 0.000587894 |
|  |  |  |  |  | C04322 | 2.727479 | 1.16397E-11 |  |  | C03577 | 1.752544 | 1.80425E-18 |
|  |  |  |  |  | C04327 | 2.310502 | 5.74245E-06 |  |  | C03594 | 0.513698 | 0.034438993 |
|  |  |  |  |  | C04409 | 0.051218 | 0.029302662 |  |  | C03626 | 0.135961 | 1.54115E-15 |
|  |  |  |  |  | C04411 | 0.345411 | 0.006155094 |  |  | C03646 | 0.348555 | 0.036038879 |
|  |  |  |  |  | C04462 | 0.260561 | 0.028060086 |  |  | C03664 | 3.35035 | 3.47721E-12 |
|  |  |  |  |  | C04479 | 0.102962 | 0.004725811 |  |  | C03665 | 0.364977 | 2.62589E-05 |
|  |  |  |  |  | C04483 | 0.088251 | 0.019483298 |  |  | C03690 | 0.196999 | 1.21032E-10 |
|  |  |  |  |  | C04489 | 0.408223 | 0.006117468 |  |  | C03740 | 14.67 | 2.54914E-15 |
|  |  |  |  |  | C04540 | 2.638502 | 2.4057E-16 |  |  | C03758 | 0.605624 | 3.23915E-06 |
|  |  |  |  |  | C04555 | 0.115641 | 6.90743E-06 |  |  | C03761 | 0.197634 | 1.35433E-13 |
|  |  |  |  |  | C04556 | 0.1805 | 0.000349725 |  |  | C03772 | 0.337063 | 4.20063E-08 |
|  |  |  |  |  | C04592 | 0.615057 | 1.36261E-13 |  |  | C03794 | 1.919823 | 0.002484271 |
|  |  |  |  |  | C04599 | 3.910272 | 0.00213154 |  |  | C03805 | 0.056849 | 1.41894E-15 |
|  |  |  |  |  | C04638 | 2.606484 | 4.3566E-10 |  |  | C03838 | 4.456365 | 2.02483E-11 |
|  |  |  |  |  | C04640 | 0.157895 | 4.40825E-05 |  |  | C03862 | 0.107552 | 0.000361644 |
|  |  |  |  |  | C04666 | 4.08792 | 7.66286E-12 |  |  | C03872 | 0.16818 | 4.75901E-06 |
|  |  |  |  |  | C04685 | 0.140353 | 3.29652E-05 |  |  | C03925 | 30.47027 | 2.86406E-10 |
|  |  |  |  |  | C04692 | 3.434113 | 1.50612E-23 |  |  | C03955 | 0.19489 | 0.000245152 |
|  |  |  |  |  | C04717 | 0.339888 | 0.001150988 |  |  | C04025 | 0.25783 | 3.1941E-17 |
|  |  |  |  |  | C04742 | 0.178926 | 0.024596847 |  |  | C04137 | 0.240591 | 0.032012273 |
|  |  |  |  |  | C04744 | 0.386007 | 0.001477094 |  |  | C04148 | 0.371866 | 0.013991537 |
|  |  |  |  |  | C04751 | 2.431285 | 0.012759408 |  |  | C04188 | 0.559813 | 0.005326416 |
|  |  |  |  |  | C04778 | 0.22496 | 0.039485962 |  |  | C04218 | 0.086889 | 4.58293E-08 |
|  |  |  |  |  | C04834 | 0.27938 | 0.000246685 |  |  | C04271 | 2.572194 | 1.61993E-10 |
|  |  |  |  |  | C04874 | 0.22691 | 0.001197757 |  |  | C04299 | 4.477105 | 0.000134689 |
|  |  |  |  |  | C04886 | 0.268203 | 0.029316243 |  |  | C04322 | 3.188958 | 2.57729E-09 |
|  |  |  |  |  | C04895 | 7.286595 | 4.5704E-21 |  |  | C04327 | 2.481014 | 8.68461E-11 |
|  |  |  |  |  | C05108 | 0.057416 | 0.021801773 |  |  | C04360 | 3.881907 | 0.004346792 |
|  |  |  |  |  | C05132 | 0.38258 | 0.000969738 |  |  | C04411 | 0.314293 | 0.000433148 |
|  |  |  |  |  | C05135 | 0.050955 | 7.46394E-05 |  |  | C04424 | 0.041377 | 0.008216843 |
|  |  |  |  |  | C05139 | 0.149062 | 0.039934317 |  |  | C04431 | 0.009471 | 0.016956142 |
|  |  |  |  |  | C05151 | 5.769205 | 3.12763E-07 |  |  | C04479 | 0.285921 | 0.019218049 |
|  |  |  |  |  | C05177 | 1.861528 | 1.35253E-07 |  |  | C04483 | 0.148884 | 0.002063267 |
|  |  |  |  |  | C05290 | 6.679219 | 0.000798495 |  |  | C04489 | 0.520141 | 0.025915149 |
|  |  |  |  |  | C05381 | 1.872401 | 1.62425E-05 |  |  | C04540 | 3.141257 | 6.50606E-19 |
|  |  |  |  |  | C05403 | 0.159546 | 0.00626097 |  |  | C04555 | 0.116469 | 1.50477E-06 |
|  |  |  |  |  | C05462 | 0.177222 | 7.56788E-08 |  |  | C04556 | 0.224328 | 1.64872E-22 |
|  |  |  |  |  | C05463 | 0.148959 | 0.047973502 |  |  | C04592 | 0.603922 | 5.3112E-11 |
|  |  |  |  |  | C05464 | 0.587434 | 0.001163405 |  |  | C04599 | 3.186274 | 0.040224247 |
|  |  |  |  |  | C05465 | 0.214104 | 0.020711115 |  |  | C04640 | 0.078445 | 2.06579E-06 |
|  |  |  |  |  | C05466 | 0.025388 | 0.000267805 |  |  | C04666 | 5.867084 | 4.82299E-12 |
|  |  |  |  |  | C05471 | 3.720246 | 0.0012029 |  |  | C04685 | 0.133947 | 1.87929E-12 |
|  |  |  |  |  | C05476 | 0.03949 | 9.82416E-07 |  |  | C04692 | 4.063207 | 5.51336E-25 |
|  |  |  |  |  | C05547 | 6.059909 | 1.81469E-12 |  |  | C04717 | 0.213492 | 1.67487E-08 |
|  |  |  |  |  | C05560 | 0.160503 | 0.007783638 |  |  | C04742 | 0.118409 | 1.47164E-05 |
|  |  |  |  |  | C05565 | 7.951209 | 2.04776E-07 |  |  | C04744 | 0.298124 | 4.18692E-05 |
|  |  |  |  |  | C05576 | 0.602359 | 1.07426E-05 |  |  | C04751 | 1.967704 | 0.04079594 |
|  |  |  |  |  | C05579 | 0.043082 | 0.025890436 |  |  | C04778 | 0.335624 | 7.29885E-08 |
|  |  |  |  |  | C05635 | 0.465026 | 0.027131859 |  |  | C04834 | 0.29412 | 9.02423E-06 |
|  |  |  |  |  | C05636 | 0.275338 | 0.006685836 |  |  | C04874 | 0.232807 | 2.86535E-19 |
|  |  |  |  |  | C05642 | 0.287726 | 0.025248007 |  |  | C04886 | 0.318719 | 5.29772E-12 |
|  |  |  |  |  | C05655 | 0.07741 | 6.77546E-05 |  |  | C04895 | 5.953956 | 1.20664E-13 |
|  |  |  |  |  | C05656 | 0.206859 | 0.003210088 |  |  | C05108 | 0.028786 | 0.031100779 |
|  |  |  |  |  | C05658 | 2.576701 | 2.73865E-09 |  |  | C05123 | 0.377623 | 0.03289217 |
|  |  |  |  |  | C05659 | 0.075551 | 0.001372493 |  |  | C05132 | 0.491162 | 0.001270981 |
|  |  |  |  |  | C05660 | 0.443088 | 4.26223E-06 |  |  | C05135 | 0.081558 | 0.014079483 |
|  |  |  |  |  | C05683 | 1.967651 | 6.6772E-09 |  |  | C05139 | 0.215048 | 0.012895666 |
|  |  |  |  |  | C05689 | 188.0696 | 4.49226E-07 |  |  | C05145 | 0.254387 | 1.13677E-07 |
|  |  |  |  |  | C05695 | 0.129388 | 0.02502255 |  |  | C05151 | 16.25342 | 5.18743E-07 |
|  |  |  |  |  | C05699 | 1.832574 | 0.003163411 |  |  | C05177 | 1.958637 | 8.84941E-08 |
|  |  |  |  |  | C05787 | 4.984224 | 5.45049E-13 |  |  | C05239 | 0.223355 | 1.63927E-05 |
|  |  |  |  |  | C05793 | 2.411258 | 1.90063E-11 |  |  | C05290 | 8.839557 | 0.00017555 |
|  |  |  |  |  | C05804 | 4.063235 | 1.07406E-08 |  |  | C05381 | 2.140429 | 2.33699E-06 |
|  |  |  |  |  | C05817 | 1.877765 | 0.040629738 |  |  | C05400 | 0.088686 | 0.018246582 |
|  |  |  |  |  | C05828 | 1.811533 | 0.000049836 |  |  | C05403 | 0.150907 | 0.0025691 |
|  |  |  |  |  | C05832 | 0.517008 | 6.29052E-05 |  |  | C05462 | 0.375872 | 0.000792285 |
|  |  |  |  |  | C05849 | 0.097281 | 0.013871082 |  |  | C05463 | 0.080417 | 8.38782E-08 |
|  |  |  |  |  | C05901 | 0.006688 | 0.036638397 |  |  | C05466 | 0.186439 | 0.01440632 |
|  |  |  |  |  | C05933 | 0.089028 | 6.37429E-06 |  |  | C05471 | 9.185767 | 9.54396E-08 |
|  |  |  |  |  | C05949 | 2.616325 | 0.003246954 |  |  | C05476 | 0.12977 | 4.20915E-07 |
|  |  |  |  |  | C05964 | 2.76562 | 0.022561796 |  |  | C05512 | 7.440127 | 0.000228671 |
|  |  |  |  |  | C06054 | 0.272208 | 3.99384E-07 |  |  | C05547 | 8.511681 | 2.17519E-13 |
|  |  |  |  |  | C06121 | 0.397225 | 0.000006281 |  |  | C05560 | 0.187007 | 0.000336743 |
|  |  |  |  |  | C06313 | 1.757872 | 0.018844109 |  |  | C05565 | 29.85964 | 3.67663E-05 |
|  |  |  |  |  | C06424 | 0.157508 | 9.45964E-05 |  |  | C05579 | 0.086202 | 0.027797382 |
|  |  |  |  |  | C06426 | 0.112692 | 0.0024766 |  |  | C05582 | 0.59498 | 0.001597006 |
|  |  |  |  |  | C06427 | 0.197447 | 0.003394565 |  |  | C05588 | 0.436645 | 0.008236977 |
|  |  |  |  |  | C06428 | 0.188388 | 1.40177E-07 |  |  | C05635 | 0.344014 | 0.009916037 |
|  |  |  |  |  | C06429 | 0.204747 | 0.002619017 |  |  | C05636 | 0.493712 | 0.039357442 |
|  |  |  |  |  | C06451 | 0.084462 | 0.032419172 |  |  | C05642 | 0.392097 | 0.041510027 |
|  |  |  |  |  | C06469 | 0.112721 | 0.006206169 |  |  | C05655 | 0.053629 | 7.36616E-05 |
|  |  |  |  |  | C06470 | 1.683586 | 2.26769E-20 |  |  | C05656 | 0.255709 | 0.000119895 |
|  |  |  |  |  | C06554 | 0.098725 | 0.027529721 |  |  | C05658 | 2.499389 | 3.82337E-14 |
|  |  |  |  |  | C06579 | 0.502178 | 0.047213775 |  |  | C05660 | 0.45823 | 2.09527E-05 |
|  |  |  |  |  | C06656 | 0.034152 | 0.000118174 |  |  | C05689 | 311.0389 | 1.0833E-08 |
|  |  |  |  |  | C06670 | 0.368994 | 6.37649E-05 |  |  | C05695 | 0.095924 | 0.000554182 |
|  |  |  |  |  | C06772 | 0.361391 | 0.008669503 |  |  | C05699 | 1.835889 | 0.021622349 |
|  |  |  |  |  | C07326 | 0.200468 | 2.19259E-07 |  |  | C05787 | 6.589447 | 1.88492E-14 |
|  |  |  |  |  | C07888 | 13.73897 | 3.21436E-12 |  |  | C05789 | 2.239114 | 3.96957E-05 |
|  |  |  |  |  | C08156 | 0.045468 | 0.001588988 |  |  | C05791 | 6.358615 | 0.000568534 |
|  |  |  |  |  | C08302 | 0.546821 | 5.17953E-06 |  |  | C05793 | 2.434451 | 3.52546E-12 |
|  |  |  |  |  | C08314 | 0.161323 | 0.002252796 |  |  | C05795 | 1.590891 | 0.000300224 |
|  |  |  |  |  | C08316 | 0.066736 | 0.005599948 |  |  | C05809 | 49.65724 | 1.96573E-05 |
|  |  |  |  |  | C08322 | 0.15985 | 0.002952386 |  |  | C05828 | 3.373968 | 1.69251E-10 |
|  |  |  |  |  | C08348 | 0.469146 | 0.004836779 |  |  | C05832 | 0.507455 | 0.001279772 |
|  |  |  |  |  | C08357 | 2.816841 | 0.005215709 |  |  | C05849 | 0.102924 | 0.001082924 |
|  |  |  |  |  | C08362 | 0.17353 | 0.002224012 |  |  | C05901 | 0.024877 | 0.001246348 |
|  |  |  |  |  | C08486 | 0.144708 | 0.000349695 |  |  | C05933 | 0.123819 | 8.74831E-05 |
|  |  |  |  |  | C10544 | 2.378713 | 8.18818E-12 |  |  | C06034 | 3.315756 | 5.31672E-05 |
|  |  |  |  |  | C10998 | 0.609313 | 0.00027673 |  |  | C06054 | 0.27854 | 5.52481E-06 |
|  |  |  |  |  | C11045 | 0.626981 | 4.55767E-05 |  |  | C06121 | 0.353483 | 1.21146E-08 |
|  |  |  |  |  | C11111 | 0.445524 | 0.006992036 |  |  | C06313 | 2.210382 | 0.011651185 |
|  |  |  |  |  | C11118 | 0.579118 | 0.001522149 |  |  | C06424 | 0.18852 | 3.7571E-07 |
|  |  |  |  |  | C11134 | 2.971381 | 6.52777E-20 |  |  | C06426 | 0.398979 | 0.000131826 |
|  |  |  |  |  | C11136 | 4.070583 | 5.11435E-09 |  |  | C06427 | 0.161071 | 0.00040294 |
|  |  |  |  |  | C11150 | 0.082317 | 0.037962141 |  |  | C06428 | 0.150416 | 5.43123E-09 |
|  |  |  |  |  | C11154 | 2.938308 | 3.72678E-19 |  |  | C06429 | 0.213011 | 5.83079E-12 |
|  |  |  |  |  | C11289 | 4.246014 | 4.68804E-08 |  |  | C06451 | 0.125497 | 1.83426E-05 |
|  |  |  |  |  | C11301 | 3.086798 | 7.07576E-15 |  |  | C06469 | 0.123959 | 2.30481E-05 |
|  |  |  |  |  | C11352 | 3.525165 | 5.06074E-15 |  |  | C06470 | 1.748573 | 3.92144E-18 |
|  |  |  |  |  | C11404 | 4.003755 | 0.000666467 |  |  | C06554 | 0.131563 | 0.000284191 |
|  |  |  |  |  | C11457 | 0.17163 | 0.039584434 |  |  | C06645 | 0.490291 | 0.001871859 |
|  |  |  |  |  | C11821 | 0.243747 | 0.026457572 |  |  | C06670 | 0.512014 | 0.002903821 |
|  |  |  |  |  | C11849 | 0.241571 | 0.01675679 |  |  | C06677 | 0.137307 | 0.034842217 |
|  |  |  |  |  | C11896 | 0.20929 | 0.00090851 |  |  | C06772 | 0.46966 | 0.015201512 |
|  |  |  |  |  | C11924 | 0.265532 | 2.89139E-09 |  |  | C07326 | 0.183994 | 1.47543E-08 |
|  |  |  |  |  | C11989 | 0.44735 | 0.005798303 |  |  | C07888 | 7.443176 | 8.32505E-10 |
|  |  |  |  |  | C11992 | 1.54532 | 7.05334E-14 |  |  | C08156 | 0.060871 | 0.000310665 |
|  |  |  |  |  | C12100 | 0.174197 | 5.66615E-07 |  |  | C08302 | 0.470365 | 2.24219E-08 |
|  |  |  |  |  | C12286 | 0.364534 | 4.17098E-06 |  |  | C08314 | 0.058284 | 5.1596E-07 |
|  |  |  |  |  | C12888 | 0.428983 | 0.001694305 |  |  | C08316 | 0.042855 | 0.020208389 |
|  |  |  |  |  | C12986 | 0.262275 | 0.000125987 |  |  | C08320 | 0.047007 | 0.009184734 |
|  |  |  |  |  | C13309 | 6.872078 | 1.76676E-18 |  |  | C08322 | 0.193637 | 9.2932E-05 |
|  |  |  |  |  | C13358 | 0.14015 | 3.9466E-09 |  |  | C08348 | 0.524878 | 0.007782207 |
|  |  |  |  |  | C13732 | 3.136535 | 1.0009E-06 |  |  | C08357 | 5.502524 | 0.000202582 |
|  |  |  |  |  | C13829 | 4.685335 | 0.004297273 |  |  | C08362 | 0.17491 | 0.000129684 |
|  |  |  |  |  | C13846 | 0.270392 | 8.34367E-06 |  |  | C08390 | 0.079993 | 2.97887E-07 |
|  |  |  |  |  | C13856 | 0.027984 | 0.004590329 |  |  | C08486 | 0.28384 | 2.53759E-11 |
|  |  |  |  |  | C13864 | 0.224617 | 0.034777979 |  |  | C10264 | 1.707349 | 4.09958E-05 |
|  |  |  |  |  | C13914 | 0.355419 | 0.000420308 |  |  | C10544 | 2.363982 | 4.8896E-12 |
|  |  |  |  |  | C14040 | 1.549374 | 0.008205562 |  |  | C10998 | 0.514057 | 1.53059E-05 |
|  |  |  |  |  | C14179 | 0.121833 | 0.008166004 |  |  | C11045 | 0.543425 | 0.000452954 |
|  |  |  |  |  | C14422 | 0.385497 | 2.47942E-08 |  |  | C11111 | 0.404445 | 3.1353E-09 |
|  |  |  |  |  | C14472 | 4.322613 | 9.98647E-11 |  |  | C11118 | 0.553153 | 6.66805E-05 |
|  |  |  |  |  | C14606 | 0.197797 | 0.008323886 |  |  | C11134 | 2.624208 | 5.02309E-16 |
|  |  |  |  |  | C14772 | 0.103619 | 4.31431E-09 |  |  | C11136 | 3.583855 | 3.76362E-07 |
|  |  |  |  |  | C14791 | 0.272698 | 0.001509467 |  |  | C11150 | 0.118629 | 0.004216575 |
|  |  |  |  |  | C14800 | 2.549643 | 1.15144E-05 |  |  | C11154 | 3.085834 | 8.19863E-21 |
|  |  |  |  |  | C14820 | 0.261381 | 0.000975898 |  |  | C11289 | 11.55167 | 1.90845E-13 |
|  |  |  |  |  | C14829 | 3.196892 | 5.33648E-07 |  |  | C11301 | 1.996736 | 4.57279E-06 |
|  |  |  |  |  | C14847 | 3.797626 | 0.00396123 |  |  | C11352 | 3.545021 | 1.39897E-17 |
|  |  |  |  |  | C14853 | 2.859353 | 2.3174E-08 |  |  | C11404 | 3.413426 | 8.33987E-05 |
|  |  |  |  |  | C14854 | 3.91434 | 1.21937E-14 |  |  | C11457 | 0.241912 | 1.05941E-12 |
|  |  |  |  |  | C14872 | 0.283142 | 0.037823139 |  |  | C11821 | 0.265364 | 1.34237E-13 |
|  |  |  |  |  | C14877 | 3.879205 | 5.55136E-13 |  |  | C11849 | 0.201252 | 4.28545E-11 |
|  |  |  |  |  | C15532 | 0.324145 | 0.003593683 |  |  | C11896 | 0.264002 | 2.90809E-11 |
|  |  |  |  |  | C15557 | 0.206701 | 0.006060235 |  |  | C11924 | 0.311856 | 2.535E-05 |
|  |  |  |  |  | C15606 | 0.104444 | 0.04715351 |  |  | C11989 | 0.405046 | 0.0074837 |
|  |  |  |  |  | C15700 | 2.810827 | 0.01506575 |  |  | C11992 | 1.531928 | 2.20761E-14 |
|  |  |  |  |  | C15802 | 0.18035 | 2.02541E-12 |  |  | C12002 | 1.745591 | 5.88388E-12 |
|  |  |  |  |  | C15886 | 1.619928 | 0.003013577 |  |  | C12100 | 0.196607 | 1.85991E-05 |
|  |  |  |  |  | C15935 | 1.998633 | 7.07876E-05 |  |  | C12106 | 0.085712 | 1.43634E-05 |
|  |  |  |  |  | C15974 | 0.154283 | 0.008552071 |  |  | C12286 | 0.365635 | 0.000254833 |
|  |  |  |  |  | C16146 | 0.581533 | 0.001393215 |  |  | C12294 | 0.631328 | 8.0731E-13 |
|  |  |  |  |  | C16155 | 0.270861 | 0.001767799 |  |  | C12986 | 0.311631 | 2.51605E-05 |
|  |  |  |  |  | C16186 | 0.497394 | 1.54311E-06 |  |  | C13309 | 5.033696 | 2.08139E-12 |
|  |  |  |  |  | C16196 | 0.232101 | 4.01851E-07 |  |  | C13358 | 0.277749 | 1.3266E-05 |
|  |  |  |  |  | C16249 | 7.219119 | 2.0251E-11 |  |  | C13732 | 2.387539 | 0.001551191 |
|  |  |  |  |  | C16261 | 0.288984 | 0.000113379 |  |  | C13790 | 0.19191 | 4.03646E-12 |
|  |  |  |  |  | C16265 | 3.759299 | 1.57726E-18 |  |  | C13831 | 0.595223 | 5.65654E-11 |
|  |  |  |  |  | C16308 | 0.532835 | 0.00833511 |  |  | C13846 | 0.255187 | 1.60329E-08 |
|  |  |  |  |  | C16358 | 0.035715 | 0.047880167 |  |  | C13856 | 0.114283 | 1.10029E-08 |
|  |  |  |  |  | C16366 | 0.324013 | 0.000089247 |  |  | C13864 | 0.202315 | 0.016541825 |
|  |  |  |  |  | C16488 | 0.225351 | 0.000828489 |  |  | C13914 | 0.323255 | 0.000988602 |
|  |  |  |  |  | C16489 | 4.514646 | 5.09102E-16 |  |  | C14040 | 2.66812 | 0.000467685 |
|  |  |  |  |  | C16497 | 0.3481 | 0.000108767 |  |  | C14179 | 0.18253 | 0.00120737 |
|  |  |  |  |  | C16613 | 0.010392 | 0.003314369 |  |  | C14218 | 0.21026 | 3.84414E-09 |
|  |  |  |  |  | C16660 | 4.095311 | 3.64143E-11 |  |  | C14422 | 0.386209 | 1.35889E-07 |
|  |  |  |  |  | C16682 | 1.797734 | 0.007789941 |  |  | C14472 | 4.056434 | 1.22291E-08 |
|  |  |  |  |  | C16741 | 0.148775 | 0.007681325 |  |  | C14606 | 0.153253 | 0.003693519 |
|  |  |  |  |  | C16884 | 0.410105 | 0.000775187 |  |  | C14772 | 0.137184 | 3.04357E-09 |
|  |  |  |  |  | C17467 | 0.149788 | 0.000746327 |  |  | C14791 | 0.273523 | 0.000328375 |
|  |  |  |  |  | C17938 | 2.708811 | 4.21694E-08 |  |  | C14800 | 2.955044 | 1.73518E-06 |
|  |  |  |  |  | C18319 | 0.438525 | 0.022370371 |  |  | C14820 | 0.186785 | 3.05407E-15 |
|  |  |  |  |  | C19691 | 1.849214 | 1.61184E-08 |  |  | C14829 | 3.864013 | 1.81798E-09 |
|  |  |  |  |  | C19910 | 0.433758 | 0.012642391 |  |  | C14847 | 38.64466 | 0.000954827 |
|  |  |  |  |  | C21298 | 0.317352 | 8.17939E-06 |  |  | C14853 | 4.134124 | 3.53666E-05 |
|  |  |  |  |  | HMDB0000333 | 0.242315 | 7.14181E-06 |  |  | C14854 | 3.925447 | 8.50743E-16 |
|  |  |  |  |  | HMDB0000354 | 0.220898 | 1.14083E-08 |  |  | C14860 | 0.350306 | 0.000418148 |
|  |  |  |  |  | HMDB0000407 | 0.239829 | 3.96529E-08 |  |  | C14872 | 0.416316 | 0.03483161 |
|  |  |  |  |  | HMDB00005 | 0.167233 | 0.011052347 |  |  | C14877 | 5.305632 | 7.41526E-11 |
|  |  |  |  |  | HMDB0000555 | 0.301833 | 0.008618767 |  |  | C15532 | 0.278037 | 6.09421E-08 |
|  |  |  |  |  | HMDB0000678 | 0.143808 | 5.26651E-10 |  |  | C15606 | 0.103326 | 0.009669877 |
|  |  |  |  |  | HMDB0000695 | 0.204229 | 2.34687E-05 |  |  | C15802 | 0.241247 | 8.57913E-15 |
|  |  |  |  |  | HMDB0000701 | 0.200843 | 8.32133E-05 |  |  | C15974 | 0.18037 | 0.000270574 |
|  |  |  |  |  | HMDB0000705 | 0.265131 | 0.00843833 |  |  | C16038 | 0.356102 | 0.002912545 |
|  |  |  |  |  | HMDB0000734 | 0.467187 | 6.50749E-06 |  |  | C16186 | 0.462469 | 7.15512E-08 |
|  |  |  |  |  | HMDB0000750 | 0.158657 | 1.77452E-05 |  |  | C16196 | 0.234617 | 7.12237E-09 |
|  |  |  |  |  | HMDB0000766 | 0.194439 | 1.00976E-15 |  |  | C16249 | 15.71633 | 1.00762E-15 |
|  |  |  |  |  | HMDB0000888 | 3.369223 | 1.86752E-12 |  |  | C16261 | 0.214601 | 2.96413E-05 |
|  |  |  |  |  | HMDB0001624 | 0.220571 | 3.67361E-06 |  |  | C16265 | 3.785078 | 8.80648E-15 |
|  |  |  |  |  | HMDB0001863 | 0.282215 | 0.000507251 |  |  | C16358 | 0.06794 | 0.000929499 |
|  |  |  |  |  | HMDB00140 | 2.838894 | 1.25633E-16 |  |  | C16366 | 0.311559 | 0.000111913 |
|  |  |  |  |  | HMDB00770 | 10.35285 | 2.96587E-05 |  |  | C16488 | 0.226809 | 3.69129E-06 |
|  |  |  |  |  |  |  |  |  |  | C16489 | 7.743756 | 1.00529E-08 |
|  |  |  |  |  |  |  |  |  |  | C16497 | 0.446613 | 0.00688207 |
|  |  |  |  |  |  |  |  |  |  | C16526 | 0.044323 | 0.014119994 |
|  |  |  |  |  |  |  |  |  |  | C16613 | 0.055418 | 0.018470106 |
|  |  |  |  |  |  |  |  |  |  | C16614 | 0.15973 | 0.004971581 |
|  |  |  |  |  |  |  |  |  |  | C16660 | 5.20621 | 5.36231E-09 |
|  |  |  |  |  |  |  |  |  |  | C16741 | 0.188191 | 9.30573E-05 |
|  |  |  |  |  |  |  |  |  |  | C16884 | 0.571507 | 0.017670566 |
|  |  |  |  |  |  |  |  |  |  | C16939 | 0.228037 | 0.010045854 |
|  |  |  |  |  |  |  |  |  |  | C17432 | 1.69656 | 0.006873821 |
|  |  |  |  |  |  |  |  |  |  | C17467 | 0.192651 | 1.32928E-12 |
|  |  |  |  |  |  |  |  |  |  | C17938 | 3.587668 | 6.90305E-06 |
|  |  |  |  |  |  |  |  |  |  | C18043 | 9.094221 | 0.000658355 |
|  |  |  |  |  |  |  |  |  |  | C18319 | 0.290897 | 0.008269862 |
|  |  |  |  |  |  |  |  |  |  | C19691 | 1.873533 | 3.80144E-06 |
|  |  |  |  |  |  |  |  |  |  | C19910 | 0.45268 | 5.99254E-05 |
|  |  |  |  |  |  |  |  |  |  | C21298 | 0.411639 | 0.000270215 |
|  |  |  |  |  |  |  |  |  |  | D06890 | 0.218515 | 0.002079613 |
|  |  |  |  |  |  |  |  |  |  | G13036 | 0.600803 | 0.030267672 |
|  |  |  |  |  |  |  |  |  |  | HMDB0000333 | 0.267693 | 0.00010824 |
|  |  |  |  |  |  |  |  |  |  | HMDB0000354 | 0.166336 | 5.44253E-12 |
|  |  |  |  |  |  |  |  |  |  | HMDB0000407 | 0.189862 | 7.51835E-07 |
|  |  |  |  |  |  |  |  |  |  | HMDB00005 | 0.216142 | 1.2375E-13 |
|  |  |  |  |  |  |  |  |  |  | HMDB0000555 | 0.363232 | 0.027449026 |
|  |  |  |  |  |  |  |  |  |  | HMDB0000622 | 0.126974 | 0.001541529 |
|  |  |  |  |  |  |  |  |  |  | HMDB0000678 | 0.137696 | 2.47364E-06 |
|  |  |  |  |  |  |  |  |  |  | HMDB0000695 | 0.416745 | 0.001185096 |
|  |  |  |  |  |  |  |  |  |  | HMDB0000701 | 0.137267 | 1.29973E-05 |
|  |  |  |  |  |  |  |  |  |  | HMDB0000734 | 0.475572 | 3.47758E-07 |
|  |  |  |  |  |  |  |  |  |  | HMDB0000750 | 0.173646 | 7.80235E-08 |
|  |  |  |  |  |  |  |  |  |  | HMDB0000766 | 0.258674 | 1.63229E-14 |
|  |  |  |  |  |  |  |  |  |  | HMDB0000888 | 4.047961 | 1.24741E-09 |
|  |  |  |  |  |  |  |  |  |  | HMDB0001624 | 0.131069 | 7.09542E-05 |
|  |  |  |  |  |  |  |  |  |  | HMDB0001863 | 0.440127 | 0.004811965 |
|  |  |  |  |  |  |  |  |  |  | HMDB0002404 | 2.802687 | 0.003771809 |
|  |  |  |  |  |  |  |  |  |  | HMDB00079 | 0.110542 | 2.99911E-05 |
|  |  |  |  |  |  |  |  |  |  | HMDB00140 | 1.597753 | 0.002072176 |
|  |  |  |  |  |  |  |  |  |  | HMDB00770 | 10.43545 | 1.50547E-07 |
